# Single-cell RNA sequencing reveals *in vivo* signatures of SARS-CoV-2-reactive T cells through ‘reverse phenotyping’

**DOI:** 10.1101/2020.12.07.20245274

**Authors:** David S. Fischer, Meshal Ansari, Karolin I. Wagner, Sebastian Jarosch, Yiqi Huang, Christoph H. Mayr, Maximilian Strunz, Niklas J. Lang, Elvira D’Ippolito, Monika Hammel, Laura Mateyka, Simone Weber, Lisa S. Wolff, Klaus Witter, Isis E. Fernandez, Gabriela Leuschner, Kathrin Milger, Marion Frankenberger, Lorenz Nowak, Katharina Heinig-Menhard, Ina Koch, Mircea G. Stoleriu, Anne Hilgendorff, Jürgen Behr, Andreas Pichlmair, Benjamin Schubert, Fabian J. Theis, Dirk H. Busch, Herbert B. Schiller, Kilian Schober

## Abstract

The *in vivo* phenotypic profile of T cells reactive to severe acute respiratory syndrome (SARS)-CoV-2 antigens remains poorly understood. Conventional methods to detect antigen-reactive T cells require *in vitro* antigenic re-stimulation or highly individualized peptide-human leukocyte antigen (pHLA) multimers. Here, we used single-cell RNA sequencing to identify and profile SARS-CoV-2-reactive T cells from Coronavirus Disease 2019 (COVID-19) patients. To do so, we induced transcriptional shifts by antigenic stimulation *in vitro* and took advantage of natural T cell receptor (TCR) sequences of clonally expanded T cells as barcodes for ‘reverse phenotyping’. This allowed identification of SARS-CoV-2-reactive TCRs and revealed phenotypic effects introduced by antigen-specific stimulation. We characterized transcriptional signatures of currently and previously activated SARS-CoV-2-reactive T cells, and showed correspondence with phenotypes of T cells from the respiratory tract of patients with severe disease in the presence or absence of virus in independent cohorts. Reverse phenotyping is a powerful tool to provide an integrated insight into cellular states of SARS-CoV-2-reactive T cells across tissues and activation states.

## MAIN TEXT

COVID-19 is a new form of viral pneumonia^1^ caused by SARS-CoV-2^2^ and is affecting 67,073,749 patients, with 1,536,072 deaths, world-wide (source: Johns Hopkins University, as of December 07 2020). Adaptive immunity is swiftly induced upon COVID-19 infection^3^. T cells play a central role in this process, and are implicated in contributing both to long-lasting immunity as well as to putative immunopathology^4,5^. A thorough understanding of T cell responses to SARS-CoV-2 is therefore urgently needed.

Immunodominant SARS-CoV-2 antigen specificities have been identified with unprecedented speed for an emerging pathogen, and phenotypic characterization of antigen-reactive T cells has quickly been performed by a plethora of studies^6,7,16,8–15^. While there is general agreement that SARS-CoV-2-reactive T cells are activated and differentiate during the course of the immune response, the extents of activation and differentiation are controversial^4,5^.

Methodologies for characterization of antigen-reactive T cells differ and thereby obscure a clear insight into T cell phenotypes. Some reports have described phenotypes of activated T cells during the course of COVID-19 without assessment of *bona fide* antigen specificity, whereas other studies restricted phenotypic characterization to antigen-reactive T cells^4^. Although such restriction to antigen-reactive T cells is clearly desired, it entails methodological challenges on its own: initial detection of antigen-reactive T cells usually requires *in vitro* re-stimulation with SARS-CoV-2 antigens. Upon re-stimulation, antigen-reactive cells can be defined through upregulation of activation markers (such as CD154) or release of effector cytokines (such as IFN*γ*)^17^. This, however, automatically also introduces major phenotypic biases for any downstream profiling since *in vitro* activated T cells show markedly different phenotypes compared to unperturbed cells (i.e. cells as they are *in vivo* – technically, any analysis outside the body is *ex vivo*, but to simplify terminology, we here refer to cells as they are ‘*in vivo*’ when cells are directly analyzed *ex vivo*, and ‘*in vitro*’, when cells are e.g. additionally re-stimulated *ex vivo*)^18^. In order to address these well-known challenges, peptide human leukocyte antigen (pHLA) multimers have been developed^19^. Through removal of specifically designed pHLA multimers (‘Streptamers’), unperturbed cellular phenotypes of antigen-reactive T cells can be even completely preserved^20^. Yet, pHLA multimer technology requires previous definition of specific epitopes and is restricted to individual HLA haplotypes. Furthermore, pHLA multimers are often difficult to generate for HLA class II-restricted CD4 T cells^21^. These limitations represent a significant technical obstacle to investigate unbiased *in vivo* phenotypic profiles of antigen-reactive T cells.

Vast resources of deeply profiled immune cells from COVID-19 patients do already exist, and are bundled e.g. by the Human Cell Atlas initiative (www.humancellatlas.org). It is difficult, however, for any single study to cover different patient disease states and sample sites, together with detailed clinical metadata and deep profiling down to single-cell resolution, including antigen-specific analyses. For example, few studies have investigated T cell states across different tissue sites (e.g. from peripheral blood and the respiratory tract) in identical COVID-19 patients^5^. Generally, antigen-specific T cells are expected to have different activation states in different tissues depending on the course of disease. When viral antigen is still present in the respiratory tract, T cells at the site of infection should show a more activated phenotype compared to peripheral blood T cells which are not being activated by the antigen at the time of analysis. In this context, difficulties to clearly define the phenotypic profile of currently and previously activated SARS-CoV-2-reactive T cells also hinder precise contextualization of T cell signatures across publicly available data sets.

Single-cell RNA sequencing (scRNA seq) allows simultaneous analysis of the global cellular transcriptome as well as identification of T cell receptor (TCR) sequences^22,23^. Recently, it has been demonstrated that scRNA seq can be used to reveal activation-induced phenotypic profiles of antigen-reactive T cells^24^. Samples can be split up after isolation from the tissue, stimulated *in vitro* in an antigen-specific manner (and, for comparison, left unstimulated), and then sequenced. The natural TCR can thereby serve as a barcode to link T cells of the activated and unstimulated condition belonging to the same *in vivo* expanded clonotype with a common antigen specificity.

We here used such ‘reverse phenotyping’ (Extended Data Fig. 1a) to identify and characterize SARS-CoV-2-reactive T cells from peripheral blood of severely diseased COVID-19 patients. To this end, we used a SARS-CoV-2 spike protein peptide mix for antigenic stimulation and autologous patient cells for antigen presentation. After scRNA seq, we detected clonotype-dependent transcriptional shifts with high robustness. We applied this method to CD8 and diverse T helper (Th) lineage subsets of CD4 T cells simultaneously – thereby testing dozens of TCRs for antigen reactivity based on a functional readout without prior knowledge of epitopes and independent from the specific HLA haplotype. Using CRISPR/Cas9-mediated orthotopic TCR replacement^25^, we generated TCR-transgenic T cells and validated the SARS-CoV-2 spike protein antigen reactivity of TCRs from selected clonotypes. Transcriptome comparisons of antigen-reactive and antigen-unreactive clonotypes in stimulated and unstimulated cells revealed systematic phenotypic biases introduced by antigenic stimulation, and allowed exploration of unperturbed *in vivo* phenotypes of SARS-CoV-2-reactive T cells. Finally, we performed an integrated analysis of respiratory material from identical and further patients from our own cohort, as well as from independent reference cohorts from Berlin^26^, Shenzhen^27^ and Chicago^28^. This investigation included 279,663 single cells from 50 COVID-19 patients with different disease severities and time points during or after viral infection, as well as healthy controls. Remarkably, transcriptomic states of respiratory T cells from virus-positive patients were most similar to *in vitro* stimulated reactive T cells from peripheral blood, consistent with a ‘hot’ phenotype driven by acute activation at the site of infection. In contrast, unperturbed *in vivo* phenotypes of antigen-reactive T cells from peripheral blood matched with ‘cold’ phenotypes found in the respiratory tract of virus-negative patients that had previously cleared the virus. Thus, we present transcriptional shifts from acute disease to resolution after virus clearance in antigen reactive CD4 and CD8 T cells during the course of COVID-19.

## Results

### Patient characteristics and experimental set-up for ‘reverse phenotyping’

We acquired respiratory material (tracheal aspirates) as well as peripheral blood mononuclear cells (PBMCs) from severely diseased patients with COVID-19 (Fig. 1a; Extended Data Table 1). Consistent with the risk profile for severe disease, seven of nine patients in this ‘Munich cohort’ were male and age ranged from 51-82 years (median 79 years). All patients were treated in an intensive care unit (ICU) and had been on a respirator for 8-38 days (median 29 days) at the time of sampling. After initial detection of SARS-CoV-2 via PCR, eight of nine patients had turned consistently virus-negative or had at least one prior negative test result at the time of sampling. Ultimately, two patients deceased and seven patients recovered. ScRNA seq (3’ transcriptomics) was performed on tracheal aspirates from all nine patients. For two patients (‘GT_3’ and ‘GT_2’), we stimulated PBMCs with SARS-CoV-2 spike protein peptide mix or left control samples unstimulated, and likewise performed scRNA seq (5’ transcriptomics and VDJ) after flow cytometry-assisted cell sorting of > 10.000 CD4 and CD8 T cells for each patient.

**Figure 1.**
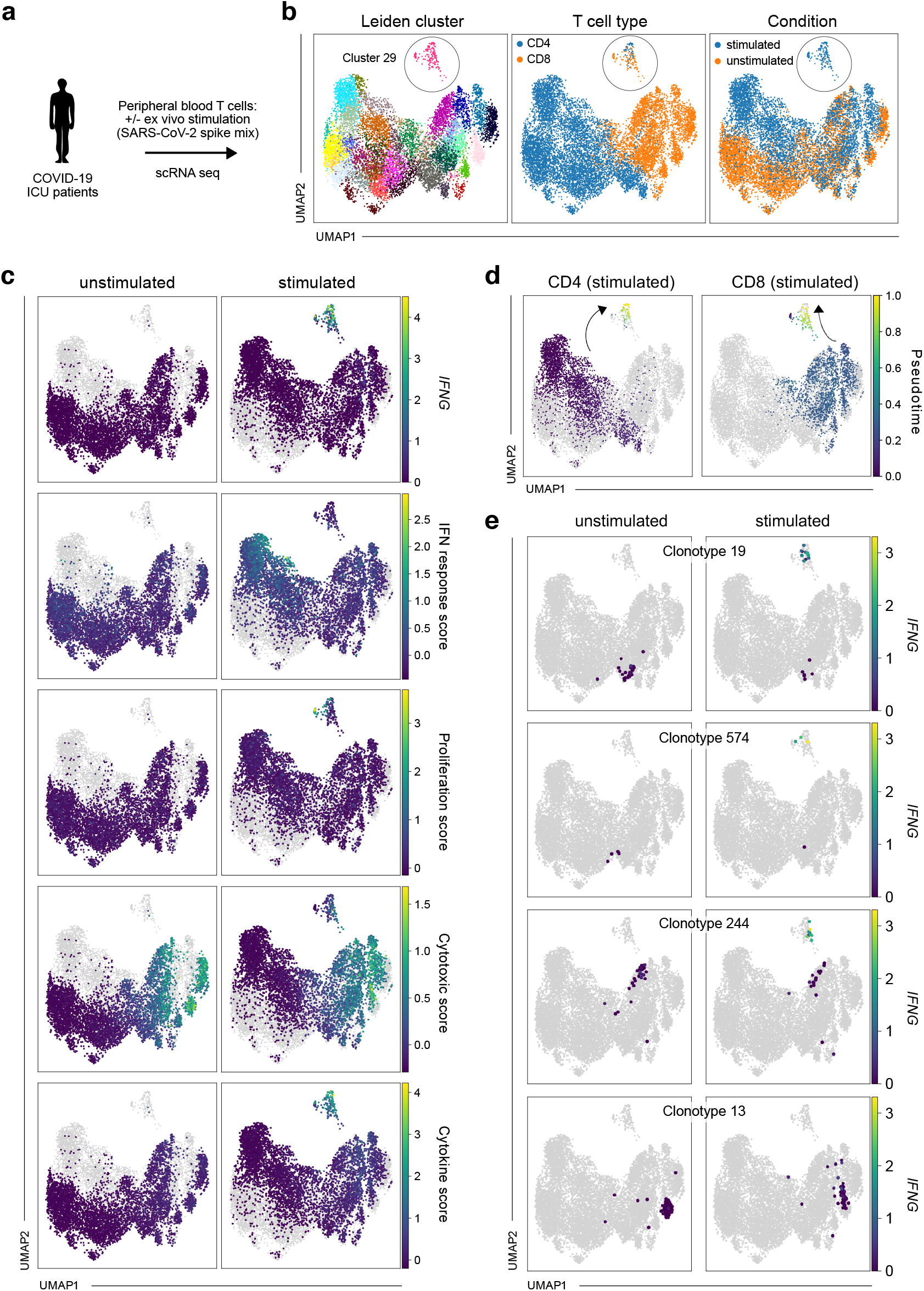
Single-cell RNA sequencing allows detection of transcriptional shifts induced by antigen-specific stimulation in TCR-barcoded clonotypes. **a**, Experimental setup; cells from tracheal aspirates and peripheral blood T cells (flow cytometry-sorted for CD4 or CD8 positivity) of intensive care unit (ICU) patients with COVID-19 were profiled by single-cell RNA sequencing (scRNA seq). Before scRNA seq, peripheral blood T cells were stimulated with SARS-CoV-2 spike protein peptide mix or left untreated. **b**, Uniform manifold approximation and projection (UMAP) of Leiden clusters (left panel), CD4 and CD8 T cells (middle panel) and stimulated and unstimulated T cells (right panel). **c**, *IFNG* expression, IFN response score, proliferation score, cytotoxic score and cytokine score (from top to bottom) in unstimulated (left panels; stimulated cells shown in grey) or stimulated (right panels; unstimulated cells shown in grey) T cells. **d**, Stimulated CD4 and CD8 T cells with highlighted pseudotime after defining the endpoint of pseudotime at the tip of cluster 29. **e**, *IFNG* expression in unstimulated or stimulated T cells for four representative clonotypes. For each clonotype, cells belonging to that clonotype are shown in an individual panel pair (cells from unstimulated condition in left panels, cells from stimulated condition in right panels), while cells not belonging to that clonotype are shown in grey.

### Antigen-induced transcriptional shifts in PBMCs

After preprocessing, uniform manifold approximation and projection (UMAP)^29^ of stimulated and unstimulated T cells identified a group of cells in both patients which was separate from all other cells (Leiden cluster 29 in patient GT_3; Fig. 1b). The cluster encompassed both CD4 and CD8 cells and consisted almost exclusively of cells from the stimulated condition. We hypothesized that this distinct stimulation-induced cluster represented antigen-reactive T cells. Indeed, only the cells from that cluster upregulated *IFNG* (the gene encoding Interferon-*γ*, IFN*γ*) after stimulation (Fig. 1c).

Antigen-reactive CD4 Th cell lineages (particularly those that are not Th1 cells) do not necessarily express IFN*γ* in response to antigen^21^. For this reason, and in order to not focus on upregulation of a single gene, we explored more holistic and unbiased antigen-reactive response scores for CD4 and CD8 T cells, as previously identified by scRNA seq after aCD3/aCD28 stimulation^18^. Upon stimulation, CD4 T cells have been described to show sequentially enriched IFN response (early activation) and proliferation (late activation) scores^18^. Accordingly, in our analysis CD4 T cells showing high proliferation scores were exclusively present upon stimulation, and only in cluster 29 (Fig. 1c). Interestingly, many CD4 cells outside the cluster (on the left side of the UMAP) underwent a general transcriptional shift through stimulation, whereas this did not occur for CD8 cells. These seemingly unspecifically shifting CD4 T cells were also the ones that showed a high IFN response (early activation) score (Fig. 1c) or e.g. expression of the IFN*γ* receptor gene *IFNGR2* (Extended Data Fig. 1b) upon stimulation. This confirms previous observations of differential CD4 transcriptomics through sensing of TCR-triggered cytokines such as IFN*γ*^18^. The source of the sensed cytokines could be antigen-reactive CD4 or CD8 T cells that are neighboring and stimulated in the culture system.

CD8 T cells have been described to undergo sequential transcriptional states upon activation that are reflected by high ‘cytotoxic’ (early activation) or ‘cytokine secretion’ scores (late activation)^18^. While in our analysis almost all CD8 T cells showed a high cytotoxic score, high cytokine scores were exclusively confined to cells belonging to the antigen-reactive cluster 29 in the stimulated condition (Fig. 1c). Differential gene expression analysis confirmed that cluster 29 CD8 T cells showed upregulation of genes as *IFNG, TNF, IL2, CCL3, CCL4* or *GZMB* (Extended Data Tables 2-3), in line with CD8 T cell activation. We next aimed to elucidate if activation-induced transcriptomic changes followed consistent gradients. To this end, we defined the tip of cluster 29 as an ‘end point’ in pseudo-time, and ordered all stimulated cells along this trajectory of transcriptomic similarity. Mean transcriptomic similarity of clonotypes to this pseudotime end-point correlated with T cell activation scores, with reactive CD4 and CD8 T cells feeding into cluster 29 from different directions (Fig. 1d). This shows that our approach could resolve a spectrum of activation states across clonotypes. We validated our findings in the second severely ill patient (‘GT_2’) with COVID-19 for which we stimulated or did not stimulate peripheral blood (PB) CD4 and CD8 T cells with SARS-CoV-2 spike protein peptide mix. Again, antigen stimulation induced transcriptional shifts giving rise to a specific ‘reactive Leiden cluster’ (in this patient cluster 36; Extended Data Fig. 2a-b). Data integration showed that the reactive Leiden clusters from both patients showed convergent phenotypes (Extended Data Fig. 2c).

### Identification of antigen-reactive clonotypes

Using the TCR information from VDJ sequencing, we next investigated whether differential transcriptional responses to antigen stimulation could be attributed to specific clonotypes. We detected CDR3*αβ* sequences in 92% of analyzed T cells (69.9% fully paired CDR3*αβ*; 3.4% CDR3*α* only; 26.7% CDR3*β* only). In order to ensure the clonotypic nature of the investigated cells, we first only included clonotypes based on identical fully paired CDR3*αβ* sequences in our analysis. For both CD4 and CD8 T cell clonotypes, we recovered highly similar cell numbers in the stimulated and non-stimulated condition (Extended Data Fig. 3a). This enabled us to identify clonotypes that underwent transcriptional shifts upon antigenic stimulation (Fig. 1e). Interestingly, we observed a heterogenous response pattern within clonotypes, with some cells moving into the antigen-reactive cluster 29 (with concomitant upregulation of *IFNG*) and other cells staying in a transcriptional state that is consistent with their *in vivo* phenotype without *in vitro* re-stimulation (Fig. 1e). Possibly, these response patterns are a result of stochastic antigen encounters in the experimental stimulation system *in vitro*. The remarkable degree of transcriptional synchronization specific for each TCR *in vivo* confirms previous observations of TCR-mediated phenotypic imprinting^30,31^ and renders high credibility to the assessment of clonotypes that are as small as a few cells per condition group (such as clonotype 574; Fig. 1e). Overall, the distinct transcriptional shifts induced by antigenic stimulation in specific clonotypes strongly suggested the presence of antigen-reactive T cells.

While mean proliferation scores (for CD4 T cells) or mean cytokine scores (for CD8 T cells) generally correlated with mean *IFNG* expression per clonotype after antigen-specific stimulation (Extended Data Fig. 3b), *IFNG* expression seemed to be the more sensitive read-out. We therefore decided to define antigen-reactive T cells by significantly enhanced *IFNG* expression after antigen-specific stimulation (Fig. 2a-b). Among clonotypes with at least three cells in each stimulation condition, we detected five and four antigen-reactive clonotypes for CD8 and CD4 T cells, respectively. Apart from clonotype 574, reactive clonotypes were large in size, consistent with previous *in vivo* activation and clonal expansion.

**Figure 2.**
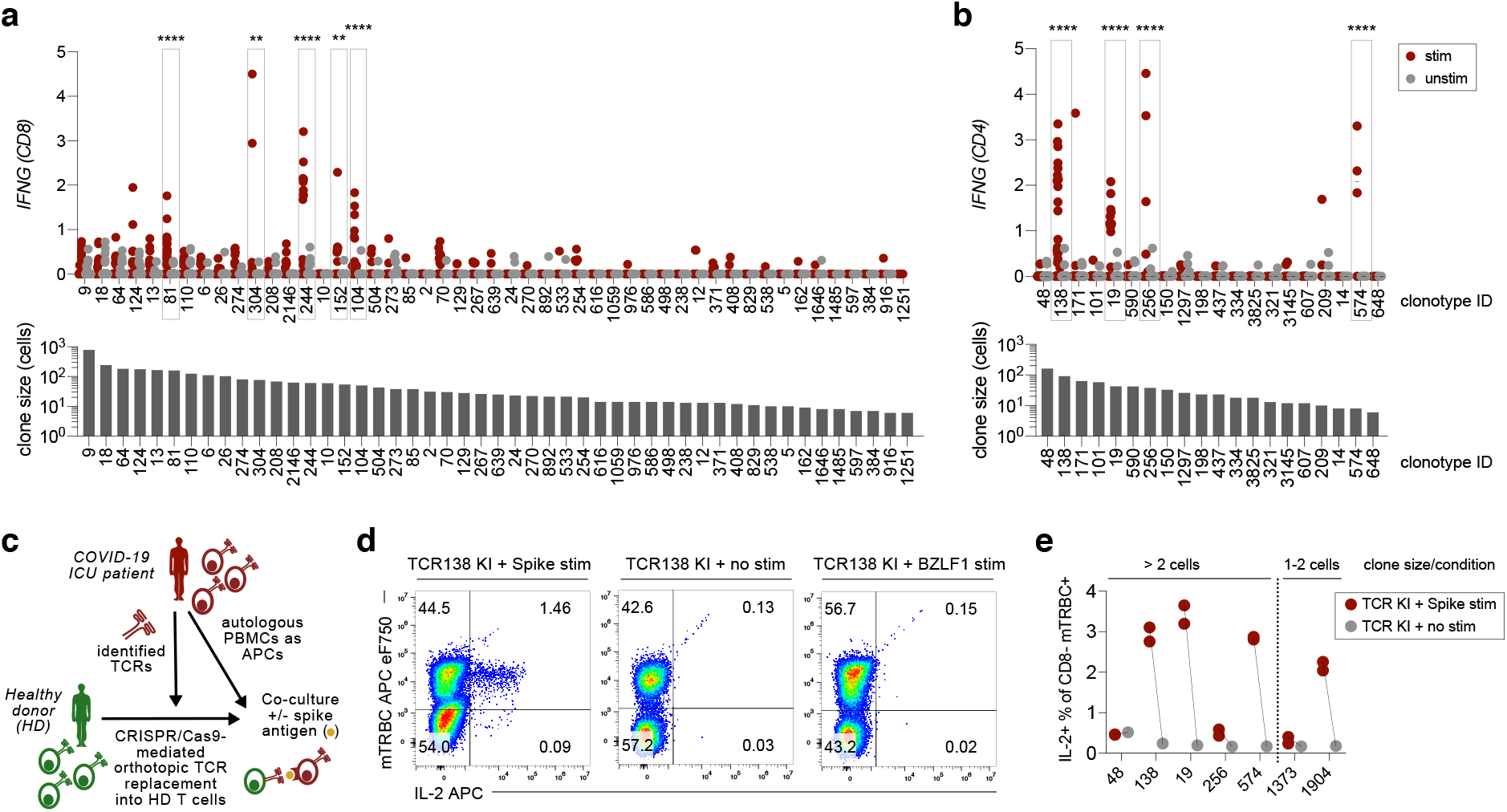
Identification and validation of SARS-CoV-2 antigen-reactive T cell receptors. **a**, Top: *IFNG* expression of CD8 clonotypes after antigenic or no stimulation. Only clonotypes with at least three cells in each condition and as defined by a unique *αβ* CDR3 sequence were included in this analysis. Antigen-reactive clonotypes (defined through statistically significant upregulation of *IFNG*) are highlighted. Bottom: Clonotype sizes in numbers of total cells analyzed. **b**, As in a, but for CD4 clonotypes. **c**, Experimental setup; T cells from healthy donors were equipped with TCRs identified from COVID-19 patients by CRISPR/Cas9-mediated orthotopic TCR replacement (OTR); transgenic T cells were co-incubated with antigen-loaded patient PBMCs and reactivity was investigated by intracellular cytokine staining. **d**, Flow-cytometric analysis of antigen-stimulated TCR-engineered T cells, 1 week after OTR; representative data are shown for TCR139 after stimulation with SARS-CoV-2 spike protein peptide mix, no stimulation and stimulation with irrelevant EBV antigen BZLF1 peptide mix (negative controls); mTRBC: murine constant region of the TCR beta chain incorporated into transgenic TCRs for detection; shown gates are pre-gated for CD3+ CD8-living lymphocytes. **e**, Quantification of spike antigen-specific reactivity for selected clonotypes tested in (**b)** as well as two additional small clonotypes; for antigen-specific transcriptional shifts detected by initial scRNA seq for the respective clones see Fig. 1e, Extended Data Fig. 5c and Extended Data Fig. 7b. Statistical analysis by two-way ANOVA (**** each for treatment effect and clonotype distribution) followed by Sidak’s multiple comparisons test * p < 0.05, ** p < 0.01, *** p < 0.001, **** p < 0.0001 (**a, b**).

This first analysis was strictly restricted to fully paired *αβ* TCRs. We also explored all clonotypes defined by unique double (CDR3*αβ*) *or* single (CDR3*α* or CDR3*β*) chain sequences (Extended Data Fig. 4). This yielded two further CD8 clonotypes (clonotypes 651 and 548). Upon further examination, these single-chain (CDR3*β* only) clonotypes shared CDR3*β* sequences with clonotypes 81 and 244, respectively, and showed highly similar *IFNG* upregulation (Extended Data Fig. 4a) and global transcriptomic states *in vivo* and upon stimulation (Extended Data Fig. 4b) compared to their fully paired partner clonotypes. We also identified an additional single-chain CD4 clonotype (clonotype 136), for which no fully paired partner clonotype existed (Extended Data Fig. 4c). These findings are best explained by technical variability of CDR3*α* sequence detection. Since we conversely did not find any false-positive pairing of TCRs sharing the same CDR3*β*, but different CDR3*α* sequences, we included these single-chain clonotypes into downstream phenotypic analyses.

Having defined reactive clonotypes through significant upregulation of *IFNG* after stimulation, we aimed to ensure we would not miss clonotypes that show antigen-dependent reactivity through other means than *IFNG* upregulation. We therefore again analyzed cytokine / cytotoxicity scores for CD8 T cells and proliferation / IFN response scores for CD4 T cells in a clonotype-dependent manner. While for CD8 T cells, no clonotype-dependent enrichment was visible after stimulation in the cytotoxicity score (early activation), in the cytokine scores (late activation) we detected broad statistically significant signals, which did not mirror global transcriptomic shifts in sanity checks and therefore seemed to reflect rather unspecific changes (e.g. compare statistically significant results for clonotype 13 in Extended Data Fig. 4a with missing reactivity in Fig. 1d). In CD4 T cells, clonotype and stimulation-dependent IFN responses (early activation) likewise yielded inconsistent results (Extended Data Fig. 4c), whereas the proliferation score (late activation) identified the same antigen-reactive clonotypes as defined through *IFNG* upregulation except for one clonotype, which was missed by the proliferation score (Extended Data Fig. 4c). *IFNG* expression of all reactive clonotypes highly correlated with end-stage transcriptional states in pseudo-time after stimulation (Extended Data Fig. 5a-b), providing further unbiased indication for consistent and global transcriptional shifts of reactive clonotypes upon antigen stimulation. Finally, targeted exploration of markers alternative to *IFNG*, such as *MKI67, TNFRSF9* (coding for CD137), IL-2, IL-4, IL-5, IL-9, IL-10, IL-13, IL-17 or IL-22 did not yield any statistically significant antigen-induced upregulation in a clonotype-dependent manner.

To capture the global antigen-specific reactivity landscape of the TCR repertoire, we calculated correlations between clonotypes in terms of the general transcriptomic shift these clonotypes underwent upon stimulation. This revealed two groups of clonotypes that showed high inter-clonotype correlations within the respective group (Extended Data Fig. 5c). The larger group consisted mostly of CD8 clonotypes assessed as non-reactive to antigen based on the lack of statistically significant *IFNG* upregulation and recruitment into cluster 29. The few clonotypes that are antigen-reactive and/or belong to cluster 29 in that larger group may represent weakly cross-reactive clonotypes. A smaller group contained almost exclusively CD4 and CD8 clonotypes that we defined as antigen-reactive based on statistically significant *IFNG* upregulation (Extended Data Fig. 4) and/or were present in the stimulation-induced cluster 29 (Fig. 1b).

In order to experimentally validate the antigen reactivity of our selected clonotypes, we generated TCR-transgenic T cells by CRISPR/Cas9-mediated orthotopic TCR replacement (OTR)^25^. Through this technology, transgenic TCRs are knocked into the endogenous TCR gene locus, thereby simultaneously placing the transgenic TCR under physiological transcriptional control and knocking-out the endogenous TCR. We equipped healthy donor T cells by OTR with identified CD4 TCRs (Fig. 2c). From our screening of clonotypes with at least three cells in each condition (Fig. 2b), we included the four TCRs defined as reactive (TCRs 138, 19, 256 and 574) and the TCR from the largest CD4 clonotype (TCR 48), which did not show antigen-induced transcriptomic changes. After TCR knock-in (KI), T cells with TCRs that were previously defined as reactive (TCRs 138, 19, 256 and 574) all showed SARS-CoV-2 spike antigen-dependent reactivity, whereas TCR 48 KI T cells did not (Fig. 2d-e). To test the sensitivity of our system, we additionally investigated TCRs from two very small clonotypes (1373 and 1904), which were not included in the initial definition of reactive clonotypes (Fig. 2b) since they had less than three cells in each condition. Remarkably, we could validate reactivity for TCR 1904, which had a transcriptional shift into cluster 29 with concomitant *IFNG* upregulation in one of two cells in the stimulated condition, whereas reactivity for TCR 1373 was missing, as predicted based on the lack of *IFNG* upregulation and no movement into cluster 29 (Fig. 2e, Extended Data Fig. 5d). Overall, these data reinforced our approach to use stimulation-induced *IFNG* upregulation as a surrogate marker to detect antigen-reactive clonotypes and functionally validated SARS-CoV-2 reactive TCRs.

### Reverse phenotyping reveals in vitro antigen re-stimulation-induced phenotypic biases in peripheral blood T cells

The identification of antigen-reactive clonotypes enabled us to perform ‘reverse phenotyping’ by ‘looking back’ at the phenotype without re-stimulation for clonotypes that undergo defined functional changes after re-stimulation, using their T cell receptor sequence as natural barcodes. In other words, this enabled us to define antigen-reactive clonotypes by a functional read-out, and then investigate the phenotype of these cells *would have had* if they had not been stimulated. Through this, we can on the one hand investigate systematic phenotypic effects that are introduced by antigenic re-stimulation *in vitro*, and on the other hand explore the unperturbed *in vivo* phenotype of antigen-reactive T cells.

Upon re-stimulation *in vitro*, antigen-reactive CD4 T cells upregulated *TNFRSF9* (encoding CD137) and effector cytokines: *IFNG, TNF, XCL1, XCL2* or *CCL3* were absent in unstimulated reactive CD4 T cells, but strongly induced upon stimulation (Fig. 3a, Extended Data Fig. 6a). *GZMB* or *CCL4* were expressed in reactive CD4 T cells (as well as in non-reactive cells) in the unstimulated condition already, but expression was boosted by stimulation for reactive CD4 T cells only (Fig. 3a). Of note, co-inhibitory molecules, such as PD-1 (*PDCD1*), *LAG3* or *TIGIT*, were almost absent on antigen-reactive cells before stimulation, and only induced thereafter, indicating that detection of these molecules in *in vitro* stimulated T cells reflects T cell activation rather than exhaustion.

**Figure 3.**
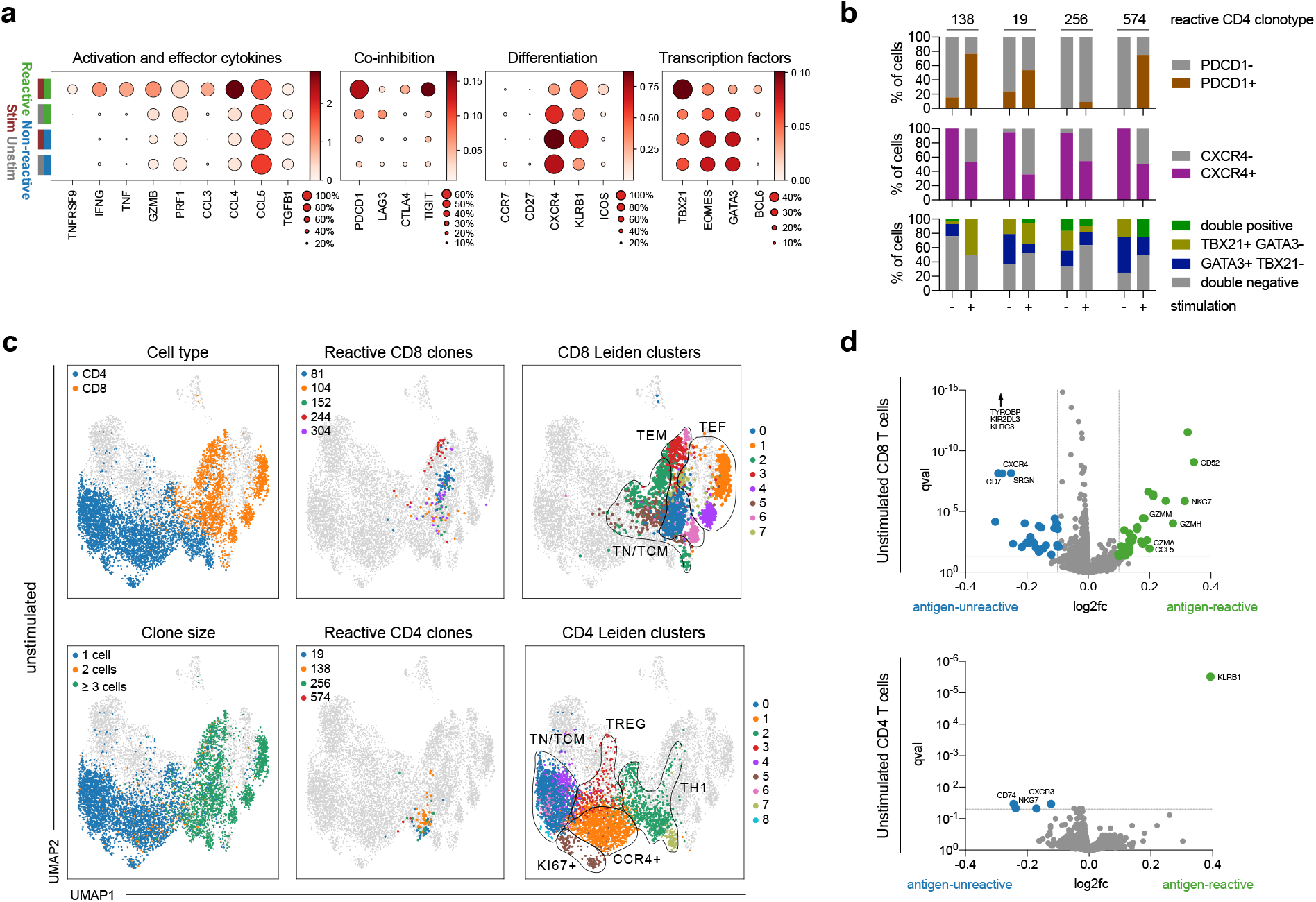
Reverse phenotyping reveals systematic biases induced by antigenic stimulation and allows precise definition of in vivo transcriptional profiles of SARS-CoV-2 antigen-reactive T cells. **a**, Dot plots of log-normalized expression of selected marker genes by clonotype group (reactive and non-reactive) and by condition (stimulated: ‘stim’, unstimulated: ‘unstim’). **b**, Fraction of gene-positive cells with >0 UMIs on the presented gene by clonotype and condition (+: stimulated, -: unstimulated). **c**, UMAPs indicating cells from unstimulated condition in colors with cells from stimulated condition shown in grey; CD4 or CD8 T cell state (top left panel), clonotype size as defined through cell number (bottom left panel); reactive CD8 clonotypes as defined in **a** (top middle panel) or reactive CD4 clonotypes as defined in **b** (bottom middle panel), with all other cells not belonging to that clonotype also shown in grey; CD8 (top right panel) and CD4 (bottom right panel) Leiden clusters, including manual annotation of cluster groups for the sake of clarity. **d**, Volcano plots of differential expression test of cells from non-reactive versus reactive clonotypes in unstimulated CD8 (top) and CD4 (bottom) T cells. For the sake of clarity, *TYROBP* (l2fc -0.48; qval 7×10^−131^), *KIR2DL3* (l2fc -0.15; qval 3×10^−60^) and *KLRC3* (l2fc -0.13; qval 3×10^−55^) are not displayed (CD8).

Indeed, many induced molecules are known to be differentially regulated after antigenic stimulation, but also serve as important determinants of more stably committed T cell phenotypes. High expression of *ICOS, PDCD1* and *TIGIT* has previously been described as a hallmark of SARS-CoV-2 specific CD4 T cells during severe disease^4^, but our findings indicate that the degree of expression can be significantly overestimated due to confounding stimulation effects. Conversely, expression of *CXCR4* or *KLRB1* would have been underestimated if cells had only been assessed after stimulation. CXCR4 is known to be more expressed in less differentiated T cells^32^, but downregulated upon T cell activation^33^. We next wondered whether and how *in vitro* stimulation with antigen also changed Th lineage-defining transcription factors, since these are particularly relevant for the assignment of CD4 T cell identity. Despite their robust *IFNG* upregulation upon antigenic stimulation, reactive CD4 T cells showed overall less Th1-defining *TBX21* (T-bet) expression *in vivo* than it would have seemed after stimulation (Fig. 3a). Vice versa, expression of *EOMES* and Th2-defining *GATA3* was downregulated in reactive CD4 cells after antigenic stimulation (Fig. 3a). Antigen stimulation also induced expression of (circulating) follicular helper T cell ((c)Tfh)-defining *BCL6* in a ‘false-positive’ manner in a few antigen-reactive CD4. Overall, we revealed that stimulation induced broad changes in transcriptional profiles, ranging from expected upregulation of activation markers to more unexpected and complex changes in markers associated with stable phenotypes.

The mentioned observations account for comparisons of stimulated vs. unstimulated and reactive vs. non-reactive clonotypes in bulk (Fig. 3a). However, individual clonotypes have also different phenotypes *a priori*, and could therefore show unique reactivity upon stimulation. To explore this further, we analyzed stimulation-induced transcriptional changes for individual reactive clonotypes. This revealed expected as well as unexpected, consistent as well as heterogenous phenotypic changes for reactive clonotypes. For example, while all reactive clonotypes upregulated *PDCD1* or downregulated *CXCR4* upon stimulation in a synchronized manner, stimulation induced clonotype-specific changes in CD4 lineage defining *TBX21* and *GATA3* (Fig. 3b). Clonotype 138 upregulated and clonotype 256 downregulated *TBX21* expression after stimulation. In addition to this inter-clonal heterogeneity, intra-clonal variability adds an additional layer of complexity (Fig. 3b), since individual clonotypes can show mixed phenotypes^34^. Indeed, all reactive CD4 clonotypes contain cells that express *TBX21* and/or *GATA3*. We could however not detect a statistical dependency between *TBX21* and *GATA3* expression (see methods section), suggesting that their expression is independent or that correlations could not be resolved.

Reactive CD8 T cells were overall less affected by antigenic stimulation than reactive CD4 T cells. After stimulation, CD8 T cells upregulated *IFNG* or *CCL4*, and also appeared more *LAG3* or *PDCD1* expressing than they were *in vivo* (Extended Data Fig. 6b). In previous studies, these latter molecules have been described to be strongly increased in SARS-CoV-2 specific CD8 T cells during severe disease^4^, providing further evidence that phenotypes of T cells after *in vitro* re-stimulation with antigen should be interpreted with caution.

To further characterize the stimulation-induced changes of antigen-reactive CD4 and CD8 clonotypes, we performed differential expression analysis (Extended Data Fig. 6c-d; Extended Data Tables 4-5). In CD4 T cells, this confirmed stimulation-induced upregulation of genes associated with induction of T cell activation^35–38^, or downregulation of genes known to be associated with repression of T cell activation^38–40^, but also revealed more unexpected differential gene expression of the Treg marker *FTH1*^41,42^, or of *TMSB4X* which is associated with effector-like Th1 cells^43,44^. In CD8 T cells, stimulation induced *GZMB* or *PRF1* most strongly, but was also associated with a prominent signature of actin (-binding) genes, which has previously described effector rather than exhausted T cells^35^. Of note, stimulation led to moderately increased detection of tissue residency signatures in reactive T cells (Extended Data Fig. 6e).

In summary, reverse phenotyping unraveled expected and unexpected phenotypic effects introduced by antigenic re-stimulation *in vitro*. The results reveal activation ‘artefacts’ and also highlight transcriptional differences between currently and previously activated T cells. Beneath the surface of global transcriptomic shifts on the population level, single-cell resolution together with TCR barcoding demonstrate additional inter- and intra-clonal heterogeneity.

### Unperturbed in vivo phenotypes of SARS-CoV-2-reactive T cells in peripheral blood

After comparing the differences between stimulated and unstimulated reactive clonotypes, we next focused on the unperturbed *in vivo* phenotypes of SARS-CoV-2-reactive T cells in further detail by comparing them with unreactive clonotypes in the unstimulated condition only. Reactive CD4 T cells selectively stemmed from a T cell effector memory (TEM) Th1-like group of cells, which was transcriptionally similar to CD8 T cells (Fig. 3c, Extended Data Fig. 7a-d). Two CD4 clusters (CD4 cluster 2 and 7) entailed Th1 cells. Both clusters were high in *IL7R* and *CXCR4*, but low in *CCR7* and *CD27*, and were the only clusters that expressed *CX3CR1*, speaking for a TEM-like phenotype. (Extended Data Fig. 7e-f). Remarkably, reactive CD4 clonotypes were exclusively found in CD4 cluster 2, but not in cluster 7. Cluster 7 consisted of cells belonging to a single clonotype (number 48, the largest clonotype found for CD4 T cells; Fig. 2b). The similar phenotype, but lacking reactivity of this clonotype could indicate that clonotype 48 recognizes an entirely different target, a different part of SARS-CoV-2 (non-spike), or it recognizes the same part, but is functionally exhausted and therefore non-reactive.

Reactive CD8 T cells were more evenly distributed across different CD8 TEM-like Leiden clusters (Fig. 3c, Extended Data Fig. 7). Of note, most CD8 T cells and the CD4 Th1 clusters showed clonal expansion, whereas few CD8 T cells and all non-Th1 CD4 clusters were clonotypically highly diverse (Fig. 3c), consistent with their overall little differentiated IL7R+ CCR7+ phenotype (Extended Data Fig. 7e).

To precisely define the *in vivo* phenotypes of SARS-CoV-2-reactive T cells in an unbiased manner, we performed differential gene expression analysis comparing reactive and non-reactive clonotypes in the unstimulated condition (Extended Data Tables 6-7). This identified *KLRB1* to be most significantly upregulated in reactive CD4 T cells (Fig. 3d). *KLRB1* encodes CD161 and is part of cytotoxic / Th 1 anti-viral T cells^45^. It has also already been described to be upregulated in SARS-CoV-2 reactive T cells after antigenic re-stimulation *in vitro*^46^. Together with IL7R expression, *KLRB1* also marks MAIT cells, but the TRAV/TRAJ expression of our antigen-reactive TCRs did not reflect invariant chains expressed by MAIT cells^31^.

Antigen-reactive CD8 T cells were characterized by high expression of granzymes, *CCL5* or the cytotoxic marker *NKG7*^18^, as well as *CD52*, target of Alemtuzumab, for which a paracrine suppressive function is described in activated CD4 T cells^47^ (Fig. 3d). Antigen-reactive CD8 T cells also were markedly lower in *TYROBP, KIR2DL3* and *KLRC3. TYROBP* encodes DAP12 and has known activating as well as inhibitory immune cell signaling roles when pairing with receptors belonging to the killer inhibitory receptors (KIR) or killer lectin like receptor (KLR) family^48^. In CD8 T cells, TYROBP and KLR transcripts have previously been shown to be downregulated after activation^49^.

Using these differentially expressed genes of reactive and non-reactive cells in the unstimulated condition, we defined transcriptional signature scores characteristic for antigen-reactive CD4 or CD8 T cells *in vivo*. These ‘*in vivo* signature scores’ overlapped to some extent with ‘stimulation-biased signature scores’ that we defined by using differentially expressed genes of reactive and non-reactive cells in the stimulated condition (Extended Data Tables 8-9), but also showed differences (Extended Data Fig. 8). For example, for CD8 cells, *GZMB* upregulation is only part of the ‘stimulation-biased signature score’, *GZMH* upregulation is part of both scores and *GZMA* and *GZMM* upregulation is only part of the ‘*in vivo* signature score’. Overall, we could precisely identify the phenotypes of currently and previously activated antigen-reactive PB T cells.

### Matching phenotypes of peripheral blood antigen-reactive T cells and the respiratory tract of COVID-19 patients

We next wondered whether we could identify T cells with our antigen-reactive transcriptional signatures in the respiratory tract. Tracheal aspirate (TA) T cells from identical as well as eight additional patients clustered in-between stimulated and unstimulated reactive clonotypes from peripheral blood (Extended Data Fig. 9a). TA CD8 T cells showed *IFNG* expression and high cytokine scores, whereas for CD4 T cells neither *IFNG* nor high proliferation scores were detectable (Extended Data Fig. 9b). These results were further corroborated by gene expression analyses of TA T cell clusters (Extended Data Fig. 10). Notably, TA CD4 T cells did not express *ZNF683*, encoding the Tissue resident memory (TRM) T cell-associated transcription factor Hobit, and expressed little *ITGA1*, encoding the TRM marker CD49a. TA CD8 T cells, in contrast, expressed *ZNF683* and ITGA1 throughout. While these TRM markers differentiated them from antigen-reactive PB CD8 T cells, TA CD8 T cells were overall very similar to reactive PB T cells – especially from the stimulated condition – with high expression of *IFNG, PDCD1* or *CD38* (Extended Data Fig. 10). These data indicated that particularly CD8 T cells from the respiratory tract of COVID-19 patients showed a phenotype that is similar to *in vitro* activated antigen-reactive CD8 T cells from peripheral blood.

To further test this hypothesis and investigate whether this was a feature that is specific for severe disease states, we integrated previously published and publicly available scRNA seq data sets from the respiratory tract of COVID-19 patients with severe disease, mild disease and healthy controls, from Berlin^26^, Shenzhen^27^ and Chicago^28^ in addition to our own Munich cohort (see methods for further information on data set integration). In total, we analyzed 279,663 cells from 50 patients (including 30,033 T cells from 28 patients; Extended Data Fig. 11a). Interestingly, stimulated antigen-reactive T cells from peripheral blood were most similar to T cells from nasopharyngeal swabs (NS), which were only present in the Berlin cohort^26^ (Extended Data Fig. 11b). However, in order to compare cell state similarities in a consistent manner and across as many patients and cohorts as possible, we excluded NS samples from further analyses, and only included samples derived from the lower airways (TA and bronchoalveolar lavage fluid, BALF). For some of the severely diseased patients, SARS-CoV-2 transcript was still detectable also in the scRNA seq data (Extended Data Fig. 12a), enabling further categorizations into ‘severe, virus-positive’ and ‘severe, virus-negative’ patients.

Stimulated CD4 or CD8 T cells from peripheral blood clustered in distinct niches that also contained respiratory T cells from severely diseased patients (Fig. 4a). To map this integrated data set in an unbiased and systematic manner, we applied Louvain clustering and then investigated cell type-dependent enrichments by calculating the share of each cell type within CD8 or CD4 T cells from peripheral blood or the respiratory tract across all Louvain clusters (Extended Data Fig. 12b-f). Louvain cluster 11 was enriched for CD8 and CD4 respiratory T cells from severely diseased patients and was at the same time the cluster that encompassed the most reactive peripheral blood clonotypes from the stimulation condition (Extended Data Fig. 12b-f). Leiden cluster 29 T cells (Fig. 1b) were also enriched in this area (Fig. 4a). We next analyzed the relative distribution of each cell type across Louvain clusters (Fig. 4b). Hierarchical clustering confirmed high proximity of respiratory CD8 T cell states to reactive peripheral blood CD4 and CD8 T cells from the stimulated condition, which were highly enriched in Louvain cluster 11. Intriguingly, PB CD8 reactive T cells clustered together with respiratory CD8 T cells from severely diseased patients particularly from Chicago and Shenzhen, which in contrast to the Berlin cohort encompassed a substantial number of virus-positive patients very early after entering the ICU. We therefore hypothesized that the phenotypic signatures of our stimulated or unperturbed reactive clonotypes could reflect, respectively, ‘hot’ (active virus replication) or ‘cold’ (virus cleared) respiratory tract environments of severely diseased patients, in which, respectively, virus was still detectable or not detectable anymore.

**Figure 4.**
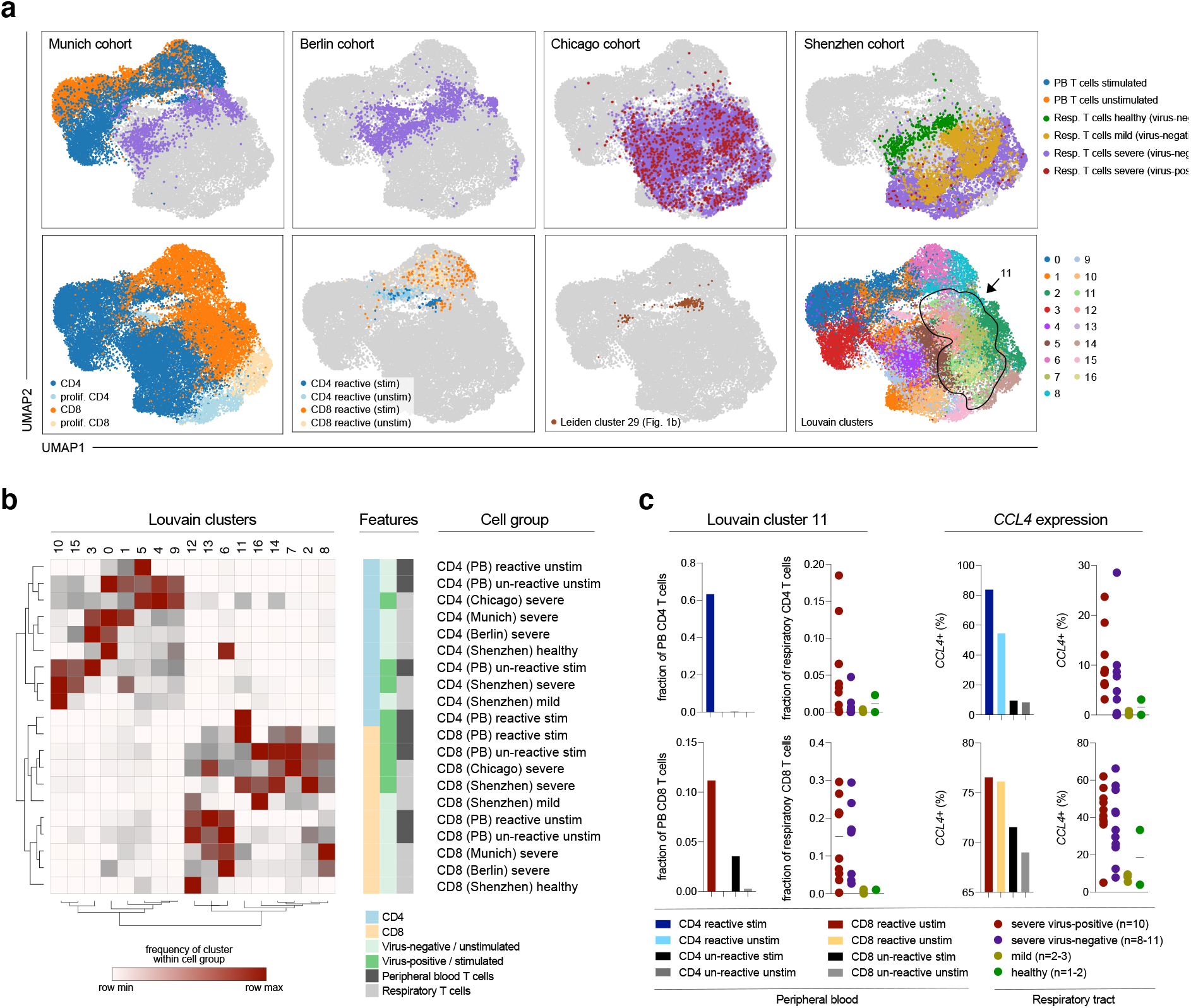
Phenotypic convergence of *in vitro* stimulated reactive T cells from peripheral blood and *in vivo* T cells from the respiratory tract of severely diseased patients. **a**, Top panels: UMAPs of previously analyzed peripheral blood (PB) T cells and T cells from tracheal aspirates (TA) of the same (GT_3) and additional patients with severe disease (Munich cohort) as well as from bronchoalveolar lavage fluid (BALF) from patients with mild disease, severe disease and healthy donors (Chua et al.^26^ - Berlin cohort; Grant et al.^28^ - Chicago cohort; Liao et al.^27^ - Shenzhen cohort); bottom panels (from left to right): CD4 or CD8 (proliferating or not) cell types, reactive PB T cells from stimulated or unstimulated conditions (see previous figures), IFNG-positive antigen-’reactive’ Leiden cluster 29 cells (see Fig. 1b), Louvain clusters. **b**, Hierarchical clustering of cell types and Louvain clusters based on ‘one minus Pearson’ correlations of deviations from baseline compositions for each cell type across Louvain clusters. PB, peripheral blood T cells. Cohort names (Munich, Chicago, Berlin, Shenzhen) refer to respiratory tract T cells. Fractions of CD4 or CD8 T cells in PBMCs or respiratory materials (Extended Data Fig. 12) served as input compositions. **c**, Fractions of Louvain cluster 11 or *CCL4* expressing cells among (from left to right and top to bottom) PB CD4 T cells, respiratory CD4 T cells, PB CD8 T cells or respiratory CD8 T cells. For respiratory T cells, data for individual patients with indicated disease stages are shown (represented by dots; n=2 healthy, n=3 mild, n=11 severe-cold, n=10 severe-hot; patients with less than < 20 cells in Louvain cluster 11 were excluded from analysis).

Based on the dominance of stimulated or unperturbed CD4 or CD8 clonotypes, we predicted Louvain clusters to be hot or cold CD4 or CD8 clusters, with cluster 11 being a generally ‘hot T’ cluster and for example the areas around cluster 5 and 13 reflecting ‘cold’ CD4 and CD8 environments, respectively (Fig. 4a-b). We then analyzed how many respiratory CD4 or CD8 T cells were present in those clusters. Unperturbed T cell signatures of reactive clonotypes were associated with ‘cold’ respiratory environments from severely diseased, but virus-negative patients. In contrast, stimulation-induced T cell signatures of reactive clonotypes were associated with ‘hot’ respiratory environments from severely diseased, virus-positive patients, as most prominently visible for Louvain cluster 11 (Fig. 4c). As an individual gene, *CCL4* was markedly upregulated in respiratory T cells from patients with severe disease, and even more so in virus-positive patients. This is reflected by higher expression in reactive T cells from peripheral blood, with particularly pronounced expression in the stimulated condition (see also Extended Data Fig. 8).

Overall, these data indicate that *ex vivo* stimulation induced a transcriptional state in antigen-reactive T cells from peripheral blood which is highly similar to T cells found in the respiratory tract from severely diseased patients with high viral loads (‘hot’ environments). In contrast, respiratory T cells from severely diseased patients in which no virus is detectable anymore (‘cold’ environments) showed a phenotype that is similar to peripheral blood antigen-reactive T cells when they are not additionally re-stimulated *ex vivo*. We could thereby interrogate connections between disease stages of individual patients and phenotypic signatures of respiratory T cells, for which currently and previously activated PB T cell subsets provided a framework.

### Modelling intercellular communication between respiratory T cells with antigen-reactive signatures and SARS-CoV-2-positive macrophages

We finally aimed to test how the definition of transcriptional signatures from antigen-reactive T cells after different antigen stimulation conditions can be leveraged across publicly available scRNA seq data sets. To this end, we investigated the intercellular communication between macrophages and respiratory T cells that bear our antigen-reactive signatures. We used scRNA seq data from the ‘Chicago cohort’^28^, for which a macrophage-T cell circuit has been described. The samples in this cohort of severely diseased patients were acquired 48 hours after intubation when virus was still detectable via scRNA seq. Macrophages in which SARS-CoV-2 transcript was detectable (either after direct infection or after phagocytosis of infected cells) seemed to sense IFNy produced by T cells and in turn released T cell-targeting cytokines^28^. These macrophages included tissue-resident alveolar macrophages (TRAM) as well as monocyte-derived alveolar macrophages (MoAM). To study the cross-talk between these macrophages and T cells with our antigen-reactive signatures systematically, we applied NicheNet (see methods). The NicheNet algorithm ranks ligands expressed by ‘sender’ cells according to their ability to induce a set of target genes in ‘receiver’ cells. Thereby, the algorithm exploits the transcriptomic signatures of receiver cells, building on prior knowledge on gene regulatory network from public databases beyond mere expression of matching ligands and receptors by sender and receiver cells, respectively^50^.

We first investigated which T cell ligands are predicted to induce the differentially expressed genes between SARS-CoV-2 transcript positive and negative macrophages. Compared to SARS-CoV-2-negative TRAM1, SARS-CoV-2-positive TRAM2 highly expressed cytokines such as *CCL2, CCL3, CCL4, CXCL9, CXCL10* and *CXCL11*, as well as molecules that have previously been described to play a role in chemotaxis (e.g. *ICAM1*) and responsiveness to IFNy (e.g. *STAT1*) in the interplay between monocytes and T cells during COVID-19^26^ (Extended Data Fig. 13). *IFNG* and *TNF* were predicted to be the most important ligands expressed by T cells that could induce the transcriptomic changes observed between SARS-CoV-2 transcript positive and negative TRAM. In T cells, the ligands IFNG and TNF were most dominantly expressed in Louvain cluster 11, which also highly expressed the predicted ligands *CCL3* and *CCL4* (Extended Data Fig. 13). The prediction pattern of additional top predicted ligands also coincided with the other hot or cold Louvain cluster regions (Fig. 4b). For example, hot CD8 Louvain clusters 2 and 7 (Fig. 4b) stood out through their high expression of the predicted ligand *CCL5* (Extended Data Fig. 13). Further, NicheNet predicted similar T cell ligands inducing the transcriptomic changes between SARS-CoV-2-positive and -negative MoAM (immature MoAM1 vs. mature MoAM2/3, respectively; Extended Data Fig. 14). In summary, unbiased prediction of T cell ligands inducing gene expression changes in SARS-CoV-2 transcript-positive macrophages identified Louvain clusters as the most dominant source of these ligands, which were initially grouped based on the presence of hot or cold T cell signatures and disease stages.

We next wondered whether SARS-CoV-2 transcript carrying macrophages would also signal back to T cells with our antigen-reactive signature in a specific manner. To reflect the ‘hot’ environment of the investigated patients in which virus was still present, we utilized NicheNet to predict which ligands expressed by Macrophages could induce the genes distinguishing reactive from unreactive T cells from the stimulated condition in CD4 and CD8 T cells. The definition of these target gene sets, including *GZMB, IFNG, TNF, CCL3* and *CCL4* in stimulated reactive CD4 T cells, led to the prediction of macrophage derived co-stimulatory ligands such as IL-15, IL-18, CCL4, CCL8 or CXCL9, which have been described to be upregulated in macrophages from the respiratory tract during COVID-19 also in the original studies on the Berlin and Shenzhen cohorts^26,27^ (Extended Data Fig. 15). For CD8 T cells as receivers, NicheNet predicted macrophage derived co-stimulatory ligands CCL2 and SPP1 (which encodes Osteopontin^51,52^) to drive *CCL3* and *CCL4* expression, as well as IL-15, IL-18, ICAM1, ADAM17, CD80 and CD86 to drive *IFNG, TNF* and *GZMB* expression (Extended Data Fig. 16). Intriguingly, most of the ligands that were predicted on the basis of target genes from our antigen-reactive CD4 and CD8 T cell signatures were preferentially expressed by the very macrophage subtypes in which SARS-CoV-2 transcript was detectable (TRAM2 and MoAM1). Overall, these data indicate a specific ligand-receptor cross-talk between respiratory T cells with antigen-reactive signatures and SARS-CoV-2-positive macrophages.

## Discussion

We here show how barcoding through natural TCR sequences can be used to identify SARS-CoV-2 antigen-reactive T cells after differential stimulation with and without antigen, followed by scRNA seq. Antigen-reactive clonotypes showed consistent transcriptomic shifts upon stimulation, as visible by induction of a distinct cluster in a UMAP projection, expression of generic T cell activation scores along continuous pseudotemporal gradients, and high degrees of inter-correlative gene expression changes. As a single marker, *IFNG* upregulation proved to reliably identify SARS-CoV-2 antigen-reactive CD4 and CD8 T cells. We validated reactive TCRs by transgenic re-expression using CRISPR/Cas9-mediated OTR. Such SARS-CoV-2 reactive TCR-transgenic T cells serve as a potential resource for future immunotherapy of COVID-19 infection. Furthermore, the presented approach represents a generic platform for identification of antigen-reactive TCRs for a plethora of *a priori* phenotypically heterogeneous clonotypes at once.

In addition to the identification of antigen-reactive clonotypes *per se*, ‘reverse phenotyping’ allowed us to investigate the phenotypic profile of antigen-reactive T cells with or without antigenic re-stimulation. This revealed systematic biases induced through re-stimulation and allowed exploration of unperturbed T cell states. By profiling antigen stimulated CD4 T cells from peripheral blood via scRNA seq, it has recently been shown that SARS-CoV-2 antigen-reactive CD4 T cells – particularly in patients with severe disease – show a particularly pronounced cytotoxic profile^9,13,15^ compared to CD4 T cells specific for other viruses^14^. We confirm these findings by demonstrating that CD4 T cells which show major transcriptional shifts upon antigenic stimulation robustly upregulate *IFNG*. However, our ‘reverse phenotyping’ approach also reveals that the classical Th1-ness (as defined by *TBX21* expression) of those cells is dramatically overestimated after stimulation, and that – *in vivo –* antigen-reactive PB T cells instead show signatures that are far more dominated by *EOMES* and *GATA3* than it appears after stimulation. The role of EOMES in CD4 T cells is less clear compared to other Th lineage defining core transcription factors, but recent evidence suggests that EOMES drives a cytotoxic signature and represses Th17 features^53,54^. Apart from Th1-polarized CD4 cells, antigen-reactive cTfh cells have been reported by several studies^4^. In our patient subset with a severe disease course, we observed *BCL6* and *ICOS* expression in antigen-reactive CD4 clonotypes upon stimulation only. While this does not exclude the possibility that cTfh play a dominant role in the adaptive immune response to SARS-CoV-2, these findings provide examples of the phenotypic biases introduced by antigen stimulation prior to phenotyping.

Both CD4 and CD8 antigen-reactive T cells showed TEM/T cell effector (TEF) phenotypes with less expression of co-inhibitory molecules and markers of terminal differentiation than would be expected based on the phenotypes after stimulation. However, at the same time, antigen-reactive CD4 and CD8 T cells were characterized by a more cytotoxic and differentiated profile in comparison to antigen-unreactive T cells when stimulation biases are accounted for. Thereby, reverse phenotyping allowed us to define more accurately the actual *in vivo* phenotypic profiles of SARS-CoV-2 antigen-reactive T cells. We performed reverse phenotyping on T cells only from peripheral blood, in a limited number of patients with severe disease, and focused on T cell reactivity to virus spike antigens. More comprehensive experimental analyses of SARS-CoV-2 antigen-reactive T cells are needed – including different time points, disease severities, and specific antigen groups – to more comprehensively map the *in vivo* phenotypic profile of SARS-CoV-2 specific T cells. Here, we provide proof-of-concept that ‘reverse phenotyping’ can be used for that purpose.

Publicly available data sets – particularly those that are bundled in a coherent manner as within the Human Cell Atlas initiative – represent a vast resource of deeply profiled immune cells from many different patients and disease contexts. However, in these data sets often no dedicated antigen-specific T cells analyses have been performed. In T cells from the respiratory system of identical as well as further patients – including patients from independent reference cohorts^26–28^ – we detected transcriptional signatures that overlapped with those we defined based on antigen-reactive T cells from peripheral blood. *In vitro* stimulated reactive T cells thereby mirrored ‘hot’ cell states from virus-positive patients with severe disease, whereas unperturbed T cell states reflected ‘cold’ cell states from virus-negative patients with severe and mild disease. Interaction of T cells with viral antigen at the site of infection could explain these findings. In the future, it will be exciting to further investigate the role of SARS-CoV-2 antigen-reactive T cells at the site of infection as well as in peripheral blood, e.g. by using TCR sequences to precisely define ontogenic relationships.

As a tool for T cell immunology, scRNA seq after differential antigen stimulation opens up new avenues for identification and characterization of antigen-reactive T cells by exposing gradients of antigen reactivity and reverse phenotyping. It thereby contributes to a deep understanding of the adaptive immunity induced by viral infection, which will be pivotal to guide enhanced and accelerated development of therapies and vaccines for emerging pathogens such as SARS-CoV-2.

## MATERIALS & METHODS

### Munich cohort patients

#### Clinical information and material

All Munich cohort patients were PCR-confirmed SARS-CoV-2 positive, admitted to the intensive care unit in the University Hospital of the Ludwig-Maximillians University, Munich (n=5), or the Asklepios Lung Clinic Munich-Gauting, Gauting (n=4), for treatment of severe COVID-19 requiring invasive, mechanical ventilation. For further clinical information see Extended Data Table 1. PBMCs and tracheal aspirate samples were taken at the end of April, 2020.

#### Consent

Written informed consent was obtained from the donors or their caregivers, usage of the blood samples was approved according to national law by the local Institutional Review Board (Ethikkommission der Medizinischen Fakultät der Ludwigs-Maximilian-Universität München; vote IDs 19-629, 19-630 and 20-259) and/or samples were used according to legal provisions defined by the German Infection Protection Act (IfSG).

### T cells from PBMCs

#### Cell culture

Peripheral blood mononuclear cells (PBMC) were isolated from EDTA whole blood by gradient density centrifugation according to manufacturer’s instructions (Biocoll, Biochrom) and frozen in FCS + 10% DMSO (Merck) for liquid nitrogen storage. After freezing-thawing procedure, T cells were cultured in RPMI 1640 (Gibco) supplemented with 5% human serum, 0.025% l-glutamine, 0.1% HEPES, 0.001% gentamycin and 0.002% streptomycin (hereafter RPMI-HS).

#### Antigen-specific stimulation and flow cytometry-assisted cell sorting prior to scRNA seq

PBMCs were stimulated with 0.6nmol of SARS-CoV-2 spike protein peptide mix (PepTivator®SARS-CoV-2 Prot_S, Miltenyi). For the unstimulated condition, PBMCs were only cultured in RPMI-HS. After stimulation for 4h at 37°C, surface staining was conducted for 20min with the following fluorochrome conjugated antibodies: CD3-APC, CD4-PE, CD56-FITC (Life Technologies), CD8-eFluor450 (Biolegend) and CD19-ECD (Beckman Coulter). Propidium iodide (Invitrogen) was used for live/dead discrimination. Flow sorting of CD4+ and CD8+ cells from the stimulated and unstimulated condition was conducted on a MoFlo Astrios EQ (Beckman Coulter) under biosafety level 3.

#### TCR DNA template design and CRISPR-Cas9 mediated TCR KI

DNA constructs for CRISPR-Cas-9-mediated HDR at TRAC locus were designed *in silico* with the following structure: 5′ homology arm (300–400 base pairs (bp)), P2A, TCR-β (including mTRBC with additional cysteine bridge^55^), T2A, TCR-α (including mTRAC with additional cysteine bridge), bGHpA tail, 3′ homology arm (300–400 bp). All HDR DNA template sequences were synthesized by Twist. CRISPR-Cas9-mediated TCR KO and KI was performed as previously described^25^ on isolated PBMCs from donor whole blood. Sequences of experimentally tested TCRs can be found in Extended Data Table 10.

#### Antigen-specific activation of TCR-engineered T cells and intracellular cytokine staining

PBMCs for autologous peptide pulsing were isolated and cultured in RPMI-HS with 50 IU/ml human IL-2 (Peprotech). On the day of antigen-specific activation of CRSIPR-Cas9-engineered T cells, autologous PBMCs were pulsed with 10 µg/ml SARS-CoV-2 spike protein peptide mix (PepTivator®SARS-CoV-2 Prot_S, Miltenyi) for 2h at RT and gentle agitation. After peptide pulsing, excess peptide was removed by washing and PBMCs were co-cultured with CRSIPR-Cas9-engineered T cells in 1:1 and 1:3 effector:target ratio and Golgi-Plug (BD Biosciences) for 4h at 37°C. Surface marker antibody staining for CD3-BV421 (BD Biosciences), CD8-PE (Invitrogen) and murine TCR β-chain-APC/Fire750 (Biolegend) was followed by permeabilization using Cytofix/Cytoperm (BD Biosciences) and intracellular staining of IL-2-APC (BD Pharmingen). Live/dead discrimination was performed by using ethidium-monoazide-bromide (Invitrogen). FACS samples were acquired on a Cytoflex (S) flow cytometer (Beckman Coulter).

### Single-cell RNA sequencing

#### Peripheral blood T cell processing

CD3+ CD4+ and CD8^+^ T cells were sorted by flow cytometry, centrifuged and the supernatant was carefully removed. Cells were resuspended in the Mastermix + 37.8 µl of water before 70 µl of the cell suspension were transferred to the chip (step 1.1 and 1.2 of the original protocol). After each step, the integrity of the pellet was checked under the microscope to ensure that all cells are loaded onto the chip. From here on, 10x experiments have been performed according to the manufacturer’s protocol (Chromium next GEM Single Cell VDJ V1.1, Rev D). Quality control has been performed with a high sensitivity DNA Kit (Agilent #5067-4626) on a Bioanalyzer 2100 as recommended in the protocol and libraries were quantified with the Qubit dsDNA hs assay kit (life technologies #Q32851). All steps have been performed using RPT filter tips (Starlab #S1181-3710) and DNA LoBind tubes (Sigma #EP0030108051, #EP0030108078, #EP0030124359).

#### Tracheal aspirate processing

Tracheal aspirates were digested with 4ml dispase (50 units/ml) (Corning, #354235) and 25 µl DNase (30 μg/ml) (Qiagen, #79254) at 37°C for 10 min with occasional shaking. The digestion was then stopped with 10 ml of ice-cold 10% FCS/PBS. To obtain single cell suspensions, the digestion mix was passed through a 70 µm cell strainer. Red blood cell lysis was performed only when necessary by incubating the cells with 3 ml RBL buffer at RT for 1 min. The cells were counted, diluted to 1,000 cells/µl and loaded on the 10x Chromium Next GEM Chip G with a targeted cell recovery of 10,000. The following steps were completed according to manufacturer’s protocol (Chromium Next GEM Single Cell 3’ Reagent Kits v3.1).

#### Next Generation Sequencing

Libraries have been pooled according to their minimal required read counts (35,000 or 50,000 reads/cell for 3’ gene expression libraries, 20,000 reads/cell for 5’ gene expression libraries and 5,000 reads/cell for TCR libraries). Illumina paired end sequencing was performed with 150 or 200 (3’ gene expression) and 100 cycles (5’ gene expression and TCR libraries) on a NovaSeq 6000.

### Single-cell RNA sequencing data analysis

#### Data processing

After sequencing, the processing of next generation sequencing reads of the scRNA-seq data was performed using CellRanger version 3.1.0 (10X Genomics) with a customized reference genome. To account for transcripts originating from the virus, the genes of the SARS-CoV-2 genome (NCBI Reference Sequence: NC_045512.2) were manually added to the hg38 (GRCh38.99) human reference genome’s GTF files. Negative strands of the viral genes were additionally added. The genome was indexed via CellRanger’s “mkref” command.

#### Initial unsupervised analysis of PB T cells

We performed unsupervised analysis for multiple subsets of the data with scanpy^56^ (v1.6.0): Both patients, patient GT_3 only, patient GT_2 only, stimulated or unstimulated condition only for both patient GT_3 and GT_2, separately. In all cases, we first filtered genes that were expressed in at least 10 cells, scaled cell-wise expression vectors to a total count of 10,000, and logp1-transformed the data and selected highly variable genes (flavor=“seurat). We then performed a principal component analysis, computing 50 components, based on which we computed a k-nearest neighbor graph (k=50). We then computed UMAPs^57^ and Leiden or Louvain clusterings^58^. The complete code for this analysis is reported in Extended Data File 1.

#### Cell type assignment of PB T cells

CD4 and CD8 expressing cells did not clearly separate in Leiden groups. Therefore, we chose a two-staged cell type assignment process: First, we assigned Leiden groups to either CD4+ or CD8+ T cells based on their relative mean expression in the group. Secondly, we re-assigned cells from clonotypes (see also: Clonotype analysis), which contained both putative CD4+ and CD8+ T cells, to the major cell type found in this clonotype.

#### Cell state assignment of PB T cells

We re-clustered CD4 and CD8 cells in the unstimulated condition separately using the Leiden algorithm to identify cellular states at higher resolution than cell types. For this purpose, we repeated the unsupervised analysis as described above on this subset of cells. Throughout the manuscript, these clusters are ordered by similarity in heatmaps and dot plots. This ordering is derived from a hierarchical clustering based on all genes with mean expression larger than 0.5 in the selected subset.

#### Clonotype assignment of PB T cells

We performed TCR sequence analysis with scirpy^59^ (v0.4). We defined clonotypes based as groups of cells with identical nucleotide CDR3 sequences of the primary TRA and TRB chains. We performed clonotype assignment for both patients separately, but across both conditions, stimulated and unstimulated, together. Clonotype sequences can be found in Extended Data Tables 11 and 12. The complete code for this analysis can is reported in Extended Data File 1.

#### Pseudotime analysis of PB T cells

The complete code for this analysis is reported in Extended Data File 1. We computed pseudotime-based scores by computing diffusion pseudotime^60^ with respect to a cell in the reactive T cell cluster in patient GT_3. This root T cell was selected based on its extremal position in the UMAP, at the tip of the reactive cluster. The nearest-neighbor graph underlying the pseudotime computation was derived as described in section initial unsupervised analysis.

#### GATA3 and TBX21 correlation

The complete code for this analysis is reported in Extended Data File 1. We investigated *GATA3* and *TBX21* correlation in the CD4 cells with two techniques: Firstly, we computed their log-normalized expression covariance. Secondly, we computed expected rates of *GATA3*-*TBX21*-, *GATA3*+*TBX21*-, *GATA3*-*TBX21*+ and *GATA3*+*TBX21*+ cells based on the marginal frequency of *GATA3*+ and *TBX21*+ cells and compared these to the observed frequencies of single- and double-positives. Here, positive cells were defined as cells with any detected UMIs of the selected gene.

#### Differential expression analysis of PB T cells

We performed differential expression analysis with diffxpy (v.0.7.4): We used Welch’s t-test on log-normalized UMIs to compare expression differences between two groups of cells. We labeled genes as differentially expressed if they had a Benjamini-Hochberg corrected p-value of less than 0.01 and a mean expression of at least 0.5.

#### Tissue residency score of PB T cells

We computed expression z-scores for the following genes associated with tissue residency^61^: *CA10, CRTAM, CX3CR1, CXCL13, CXCR6, DUSP6, FAM65B, IL2, IL10, IL23R, ITGA1, ITGAE, KCNK5, KLF2, KLF3, KRT72, KRT73, NPDC1, PDCD1, PTGDS, RAP1GAP2, RGS1, S1PR1, SBK1, SELL, SOX13, STK38, TSPAN18, TTC16, TTYH2*. We aggregated these cell- and gene-wise z-scores to a cell-wise tissue residency score by taking the mean across all gene-wise z-scores.

#### Unsupervised analysis of TA data

The count matrices output by CellRanger were again analyzed using Scanpy^56^ (v.1.6.0). For barcode filtering, we excluded barcodes with less than 200 detected genes. A high proportion (> 10%) of transcript counts derived from mitochondria-encoded genes may indicate low cell quality, and we removed these unqualified cells from downstream analysis. The number of unique molecular identifiers (UMIs) was explored via patient wise violin plots, and individual upper cut-offs were applied ranging from 5,000 to 80,000 UMI counts. Genes were only considered if they were expressed in at least 3 cells in the data set. As we observed a certain degree of ambient RNA bias, we applied SoupX version 1.3.6^62^ to lessen this effect with mostly default parameters, setting the contamination fraction manually to 0.3.

The expression matrix was normalized using scran’s normalization approach^63^, in which size factors are calculated and used to scale the counts in each cell. The data were then log1p-transformed and the top 4,000 variable gene were selected for each patient individually (flavor=“seurat). Genes that were listed as variable in at least 3 patients (5,984 genes after removing cell cycle genes) were used as basis for the principal component analysis. BBKNN^64^ was employed to construct a batch-corrected neighborhood graph using the first 20 components and individual patient label as batch key. Unsupervised clustering was performed with Louvain at resolution 3 and cells coarsely annotated based on manual exploration of known marker genes. As T cell subtypes were of particular interest, we subdivided the data set to *CD3E* high cluster and refined the annotation after re-clustering and assessment of mean expression of *CD8A, CD8B* and *CD4* in the respective cluster.

### Computational data integration

#### Computational data integration - Munich TA and PBMC

In order to compare the results from the PBMC data set with *in vivo* patient samples, the normalized, log1p-transformed matrices of both objects after initial filtering were concatenated. The variable gene selection was executed anew with more stringent parameters on this combined object. The top 800 genes (flavor= “cell_ranger”) were selected for both data sets separately, resulting in 183 variable genes labelled as such in both calculations. PCA and the neigbourhood graph was re-calculated using 50 components and 15 neighbors. The cell type annotation established on the separate objects was retained.

#### Computational data integration – include recent Covid-19 related BALF data sets

To test the generalizability of our results on independent cohorts and in order to include varying levels of severity and healthy controls in our analysis, we combined our single cell data (Munich cohort) with recently published Covid-19 related data sets. The raw count matrices and meta data from the Shenzhen^27^, Chicago^28^, and Berlin^26^ cohorts were downloaded and pre-processed separately with Scanpy (v1.6.0). Briefly, for each of the data sets cells that were present in the final provided object were retained. Additionally, we re-calculated the number of counts and genes per cell and applied following thresholds: only cells with more than 200 genes and less than 15% mitochondrial transcripts were kept. We assessed the number of UMIs with sample-wise violin plots as the number of detected transcripts varied across the cohorts. Following upper thresholds were chosen: 6,000 for the Shenzhen cohort, 30,000 for the Chicago cohort, and 200,000 for the Berlin cohort.

The expression matrices of each data set were normalized and log1p-transformed separately as described above for the Munich Tracheal Aspirates. For a first lighter batch correction we defined the list of variable genes in a way that decreased cohort-specific effect as follows. First, highly variable genes were selected (flavor=“cell_ranger”) for each sample separately, returning the top 4,000 variable genes per individual. For the Munich, Shenzhen, and Chicago cohorts, we considered a gene as highly variable if it is labelled as such in at least 3 patients of the respective cohort. As the Berlin cohort contained far more patients, we increased the threshold to 6 patients.

After quality control the ambient RNA contamination was assessed and removed using SoupX^62^ with mostly default parameters, setting the contamination fraction manually to 0.3.

Next, the preprocessed count matrices from the individual data sets were merged and the highly variable genes were set to the intersection of the four cohort-wise lists. After excluding genes associated to the cell cycle, 1,370 variable genes remained, which were used as input to PCA (n=50).

The published cell type assignments from each cohort were used further, the labels were harmonized as some annotations for the same cell type showed slight changes in spelling. Cells with unclear annotation were removed from the analysis (“Doublets”, “hybrid”, “unknown_epithelial”, “IRC”).

For visualization of the concatenated data sets, a UMAP and a batch-corrected neighborhood graph was constructed via BBKNN^64^ using 10 neighbors within each batch with *cohort* used as batch key.

### Computational data integration – T cell subtypes

The cells with the following annotations were used to subset to a T cell only data set: “CD8 T, CCR7+ T, Proliferating T, Treg” (Shenzhen cohort^27^), “CD4 T cells, CD8 T cells, Proliferating CD4 T cells, Proliferating CD8 T cells, Tregs” (Chicago cohort^28^) and “CTL, Treg” (Berlin cohort^26^). For the Berlin cohort, only T cells from BALF samples were used further, cells originating from nasal swabs are only shown in the data set encompassing all cell types. The cell type annotation for the Shenzhen and Berlin cohort did not distinguish between CD4 and CD8 T cells, and was therefore refined in an additional step. After re-calculating of the principal components and Louvain clustering, manual examination of CD8A, CD8B, CD4A, CD3E expression enabled a more fine-grained annotation of the subtypes comparable with the Munich cohort annotation. Using the variable gene list established on the full integrated data set, the PCA was re-calculated and the batch corrected neighborhood graph re-constructed (n_pcs = 50, neighbors_within_batch = 20, batch_key = “data_set”).

### Hierarchical clustering of cell type frequencies across Louvain clusters

Hierarchical clustering of cell type frequencies across Louvain clusters was performed with Morpheus (https://software.broadinstitute.org/morpheus). Both Louvain clusters and cell types were clustered based on ‘one minus Pearson’ correlations.

### NicheNet analysis

We performed NicheNet analysis on the scRNA seq reference cohort from Chicago using the R (version 3.6.3) packages nichenetr^50^ (version 1.0) and Seurat^65^ (version 3.2): To analyze cell-cell communication between Macrophages and CD4 or CD8 T cells respectively, we adopted the cell type annotations from the Chicago dataset and defined the five distinct Macrophage subpopulations contained therein (MoAM1, MoAM2, MoAM3, TRAM1 and TRAM2) as sender cell types and all CD4 or CD8 T cells as receiver cell types, respectively. We then defined the genes comprising the different ‘in vivo signature scores’ for CD4 and CD8 T cells as gene set of interest for CD4 and CD8 T cells, respectively.

To analyze cell-cell communication between individual T cell subsets and Tissue-resident Macrophages (TRAM) or Monocyte-derived Alveolar Macrophages (MoAM) respectively, we used the resulting 17 Louvain clusters from the integrated analysis as sender cell types and all TRAMs or MoAMs (see original annotation from the Chicago reference cohort) as receiver cell types. To define a gene set of interest, we performed differential expression testing at the gene level between TRAM1 vs TRAM2 and immature MoAMs (MoAM1) vs mature MoAMs (MoAM2 and MoAM3) using a Wilcoxon Rank Sum test with Seurat’s FindMarkers() function. We only included genes with a log-fold change of >0.25, if they were expressed in > 10% of all cells, and if their Holm-Bonferroni adjusted p-value was < 0.05.

For all NicheNet analyses we identified a list of potentially active ligands expressed by sender cell types and ranked them according to their ability (Pearson’s correlation coefficient) to predict the gene set of interest. We then selected the 12 top-ranked ones for subsequent analysis. The complete code for this analysis is reported in Extended Data File 1.

## Data Availability

All data generated or analyzed during this study are included in this article and its supplementary information files. Additional raw data are available from the corresponding authors upon reasonable request.

## EXTENDED DATA

**Extended Data Figure 1.**
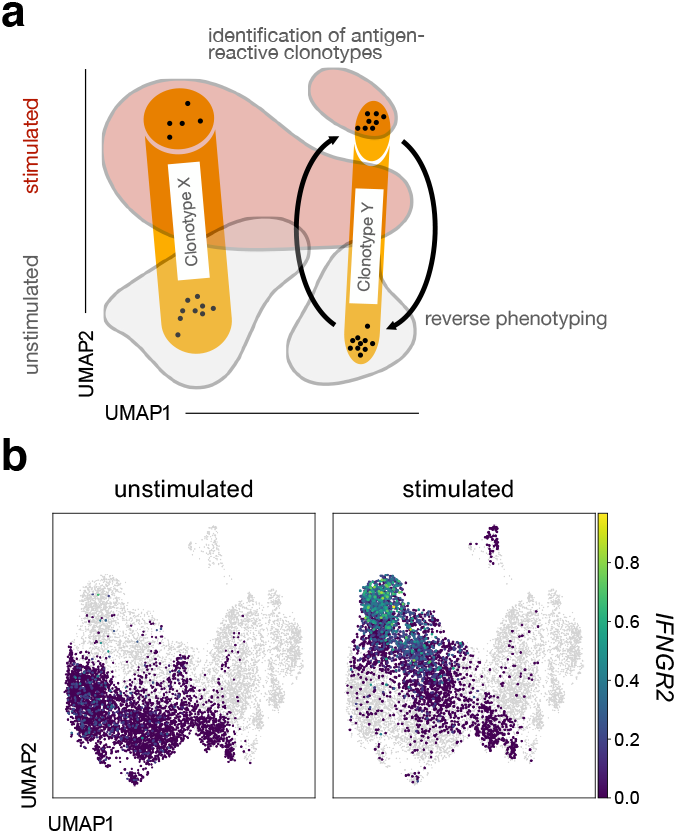
Scheme of experimental approach and *IFGNR2* expression in CD4 T cells. **a**, Scheme of experimental approach to use scRNA seq of differentially stimulated cells and TCRs as natural barcodes for cells belonging to shared clonotypes for identification of antigen-reactive clonotypes and reverse phenotyping. **b**, UMAP of all T cells, with CD4 T cells from the stimulated and unstimulated condition, highlighted separately with log-normalized *IFGNR2* expression superimposed. Data are shown for patient GT_3.

**Extended Data Figure 2.**
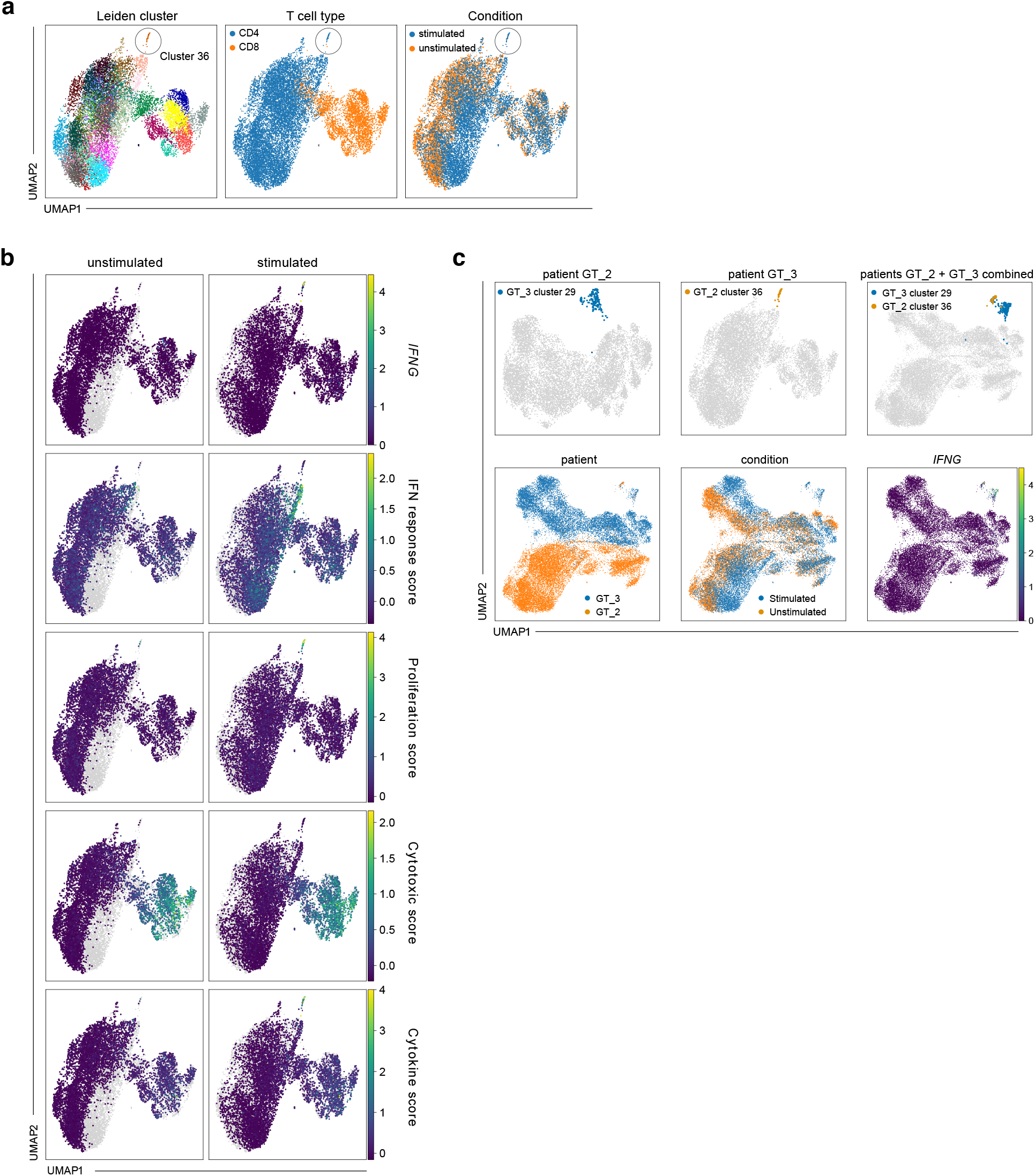
Unsupervised analysis of PBMC of patient GT_2. **a, b**, UMAPs of all T cells of patient GT_2 with Leiden cluster, cell type and condition superimposed (**a**) and log-normalized *IFNG* expression and selected scores superimposed (**b**). **c**, Individual UMAPs from patient GT_3, GT_2 as well as integrated UMAP from both patients with stimulation-reactive Leiden clusters, patient ID, condition and *IFNG* expression highlighted.

**Extended Data Figure 3.**
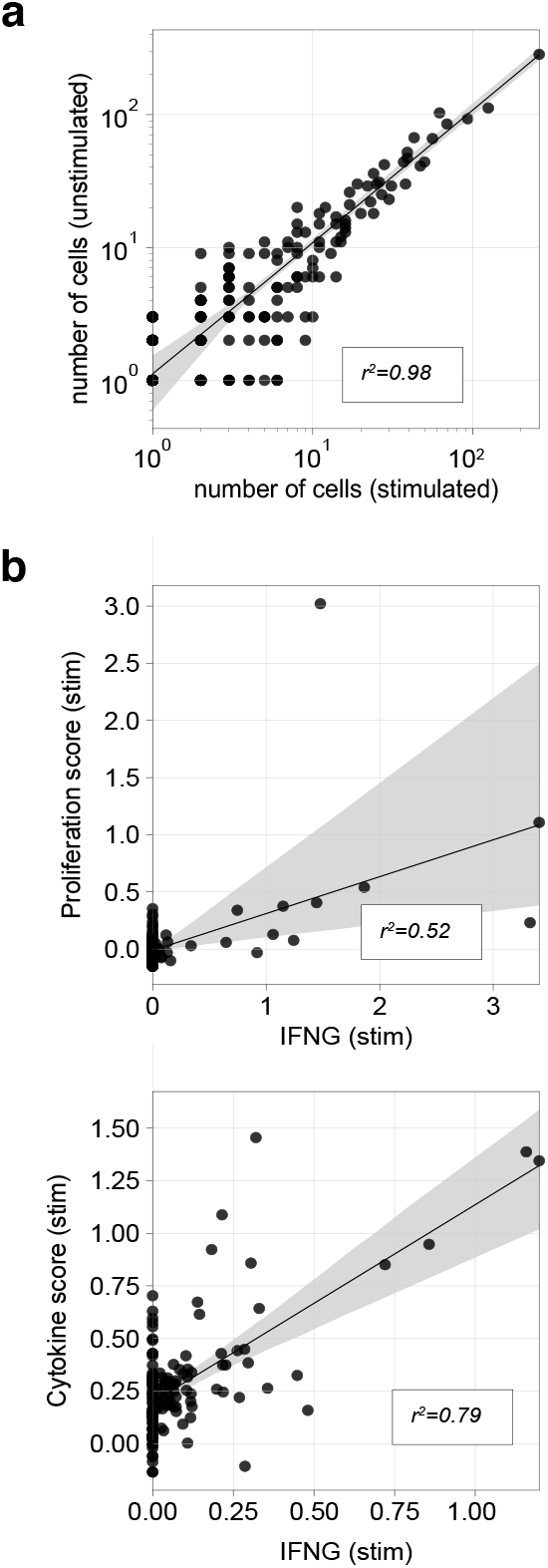
Clonotype summary statistics. **a**, Scatter plot showing number of cells of clonotype in stimulated condition versus number of cells in the unstimulated condition. **b**, Mean log-normalized *IFNG* expression versus mean proliferation score in CD4 T cells (top) or versus mean cytokine score in CD8 T cells (bottom) in stimulated condition by clonotype. Data are shown for patient GT_3.

**Extended Data Figure 4.**
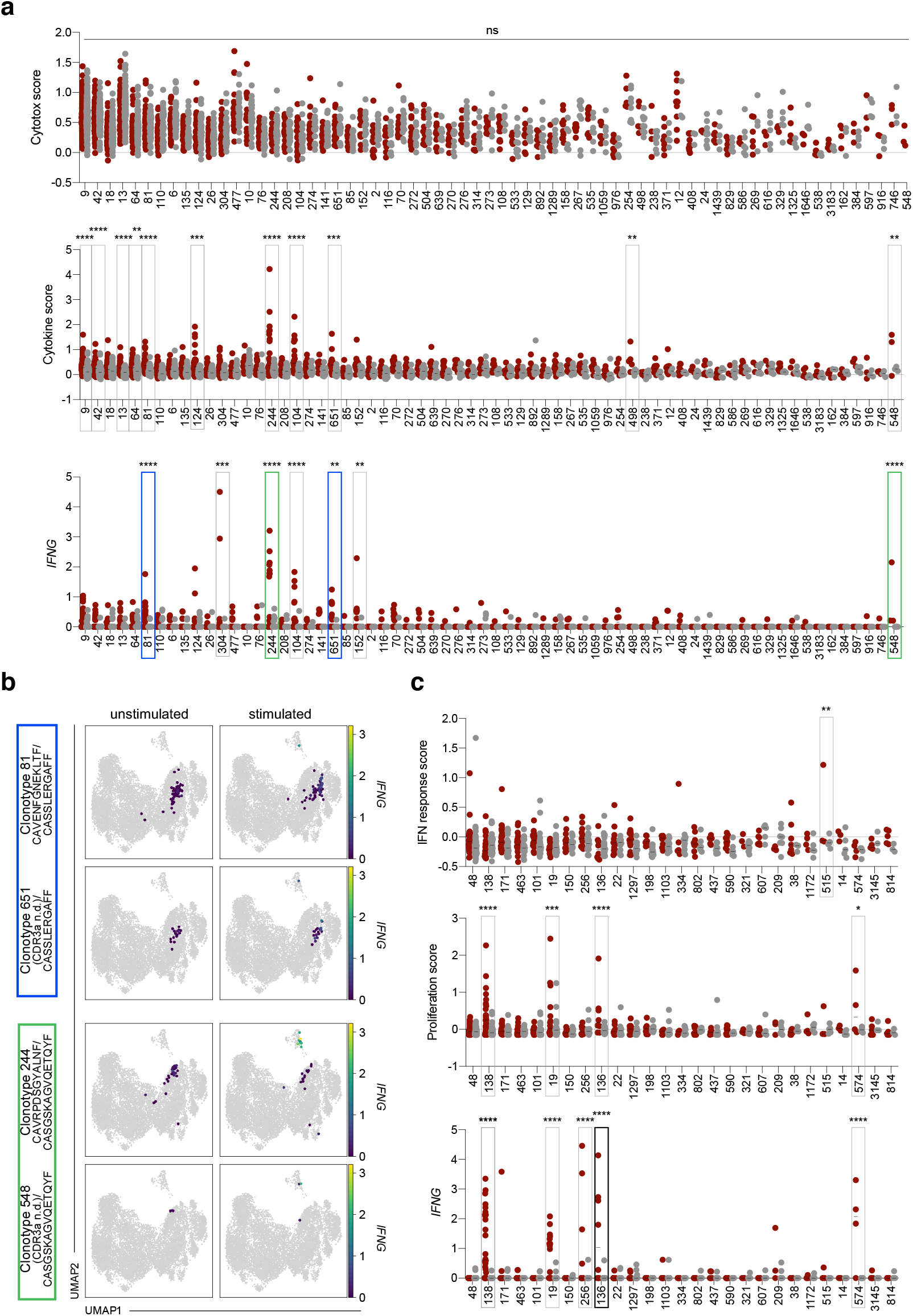
Clonotype-wise activation statistics. **a**, Cell-wise cytotoxic score, cytokine score, and log-normalized *IFNG* expression by clonotype in CD8 T cells. **b**, UMAPs of all T cells with cells from selected CD8 clonotypes highlighted with log-normalized *IFNG* expression superimposed. **c**, Cell-wise IFN response score, proliferation score, and log-normalized *IFNG* expression by clonotype in CD4 T cells. In contrast to Fig. 2a-b, additionally clonotypes that were defined through a single TCR chain were incorporated into the analyses; as in Fig. 2a-b, only clonotypes with at least three cells in each condition were considered (a-c); reactive single-chain clonotypes are highlighted in blue/green (a) and black (c); reactive partner clonotypes based on shared CDR3a sequences are shown in corresponding colors (a). Data are shown for patient GT_3.

**Extended Data Figure 5.**
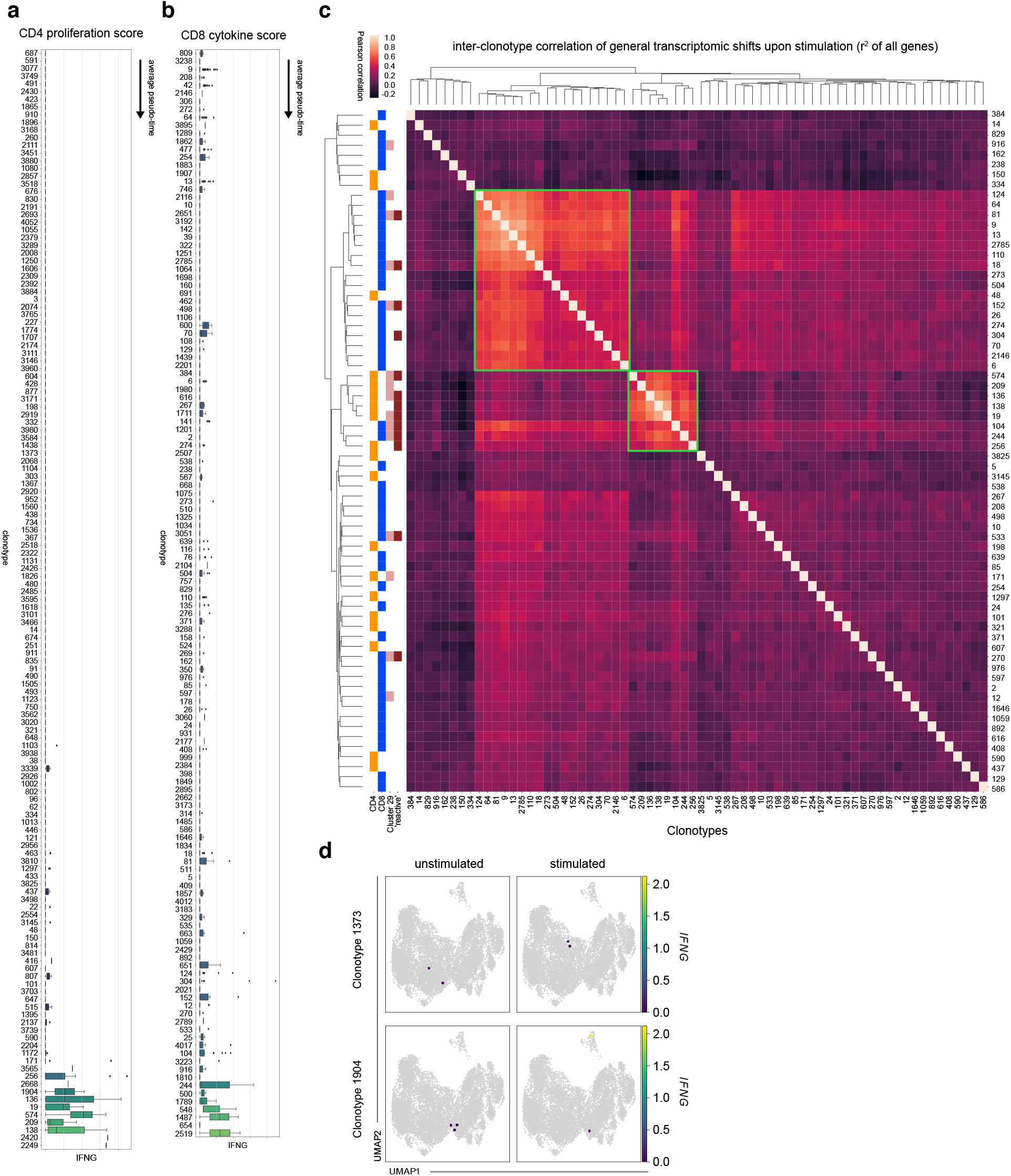
Pseudotemporal interpolation of clonotype activation and stimulation-induced shifts for two small clonotypes. **a, b**, Distribution of CD4 proliferation score (**a**) and CD8 cytokine score (**b**) of clonotypes ordered by mean pseudotime. Considered were clonotypes with at least one cell in each condition. The center of each boxplot is the sample median; the whiskers extend from the upper (lower) hinge to the largest (smallest) data point no further than 1.5 times the interquartile range from the upper (lower) hinge. Data are shown for patient GT_3. **c**, Pearson correlation of stimulation vectors per clonotype. On the left, clonotypes are assigned to CD4+ T cells (CD4), CD8+ T cells (CD8), to having cells in the IFNG+ reactive cluster 29 (Fig. 1b) and as to whether the clonotype was identified as ‘reactive’ (Fig. 2a-b). Stimulation vectors reflect the difference between mean log-normalized counts of the 2000 highest expressed genes between stimulated and unstimulated condition for each clonotype that had at least three cells in each condition. Clonotypes were clustered using hierarchical clustering. **d**, *IFNG* expression in unstimulated or stimulated T cells for indicated clonotypes. For each clonotype, cells belonging to that clonotype are shown in an individual panel pair (cells from unstimulated condition in left panels, cells from stimulated condition in right panels), while cells not belonging to that clonotype are shown in grey.

**Extended Data Figure 6.**
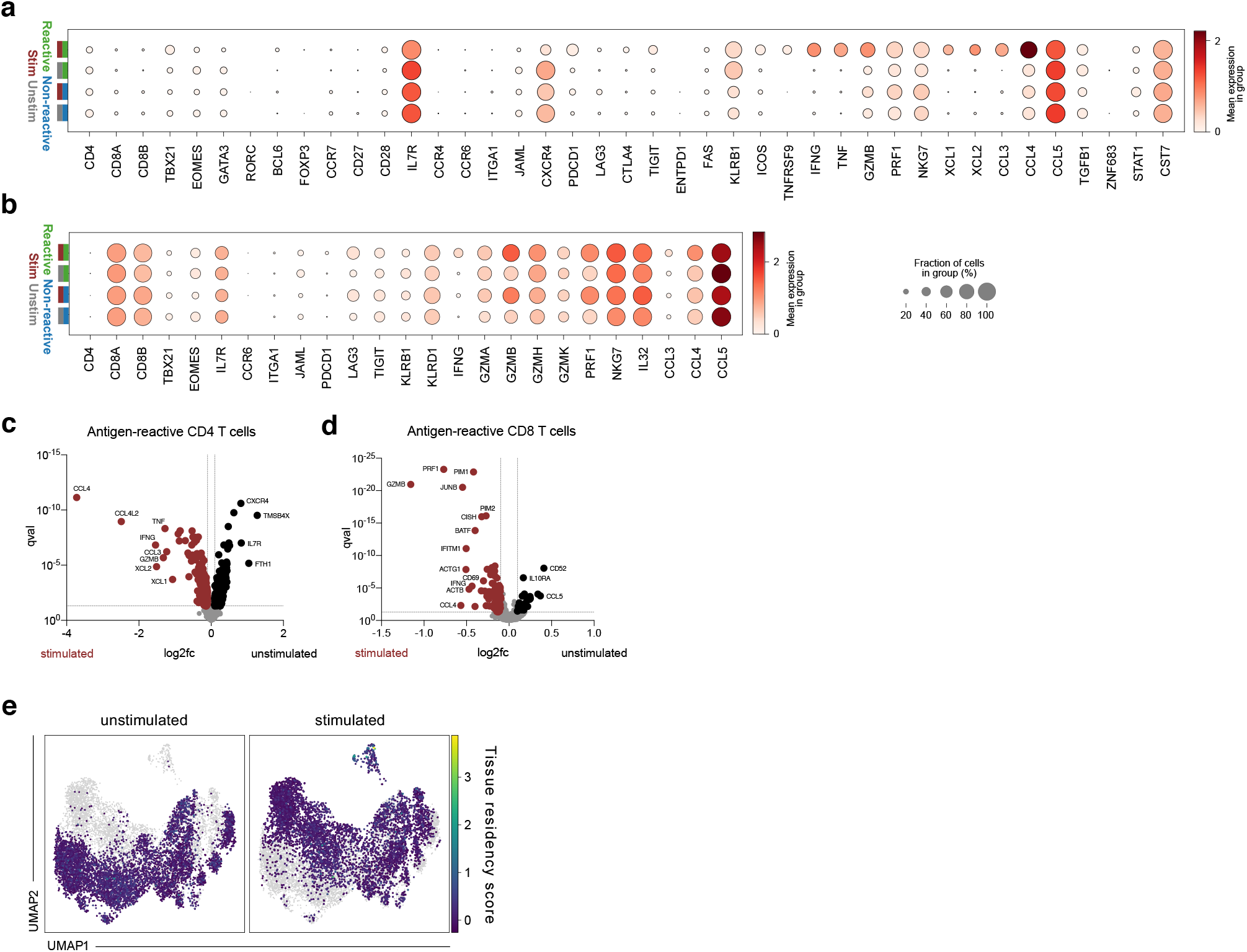
In vivo and stimulation-biased phenotypes of antigen-reactive T cells. **a, b**, Dot plots of log-normalized expression of selected marker genes by clonotype group (reactive and non-reactive) and by condition (stimulated: ‘stim’, unstimulated: ‘unstim’). **c, d**, Volcano plots of differential expression test of stimulated condition versus unstimulated condition in antigen reactive CD4 T cells (**c**) and CD8 T cells (**d**). **e**, UMAPs of all T cells with tissue residency score superimposed. For definition of tissue residency scores see methods section as well as reference^61^. Data are shown for patient GT_3.

**Extended Data Figure 7.**
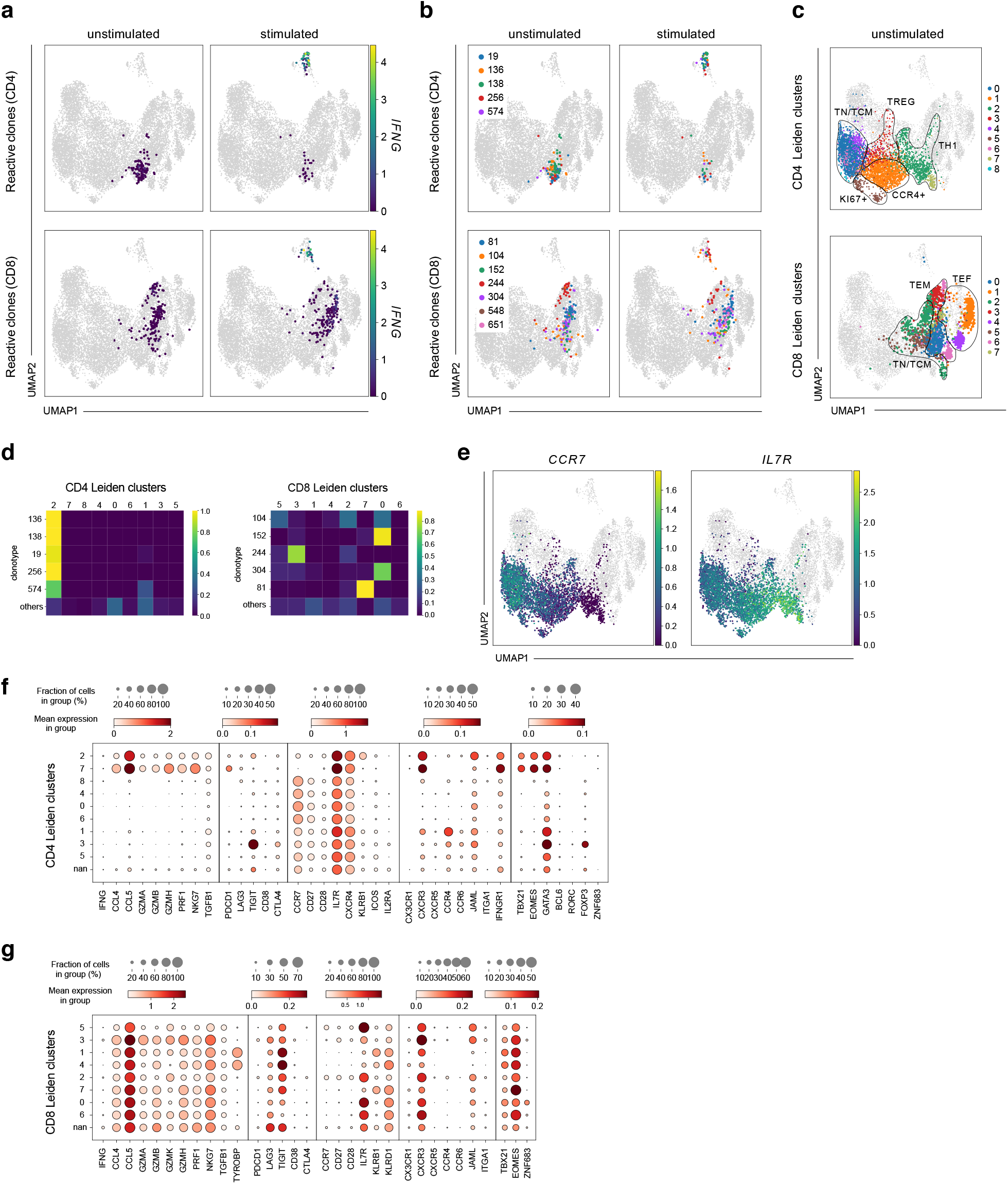
Phenotypic characterization of unstimulated T cells. **a, b**, UMAPs of all T cells, with cells from reactive clonotypes highlighted by cell type and with log-normalized *IFNG* expression (**a**) or clonotype identity (**b**) superimposed. **c**, UMAPs of all T cells from unstimulated condition highlighted and cell type-specific (CD4 or CD8) Leiden cluster annotation superimposed (UMAPs are identical to some of the UMAPs shown in Fig. 3c, but are shown here again for the sake of clarity (b-c)). **d**, Fraction of cells per clonotype in each Leiden cluster from (c) for both CD4 and CD8 T cells. **e**, UMAPs of all T cells with unstimulated CD4 T cells highlighted and log-normalized *CCR7* and *IL7R* expression superimposed. **f, g**, Dot plots of log-normalized expression of selected marker genes by cell type-specific Leiden cluster for CD4 T cells (**f**) and CD8 T cells (**g**). Data are shown for patient GT_3.

**Extended Data Figure 8.**
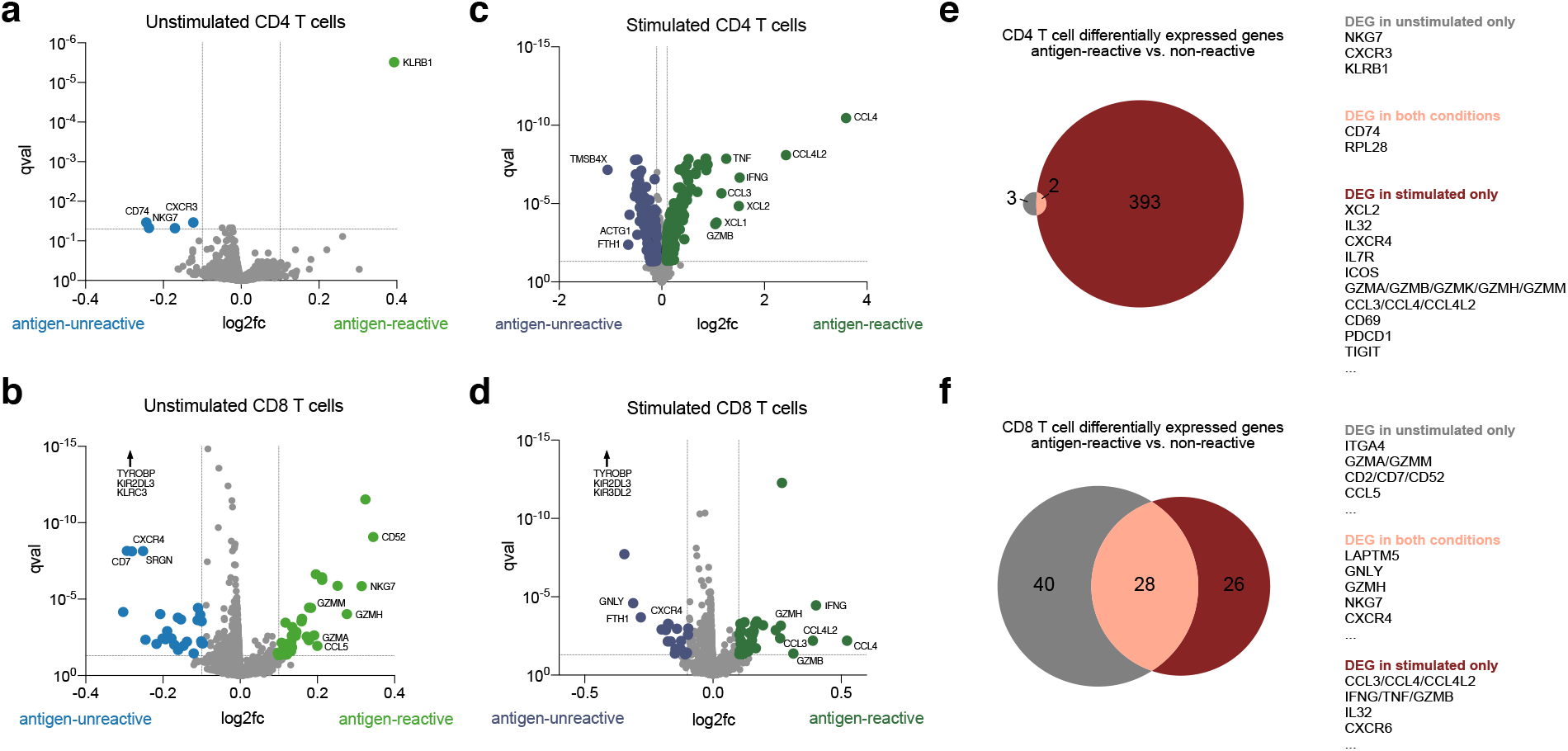
In vivo and stimulation-biased phenotypic signature scores of antigen-reactive T cells. **a-d**, Volcano plots of differential expression test of cells from non-reactive versus reactive clonotypes in unstimulated CD4 (**a**) and CD8 (**b**) T cells, and stimulated CD4 (**c**) and CD8 (**d**) T cells. For the sake of clarity, *TYROBP* (l2fc -0.48; qval 7×10^−131^), *KIR2DL3* (l2fc -0.15; qval 3×10^−60^) and *KLRC3* (l2fc -0.13; qval 3×10^−55^) are not displayed for unstimulated CD8 T cells, as well as *TYROBP* (l2fc -0.45; qval 1×10^−110^), *KIR2DL3* (l2fc -0.25; qval 1×10^−78^) and *KIR3DL2* (l2fc -0.10; qval 8×10^−22^) for stimulated CD8 T cells. **e, f**, Venn diagrams of overlaps of differentially expressed genes between tests from panel a, c (**e**) and b, d (**f**). Data are shown for patient GT_3.

**Extended Data Figure 9.**
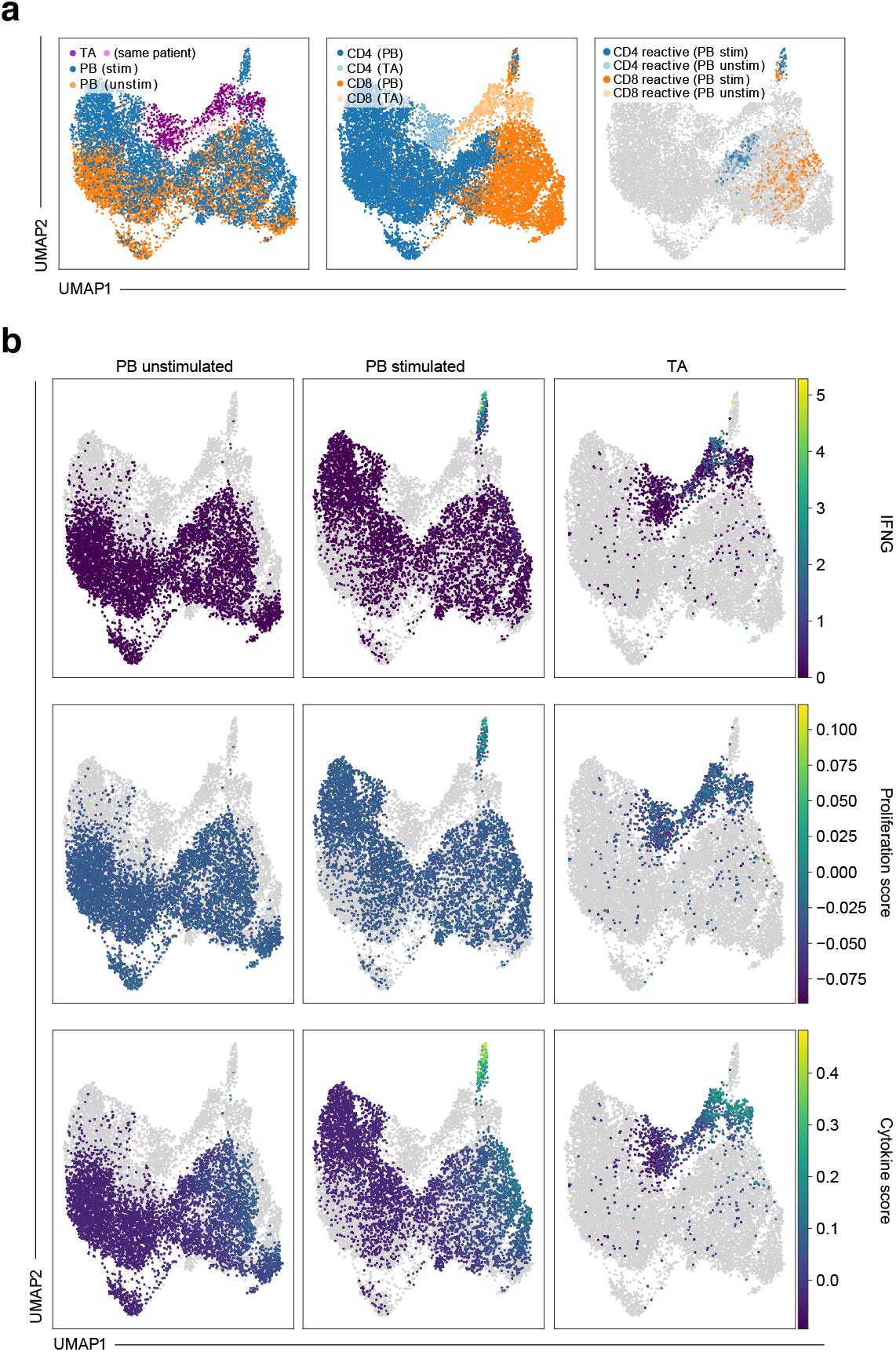
Characterization of cells from tracheal aspirate samples in context of stimulated and unstimulated PB T cells. **a**, UMAP of all PB T cells (stimulated and unstimulated) of patient GT_3 and cells from tracheal aspirate samples (TA) with sample, cell type and reactivity label in superimposed. **b**, UMAP as in a) with *IFNG* expression, proliferation score and cytokine score superimposed per data subset.

**Extended Data Figure 10.**
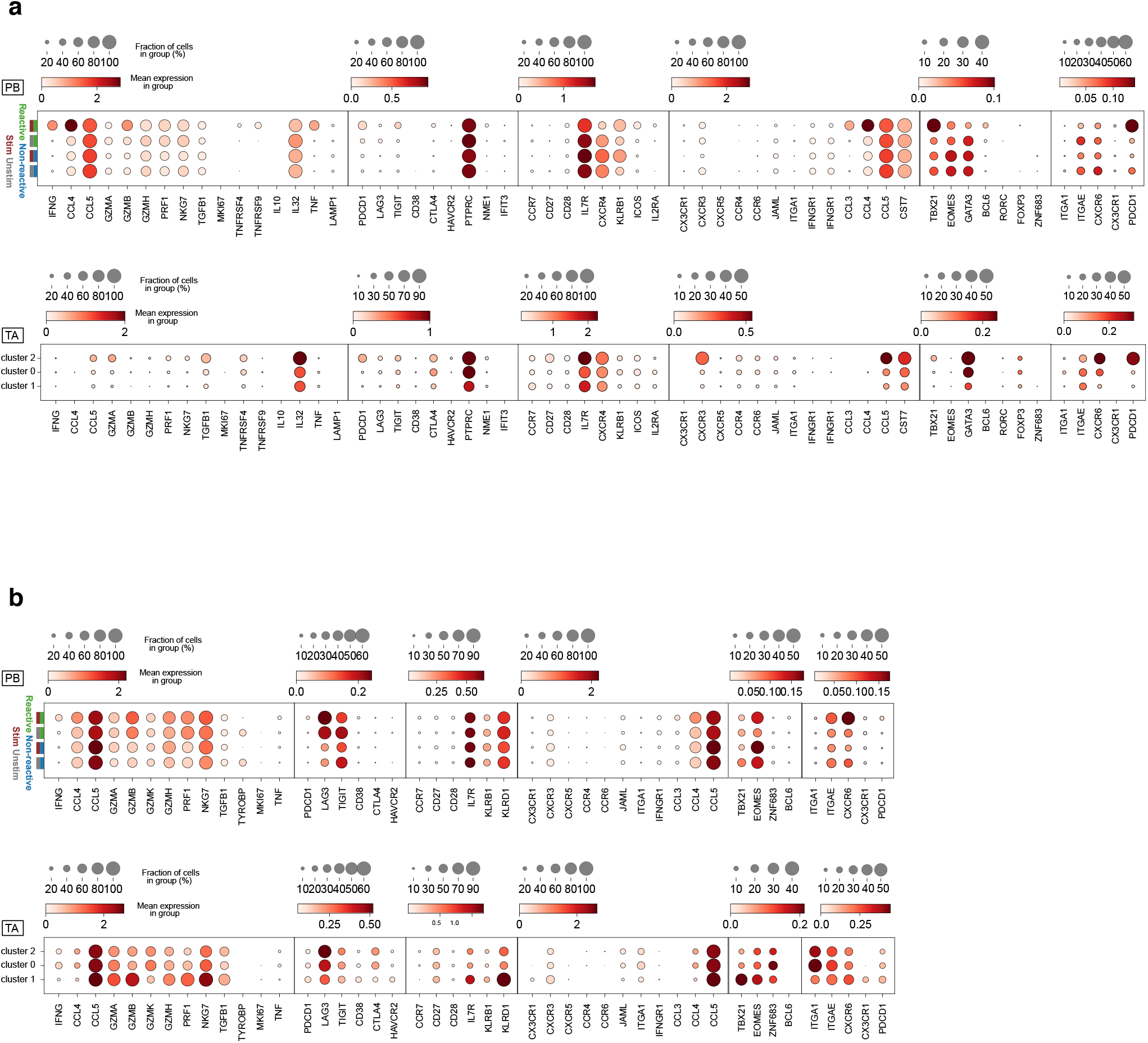
Characterization of clusters of cells from tracheal aspirate in context of stimulated and unstimulated PB T cells. Shown are dot plots for CD4 T cells (**a)** and CD8 T cells **(b)**. The upper dot plot in each panel contains grouping of PB T cells into reactive and non-reactive T cells within each condition (stimulated and unstimulated). The lower dot plot in each panel contains a clustering of the tracheal aspirate T cells.

**Extended Data Figure 11.**
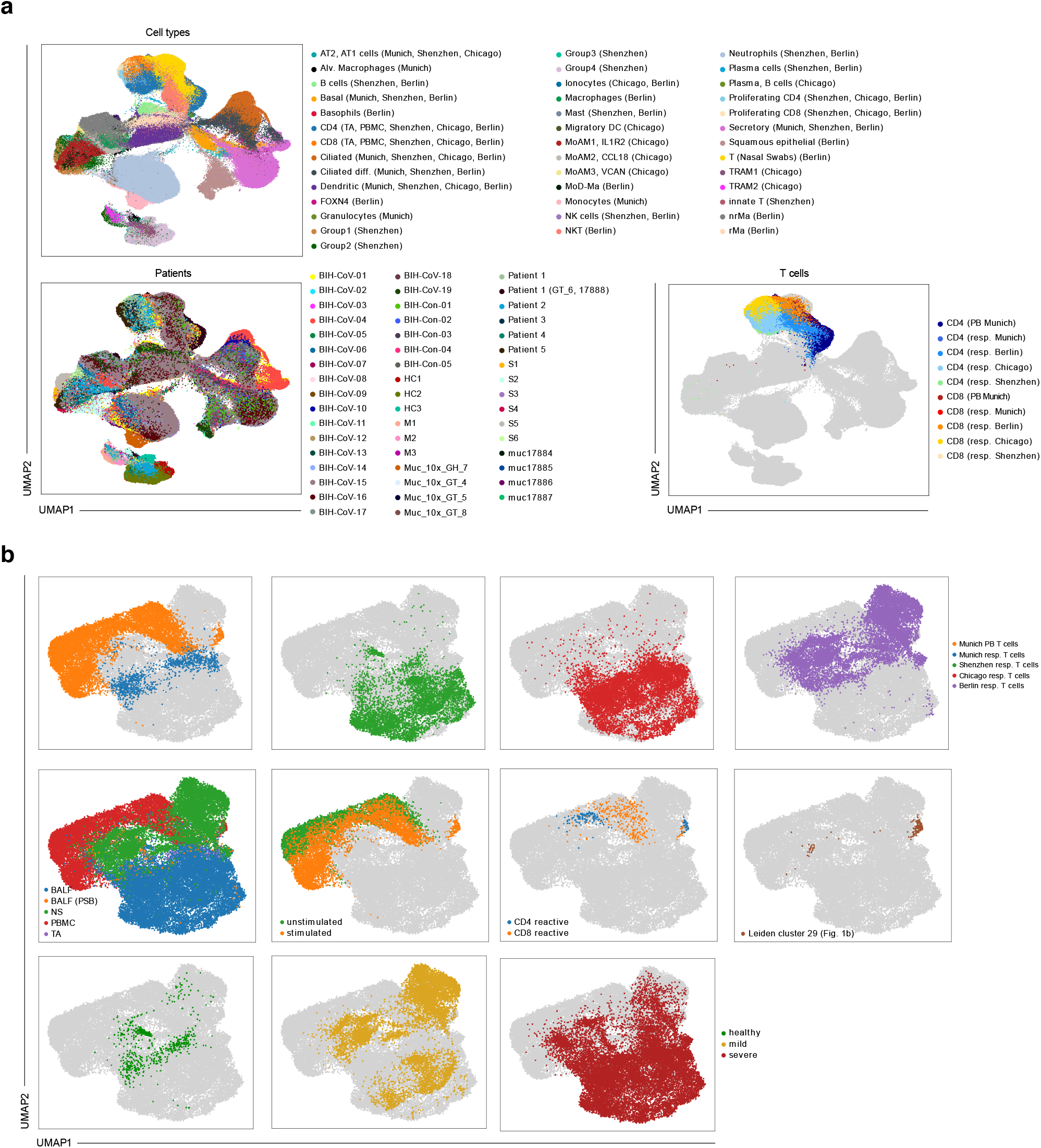
Joined embedding of cells from all cohorts. **a**, UMAP computed on all cells from samples from PB and respiratory tract samples of Munich, Chicago, Shenzhen and Berlin cohorts with cell type and sample (top), patient (bottom left) and highlighted T cells (bottom right) superimposed, **b**, UMAP computed on all T cells from all samples with cohort, sample source, condition, reactivity labels in PB T cells, IFNG+ cluster annotation from PB T cell analysis (Leiden cluster 29) and disease state in other cohorts (healthy, mild, severe) superimposed (from top to bottom and left to right).

**Extended Data Figure 12.**
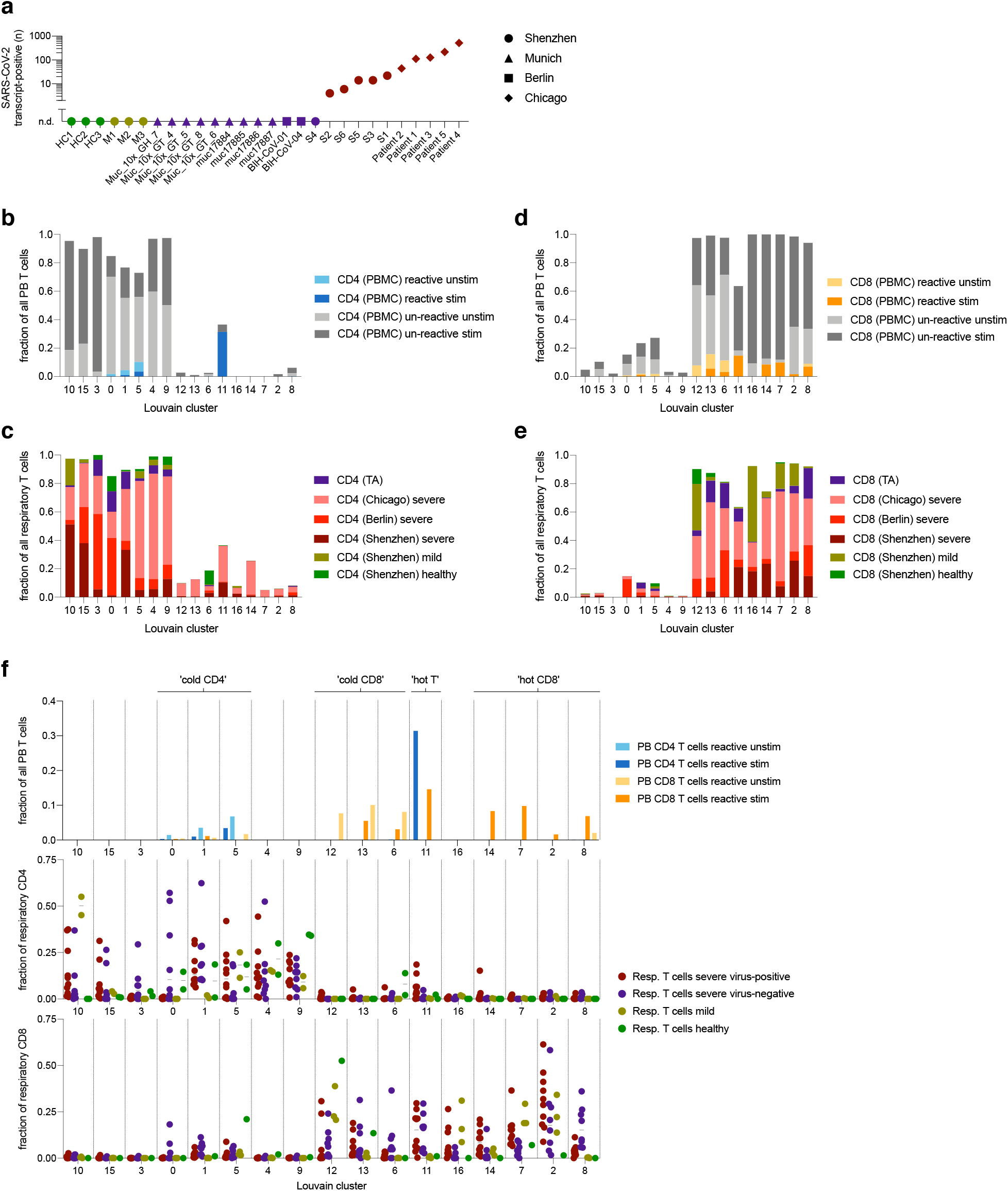
Characterization of clustering of joined T cell data set. **a**, Number of SARS-CoV-2 transcript-positive cells per patient among all (including non-T) cells in indicated scRNA seq data. **b-e**, Shown are fractions of cells with particular phenotypes per Louvain clustering, after clustering on all cells from samples from PB and respiratory tract samples of Munich, Chicago, Shenzhen and Berlin cohorts. Shown are reactive and un-reactive cells in stimulated (stim) and unstimulated (unstim) condition for both CD4 (b) and CD8 (c) PB T cells and fraction of cells in cohort for each CD4 (d) and CD8 (e) respiratory tract T cells. **f**, Fractions of all PB (top panel), of respiratory CD4 (middle panel) or of respiratory CD8 (bottom panel) T cells in individual patients (dots) from indicated four disease stages per indicated Louvain cluster. Patients with less than < 20 cells for each panel were excluded from analysis.

**Extended Data Figure 13.**
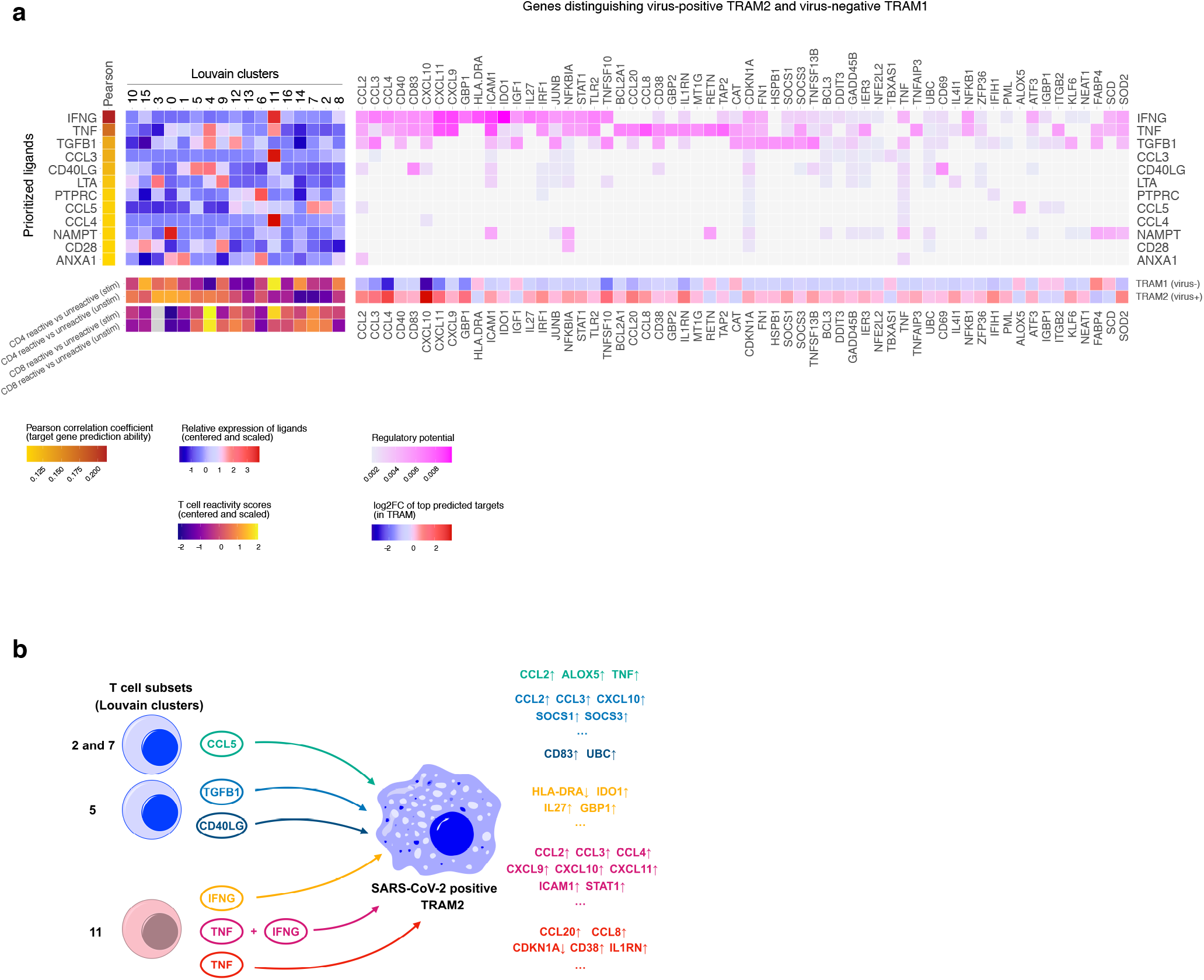
Cell-cell communication between individual T cell subsets and Tissue-resident Macrophages (TRAMs) from the respiratory tract of patients with severe COVID-19 in the scRNA-seq reference cohort from Chicago. **a**, Heatmap representation of the NicheNet analysis^50^ of ligand-target pairs regulating genes differentially expressed genes between virus-negative TRAM1 and virus-positive TRAM2. From left to right: (1) potential ligands expressed by respiratory tract T cells ranked according to their ability (Pearson correlation coefficient) to predict the gene expression changes observed between TRAM1 and TRAM2, (2) relative expression of the top ranked ligands in individual T cell subsets (Louvain clusters identified by unsupervised clustering of the integrated data set, see Fig. 4b) (top) and mean ‘in vivo signature scores’ per individual T cell subset (stimulated: ‘stim’, unstimulated: ‘unstim’), (3) potential targets of the top ranked ligands and their regulatory potential in TRAMs (top) and their log2-fold change in gene expression between TRAM1and TRAM2 (bottom). **b**, Graphical summary of results from (a) with exemplary sender cell groups/ligands and target genes.

**Extended Data Figure 14.**
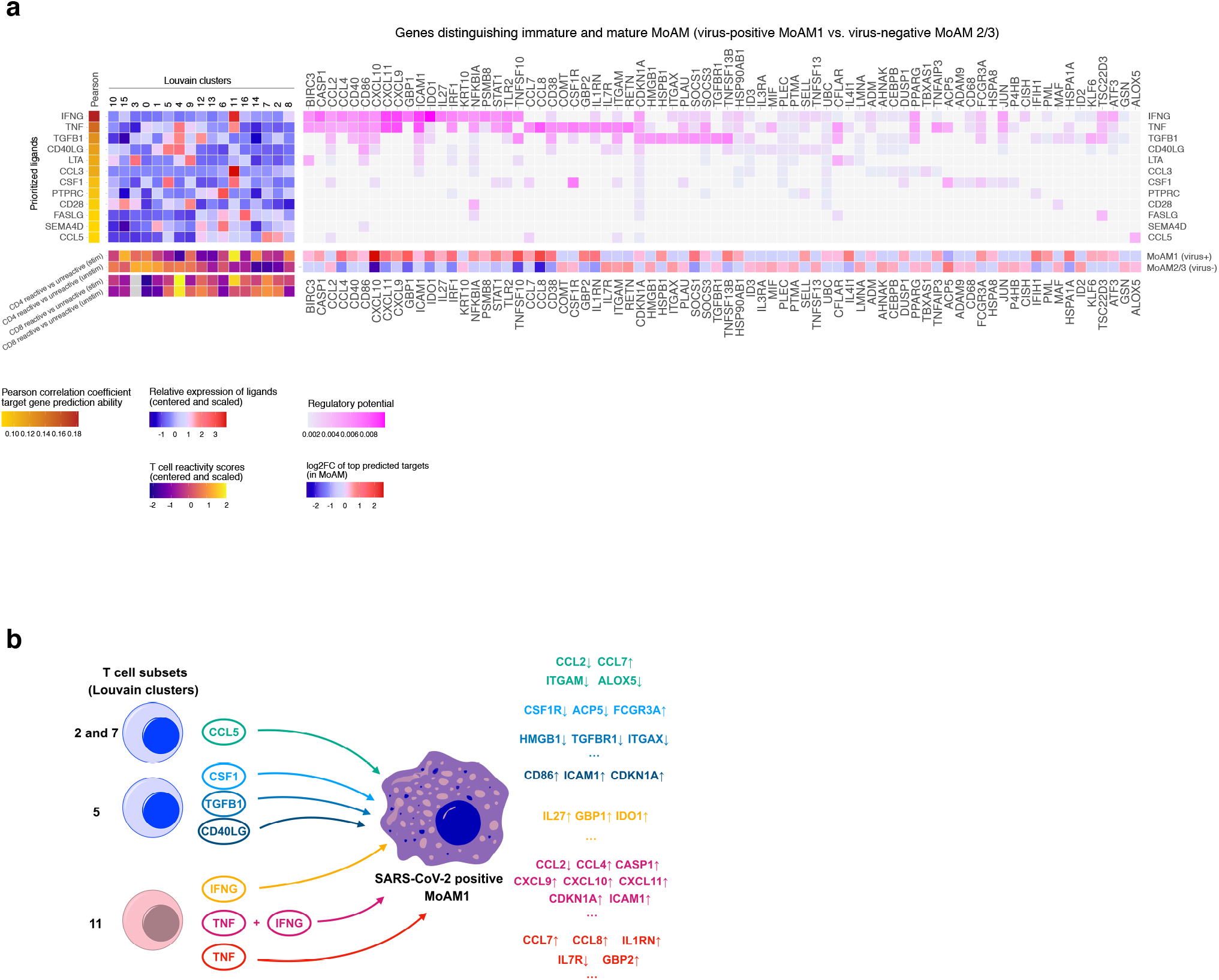
Cell-cell communication between individual T cell subsets and Monocyte-derived Macrophages (MoAMs) from the respiratory tract of patients with severe COVID-19 in the scRNA seq reference cohort from Chicago. **a**, Heatmap representation of the NicheNet analysis^50^ of ligand-target pairs regulating genes differentially expressed genes between virus-negative MoAM2/3 and virus-positive MoAM1. From left to right: (1) potential ligands expressed by respiratory tract T cells ranked according to their ability (Pearson correlation coefficient) to predict the gene expression changes observed between virus-negative MoAM2/3 and virus-positive MoAM1, (2) relative expression of the top ranked ligands in individual T cell subsets (Louvain clusters identified by unsupervised clustering of the integrated data set, see Fig. 4b) (top) and mean ‘in vivo signature scores’ per individual T cell subset (stimulated: ‘stim’, unstimulated: ‘unstim’), (3) potential targets of the top ranked ligands and their regulatory potential in MoAMs (top) and their log2-fold change in gene expression between virus-negative MoAM2/3 and virus-positive MoAM1 (bottom). **b**, Graphical summary of results from (a) with exemplary sender cell groups/ligands and target genes.

**Extended Data Figure 15.**
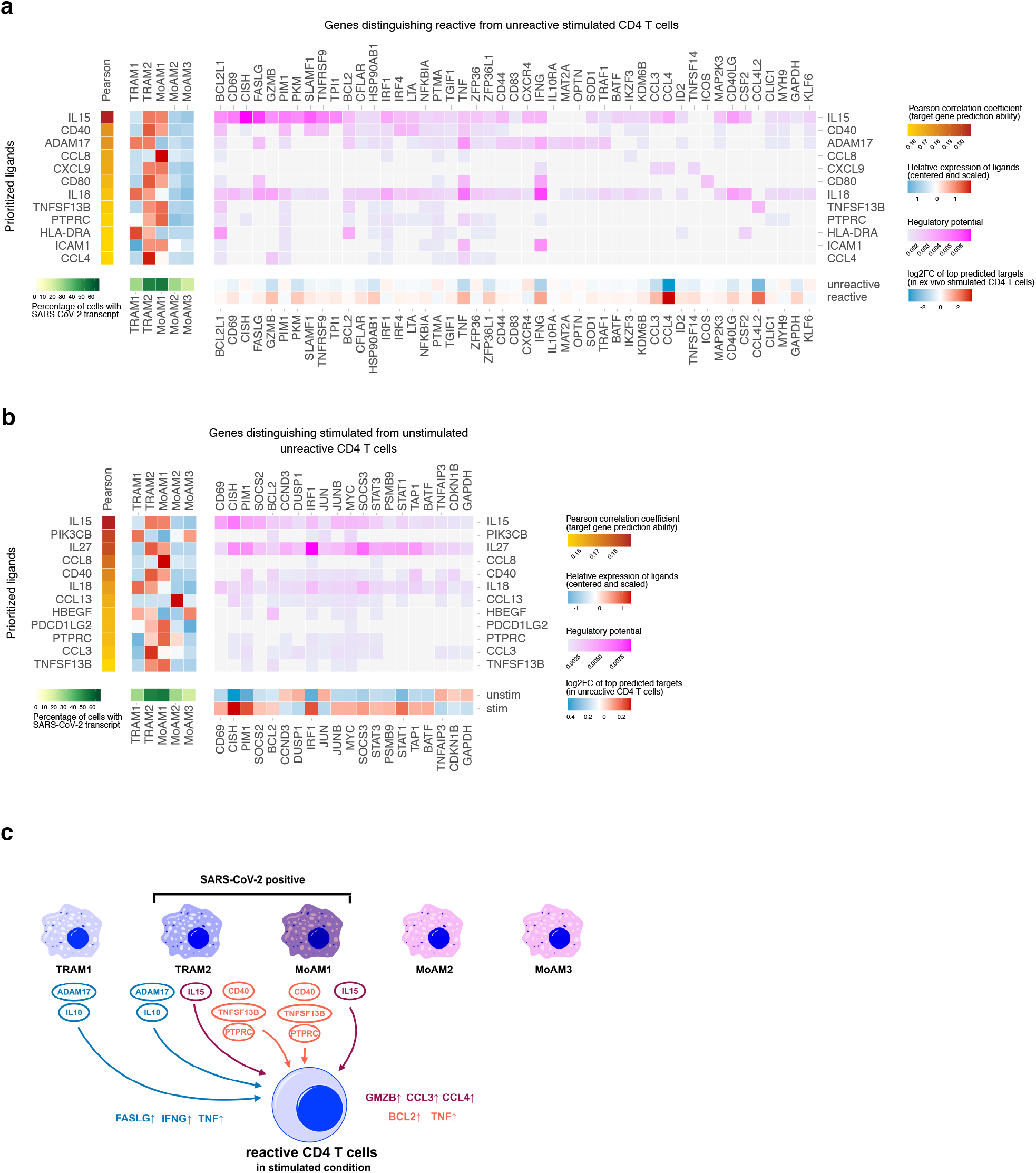
Cell-cell communication between macrophage subpopulations and CD4 T cells from patients with severe COVID-19 in the scRNA seq reference cohort from Chicago. **a, b**, Heatmap representation of the NicheNet analysis^50^ of ligand-target pairs regulating genes differentially expressed genes between **(a)** reactive and unreactive CD4 T cells after *in vitro* stimulation SARS-CoV-2 spike protein peptide mix and, to obtain an overview on unspecific ‘bystander’ effects present in an inflammatory environment, **(b)** stimulated and unstimulated unreactive CD 4 T cells. From left to right: (1) potential ligands expressed by macrophages ranked according to their ability (Pearson correlation coefficient) to predict the gene expression changes observed *in vitro*, (2) relative expression of the top ranked ligands in macrophage subpopulations (top) and the percentage of SARS-CoV-2 infected cells (SARS-CoV-2 genes per cell ≥ 1) per macrophage subpopulation (bottom), (3) potential targets of the top ranked ligands and their regulatory potential in CD4 T cells (top) and their log2-fold change in gene expression (bottom) between **(a)** reactive and unreactive *in vitro* stimulated CD4 T cells and **(b)** stimulated and unstimulated unreactive CD4 T cells. **c**, Graphical summary of results from (a) with exemplary ligands and target genes.

**Extended Data Figure 16.**
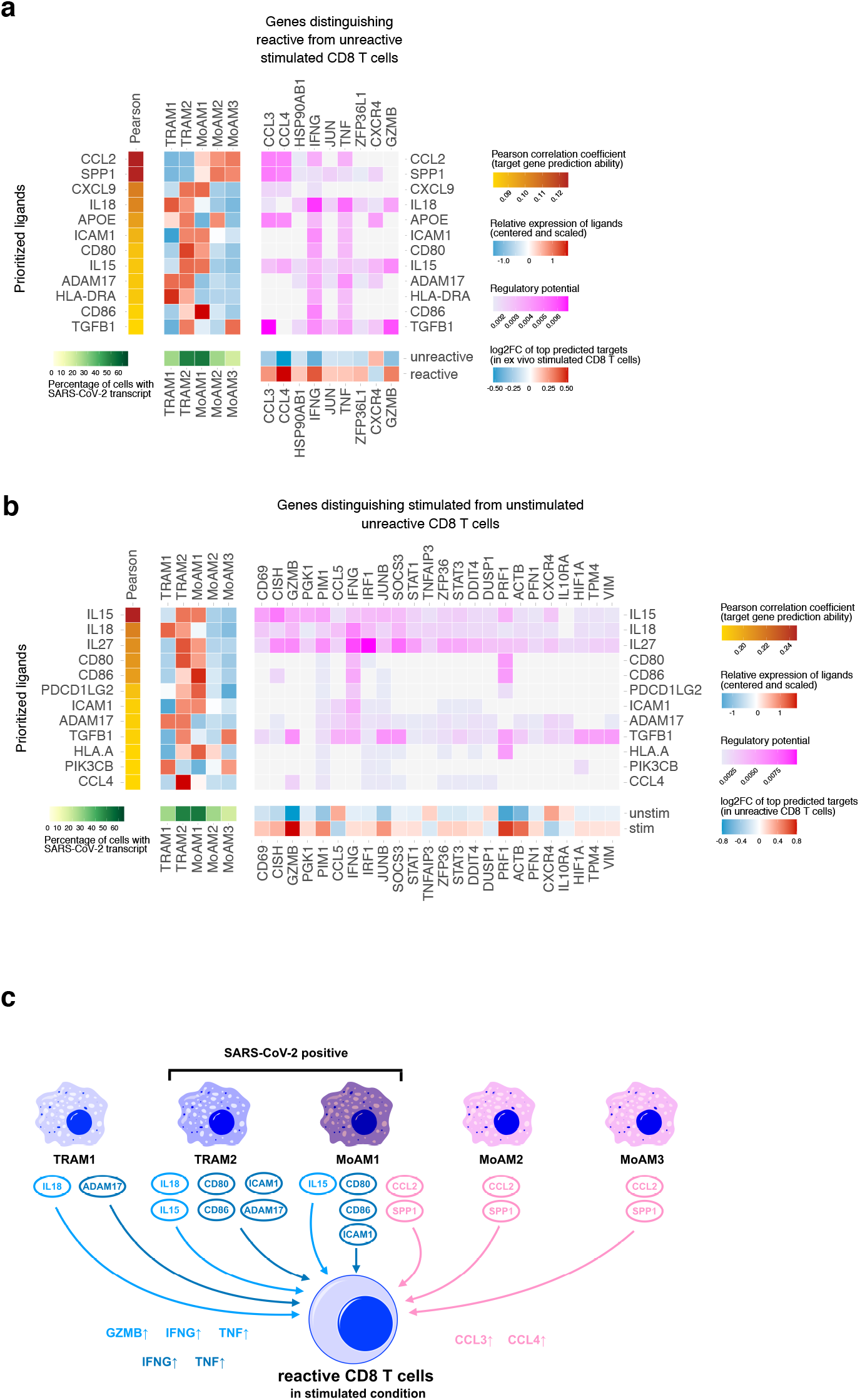
Cell-cell communication between macrophage subpopulations and CD8 T cells from patients with severe COVID-19 in the scRNA seq reference cohort from Chicago. **a, b**, Heatmap representation of the NicheNet analysis^50^ of ligand-target pairs regulating genes differentially expressed genes between **(a)** reactive and unreactive CD8 T cells after *in vitro* stimulation SARS-CoV-2 spike antigen peptide mix and, to obtain an overview on unspecific ‘bystander’ effects present in an inflammatory environment, **(b)** stimulated and unstimulated unreactive CD 4 T cells. From left to right: (1) potential ligands expressed by macrophages ranked according to their ability (Pearson correlation coefficient) to predict the gene expression changes observed *in vitro*, (2) relative expression of the top ranked ligands in macrophage subpopulations (top) and the percentage of SARS-CoV-2 infected cells (SARS-CoV-2 genes per cell ≥ 1) per macrophage subpopulation (bottom), (3) potential targets of the top ranked ligands and their regulatory potential in CD8 T cells (top) and their log2-fold change in gene expression (bottom) between **(a)** reactive and unreactive *in vitro* stimulated CD8 T cells and **(b)** stimulated and unstimulated unreactive CD8 T cells. **c**, Graphical summary of results from (a) with exemplary ligands and target genes.

**Extended Data Table 1. Clinical information on patients from Munich cohort**.

**Extended Data Table 2. Differential gene expression results of CD8 T cells in IFNG cluster compared with other CD8 T cells in stimulated condition**.

**Extended Data Table 3. Differential gene expression results of CD4 T cells in IFNG cluster compared with other CD4 T cells in stimulated condition**.

**Extended Data Table 4. Differential gene expression results of SARS-CoV-2-reactive CD4 T cells unstimulated versus stimulated condition**.

**Extended Data Table 5. Differential gene expression results of SARS-CoV-2-reactive CD8 T cells unstimulated versus stimulated condition**.

**Extended Data Table 6. Differential gene expression results of unstimulated CD4 T cells between SARS-CoV-2-reactive and non-reactive cells**.

**Extended Data Table 7. Differential gene expression results of unstimulated CD8 T cells between SARS-CoV-2-reactive and non-reactive cells**.

**Extended Data Table 8. Differential gene expression results of stimulated CD4 T cells between SARS-CoV-2-reactive and non-reactive cells**.

**Extended Data Table 9. Differential gene expression results of stimulated CD8 T cells between SARS-CoV-2-reactive and non-reactive cells**.

**Extended Data Table 10. Clonotypes selected for orthotopic TCR replacement**.

**Extended Data Table 11. Clonotypes discovered in GT_2 after filtering**.

**Extended Data Table 12. Clonotypes discovered in GT_3 after filtering**.

**Extended Data File 1. Code for reproduction of analysis**.

## ACKNOWLEDGMENTS

We thank members of the Schiller and Busch laboratories as well as Veit Buchholz and Michael Neuenhahn for experimental and analytical help as well as critical discussions. We also gratefully acknowledge the provision of human biomaterial and clinical data from the CPC-M bioArchive and its partners at the Asklepios Biobank Gauting, the Klinikum der Universität München and The Ludwig-Maximilians-Universität München.

K.S., D.H.B. and K.I.W. acknowledge support by the German National Network of University Medicine of the Federal Ministry of Education and Research (BMBF; NaFoUniMedCovid19, 01KX2021; COVIM), as well as the German Center for Infection Research (DZIF) and the German Research Foundation (DFG; SFB 1321/TP17; SFB 1054/B09 and SFB 1371/TP04). H.B.S. and M.A. acknowledge support by the German Center for Lung Research (DZL), the Helmholtz Association, the BMBF project Single Cell Genomics Network Germany, the European Union’s Horizon 2020 research and innovation program (grant agreement 874656). F.J.T. acknowledges support by the BMBF (01IS18036B and 01IS18053A) and the Chan Zuckerberg Initiative (2019-207271 and 2019-207271) and by the Helmholtz Association’s Initiative and Networking Fund through Helmholtz AI (ZT-I-PF-5-01). D.S.F. acknowledges support from a German Research Foundation (DFG) fellowship through the Graduate School of Quantitative Biosciences Munich (QBM) [GSC 1006] and by the Joachim Herz Stiftung.

## AUTHOR CONTRIBUTIONS

K.S., H.B.S., D.H.B., F.J.T. and B.S. conceptualized the study and developed methodology; D.S.F., M.A., and N.J.L. developed and performed software analyses; K.S., D.S.F., A.M., and K.I.W. conducted formal analysis of the data; K.I.W., S.J. Y.H., C.H.M., M.S., M.H., E.D., L.M., S.W. and L.S.W. performed experiments; A.H., I.E.F., G.L., K.M., J.B., M.F., L.N., K.H., I.K. and M.G.S. acquired patient data and samples and performed medical evaluation and analysis; H.B.S., F.J.T., A.P. K.W. contributed resources; K.S. wrote the manuscript; all authors read and approved the manuscript; H.B.S., D.H.B. and F.J.T. acquired most of the funding; K.S. and H.B.S. supervised the study and administered the project.

## CODE AVAILABILITY

For scRNA seq data analysis algorithms see methods section and jupyter notebooks in Extended Data File 1. Additional information is available from the corresponding authors upon reasonable request.

## MATERIALS & CORRESPONDANCE

Kilian Schober, Institute for Medical Microbiology, Immunology and Hygiene, Technische Universität München (TUM), Munich, Germany; or Herbert B. Schiller, Institute of Lung Biology and Disease, Comprehensive Pneumology Center, Helmholtz Zentrum München, Neuherberg, Germany.

