## Extended Data File 1 for "Single-cell RNA sequencing reveals *in vivo* signatures of SARS-CoV-2-reactive T cells through ‘reverse phenotyping’": 1_basic_analysis.ipynb

```
# %% [markdown]
# Load packages

# %% [code]
import anndata
import diffxpy.api as de
import numpy as np
import scanpy as sc
import scirpy as ir
import scipy.stats
import sys
import pandas as pd
import seaborn as sb
import matplotlib.pyplot as plt
from matplotlib import rcParams
from matplotlib import colors

np.random.seed(seed=1)

# %% [code]
import warnings
warnings.filterwarnings('ignore')

# %% [code]
sc.settings.verbosity = 3 # amount of output
pd.options.display.max_columns = 999

# Put your name here and comment other out.
user = "david"

# Directories on your machine with backslash at the end.
if user == "david_server":
    data_dir_vdj = '/storage/scratch/users/xavier.pastor/projects/20200727_Covid19/data/10x_vdj/cellranger_vdj/'
    data_dir_rna = '/storage/groups/bcf/projects/SingleCell/10x/projects/schiller/200724_Covid19_stimulated/cellranger/'
    data_dir_ta = None
    data_dir_ta_objects = None
    dir_out = '/storage/groups/ml01/workspace/david.fischer/covid19tcr/results/'
    dir_data = '/storage/groups/ml01/workspace/david.fischer/covid19tcr/data/'
    dir_data_liao = '/storage/groups/ml01/workspace/david.fischer/covid19tcr/data/GSE145926_RAW/'
    dir_data_lee = '/storage/groups/ml01/workspace/david.fischer/covid19tcr/data/2020_Lee_CovidFlu/'
elif user == "david":
    data_dir_vdj = '/Users/david.fischer/phd/data/covidtcr/data/pbmc/cellranger_vdj/'
    data_dir_rna = '/Users/david.fischer/phd/data/covidtcr/data/pbmc/cellranger/'
    data_dir_ta = '/Users/david.fischer/phd/data/covidtcr/data/ta/cellranger/'
    data_dir_ta_objects = '/Users/david.fischer/phd/data/covidtcr/data/ta/'
    dir_out = '/Users/david.fischer/phd/data/covidtcr/results/'
    dir_data = '/Users/david.fischer/phd/data/covidtcr/data/'
    dir_data_liao = '/Users/david.fischer/phd/datasets/2020_Liao_SARSCOV2_TCR/GSE145926_RAW/'
    dir_data_lee = '/Users/david.fischer/phd/datasets/2020_Lee_CovidFlu/'

# Make sure that these folders exist:
dir_panels = dir_out + 'panels/'
dir_tables = dir_out + 'tables/'
dir_anndata = dir_out + 'anndata/'
dir_genesets = dir_data + "genesets/"
dir_reference = dir_data + "reference/"

sc.logging.print_versions()
sc.settings.set_figure_params(dpi=80, scanpy=True)
sc.settings.figdir = dir_panels
print(sys.version)

# %% [code]
bool_recomp = True
bool_recluster = False
bool_recomp_dist = False

# %% [markdown]
# Read data

# %% [markdown]
## PBMC

# %% [code]
sample_ids_3 = [  # these have very few TCR observations
    "Sample_20063a009_01",
    "Sample_20063a010_01",
    "Sample_20063a011_01",
    "Sample_20063a012_01"
]
sample_ids_5 = [  # these have very low RNA counts
    "Sample_20063a013_01",
    "Sample_20063a014_01",
    "Sample_20063a015_01",
    "Sample_20063a016_01"
]
if bool_recomp:
    adata_ls = {}
    for x, y in zip(sample_ids_3, sample_ids_5):
        adata_rna = sc.read_10x_h5(data_dir_rna+x+"/count_matrices/filtered_feature_bc_matrix.h5")
        adata_tcr = ir.io.read_10x_vdj(data_dir_vdj+y+"/outs/all_contig_annotations.json", filtered=True)
        ir.pp.merge_with_tcr(adata_rna, adata_tcr)
        adata_rna.obs["sample_rna"] = x
        adata_rna.obs["sample_tcr"] = y
        adata_rna.var_names_make_unique()
        # Load cellranger clonotypes:
        tab = pd.read_csv(data_dir_vdj + y + "/outs/filtered_contig_annotations.csv")
        tab["barcode_adjusted"] = ["-".join([xx, x]) for xx in tab["barcode"].values]
        adata_rna.obs["clonotype_cellranger"] = [
            tab.loc[tab["barcode_adjusted"].values == xx, 'raw_clonotype_id'].values[0] + "_" + x
            if xx in tab["barcode_adjusted"].values else "none" 
            for xx in adata_rna.obs_names
        ]
        adata_ls[x] = adata_rna
    adata_raw_p1 = adata_ls[sample_ids_3[0]].concatenate(
        [adata_ls[sample_ids_3[1]]],
        batch_key="concatenation", batch_categories=sample_ids_3[:2]
    )
    adata_raw_p1.obs["patient"] = "p1"
    adata_raw_p1.obs["condition"] = [
        {
            "Sample_20063a009_01": "unstimulated",
            "Sample_20063a010_01": "stimulated",
            "Sample_20063a011_01": "unstimulated",
            "Sample_20063a012_01": "stimulated"
        }[x] for x in adata_raw_p1.obs["sample_rna"].values
    ]
    adata_raw_p2 = adata_ls[sample_ids_3[2]].concatenate(
        [adata_ls[sample_ids_3[3]]],
        batch_key="concatenation", batch_categories=sample_ids_3[2:]
    )
    adata_raw_p2.obs["patient"] = "p2"
    adata_raw_p2.obs["condition"] = [
        {
            "Sample_20063a009_01": "unstimulated",
            "Sample_20063a010_01": "stimulated",
            "Sample_20063a011_01": "unstimulated",
            "Sample_20063a012_01": "stimulated"
        }[x] for x in adata_raw_p2.obs["sample_rna"].values
    ]
    adata_raw_p1.raw = adata_raw_p1.copy()
    adata_raw_p2.raw = adata_raw_p2.copy()
else:
    adata_raw_p1 = sc.read(dir_anndata+'adata_raw_p1.h5ad') 
    adata_raw_p2 = sc.read(dir_anndata+'adata_raw_p2.h5ad') 

# %% [markdown]
## TA

# %% [code]
sample_ids = {
    'Muc_10x_GH_7': "other",
    'Muc_10x_GT_5': "other",
    'Muc_10x_GT_6': "P1",
    'Muc_10x_GT_8': "other",
    'MUC17884': "other",
    'MUC17885': "other",
    'MUC17886': "other",
    'MUC17887': "other",
    'MUC17888': "P1",
}
if bool_recomp:
    adata_ls = {}
    for k, v in sample_ids.items():
        adata_rna = sc.read_10x_h5(data_dir_ta+k+"/count_matrices/filtered_feature_bc_matrix.h5")
        adata_rna.obs["patient"] = v
        adata_rna.var_names_make_unique()
        adata_ls[k] = adata_rna
    adata_raw_ta = adata_ls[list(sample_ids.keys())[0]].concatenate(
        [adata_ls[x] for x in list(sample_ids.keys())[1:]],
        batch_key="concatenation", batch_categories=list(sample_ids.keys())
    )
    adata_raw_ta.raw = adata_raw_ta.copy()
else:
    adata_raw_ta = sc.read(dir_anndata+'adata_raw_ta.h5ad')

# %% [code]
adata_p1_withta = sc.read_h5ad(data_dir_ta_objects+"Covid19_TAs_PBMCs.h5ad")

# %% [code]
np.unique(np.asarray(adata_p1_withta.layers["counts"].todense()))

# %% [code]
adata_p1_withta.layers["lognormalised"] = np.log(
    adata_p1_withta.layers["counts"] / np.sum(adata_p1_withta.layers["counts"], axis=1) * 10000 +1
)

# %% [markdown]
## Define gene sets

# %% [code]
markers_cd4 = {
    "CD4 cytokines": [
        "IFNG",
        "CCL4",
        "CCL5",
        "GZMA",
        "GZMB",
        "GZMH",
        "PRF1",
        "NKG7",
        "TGFB1",
    ],
    "CD4 co-inhibition": [
        "PDCD1",
        "LAG3",
        "TIGIT",
        "CD38",
        "CTLA4",
    ],
    "CD4 Differentiation": [
        "CCR7",
        "CD27",
        "CD28",
        "IL7R",
        "CXCR4",
        "KLRB1",
        "ICOS",
        "IL2RA",
    ],
    "CD4 chemokines and sensing": [
        "CX3CR1",
        "CXCR3",
        "CXCR5",
        "CCR4",
        "CCR6",
        "JAML",
        "ITGA1",
        "IFNGR1",
    ],
    "CD4 transcription factors": [
        "TBX21",
        "EOMES",
        "GATA3",
        "BCL6",
        "RORC",
        "FOXP3",
        "ZNF683",
    ],
    "TRM": [
        "ITGA1",
        "CXCR6",
        "ITGAE",
        "CXCR6",
        "CX3CR1",
        "PDCD1"
    ]
}
markers_cd4_ta = {
    "CD4 cytokines": markers_cd4["CD4 cytokines"] + [
        "MKI67", 
        "TNFRSF4", 
        "TNFRSF9",
        "IL10",
        "IL32", 
        "TNF", 
        "LAMP1",
        "IL1B",
    ],
    "CD4 co-inhibition": markers_cd4["CD4 co-inhibition"] + [
        "HAVCR2", "PTPRC", "NME1", "IFIT3"
    ],
    "CD4 Differentiation": markers_cd4["CD4 Differentiation"],
    "CD4 chemokines and sensing": markers_cd4["CD4 chemokines and sensing"] + [
        "IFNGR1", "CCL3", "CCL4", "CCL5", "CST7"
    ],
    "CD4 transcription factors": markers_cd4["CD4 transcription factors"],
    "TRM": markers_cd4["TRM"]
}

# %% [code]
markers_cd8 = {
    "CD8 cytokines": [
        "IFNG",
        "CCL4",
        "CCL5",
        "GZMA",
        "GZMB",
        "GZMK",
        "GZMH",
        "PRF1",
        "NKG7",
        "TGFB1",
        "TYROBP",
    ],
    "CD8 co-inhibition": [
        "PDCD1",
        "LAG3",
        "TIGIT",
        "CD38",
        "CTLA4",
    ],
    "CD8 Differentiation": [
        "CCR7",
        "CD27",
        "CD28",
        "IL7R",
        "KLRB1",
        "KLRD1",
    ],
    "CD8 chemokines and sensing": [
        "CX3CR1",
        "CXCR3",
        "CXCR5",
        "CCR4",
        "CCR6",
        "JAML",
        "ITGA1",
    ],
    "CD8 transcription factors": [
        "TBX21",
        "EOMES",
        "ZNF683",
    ],
    "TRM": [
        "ITGA1",
        "CXCR6",
        "ITGAE",
        "CXCR6",
        "CX3CR1",
        "PDCD1"
    ]
}
markers_cd8_ta = {
    "CD8 cytokines": markers_cd8["CD8 cytokines"] + [
        "MKI67", 
        "TNF", 
    ],
    "CD8 co-inhibition": markers_cd8["CD8 co-inhibition"] + [
        "HAVCR2", 
    ],
    "CD8 Differentiation": markers_cd8["CD8 Differentiation"],
    "CD8 chemokines and sensing": markers_cd8["CD8 chemokines and sensing"] + [
        "IFNGR1", "CCL3", "CCL4", "CCL5"
    ],
    "CD8 transcription factors": markers_cd8["CD8 transcription factors"] + [
        "BCL6"
    ],
    "TRM": markers_cd8["TRM"]
}

# %% [code]
markers_2c = {
    "activation": [
        "TNFRSF9",
        "IFNG",
        "TNF",
        "GZMB",
        "PRF1",
        "NKG7",
        "XCL1",
        "XCL2",
        "CCL3",
        "CCL4",
        "CCL5",
        "CST7",
        "STAT1",
        "TGFB1",
    ],
    "co-inhibition":  [
        "PDCD1",
        "LAG3",
        "CTLA4",
        "TIGIT",
    ],
    "differentiation":  [
        "CCR7",
        "CD27",
        "CD28",
        "CXCR4",
        "KLRB1",
        "ICOS",
    ],
    "tf": [
        "TBX21", #T-bet
        "EOMES",
        "GATA3",
        "BCL6",
        "RORC",
        "FOXP3",
        "ZNF683",
    ],
}

# %% [code]
markers_s6ab = {
    "cd4": [
        "CD4",
        "CD8A",
        "CD8B",
        "TBX21",
        "EOMES",
        "GATA3",
        "RORC",
        "BCL6",
        "FOXP3",
        "CCR7",
        "CD27",
        "CD28",
        "IL7R",
        "CCR4",
        "CCR6",
        "ITGA1",
        "JAML",
        "CXCR4",
        "PDCD1",
        "LAG3",
        "CTLA4",
        "TIGIT",
        "ENTPD1",
        "FAS",
        "KLRB1",
        "ICOS",
        "TNFRSF9",
        "IFNG",
        "TNF",
        "GZMB",
        "PRF1",
        "NKG7",
        "XCL1",
        "XCL2",
        "CCL3",
        "CCL4",
        "CCL5",
        "TGFB1",
        "ZNF683", 
        "STAT1",
        "CST7"
    ],
    "cd8": [
        "CD4",
        "CD8A",
        "CD8B",
        "TBX21",
        "EOMES",
        "IL7R",
        "CCR6",
        "ITGA1",
        "JAML",
        "PDCD1",
        "LAG3",
        "TIGIT",
        "KLRB1",
        "KLRD1",
        "IFNG",
        "GZMA",
        "GZMB",
        "GZMH",
        "GZMK",
        "PRF1",
        "NKG7",
        "IL32",
        "CCL3",
        "CCL4",
        "CCL5",
    ]
}

# %% [markdown]
# PBMC processing

# %% [markdown]
## Cell filtering

# %% [markdown]
### Basic stats

# %% [markdown]
#### P1

# %% [code]
if bool_recomp:
    adata_raw_p1.obs['n_genes'] = np.sum(adata_raw_p1.X > 0, axis=1)
    adata_raw_p1.obs['n_counts'] = np.sum(adata_raw_p1.X, axis=1)
    mt_gene_mask = [gene.lower().startswith('mt-') for gene in adata_raw_p1.var_names]
    adata_raw_p1.obs['mt_frac'] = np.asarray(adata_raw_p1[:,mt_gene_mask].X.sum(axis=1)).flatten() / \
        adata_raw_p1.obs['n_counts'].values

# %% [code]
sc.pl.scatter(adata_raw_p1, 'n_counts', 'n_genes', color='mt_frac', size=5)
sc.pl.scatter(adata_raw_p1[adata_raw_p1.obs['n_counts']<2000], 'n_counts', 'n_genes', color='mt_frac', size=5)

# %% [code]
adata_raw_p1.obs["CD3_all_genes"] = np.asarray(np.sum(adata_raw_p1[:, ["CD3D", "CD3E", "CD3G"]].X.todense(), axis=1)).flatten()
sc.pl.scatter(adata_raw_p1, 'n_counts', 'CD3_all_genes', color='mt_frac', size=5)

# %% [code]
if bool_recomp:
    sc.pp.filter_cells(adata_raw_p1, min_genes=200)
    sc.pp.filter_cells(adata_raw_p1, max_counts=15000)

# %% [markdown]
#### P2

# %% [code]
if bool_recomp:
    adata_raw_p2.obs['n_genes'] = np.sum(adata_raw_p2.X > 0, axis=1)
    adata_raw_p2.obs['n_counts'] = np.sum(adata_raw_p2.X, axis=1)
    mt_gene_mask = [gene.lower().startswith('mt-') for gene in adata_raw_p2.var_names]
    adata_raw_p2.obs['mt_frac'] = np.asarray(adata_raw_p2[:,mt_gene_mask].X.sum(axis=1)).flatten() / \
        adata_raw_p2.obs['n_counts'].values

# %% [code]
sc.pl.scatter(adata_raw_p2, 'n_counts', 'n_genes', color='mt_frac', size=5)
sc.pl.scatter(adata_raw_p2[adata_raw_p2.obs['n_counts']<2000], 'n_counts', 'n_genes', color='mt_frac', size=5)

# %% [code]
adata_raw_p2.obs["CD3_all_genes"] = np.asarray(np.sum(adata_raw_p2[:, ["CD3D", "CD3E", "CD3G"]].X.todense(), axis=1)).flatten()
sc.pl.scatter(adata_raw_p2, 'n_counts', 'CD3_all_genes', color='mt_frac', size=5)

# %% [code]
if bool_recomp:
    sc.pp.filter_cells(adata_raw_p2, min_genes=200)
    sc.pp.filter_cells(adata_raw_p2, max_counts=15000)

# %% [markdown]
### Cluster-based refined filtering

# %% [code]
if bool_recomp:
    sc.pp.filter_genes(adata_raw_p1, min_cells=10)
    sc.pp.normalize_per_cell(adata_raw_p1, counts_per_cell_after=1000)
    sc.pp.log1p(adata_raw_p1)
    sc.pp.pca(adata_raw_p1)
    sc.external.pp.bbknn(adata_raw_p1, batch_key="condition")
    sc.tl.umap(adata_raw_p1)
    if bool_recluster == True:
        sc.tl.leiden(adata_raw_p1, resolution=3., random_state=0)
        if bool_recluster:
            pd.DataFrame(adata_raw_p1.obs).to_csv(path_or_buf=dir_anndata+"adata_raw_p1_obs.csv")
    else:
        obs = pd.read_csv(dir_anndata+'adata_raw_p1_obs.csv')
        adata_raw_p1.obs['leiden'] = pd.Categorical(
            obs['leiden'].values.astype(str), 
            categories=np.sort(np.unique(obs['leiden'].values.astype(str)))
        )
    
    sc.pp.filter_genes(adata_raw_p2, min_cells=10)
    sc.pp.normalize_per_cell(adata_raw_p2, counts_per_cell_after=1000)
    sc.pp.log1p(adata_raw_p2)
    sc.pp.pca(adata_raw_p2)
    sc.external.pp.bbknn(adata_raw_p2, batch_key="condition")
    sc.tl.umap(adata_raw_p2)
    if bool_recluster == True:
        sc.tl.leiden(adata_raw_p2, resolution=3., random_state=0)
        if bool_recluster:
            pd.DataFrame(adata_raw_p2.obs).to_csv(path_or_buf=dir_anndata+"adata_raw_p2_obs.csv")
    else:
        obs = pd.read_csv(dir_anndata+'adata_raw_p2_obs.csv')
        adata_raw_p2.obs['leiden'] = pd.Categorical(
            obs['leiden'].values.astype(str), 
            categories=np.sort(np.unique(obs['leiden'].values.astype(str)))
        )

# %% [code]
if bool_recomp:
    ir.tl.chain_pairing(adata_raw_p1)
    ir.tl.chain_pairing(adata_raw_p2)

# %% [markdown]
#### P1

# %% [code]
if bool_recomp:
    sc.pl.umap(adata_raw_p1, color=["sample_rna"])
    sc.pl.umap(adata_raw_p1, color=["condition"])
    sc.pl.umap(adata_raw_p1, color=["leiden"])
    sc.pl.umap(adata_raw_p1, color=["n_counts", "has_tcr", "multi_chain"])
    sc.pl.umap(adata_raw_p1, color=["CD8A", "CD3E", "IFNG"], use_raw=False)

# %% [code]
if bool_recomp:
    fig, ax = plt.subplots(1,1,figsize=(10,4))
    sb.violinplot(
        data=adata_raw_p1.obs,
        y="n_counts",
        x="leiden",
        ax=ax
    )
    sc.pl.dotplot(
        adata_raw_p1, 
        var_names=[
            "CD4", "CD8A", "CD8B", "CD3D", "CD3E", "CD3G", "IFNG"
        ], 
        groupby="leiden", 
        use_raw=False,
    )

# %% [code]
if bool_recomp:
    if user == "david":
        adata_raw_p1 = adata_raw_p1[[
            x not in ["39"]
            for x in adata_raw_p1.obs["leiden"].values
        ]].copy()
    else:
        adata_raw_p1 = adata_raw_p1[[
            x not in ["28"]
            for x in adata_raw_p1.obs["leiden"].values
        ]].copy()
    sc.pl.umap(adata_raw_p1, color=["n_counts", "multi_chain"])

# %% [markdown]
#### P2

# %% [code]
if bool_recomp:
    sc.pl.umap(adata_raw_p2, color=["sample_rna"])
    sc.pl.umap(adata_raw_p2, color=["condition"])
    sc.pl.umap(adata_raw_p2, color=["leiden"])
    sc.pl.umap(adata_raw_p2, color=["n_counts", "has_tcr", "multi_chain"])
    sc.pl.umap(adata_raw_p2, color=["CD8A", "CD3E", "IFNG"], use_raw=False)

# %% [code]
if bool_recomp:
    sc.pl.dotplot(
        adata_raw_p2, 
        var_names=[
            "CD4", "CD8A", "CD8B", "CD3D", "CD3E", "CD3G", "IFNG"
        ], 
        groupby="leiden", 
        use_raw=False,
    )

# %% [code]
if bool_recomp:
    if user == "david":
        adata_raw_p2 = adata_raw_p2[[
            x not in ["29", "35", "37"]
            for x in adata_raw_p2.obs["leiden"].values
        ]].copy()
    else:
        adata_raw_p2 = adata_raw_p2[[
            x not in ["34", "38"]
            for x in adata_raw_p2.obs["leiden"].values
        ]].copy()
    sc.pl.umap(adata_raw_p2, color=["n_counts", "multi_chain"])

# %% [markdown]
### TCR-based

# %% [code]
adata_raw_p1.obs.groupby(["has_tcr", "multi_chain"]).size()

# %% [code]
adata_raw_p2.obs.groupby(["has_tcr", "multi_chain"]).size()

# %% [code]
# Only keep cells with reconstructed TCR
adata_raw_p1 = adata_raw_p1[adata_raw_p1.obs["has_tcr"].values == "True", :].copy()
adata_raw_p2 = adata_raw_p2[adata_raw_p2.obs["has_tcr"].values == "True", :].copy()

# %% [markdown]
## Unsupervised analysis

# %% [markdown]
### Compute anlyses

# %% [code]
if bool_recomp:
    adata_raw = adata_raw_p1.concatenate(adata_raw_p2)
    adata_raw.obs_names = ["-".join(x.split("-")[:-1]) for x in adata_raw.obs_names]
    adata_proc = adata_raw.raw.to_adata()
    adata_proc.raw = adata_raw.raw.to_adata()
    adata_proc_p1 = adata_raw_p1.raw.to_adata()
    adata_proc_p1.raw = adata_raw_p1.raw.to_adata()
    adata_proc_p2 = adata_raw_p2.raw.to_adata()
    adata_proc_p2.raw = adata_raw_p2.raw.to_adata()
    
    adata_proc_p1_cor = adata_raw_p1.raw.to_adata()
    adata_proc_p1_cor.raw = adata_raw_p1.raw.to_adata()
    adata_proc_p2_cor = adata_raw_p2.raw.to_adata()
    adata_proc_p2_cor.raw = adata_raw_p2.raw.to_adata()
    
    adata_proc_p1_unstim = adata_raw_p1[adata_raw_p1.obs["condition"].values == "unstimulated"].raw.to_adata()
    adata_proc_p1_unstim.raw = adata_raw_p1[adata_raw_p1.obs["condition"].values == "unstimulated"].raw.to_adata()
    adata_proc_p2_unstim = adata_raw_p2[adata_raw_p2.obs["condition"].values == "unstimulated"].raw.to_adata() 
    adata_proc_p2_unstim.raw = adata_raw_p2[adata_raw_p2.obs["condition"].values == "unstimulated"].raw.to_adata()
    
    adata_proc_p1_stim = adata_raw_p1[adata_raw_p1.obs["condition"].values == "stimulated"].raw.to_adata()
    adata_proc_p1_stim.raw = adata_raw_p1[adata_raw_p1.obs["condition"].values == "stimulated"].raw.to_adata()
    adata_proc_p2_stim = adata_raw_p2[adata_raw_p2.obs["condition"].values == "stimulated"].raw.to_adata() 
    adata_proc_p2_stim.raw = adata_raw_p2[adata_raw_p2.obs["condition"].values == "stimulated"].raw.to_adata()

# %% [markdown]
Full uncorrected:

# %% [code]
if bool_recomp:
    sc.pp.filter_genes(adata_proc, min_cells=10)
    sc.pp.normalize_per_cell(adata_proc, counts_per_cell_after=1000)
    sc.pp.log1p(adata_proc)
    #sc.pp.highly_variable_genes(adata_proc)
    sc.pp.pca(adata_proc)
    sc.pp.neighbors(adata_proc, knn=50)
    sc.tl.umap(adata_proc)
    if bool_recluster == True:
        sc.tl.leiden(adata_proc, resolution=1., random_state=0)
        pd.DataFrame(adata_proc.obs).to_csv(path_or_buf=dir_anndata+"adata_proc_obs.csv")
    else:
        obs = pd.read_csv(dir_anndata+'adata_proc_obs.csv')
        adata_proc.obs['leiden'] = pd.Categorical(
            obs['leiden'].values.astype(str), 
            categories=np.sort(np.unique(obs['leiden'].values.astype(str)))
        )

    
    sc.pp.filter_genes(adata_proc_p1, min_cells=10)
    sc.pp.normalize_per_cell(adata_proc_p1, counts_per_cell_after=1000)
    sc.pp.log1p(adata_proc_p1)
    #sc.pp.highly_variable_genes(adata_proc_p1)
    sc.pp.pca(adata_proc_p1)
    sc.pp.neighbors(adata_proc_p1, knn=50)
    sc.tl.umap(adata_proc_p1)
    if bool_recluster == True:
        sc.tl.leiden(adata_proc_p1, resolution=3., random_state=0)
        pd.DataFrame(adata_proc_p1.obs).to_csv(path_or_buf=dir_anndata+"adata_proc_p1_obs.csv")
    else:
        obs = pd.read_csv(dir_anndata+'adata_proc_p1_obs.csv')
        adata_proc_p1.obs['leiden'] = pd.Categorical(
            obs['leiden'].values.astype(str), 
            categories=np.sort(np.unique(obs['leiden'].values.astype(str)))
        )
    
    sc.pp.filter_genes(adata_proc_p2, min_cells=10)
    sc.pp.normalize_per_cell(adata_proc_p2, counts_per_cell_after=1000)
    sc.pp.log1p(adata_proc_p2)
    #sc.pp.highly_variable_genes(adata_proc_p2)
    sc.pp.pca(adata_proc_p2)
    sc.pp.neighbors(adata_proc_p2, knn=50)
    sc.tl.umap(adata_proc_p2)
    if bool_recluster == True:
        sc.tl.leiden(adata_proc_p2, resolution=3., random_state=0)
        pd.DataFrame(adata_proc_p2.obs).to_csv(path_or_buf=dir_anndata+"adata_proc_p2_obs.csv")
    else:
        obs = pd.read_csv(dir_anndata+'adata_proc_p2_obs.csv')
        adata_proc_p2.obs['leiden'] = pd.Categorical(
            obs['leiden'].values.astype(str), 
            categories=np.sort(np.unique(obs['leiden'].values.astype(str)))
        )
else:
    adata_proc = sc.read(dir_anndata+'adata_proc.h5ad') 
    adata_proc_p1 = sc.read(dir_anndata+'adata_proc_p1.h5ad') 
    adata_proc_p2 = sc.read(dir_anndata+'adata_proc_p2.h5ad') 

# %% [markdown]
Full corrected:

# %% [code]
if bool_recomp:
    sc.pp.filter_genes(adata_proc_p1_cor, min_cells=10)
    sc.pp.normalize_per_cell(adata_proc_p1_cor, counts_per_cell_after=1000)
    sc.pp.log1p(adata_proc_p1_cor)
    #sc.pp.highly_variable_genes(adata_proc)
    sc.pp.pca(adata_proc_p1_cor)
    sc.external.pp.bbknn(adata_proc_p1_cor, batch_key="condition")
    sc.tl.umap(adata_proc_p1_cor)
    if bool_recluster == True:
        sc.tl.leiden(adata_proc_p1_cor, resolution=3., random_state=0)
        pd.DataFrame(adata_proc_p1_cor.obs).to_csv(path_or_buf=dir_anndata+"adata_proc_p1_cor_obs.csv")
    else:
        obs = pd.read_csv(dir_anndata+'adata_proc_p1_cor_obs.csv')
        adata_proc_p1_cor.obs['leiden'] = pd.Categorical(
            obs['leiden'].values.astype(str), 
            categories=np.sort(np.unique(obs['leiden'].values.astype(str)))
        )
    
    sc.pp.filter_genes(adata_proc_p2_cor, min_cells=10)
    sc.pp.normalize_per_cell(adata_proc_p2_cor, counts_per_cell_after=1000)
    sc.pp.log1p(adata_proc_p2_cor)
    #sc.pp.highly_variable_genes(adata_proc)
    sc.pp.pca(adata_proc_p2_cor)
    sc.external.pp.bbknn(adata_proc_p2_cor, batch_key="condition")
    sc.tl.umap(adata_proc_p2_cor)
    if bool_recluster == True:
        sc.tl.leiden(adata_proc_p2_cor, resolution=3., random_state=0)
        pd.DataFrame(adata_proc_p2_cor.obs).to_csv(path_or_buf=dir_anndata+"adata_proc_p2_cor_obs.csv")
    else:
        obs = pd.read_csv(dir_anndata+'adata_proc_p2_cor_obs.csv')
        adata_proc_p2_cor.obs['leiden'] = pd.Categorical(
            obs['leiden'].values.astype(str), 
            categories=np.sort(np.unique(obs['leiden'].values.astype(str)))
        )
else:
    adata_proc_p1_cor = sc.read(dir_anndata+'adata_proc_p1_cor.h5ad') 
    adata_proc_p2_cor = sc.read(dir_anndata+'adata_proc_p2_cor.h5ad') 

# %% [markdown]
Unstimulated only:

# %% [code]
if bool_recomp:    
    sc.pp.filter_genes(adata_proc_p1_unstim, min_cells=10)
    sc.pp.normalize_per_cell(adata_proc_p1_unstim, counts_per_cell_after=1000)
    sc.pp.log1p(adata_proc_p1_unstim)
    #sc.pp.highly_variable_genes(adata_proc_p1_unstim)
    sc.pp.pca(adata_proc_p1_unstim)
    sc.pp.neighbors(adata_proc_p1_unstim, knn=50)
    sc.tl.umap(adata_proc_p1_unstim)
    if bool_recluster == True:
        sc.tl.leiden(adata_proc_p1_unstim, resolution=3., random_state=0)
        pd.DataFrame(adata_proc_p1_unstim.obs).to_csv(path_or_buf=dir_anndata+"adata_proc_p1_unstim_obs.csv")
    else:
        obs = pd.read_csv(dir_anndata+'adata_proc_p1_unstim_obs.csv')
        adata_proc_p1_unstim.obs['leiden'] = pd.Categorical(
            obs['leiden'].values.astype(str), 
            categories=np.sort(np.unique(obs['leiden'].values.astype(str)))
        )
    
    sc.pp.filter_genes(adata_proc_p2_unstim, min_cells=10)
    sc.pp.normalize_per_cell(adata_proc_p2_unstim, counts_per_cell_after=1000)
    sc.pp.log1p(adata_proc_p2_unstim)
    #sc.pp.highly_variable_genes(adata_proc_p2_unstim)
    sc.pp.pca(adata_proc_p2_unstim)
    sc.pp.neighbors(adata_proc_p2_unstim, knn=50)
    sc.tl.umap(adata_proc_p2_unstim)
    if bool_recluster == True:
        sc.tl.leiden(adata_proc_p2_unstim, resolution=3., random_state=0)
        pd.DataFrame(adata_proc_p2_unstim.obs).to_csv(path_or_buf=dir_anndata+"adata_proc_p2_unstim_obs.csv")
    else:
        obs = pd.read_csv(dir_anndata+'adata_proc_p2_unstim_obs.csv')
        adata_proc_p2_unstim.obs['leiden'] = pd.Categorical(
            obs['leiden'].values.astype(str), 
            categories=np.sort(np.unique(obs['leiden'].values.astype(str)))
        )
else:
    adata_proc_p1_unstim = sc.read(dir_anndata+'adata_proc_p1_unstim.h5ad') 
    adata_proc_p2_unstim = sc.read(dir_anndata+'adata_proc_p2_unstim.h5ad') 

# %% [markdown]
Stimulated only:

# %% [code]
if bool_recomp:    
    sc.pp.filter_genes(adata_proc_p1_stim, min_cells=10)
    sc.pp.normalize_per_cell(adata_proc_p1_stim, counts_per_cell_after=1000)
    sc.pp.log1p(adata_proc_p1_stim)
    #sc.pp.highly_variable_genes(adata_proc_p1_stim)
    sc.pp.pca(adata_proc_p1_stim)
    sc.pp.neighbors(adata_proc_p1_stim, knn=50)
    sc.tl.umap(adata_proc_p1_stim)
    if bool_recluster == True:
        sc.tl.leiden(adata_proc_p1_stim, resolution=3., random_state=0)
        pd.DataFrame(adata_proc_p1_stim.obs).to_csv(path_or_buf=dir_anndata+"adata_proc_p1_stim_obs.csv")
    else:
        obs = pd.read_csv(dir_anndata+'adata_proc_p1_stim_obs.csv')
        adata_proc_p1_stim.obs['leiden'] = pd.Categorical(
            obs['leiden'].values.astype(str), 
            categories=np.sort(np.unique(obs['leiden'].values.astype(str)))
        )
    
    sc.pp.filter_genes(adata_proc_p2_stim, min_cells=10)
    sc.pp.normalize_per_cell(adata_proc_p2_stim, counts_per_cell_after=1000)
    sc.pp.log1p(adata_proc_p2_stim)
    #sc.pp.highly_variable_genes(adata_proc_p2_stim)
    sc.pp.pca(adata_proc_p2_stim)
    sc.pp.neighbors(adata_proc_p2_stim, knn=50)
    sc.tl.umap(adata_proc_p2_stim)
    if bool_recluster == True:
        sc.tl.leiden(adata_proc_p2_stim, resolution=3., random_state=0)
        pd.DataFrame(adata_proc_p2_stim.obs).to_csv(path_or_buf=dir_anndata+"adata_proc_p2_stim_obs.csv")
    else:
        obs = pd.read_csv(dir_anndata+'adata_proc_p2_stim_obs.csv')
        adata_proc_p2_stim.obs['leiden'] = pd.Categorical(
            obs['leiden'].values.astype(str), 
            categories=np.sort(np.unique(obs['leiden'].values.astype(str)))
        )
else:
    adata_proc_p1_stim = sc.read(dir_anndata+'adata_proc_p1_stim.h5ad') 
    adata_proc_p2_stim = sc.read(dir_anndata+'adata_proc_p2_stim.h5ad') 

# %% [markdown]
### Full data set

# %% [markdown]
#### Overview full dataset

# %% [code]
sc.pl.umap(adata_proc, color=["sample_rna"])
sc.pl.umap(adata_proc, color=["condition"])
sc.pl.umap(adata_proc, color=["patient"])
sc.pl.umap(adata_proc, color=["n_counts"])

# %% [code]
sc.pl.umap(adata_proc, color=["CD3D", "CD3E", "CD3G"])
sc.pl.umap(adata_proc, color=["CD4", "CD8A", "CD8B"])
sc.pl.umap(adata_proc, color=["GZMA", "GZMB"])
sc.pl.umap(adata_proc, color=["IFNG"])

# %% [markdown]
### P1

# %% [markdown]
#### Unstimulated only

# %% [code]
sc.pl.umap(adata_proc_p1_unstim, color=["sample_rna"])
sc.pl.umap(adata_proc_p1_unstim, color=["leiden"])
sc.pl.umap(adata_proc_p1_unstim, color=["n_counts"])

# %% [code]
sc.pl.umap(adata_proc_p1_unstim, color=["CD3D", "CD3E", "CD3G"], use_raw=False)
sc.pl.umap(adata_proc_p1_unstim, color=["CD4", "CD8A", "CD8B"], use_raw=False)

# %% [markdown]
#### Full corrected

# %% [code]
sc.pl.umap(adata_proc_p1_cor, color=["sample_rna"])
sc.pl.umap(adata_proc_p1_cor, color=["condition"])
sc.pl.umap(adata_proc_p1_cor, color=["leiden"])
sc.pl.umap(adata_proc_p1_cor, color=["n_counts"])

# %% [code]
sc.pl.umap(adata_proc_p1_cor, color=["CD3D", "CD3E", "CD3G"], use_raw=False)
sc.pl.umap(adata_proc_p1_cor, color=["CD4", "CD8A", "CD8B"], use_raw=False)
sc.pl.umap(adata_proc_p1_cor, color=["IFNG"], use_raw=False)

# %% [markdown]
#### Full raw

# %% [code]
sc.pl.umap(adata_proc_p1, color=["sample_rna"])
sc.pl.umap(adata_proc_p1, color=["condition"])
sc.pl.umap(adata_proc_p1, color=["leiden"])
sc.pl.umap(adata_proc_p1, color=["n_counts"])

# %% [code]
sc.pl.umap(adata_proc_p1, color=["CD3D", "CD3E", "CD3G"], use_raw=False)
sc.pl.umap(adata_proc_p1, color=["CD4", "CD8A", "CD8B"], use_raw=False)

# %% [markdown]
### P2

# %% [markdown]
#### Unstimulated only

# %% [code]
sc.pl.umap(adata_proc_p2_unstim, color=["sample_rna"])
sc.pl.umap(adata_proc_p2_unstim, color=["leiden"])
sc.pl.umap(adata_proc_p2_unstim, color=["n_counts"])

# %% [code]
sc.pl.umap(adata_proc_p2_unstim, color=["CD3D", "CD3E", "CD3G"], use_raw=False)
sc.pl.umap(adata_proc_p2_unstim, color=["CD4", "CD8A", "CD8B"], use_raw=False)

# %% [markdown]
#### Full corrected

# %% [code]
sc.pl.umap(adata_proc_p2_cor, color=["sample_rna"])
sc.pl.umap(adata_proc_p2_cor, color=["condition"])
sc.pl.umap(adata_proc_p2_cor, color=["leiden"])
sc.pl.umap(adata_proc_p2_cor, color=["n_counts"])

# %% [code]
sc.pl.umap(adata_proc_p2_cor, color=["CD3D", "CD3E", "CD3G"], use_raw=False)
sc.pl.umap(adata_proc_p2_cor, color=["CD4", "CD8A", "CD8B"], use_raw=False)
sc.pl.umap(adata_proc_p2_cor, color=["IFNG"], use_raw=False)

# %% [markdown]
#### Full

# %% [code]
sc.pl.umap(adata_proc_p2, color=["sample_rna"])
sc.pl.umap(adata_proc_p2, color=["condition"])
sc.pl.umap(adata_proc_p2, color=["leiden"])
sc.pl.umap(adata_proc_p2, color=["n_counts"])

# %% [code]
sc.pl.umap(adata_proc_p2, color=["CD3D", "CD3E", "CD3G"], use_raw=False)
sc.pl.umap(adata_proc_p2, color=["CD4", "CD8A", "CD8B"], use_raw=False)

# %% [markdown]
## Carry over clustering into *proc

# %% [code]
adata_proc_p1.obs["leiden_corrected"] = adata_proc_p1_cor[adata_proc_p1.obs_names].obs["leiden"].values
adata_proc_p2.obs["leiden_corrected"] = adata_proc_p2_cor[adata_proc_p2.obs_names].obs["leiden"].values

# %% [code]
adata_proc_p1.obs["leiden_unstim"] = [
    adata_proc_p1_unstim[x].obs["leiden"].values[0]
    if x in adata_proc_p1_unstim.obs_names 
    else "none"
    for x in adata_proc_p1.obs_names
]
adata_proc_p2.obs["leiden_unstim"] = [
    adata_proc_p2_unstim[x].obs["leiden"].values[0]
    if x in adata_proc_p2_unstim.obs_names else "none"
    for x in adata_proc_p2.obs_names
]

# %% [markdown]
## Cell type annotation

# %% [markdown]
### Load signatures

# %% [code]
tcell_signatures = pd.read_table(
    dir_reference+"41467_2019_12464_MOESM7_ESM.csv",
    skiprows=1, header=0, sep=";"
).iloc[:, 3:]
tcell_signatures = {
    k: [x for x in v if x in adata_proc_p1.var_names]
    for k, v in tcell_signatures.items()
}

# %% [code]
rs = de.enrich.RefSets(fn=dir_genesets+"c5.bp.v5.2.symbols.gmt")

# %% [code]
[x.id for x in rs.sets if "t_cell_receptor" in x.id.lower()]

# %% [code]
tcr_signalling = [x for x in rs.sets if x.id == "GO_T_CELL_RECEPTOR_SIGNALING_PATHWAY"][0]
tcr_signalling_genes = tcr_signalling.genes
print("%i: %s" % (len(tcr_signalling_genes), tcr_signalling_genes))

# %% [markdown]
### Cd4 / Cd8

# %% [code]
adata_proc_p1_cor.obs["CD8_all_genes"] = np.sum(np.asarray(adata_proc_p1_cor[:, ["CD8A", "CD8B"]].X.todense()), axis=1)
adata_proc_p2_cor.obs["CD8_all_genes"] = np.sum(np.asarray(adata_proc_p2_cor[:, ["CD8A", "CD8B", "CD8B2"]].X.todense()), axis=1)

# %% [code]
sc.pl.umap(adata_proc_p1_cor, color=["leiden", "CD8_all_genes"])
sc.pl.umap(adata_proc_p2_cor, color=["leiden", "CD8_all_genes"])

# %% [code]
sc.pl.violin(
    adata_proc_p1_cor, 
    keys=["CD8_all_genes"], 
    groupby="leiden", 
    use_raw=False,
    rotation=90
)
sc.pl.violin(
    adata_proc_p2_cor, 
    keys=["CD8_all_genes"], 
    groupby="leiden", 
    use_raw=False,
    rotation=90
)

# %% [code]
cluster_map = dict([
    (x, ["Cd4", "Cd8"][int(adata_proc_p1_cor.obs.loc[adata_proc_p1_cor.obs["leiden"].values == x, "CD8_all_genes"].values.mean() > 0.5)])
    for x in np.unique(adata_proc_p1_cor.obs["leiden"].values)
])
adata_proc_p1_cor.obs["cell_type"] = [
    cluster_map[x] for x in adata_proc_p1_cor.obs["leiden"].values
]

# %% [code]
sc.pl.umap(adata_proc_p1_cor, color=["cell_type", "CD8_all_genes", "CD8A", "CD8B"])

# %% [code]
cluster_map = dict([
    (x, ["Cd4", "Cd8"][int(adata_proc_p2_cor.obs.loc[adata_proc_p2_cor.obs["leiden"].values == x, "CD8_all_genes"].values.mean() > 0.5)])
    for x in np.unique(adata_proc_p2_cor.obs["leiden"].values)
])
adata_proc_p2_cor.obs["cell_type"] = [
    cluster_map[x] for x in adata_proc_p2_cor.obs["leiden"].values
]

# %% [code]
sc.pl.umap(adata_proc_p2_cor, color=["cell_type", "CD8_all_genes", "CD8A", "CD8B"])

# %% [code]
adata_proc_p1.obs["cell_type"] = adata_proc_p1_cor[adata_proc_p1.obs_names].obs["cell_type"].values
adata_proc_p2.obs["cell_type"] = adata_proc_p2_cor[adata_proc_p2.obs_names].obs["cell_type"].values

# %% [code]
adata_proc_p1.obs["CD8_all_genes"] = np.sum(np.asarray(adata_proc_p1[:, ["CD8A", "CD8B"]].X.todense()), axis=1)
adata_proc_p2.obs["CD8_all_genes"] = np.sum(np.asarray(adata_proc_p2[:, ["CD8A", "CD8B", "CD8B2"]].X.todense()), axis=1)

# %% [code]
sc.pl.umap(adata_proc_p1, color=["cell_type", "CD8_all_genes", "CD8A", "CD8B"])

# %% [code]
sc.pl.umap(adata_proc_p2, color=["cell_type", "CD8_all_genes", "CD8A", "CD8B"])

# %% [code]
adata_proc_p1_unstim.obs["cell_type"] = adata_proc_p1[adata_proc_p1_unstim.obs_names].obs["cell_type"].values
adata_proc_p2_unstim.obs["cell_type"] = adata_proc_p2[adata_proc_p2_unstim.obs_names].obs["cell_type"].values

# %% [markdown]
### Score cells based on signatures

# %% [code]
adata_proc_p1.obs["tcr_signalling_score"] = np.mean(
    (
        adata_proc_p1[:, [x for x in tcr_signalling_genes if x in adata_proc_p1.var_names]].X - 
        adata_proc_p1[:, [x for x in tcr_signalling_genes if x in adata_proc_p1.var_names]].X.mean(axis=0)
    ) / adata_proc_p1[:, [x for x in tcr_signalling_genes if x in adata_proc_p1.var_names]].X.todense().std(axis=0),
    axis=1
)
for k, v in tcell_signatures.items():
    adata_proc_p1.obs[k.lower() + "_score"] = np.mean(
        (
            adata_proc_p1[:, [x for x in v if x in adata_proc_p1.var_names]].X - 
            adata_proc_p1[:, [x for x in v if x in adata_proc_p1.var_names]].X.mean(axis=0)
        ) / adata_proc_p1[:, [x for x in v if x in adata_proc_p1.var_names]].X.todense().std(axis=0),
        axis=1
    )

# %% [code]
adata_proc_p2.obs["tcr_signalling_score"] = np.mean(
    (
        adata_proc_p2[:, [x for x in tcr_signalling_genes if x in adata_proc_p2.var_names]].X - 
        adata_proc_p2[:, [x for x in tcr_signalling_genes if x in adata_proc_p2.var_names]].X.mean(axis=0)
    ) / adata_proc_p2[:, [x for x in tcr_signalling_genes if x in adata_proc_p2.var_names]].X.todense().std(axis=0),
    axis=1
)
for k, v in tcell_signatures.items():
    adata_proc_p2.obs[k.lower() + "_score"] = np.mean(
        (
            adata_proc_p2[:, [x for x in v if x in adata_proc_p2.var_names]].X - 
            adata_proc_p2[:, [x for x in v if x in adata_proc_p2.var_names]].X.mean(axis=0)
        ) / adata_proc_p2[:, [x for x in v if x in adata_proc_p2.var_names]].X.todense().std(axis=0),
        axis=1
    )

# %% [markdown]
Tissue residency score:

# %% [code]
residence_score_gene_list = [
    "CA10", "CRTAM", "CX3CR1", "CXCL13", "CXCR6", "DUSP6", 
    "FAM65B", "IL2", "IL10", "IL23R", "ITGA1", "ITGAE", 
    "KCNK5", "KLF2", "KLF3", "KRT72", "KRT73", "NPDC1", 
    "PDCD1", "PTGDS", "RAP1GAP2", "RGS1", "S1PR1", "SBK1", 
    "SELL", "SOX13", "STK38", "TSPAN18", "TTC16", "TTYH2"
]
print(len(residence_score_gene_list))
residence_score_gene_list_filtered = [
    x for x in residence_score_gene_list
    if x in adata_proc.var_names
]
print(len(residence_score_gene_list_filtered))

adata_proc_p1.obs["tissue_residency_score"] = np.mean(
    (
        adata_proc_p1[:, [x for x in residence_score_gene_list if x in adata_proc_p1.var_names]].X - 
        adata_proc_p1[:, [x for x in residence_score_gene_list if x in adata_proc_p1.var_names]].X.mean(axis=0)
    ) / adata_proc_p1[:, [x for x in residence_score_gene_list if x in adata_proc_p1.var_names]].X.todense().std(axis=0),
    axis=1
)
adata_proc_p2.obs["tissue_residency_score"] = np.mean(
    (
        adata_proc_p2[:, [x for x in residence_score_gene_list if x in adata_proc_p2.var_names]].X - 
        adata_proc_p2[:, [x for x in residence_score_gene_list if x in adata_proc_p2.var_names]].X.mean(axis=0)
    ) / adata_proc_p2[:, [x for x in residence_score_gene_list if x in adata_proc_p2.var_names]].X.todense().std(axis=0),
    axis=1
)

# %% [markdown]
### Plot

# %% [code]
fig, ax = plt.subplots(figsize=(5,5))
sc.pl.umap(
    adata_proc_p1, 
    ax=ax, show=False
)
sc.pl.umap(
    adata_proc_p1[adata_proc_p1.obs["condition"].values == "unstimulated"], 
    color=["cell_type"], 
    ax=ax, show=False
)
plt.show()

# %% [code]
fig, ax = plt.subplots(figsize=(5,5))
sc.pl.umap(
    adata_proc_p2, 
    ax=ax, show=False
)
sc.pl.umap(
    adata_proc_p2[adata_proc_p2.obs["condition"].values == "unstimulated"], 
    color=["cell_type"], 
    ax=ax, show=False
)
plt.show()

# %% [markdown]
### Scores on raw data umap

# %% [code]
score_to_plot = [k.lower() + "_score" for k in tcell_signatures.keys()] + ["tissue_residency_score"] + \
    ["IFNG", "NME1", "IFIT1"]
fig, ax = plt.subplots(nrows=1, ncols=len(score_to_plot), figsize=(len(score_to_plot)*4,4))
for i, x in enumerate(score_to_plot):
    sc.pl.umap(
        adata_proc_p1, 
        color=x, 
        ax=ax[i], show=False
    )
plt.tight_layout()
plt.savefig(dir_panels+"umap_p1_all.pdf")
plt.close()

# %% [code]
score_to_plot = [k.lower() + "_score" for k in tcell_signatures.keys()] + ["tissue_residency_score"] + \
    ["IFNG", "NME1", "IFIT1"]
fig, ax = plt.subplots(nrows=1, ncols=len(score_to_plot), figsize=(len(score_to_plot)*4,4))
for i, x in enumerate(score_to_plot):
    sc.pl.umap(
        adata_proc_p2, 
        color=x, 
        ax=ax[i], show=False
    )
plt.tight_layout()
plt.close()

# %% [code]
score_to_plot = ["IFNG"] + [k.lower() + "_score" for k in tcell_signatures.keys()]
fig, ax = plt.subplots(nrows=5, ncols=2, figsize=(4*2, 4*5))
for i, x in enumerate(score_to_plot):
    sc.pl.umap(
        adata_proc_p1, 
        ax=ax[i, 0], show=False
    )
    sc.pl.umap(
        adata_proc_p1[adata_proc_p1.obs["condition"].values == "unstimulated"], 
        color=x, 
        vmin=np.min(adata_proc_p1.obs[x].values) if x in adata_proc_p1.obs.columns else np.min(adata_proc_p1[:, x].X),
        vmax=np.max(adata_proc_p1.obs[x].values) if x in adata_proc_p1.obs.columns else np.max(adata_proc_p1[:, x].X),
        use_raw=False, ax=ax[i, 0], show=False
    )
    sc.pl.umap(
        adata_proc_p1, 
        ax=ax[i, 1], show=False
    )
    sc.pl.umap(
        adata_proc_p1[adata_proc_p1.obs["condition"].values == "stimulated"], 
        color=x, 
        vmin=np.min(adata_proc_p1.obs[x].values) if x in adata_proc_p1.obs.columns else np.min(adata_proc_p1[:, x].X),
        vmax=np.max(adata_proc_p1.obs[x].values) if x in adata_proc_p1.obs.columns else np.max(adata_proc_p1[:, x].X),
        use_raw=False, ax=ax[i, 1], show=False
    )
    if i == 0:
        ax[i, 0].set_title("unstimulated")
        ax[i, 1].set_title("stimulated")
    else:
        ax[i, 0].set_title(" ")
        ax[i, 1].set_title(" ")
    ax[i, 0].set_xlabel(" ")
    ax[i, 1].set_xlabel(" ")
    ax[i, 0].set_ylabel(" ")
    ax[i, 1].set_ylabel(" ")
plt.subplots_adjust(wspace=0.1, hspace=0.05)
plt.savefig(dir_panels + "fig1c.pdf")
plt.show()

# %% [code]
score_to_plot = ["tissue_residency_score"]
fig, ax = plt.subplots(ncols=2, nrows=len(score_to_plot), figsize=(2*4, len(score_to_plot)*4))
ax = np.expand_dims(ax, axis=0)
for i, x in enumerate(score_to_plot):
    sc.pl.umap(
        adata_proc_p1, 
        ax=ax[i, 0], show=False
    )
    sc.pl.umap(
        adata_proc_p1[adata_proc_p1.obs["condition"].values == "unstimulated"], 
        color=x,   
        vmin=np.min(adata_proc_p1.obs[x].values) if x in adata_proc_p1.obs.columns else np.min(adata_proc_p1[:, x].X),
        vmax=np.max(adata_proc_p1.obs[x].values) if x in adata_proc_p1.obs.columns else np.max(adata_proc_p1[:, x].X),
        use_raw=False, ax=ax[i, 0], show=False
    )
    
    sc.pl.umap(
        adata_proc_p1, 
        ax=ax[i, 1], show=False
    )
    sc.pl.umap(
        adata_proc_p1[adata_proc_p1.obs["condition"].values == "stimulated"], 
        color=x,   
        vmin=np.min(adata_proc_p1.obs[x].values) if x in adata_proc_p1.obs.columns else np.min(adata_proc_p1[:, x].X),
        vmax=np.max(adata_proc_p1.obs[x].values) if x in adata_proc_p1.obs.columns else np.max(adata_proc_p1[:, x].X),
        use_raw=False, ax=ax[i, 1], show=False
    )
    if i == 0:
        ax[i, 0].set_title("unstimulated")
        ax[i, 1].set_title("stimulated")
    else:
        ax[i, 0].set_title(" ")
        ax[i, 1].set_title(" ")
    ax[i, 0].set_xlabel(" ")
    ax[i, 1].set_xlabel(" ")
    ax[i, 0].set_ylabel(" ")
    ax[i, 1].set_ylabel(" ")
plt.tight_layout()
plt.subplots_adjust(wspace=0.1, hspace=0.05)
plt.savefig(dir_panels+"sfig6d.pdf")
plt.show()

# %% [code]
score_to_plot = ["IFNG"] + [k.lower() + "_score" for k in tcell_signatures.keys()]
fig, ax = plt.subplots(nrows=5, ncols=2, figsize=(4*2, 4*5))
for i, x in enumerate(score_to_plot):
    sc.pl.umap(
        adata_proc_p2, 
        ax=ax[i, 0], show=False
    )
    sc.pl.umap(
        adata_proc_p2[adata_proc_p2.obs["condition"].values == "unstimulated"], 
        color=x, 
        vmin=np.min(adata_proc_p2.obs[x].values) if x in adata_proc_p2.obs.columns else np.min(adata_proc_p2[:, x].X),
        vmax=np.max(adata_proc_p2.obs[x].values) if x in adata_proc_p2.obs.columns else np.max(adata_proc_p2[:, x].X),
        use_raw=False, ax=ax[i, 0], show=False
    )
    sc.pl.umap(
        adata_proc_p2, 
        ax=ax[i, 1], show=False
    )
    sc.pl.umap(
        adata_proc_p2[adata_proc_p2.obs["condition"].values == "stimulated"], 
        color=x, 
        vmin=np.min(adata_proc_p2.obs[x].values) if x in adata_proc_p2.obs.columns else np.min(adata_proc_p2[:, x].X),
        vmax=np.max(adata_proc_p2.obs[x].values) if x in adata_proc_p2.obs.columns else np.max(adata_proc_p2[:, x].X),
        use_raw=False, ax=ax[i, 1], show=False
    )
    if i == 0:
        ax[i, 0].set_title("unstimulated")
        ax[i, 1].set_title("stimulated")
    else:
        ax[i, 0].set_title(" ")
        ax[i, 1].set_title(" ")
    ax[i, 0].set_xlabel(" ")
    ax[i, 1].set_xlabel(" ")
    ax[i, 0].set_ylabel(" ")
    ax[i, 1].set_ylabel(" ")
plt.subplots_adjust(wspace=0.1, hspace=0.05)
plt.savefig(dir_panels + "sfig2b.pdf")
plt.show()

# %% [markdown]
### P1

# %% [code]
sc.pl.umap(
    adata_proc_p1, color="leiden",
)
fig, ax = plt.subplots(figsize=(20, 4))
sb.boxplot(
    y=np.asarray(adata_proc_p1[:, "IFNG"].X.todense()).flatten(), 
    x=adata_proc_p1.obs["leiden"].values, 
    ax=ax
)

# %% [code]
if user == "david":
    ifng_clusters = ["29"] #["30", "34"]
else:
    ifng_clusters = ["27"]
adata_proc_p1.uns["ifng_clusters"] = ifng_clusters
adata_proc_p1.obs["ifng_pos_cluster"] = [
    x in adata_proc_p1.uns["ifng_clusters"] 
    for x in adata_proc_p1.obs["leiden"].values
]

# %% [code]
fig, ax = plt.subplots(nrows=1, ncols=1, figsize=(5, 5))
sc.pl.umap(
    adata_proc_p1,
    ax=ax,
    show=False,
)
sc.pl.umap(
    adata_proc_p1[[
        x in adata_proc_p1.uns["ifng_clusters"] 
        for x in adata_proc_p1.obs["leiden"].values
    ]],
    size=30,
    color="IFNG",
    use_raw=False,
    ax=ax,
    show=False,
)
plt.tight_layout()
plt.show()

# %% [markdown]
#### Subcluster

# %% [code]
if bool_recomp:
    adata_temp = adata_proc_p1[adata_proc_p1.obs["ifng_pos_cluster"].values == True].copy()
    sc.pp.neighbors(adata_temp)
    sc.tl.leiden(adata_temp, resolution=0.5)
    adata_proc_p1.obs["leiden_subclustered"] = [
        "subclustered_" + str(adata_temp[x].obs["leiden"].values[0]) 
        if y == True else z
        for x, y, z in zip(
            adata_proc_p1.obs_names,
            adata_proc_p1.obs["ifng_pos_cluster"].values,
            adata_proc_p1.obs["leiden"].values
        )
    ]
    del adata_temp

# %% [code]
sc.pl.umap(adata_proc_p1, color="leiden_subclustered")

fig, ax = plt.subplots(nrows=1, ncols=1, figsize=(7, 5))
sc.pl.umap(
    adata_proc_p1,
    ax=ax,
    show=False,
)
sc.pl.umap(
    adata_proc_p1[adata_proc_p1.obs["ifng_pos_cluster"].values == True],
    size=30,
    color="leiden_subclustered",
    use_raw=False,
    ax=ax,
    show=False,
)
plt.tight_layout()
plt.show()

# %% [markdown]
### P2

# %% [code]
sc.pl.umap(
    adata_proc_p2, color="leiden_corrected",
)
fig, ax = plt.subplots(figsize=(20, 4))
sb.boxplot(
    y=np.asarray(adata_proc_p2[:, "IFNG"].X.todense()).flatten(), 
    x=adata_proc_p2.obs["leiden_corrected"].values, 
    ax=ax
)

# %% [code]
sc.pl.umap(
    adata_proc_p2, color="leiden",
)
fig, ax = plt.subplots(figsize=(20, 4))
sb.boxplot(
    y=np.asarray(adata_proc_p2[:, "IFNG"].X.todense()).flatten(), 
    x=adata_proc_p2.obs["leiden"].values, 
    ax=ax
)

# %% [code]
if user == "david":
    ifng_clusters = ["36"]
else:
    ifng_clusters = ["32"]
adata_proc_p2.uns["ifng_clusters"] = ifng_clusters
adata_proc_p2.obs["ifng_pos_cluster"] = [
    x in adata_proc_p2.uns["ifng_clusters"] 
    for x in adata_proc_p2.obs["leiden"].values
]

# %% [code]
fig, ax = plt.subplots(nrows=1, ncols=1, figsize=(5, 5))
sc.pl.umap(
    adata_proc_p2,
    ax=ax,
    show=False,
)
sc.pl.umap(
    adata_proc_p2[[
        x in adata_proc_p2.uns["ifng_clusters"] 
        for x in adata_proc_p2.obs["leiden"].values
    ]],
    size=20,
    color="IFNG",
    use_raw=False,
    ax=ax,
    show=False,
)
plt.tight_layout()
plt.show()

# %% [markdown]
### Plot both patients

# %% [code]
fig, ax = plt.subplots(nrows=2, ncols=3, figsize=(12, 8))
sc.pl.umap(
    adata_proc_p1,
    ax=ax[0, 0],
    show=False,
)
sc.pl.umap(
    adata_proc_p1[[
        x in adata_proc_p1.uns["ifng_clusters"] 
        for x in adata_proc_p1.obs["leiden"].values
    ]],
    size=30,
    color="leiden",
    palette=[sb.color_palette("colorblind")[0]],
    use_raw=False,
    ax=ax[0, 0],
    show=False,
)

sc.pl.umap(
    adata_proc_p2,
    ax=ax[0, 1],
    show=False,
)
sc.pl.umap(
    adata_proc_p2[[
        x in adata_proc_p2.uns["ifng_clusters"] 
        for x in adata_proc_p2.obs["leiden"].values
    ]],
    size=30,
    color="leiden",
    palette=[sb.color_palette("colorblind")[1]],
    use_raw=False,
    ax=ax[0, 1],
    show=False,
)

sc.pl.umap(
    adata_proc,
    ax=ax[0, 2],
    show=False,
)
sc.pl.umap(
    adata_proc[
        [
            adata_proc_p1.obs.loc[x, "leiden"] in adata_proc_p1.uns["ifng_clusters"]
            if x in adata_proc_p1.obs_names else False
            for x in adata_proc.obs_names
        ], :
    ],
    size=30,
    color="patient",
    palette=[sb.color_palette("colorblind")[0]],
    use_raw=False,
    ax=ax[0, 2],
    show=False,
)
sc.pl.umap(
    adata_proc[
        [
            adata_proc_p2.obs.loc[x, "leiden"] in adata_proc_p2.uns["ifng_clusters"] 
            if x in adata_proc_p2.obs_names else False
            for x in adata_proc.obs_names
        ], :
    ],
    size=30,
    color="patient",
    use_raw=False,
    palette=[sb.color_palette("colorblind")[1]],
    ax=ax[0, 2],
    show=False,
)

sc.pl.umap(
    adata_proc,
    color="patient",
    ax=ax[1, 0],
    show=False,
)
sc.pl.umap(
    adata_proc,
    color="condition",
    ax=ax[1, 1],
    show=False,
)
sc.pl.umap(
    adata_proc,
    color="IFNG",
    use_raw=False,
    ax=ax[1, 2],
    show=False,
)
ax[0, 0].set_title("GT_2")
ax[0, 1].set_title("GT_3")
ax[0, 2].set_title("GT_2 + GT_3")
ax[0, 0].set_xlabel(" ")
ax[0, 1].set_xlabel(" ")
ax[0, 2].set_xlabel(" ")
ax[0, 0].set_ylabel(" ")
ax[0, 1].set_ylabel(" ")
ax[0, 2].set_ylabel(" ")

ax[1, 0].set_xlabel(" ")
ax[1, 1].set_xlabel(" ")
ax[1, 2].set_xlabel(" ")
ax[1, 0].set_ylabel(" ")
ax[1, 1].set_ylabel(" ")
ax[1, 2].set_ylabel(" ")

ax[0, 0].legend_.remove()
ax[0, 1].legend_.remove()
ax[0, 2].legend_.remove()

ax[1, 0].legend_.remove()
ax[1, 1].legend_.remove()
#ax[1, 2].legend_.remove()

plt.subplots_adjust(wspace=0.1, hspace=0.05)
plt.tight_layout()
plt.savefig(dir_panels+"sfig2c.pdf")
plt.show()

# %% [markdown]
## Clear objects

# %% [code]
if bool_recomp:
    sc.write(dir_anndata+'adata_raw.h5ad', adata_raw)
    sc.write(dir_anndata+'adata_raw_p1.h5ad', adata_raw_p1)
    sc.write(dir_anndata+'adata_raw_p2.h5ad', adata_raw_p2)
    sc.write(dir_anndata+'adata_proc_p1_cor.h5ad', adata_proc_p1_cor)
    sc.write(dir_anndata+'adata_proc_p2_cor.h5ad', adata_proc_p2_cor)
    sc.write(dir_anndata+'adata_proc_p1_stim.h5ad', adata_proc_p1_unstim)
    sc.write(dir_anndata+'adata_proc_p2_stim.h5ad', adata_proc_p2_unstim)
    sc.write(dir_anndata+'adata_proc_p1_unstim.h5ad', adata_proc_p1_unstim)
    sc.write(dir_anndata+'adata_proc_p2_unstim.h5ad', adata_proc_p2_unstim)
    
del adata_raw
del adata_raw_p1
del adata_raw_p2

import gc
gc.collect()

# %% [markdown]
# TA processing

# %% [markdown]
## Cell filtering

# %% [markdown]
### Basic stats

# %% [code]
if bool_recomp:
    adata_raw_ta.obs['n_genes'] = np.sum(adata_raw_ta.X > 0, axis=1)
    adata_raw_ta.obs['n_counts'] = np.sum(adata_raw_ta.X, axis=1)
    mt_gene_mask = [gene.lower().startswith('mt-') for gene in adata_raw_ta.var_names]
    adata_raw_ta.obs['mt_frac'] = np.asarray(adata_raw_ta[:,mt_gene_mask].X.sum(axis=1)).flatten() / \
        adata_raw_ta.obs['n_counts'].values

# %% [code]
sc.pl.scatter(adata_raw_ta, 'n_counts', 'n_genes', color='mt_frac', size=5)
sc.pl.scatter(adata_raw_ta[adata_raw_ta.obs['n_genes']<1000], 'n_counts', 'n_genes', color='mt_frac', size=5)

# %% [code]
if bool_recomp:
    sc.pp.filter_cells(adata_raw_ta, min_genes=200)
    adata_raw_ta = adata_raw_ta[adata_raw_ta.obs["mt_frac"].values <= 0.2].copy()

# %% [markdown]
### Exploratory analysis

# %% [code]
if bool_recomp:
    sc.pp.filter_genes(adata_raw_ta, min_cells=10)
    sc.pp.normalize_per_cell(adata_raw_ta, counts_per_cell_after=1000)
    sc.pp.log1p(adata_raw_ta)
    #sc.pp.highly_variable_genes(adata_raw_ta, flavor="seurat")
    sc.pp.pca(adata_raw_ta)
    sc.external.pp.bbknn(adata_raw_ta, batch_key="concatenation")
    sc.tl.umap(adata_raw_ta)
    if bool_recluster == True:
        sc.tl.leiden(adata_raw_ta, resolution=1., random_state=0)
        pd.DataFrame(adata_raw_ta.obs).to_csv(path_or_buf=dir_anndata+"adata_raw_ta_obs.csv")
    else:
        obs = pd.read_csv(dir_anndata+'adata_raw_ta_obs.csv')
        adata_raw_ta.obs['leiden'] = pd.Categorical(
            obs['leiden'].values.astype(str), 
            categories=np.sort(np.unique(obs['leiden'].values.astype(str)))
        )

# %% [code]
sc.pl.umap(adata_raw_ta, color=["leiden"])
sc.pl.umap(adata_raw_ta, color=["CD3D", "CD3E", "CD3G"])
sc.pl.umap(adata_raw_ta, color=["CD4", "CD8A", "CD8B", "CD68"])

# %% [code]
sc.pl.dotplot(
    adata_raw_ta, groupby="leiden", 
    var_names=["CD3D", "CD3E", "CD3G", "CD4", "CD8A", "CD8B", "CD68", "TRAC"]
)

# %% [code]
if bool_recomp:
    if user == "david":
        tcell_cluster = ["6"] #["7"]
    else:
        assert False
    # Look for cells in right cluster that have either CD3 or CD4 / CD8 expressed
    adata_raw_ta.obs["cell_type_new"] = [
        "Cd8" if y in tcell_cluster and adata_raw_ta[x, ["CD3D", "CD3E", "CD3G"]].X.sum() > 0 and adata_raw_ta[x, ["CD8A", "CD8B"]].X.sum() > 0
        else "Cd4" if y in tcell_cluster and adata_raw_ta[x, ["CD3D", "CD3E", "CD3G"]].X.sum() > 0
        else "other"
        for x, y in zip(
            adata_raw_ta.obs_names,
            adata_raw_ta.obs["leiden"].values
        )
    ]

# %% [code]
sc.pl.dotplot(adata_raw_ta, groupby="cell_type_new", var_names=["CD3D", "CD3E", "CD3G", "CD4", "CD8A", "CD8B"])

# %% [code]
sc.pl.umap(adata_raw_ta, color="cell_type_new")

# %% [code]
if bool_recomp:
    if False:
        adata_proc_ta_tcells = adata_raw_ta[[x in ["Cd4", "Cd8"] for x in adata_raw_ta.obs["cell_type_new"]]].copy()
        adata_proc_ta_tcells["cell_type"] = adata_proc_ta_tcells["cell_type_new"].values
    else:  # predefined T cell selection
        #adata_raw_ta.obs_names = [
        #    "-".join(x.split("-")[2:]) + "_" + x.split("-")[0]
        #    for x in adata_raw_ta.obs_names
        #]
        #adata_raw_ta.obs_names = [
        #    x.upper()
        #    for x in adata_raw_ta.obs_names
        #]
        #adata_p1_withta.obs_names = [
        #    x.upper()
        #    for x in adata_p1_withta.obs_names
        #]
        adata_proc_ta_tcells = adata_p1_withta[
            adata_p1_withta.obs["grouping"].values == "Covid-19"
        ].copy()
    sc.pp.highly_variable_genes(adata_proc_ta_tcells)
    sc.pp.pca(adata_proc_ta_tcells)
    sc.pp.neighbors(adata_proc_ta_tcells)
    sc.tl.umap(adata_proc_ta_tcells)

# %% [code]
# Subcluster Cd4 and Cd8 separately:
if bool_recomp:
    adata_temp = adata_proc_ta_tcells[
        adata_proc_ta_tcells.obs["cell_type"] == "Cd4"
    ].copy()
    sc.pp.highly_variable_genes(adata_temp)
    sc.pp.pca(adata_temp)
    sc.pp.neighbors(adata_temp)
    if bool_recluster == True or True:
        sc.tl.leiden(adata_temp, resolution=0.3, random_state=0)
        pd.DataFrame(adata_temp.obs).to_csv(path_or_buf=dir_anndata+"adata_raw_ta_cd4_tcells_obs.csv")
        obs = adata_temp.obs
    else:
        obs = pd.read_csv(dir_anndata+'adata_raw_ta_cd4_tcells_obs.csv')
    adata_proc_ta_tcells.obs['leiden_cd4_ta'] = [
        obs.loc[x, "leiden"] if y == "Cd4" and x in obs.index else "other"
        for x, y in zip(
            adata_proc_ta_tcells.obs_names,
            adata_proc_ta_tcells.obs['cell_type'].values
        )
    ]
    
    adata_temp = adata_proc_ta_tcells[
        adata_proc_ta_tcells.obs["cell_type"] == "Cd8"
    ].copy()
    sc.pp.highly_variable_genes(adata_temp)
    sc.pp.pca(adata_temp)
    sc.pp.neighbors(adata_temp)
    if bool_recluster == True or True:
        sc.tl.leiden(adata_temp, resolution=0.3, random_state=0)
        pd.DataFrame(adata_temp.obs).to_csv(path_or_buf=dir_anndata+"adata_raw_ta_cd8_tcells_obs.csv")
        obs = adata_temp.obs
    else:
        obs = pd.read_csv(dir_anndata+'adata_raw_ta_cd8_tcells_obs.csv')
    adata_proc_ta_tcells.obs['leiden_cd8_ta'] = [
        obs.loc[x, "leiden"] if y == "Cd8" and x in obs.index else "other"
        for x, y in zip(
            adata_proc_ta_tcells.obs_names,
            adata_proc_ta_tcells.obs['cell_type'].values
        )
    ]
    
    adata_proc_ta_tcells.obs["leiden_all"] = [
        "cd4_" + x if z == "Cd4" else "cd8_" + y
        for x, y,z in zip(
            adata_proc_ta_tcells.obs['leiden_cd4_ta'].values,
            adata_proc_ta_tcells.obs['leiden_cd8_ta'].values,
            adata_proc_ta_tcells.obs['cell_type'].values
        )
    ]

# %% [code]
gene_ids = adata_proc_ta_tcells.var_names[np.asarray(adata_proc_ta_tcells[
    adata_proc_ta_tcells.obs["leiden_cd4_ta"].values != "other"
].X.mean(axis=0)).flatten() > np.log(1.0+0.5)]  # >logp1(mean = 0.5)
tab = pd.concat([
    adata_proc_ta_tcells[
        adata_proc_ta_tcells.obs["leiden_cd4_ta"].values != "other"
    ].obs["leiden_cd4_ta"],
    pd.DataFrame(
        adata_proc_ta_tcells[
            adata_proc_ta_tcells.obs["leiden_cd4_ta"].values != "other",
            gene_ids
        ].X.toarray(),
        columns=gene_ids,
        index=adata_proc_ta_tcells[adata_proc_ta_tcells.obs["leiden_cd4_ta"].values != "other"].obs.index
    ),
], axis=1).groupby("leiden_cd4_ta").mean().reset_index().sort_values("leiden_cd4_ta")
tab.index = tab["leiden_cd4_ta"]
leiden_clusters_ta_cd4 = np.unique(tab["leiden_cd4_ta"].values)
g = sb.clustermap(tab.iloc[:, 1:], figsize=(20,20))
plt.close()
idx_leiden_ta_cd4 = g.dendrogram_row.reordered_ind
leiden_clusters_ta_cd4_ordered = np.array(leiden_clusters_ta_cd4)[idx_leiden_ta_cd4]

adata_proc_ta_tcells.obs["leiden_cd4_ta"] = pd.Categorical(
    adata_proc_ta_tcells.obs["leiden_cd4_ta"].values,
    categories=np.concatenate([leiden_clusters_ta_cd4_ordered, ["other"]])
)

# %% [code]
gene_ids = adata_proc_ta_tcells.var_names[np.asarray(adata_proc_ta_tcells[
    adata_proc_ta_tcells.obs["leiden_cd8_ta"].values != "other"
].X.mean(axis=0)).flatten() > np.log(1.0+0.5)]  # >logp1(mean = 0.5)
tab = pd.concat([
    adata_proc_ta_tcells[
        adata_proc_ta_tcells.obs["leiden_cd8_ta"].values != "other"
    ].obs["leiden_cd8_ta"],
    pd.DataFrame(
        adata_proc_ta_tcells[
            adata_proc_ta_tcells.obs["leiden_cd8_ta"].values != "other",
            gene_ids
        ].X.toarray(),
        columns=gene_ids,
        index=adata_proc_ta_tcells[adata_proc_ta_tcells.obs["leiden_cd8_ta"].values != "other"].obs.index
    ),
], axis=1).groupby("leiden_cd8_ta").mean().reset_index().sort_values("leiden_cd8_ta")
tab.index = tab["leiden_cd8_ta"]
leiden_clusters_ta_cd8 = np.unique(tab["leiden_cd8_ta"].values)
g = sb.clustermap(tab.iloc[:, 1:], figsize=(20,20))
plt.close()
idx_leiden_ta_cd8 = g.dendrogram_row.reordered_ind
leiden_clusters_ta_cd8_ordered = np.array(leiden_clusters_ta_cd8)[idx_leiden_ta_cd8]

adata_proc_ta_tcells.obs["leiden_cd8_ta"] = pd.Categorical(
    adata_proc_ta_tcells.obs["leiden_cd8_ta"].values,
    categories=np.concatenate([leiden_clusters_ta_cd8_ordered, ["other"]])
)
adata_p1_withta.obs["leiden_all_ta"] = pd.Categorical(
    [
        adata_proc_ta_tcells.obs.loc[x, "leiden_all"] if x in adata_proc_ta_tcells.obs_names 
        else y for x, y in zip(
            adata_p1_withta.obs_names,
            adata_p1_withta.obs["cell_type"].values,
        )
    ]
)

# %% [code]
sc.pl.umap(adata_proc_ta_tcells, color=["CD3D", "CD3E", "CD3G"], use_raw=False)
sc.pl.umap(adata_proc_ta_tcells, color=["CD4", "CD8A", "CD8B"], use_raw=False)
sc.pl.umap(adata_proc_ta_tcells, color=["leiden_all"], use_raw=False)

fig, ax = plt.subplots(nrows=1, ncols=1, figsize=(5,5))
sc.pl.umap(adata_proc_ta_tcells, ax=ax, show=False)
sc.pl.umap(
    adata_proc_ta_tcells[adata_proc_ta_tcells.obs["leiden_cd4_ta"].values != "other"], 
    color=["leiden_cd4_ta"], 
    use_raw=False, 
    ax=ax, show=False
)
plt.savefig(dir_panels + "sfigx_ta_cd4_states.pdf")
plt.show()

fig, ax = plt.subplots(nrows=1, ncols=1, figsize=(5,5))
sc.pl.umap(adata_proc_ta_tcells, ax=ax, show=False)
sc.pl.umap(
    adata_proc_ta_tcells[adata_proc_ta_tcells.obs["leiden_cd8_ta"].values != "other"], 
    color=["leiden_cd8_ta"], 
    use_raw=False,
    ax=ax, show=False
)
plt.savefig(dir_panels + "sfigx_ta_cd8_states.pdf")
plt.show()

fig, ax = plt.subplots(nrows=1, ncols=1, figsize=(5,5))
sc.pl.umap(adata_proc_ta_tcells, ax=ax, show=False)
sc.pl.umap(adata_proc_ta_tcells, color=["cell_type"], use_raw=False, ax=ax, show=False)
plt.savefig(dir_panels + "sfigx_ta_celltypes.pdf")
plt.show()

# %% [code]
adata_proc_ta_tcells.obs.groupby("leiden_all").size()

# %% [code]
sc.pl.dotplot(
    adata_proc_ta_tcells, 
    var_names=["CD3D", "CD3E", "CD4", "CD8A", "CD8B"], 
    groupby="leiden_all",
)

# %% [code]
for k, v in markers_cd4_ta.items():
    sc.pl.dotplot(
        adata_proc_ta_tcells[adata_proc_ta_tcells.obs["cell_type"].values == "Cd4"], 
        var_names=[x for x in v if x in adata_proc_ta_tcells.var_names], 
        groupby="leiden_cd4_ta" ,
        use_raw=False,
        save="sfig10x_ta_" + "Cd4".lower() + "_tcells_leiden_" + k + ".pdf",
        show=False
    )
for k, v in markers_cd8_ta.items():
    sc.pl.dotplot(
        adata_proc_ta_tcells[adata_proc_ta_tcells.obs["cell_type"].values == "Cd8"], 
        var_names=[x for x in v if x in adata_proc_ta_tcells.var_names], 
        groupby="leiden_cd8_ta", 
        use_raw=False,
        save="sfig10x_ta_" + "Cd8".lower() + "_tcells_leiden_" + k + ".pdf",
        show=False
    )

# %% [markdown]
# QC TCR

# %% [code]
adata_proc_p1.obs.groupby("chain_pairing").size()

# %% [code]
adata_proc_p2.obs.groupby("chain_pairing").size()

# %% [code]
ir.pl.group_abundance(
    adata_proc_p1, 
    groupby="chain_pairing", 
    target_col="condition",
)
ir.pl.group_abundance(
    adata_proc_p2, 
    groupby="chain_pairing", 
    target_col="condition",
)

# %% [code]
sc.pl.umap(adata_proc_p1, color=["multi_chain", "chain_pairing"])
sc.pl.umap(adata_proc_p2, color=["multi_chain", "chain_pairing"])

# %% [code]
adata_proc_p1.obs.groupby(["sample_rna", "chain_pairing"]).size().reset_index()

# %% [code]
adata_proc_p2.obs.groupby(["sample_rna", "chain_pairing"]).size().reset_index()

# %% [markdown]
# TCR processing

# %% [markdown]
## AA distances

# %% [code]
# Define neighbours on aa sequence
if bool_recomp_dist:
    ir.pp.tcr_neighbors(
        adata_proc_p1,
        metric="alignment",
        sequence="aa",
        cutoff=50,
        receptor_arms="all",
        dual_tcr="primary_only",
    )
    adata_proc_p1_int = adata_proc_p1.copy()
    for k in ["repertoire_overlap", "clonotype_imbalance"]:  # cannot save
        if k in adata_proc_p1_int.uns.keys():
            del adata_proc_p1_int.uns[k]
    sc.write(dir_anndata+'adata_proc_p1_intermediate.h5ad', adata_proc_p1_int)
    del adata_proc_p1_int
else:
    adata_proc_p1_int = sc.read(dir_anndata+'adata_proc_p1_intermediate.h5ad') 
    adata_proc_p1.uns['tcr_neighbors_aa_alignment'] = adata_proc_p1_int.uns['tcr_neighbors_aa_alignment']
    del adata_proc_p1_int

# %% [code]
if bool_recomp_dist:
    ir.pp.tcr_neighbors(
        adata_proc_p2,
        metric="alignment",
        sequence="aa",
        cutoff=50,
        receptor_arms="all",
        dual_tcr="primary_only",
    )
    adata_proc_p2_int = adata_proc_p2.copy()
    for k in ["repertoire_overlap", "clonotype_imbalance"]:  # cannot save
        if k in adata_proc_p2.uns.keys():
            del adata_proc_p2.uns[k]
    sc.write(dir_anndata+'adata_proc_p2_intermediate.h5ad', adata_proc_p2_int)
    del adata_proc_p2_int
else:
    adata_proc_p2_int = sc.read(dir_anndata+'adata_proc_p2_intermediate.h5ad') 
    adata_proc_p2.uns['tcr_neighbors_aa_alignment'] = adata_proc_p2_int.uns['tcr_neighbors_aa_alignment']
    del adata_proc_p2_int

# %% [markdown]
## Nt distances

# %% [markdown]
### Primary chain

# %% [code]
# Define neighbours on nt sequence
if bool_recomp:
    ir.pp.tcr_neighbors(
        adata_proc_p1, 
        metric='identity',
        sequence="nt",
        receptor_arms="all", 
        dual_tcr="primary_only"
    )
    ir.tl.define_clonotypes(adata_proc_p1)
    
    ir.pp.tcr_neighbors(
        adata_proc_p2, 
        metric='identity',
        sequence="nt",
        receptor_arms="all", 
        dual_tcr="primary_only"
    )
    ir.tl.define_clonotypes(adata_proc_p2)

# %% [code]
if bool_recomp:
    ir.tl.define_clonotype_clusters(
        adata_proc_p1, partitions="connected", sequence="aa", metric="alignment"
    )
    ir.tl.define_clonotype_clusters(
        adata_proc_p1, 
        sequence="aa", 
        metric="alignment",
        same_v_gene="primary_only",
        key_added="ct_cluster_aa_alignment_same_v",
    )
    ir.tl.clonotype_network(adata_proc_p1, min_size=4, sequence="aa", metric="alignment")
    ir.tl.clonal_expansion(adata_proc_p1)
    
    ir.tl.define_clonotype_clusters(
        adata_proc_p2, partitions="connected", sequence="aa", metric="alignment"
    )
    ir.tl.define_clonotype_clusters(
        adata_proc_p2, 
        sequence="aa", 
        metric="alignment",
        same_v_gene="primary_only",
        key_added="ct_cluster_aa_alignment_same_v",
    )
    ir.tl.clonotype_network(adata_proc_p2, min_size=4, sequence="aa", metric="alignment")
    ir.tl.clonal_expansion(adata_proc_p2)

# %% [code]
sc.pl.umap(adata_proc_p1, color=["clonal_expansion", "clonotype_size"])
sc.pl.umap(adata_proc_p2, color=["clonal_expansion", "clonotype_size"])

# %% [code]
fig, ax = plt.subplots(figsize=(5.15,5))
sc.pl.umap(
    adata_proc_p1, 
    ax=ax, show=False
)
sc.pl.umap(
    adata_proc_p1[adata_proc_p1.obs["condition"].values == "unstimulated"], 
    color=["clonal_expansion"], 
    ax=ax, show=False
)
plt.tight_layout()
plt.savefig(dir_panels + "fig3c4.pdf")
plt.show()

# %% [code]
fig, ax = plt.subplots(figsize=(5,5))
sc.pl.umap(
    adata_proc_p2, 
    ax=ax, show=False
)
sc.pl.umap(
    adata_proc_p2[adata_proc_p2.obs["condition"].values == "unstimulated"], 
    color=["clonal_expansion"], 
    ax=ax, show=False
)
plt.show()

# %% [code]
ir.pl.clonal_expansion(adata_proc_p1, groupby="leiden_corrected", clip_at=4, normalize=False)
ir.pl.clonal_expansion(adata_proc_p1, "leiden_corrected")
ax = ir.pl.alpha_diversity(adata_proc_p1, groupby="leiden_corrected")

# %% [code]
ir.pl.clonal_expansion(adata_proc_p2, groupby="leiden_corrected", clip_at=4, normalize=False)
ir.pl.clonal_expansion(adata_proc_p2, "leiden_corrected")
ax = ir.pl.alpha_diversity(adata_proc_p2, groupby="leiden_corrected")

# %% [code]
ir.pl.spectratype(adata_proc_p1, color="leiden_corrected", viztype="bar", fig_kws={"dpi": 120})
ir.pl.spectratype(adata_proc_p2, color="leiden_corrected", viztype="bar", fig_kws={"dpi": 120})

# %% [markdown]
### All chains

# %% [code]
# Define neighbours on nt sequence
if bool_recomp:
    adata_proc_p1_temp = adata_proc_p1.copy()
    ir.pp.tcr_neighbors(
        adata_proc_p1_temp, 
        sequence="nt",
        receptor_arms="all", 
        dual_tcr="all"
    )
    ir.tl.define_clonotypes(adata_proc_p1_temp)
    adata_proc_p1.obs["clonotype_all_chains"] = adata_proc_p1_temp.obs["clonotype"].values
    del adata_proc_p1_temp
    
    adata_proc_p2_temp = adata_proc_p2.copy()
    ir.pp.tcr_neighbors(
        adata_proc_p2_temp, 
        sequence="nt",
        receptor_arms="all", 
        dual_tcr="all"
    )
    ir.tl.define_clonotypes(adata_proc_p2_temp)
    adata_proc_p2.obs["clonotype_all_chains"] = adata_proc_p2_temp.obs["clonotype"].values
    del adata_proc_p2_temp

# %% [markdown]
## Add clonotypes into sub objects

# %% [code]
adata_proc_p1_cor.obs["clonotype"] = adata_proc_p1[adata_proc_p1_cor.obs_names].obs["clonotype"].values
adata_proc_p2_cor.obs["clonotype"] = adata_proc_p2[adata_proc_p2_cor.obs_names].obs["clonotype"].values

adata_proc_p1_cor.obs["clonotype_all_chains"] = adata_proc_p1[adata_proc_p1_cor.obs_names].obs["clonotype_all_chains"].values
adata_proc_p2_cor.obs["clonotype_all_chains"] = adata_proc_p2[adata_proc_p2_cor.obs_names].obs["clonotype_all_chains"].values

# %% [code]
adata_proc_p1_stim.obs["clonotype"] = adata_proc_p1[adata_proc_p1_stim.obs_names].obs["clonotype"].values
adata_proc_p2_stim.obs["clonotype"] = adata_proc_p2[adata_proc_p2_stim.obs_names].obs["clonotype"].values

adata_proc_p1_unstim.obs["clonotype"] = adata_proc_p1[adata_proc_p1_unstim.obs_names].obs["clonotype"].values
adata_proc_p2_unstim.obs["clonotype"] = adata_proc_p2[adata_proc_p2_unstim.obs_names].obs["clonotype"].values

# %% [code]
adata_proc_p1_stim.obs["clonotype_all_chains"] = adata_proc_p1[adata_proc_p1_stim.obs_names].obs["clonotype_all_chains"].values
adata_proc_p2_stim.obs["clonotype_all_chains"] = adata_proc_p2[adata_proc_p2_stim.obs_names].obs["clonotype_all_chains"].values

adata_proc_p1_unstim.obs["clonotype_all_chains"] = adata_proc_p1[adata_proc_p1_unstim.obs_names].obs["clonotype_all_chains"].values
adata_proc_p2_unstim.obs["clonotype_all_chains"] = adata_proc_p2[adata_proc_p2_unstim.obs_names].obs["clonotype_all_chains"].values

# %% [markdown]
# Clonotype analysis

# %% [markdown]
## Number of clonotypes per sample

# %% [markdown]
### Primary chain clonotypes

# %% [code]
adata_proc_p1.obs["bc"] = adata_proc_p1.obs.index
adata_proc_p2.obs["bc"] = adata_proc_p2.obs.index

# %% [code]
df = adata_proc_p1.obs.groupby(["condition", "cell_type"])["clonotype"].unique().reset_index()
df["n_clonotypes"] = [len(x) for x in df["clonotype"].values]
print(df[["condition", "cell_type", "n_clonotypes"]])
print(adata_proc_p1.obs.groupby(["condition", "cell_type"]).size().reset_index())

# %% [code]
df = adata_proc_p2.obs.groupby(["condition", "cell_type"])["clonotype"].unique().reset_index()
df["n_clonotypes"] = [len(x) for x in df["clonotype"].values]
print(df[["condition", "cell_type", "n_clonotypes"]])
print(adata_proc_p2.obs.groupby(["condition", "cell_type"]).size().reset_index())

# %% [markdown]
### All chains clonotypes

# %% [code]
df = adata_proc_p1.obs.groupby(["condition", "cell_type"])["clonotype_all_chains"].unique().reset_index()
df["n_clonotypes"] = [len(x) for x in df["clonotype_all_chains"].values]
print(df[["condition", "cell_type", "n_clonotypes"]])
print(adata_proc_p1.obs.groupby(["condition", "cell_type"]).size().reset_index())

# %% [code]
df = adata_proc_p2.obs.groupby(["condition", "cell_type"])["clonotype_all_chains"].unique().reset_index()
df["n_clonotypes"] = [len(x) for x in df["clonotype_all_chains"].values]
print(df[["condition", "cell_type", "n_clonotypes"]])
print(adata_proc_p2.obs.groupby(["condition", "cell_type"]).size().reset_index())

# %% [markdown]
## Summarize clonotypes

# %% [markdown]
### Overlap scirpy to cellranger

# %% [markdown]
#### Primary chain clonotypes

# %% [code]
plt.hist(
    adata_proc_p1.obs[["clonotype", "clonotype_cellranger"]].\
        drop_duplicates().groupby("clonotype").size()
)

# %% [code]
plt.hist(
    adata_proc_p2.obs[["clonotype", "clonotype_cellranger"]].\
        drop_duplicates().groupby("clonotype").size()
)

# %% [markdown]
#### All chains clonotypes

# %% [code]
plt.hist(
    adata_proc_p1.obs[["clonotype_all_chains", "clonotype_cellranger"]].\
        drop_duplicates().groupby("clonotype_all_chains").size()
)

# %% [code]
plt.hist(
    adata_proc_p2.obs[["clonotype_all_chains", "clonotype_cellranger"]].\
        drop_duplicates().groupby("clonotype_all_chains").size()
)

# %% [markdown]
### Ordered by size

# %% [code]
ax = ir.pl.group_abundance(
    adata_proc_p1, 
    groupby="clonotype", 
    target_col="condition", 
    max_cols=30, 
    fig_kws={"figsize": (15, 5)}
)
ax = ir.pl.group_abundance(
    adata_proc_p2, 
    groupby="clonotype", 
    target_col="condition", 
    max_cols=30, 
    fig_kws={"figsize": (15, 5)}
)

# %% [code]
ir.pl.group_abundance(
    adata_proc_p1, 
    groupby="clonotype", 
    target_col="leiden_corrected", 
    max_cols=50, 
    figsize=(10, 5)
)
ir.pl.group_abundance(
    adata_proc_p2, 
    groupby="clonotype", 
    target_col="leiden_corrected", 
    max_cols=50, 
    figsize=(10, 5)
)

# %% [code]
ir.pl.group_abundance(
    adata_proc_p1, 
    groupby="clonotype", 
    target_col="cell_type", 
    max_cols=50, 
    figsize=(10, 5)
)
ir.pl.group_abundance(
    adata_proc_p2, 
    groupby="clonotype", 
    target_col="cell_type", 
    max_cols=50, 
    figsize=(10, 5)
)

# %% [markdown]
### Subselect clonotypes

# %% [code]
genes_to_record_in_clonotype_table = [
    "TNF",
    "IL2",
    "IL4",
    "IL5",
    "IL17",
    "IL10",
    "IL13",
    "IL21",
    "IL22",
    
    "FOXP3",
    "IL2RA",
    "IL12RB1",
    "IL18R1",
    "IL27RA",
    "IFNGR1", 
    "IFNGR2",
    "IL2RA",
    "KLRC1",  #NKG2A
    "PDCD1",  # PD1
    "HAVCR2",  # TIM3
    "CTLA4",
    "GZMA", 
    "GZMB",
    "TIGIT",
    "LAG3", 
    "ICOS", 
    "CD25",
    "CD38", 
    "ENTPD1",  # CD39
    "MKI67",  # Ki67
    "NME1",
    "IFIT1",
    "IFIT3",
    
    "TNFRSF4", 
    "TNFRSF9",
]

# %% [markdown]
#### Primary chain clonotypes

# %% [code]
clonotypes_p1 = [
    x for x in np.unique(adata_proc_p1.obs["clonotype"].values)
    if np.sum(np.logical_and(
        adata_proc_p1.obs["clonotype"].values == x,
        adata_proc_p1.obs["condition"].values == "stimulated"
    )) >= 1 and np.sum(np.logical_and(
        adata_proc_p1.obs["clonotype"].values == x,
        adata_proc_p1.obs["condition"].values == "unstimulated"
    )) >= 1
]
tab_clonotypes_p1_filtered = []
adata_temp = adata_proc_p1.copy()
keys = list(adata_temp.uns.keys())
for k in keys:
    del adata_temp.uns[k]
for x in clonotypes_p1:
    adata = adata_temp[adata_temp.obs["clonotype"].values == x]
    tab_clonotypes_p1_filtered.append(pd.DataFrame(dict(list({
        "clonotype": x,
        "clonotypes_all_chains": [np.unique(adata.obs["clonotype_all_chains"].values)],
        "exclusive_clonotype_map": len(np.unique(adata.obs["clonotype_all_chains"].values)) == 1,
        "size": adata.shape[0],
        "n_cd4": np.sum(adata.obs["cell_type"].values == "Cd4"),
        "n_cd8": np.sum(adata.obs["cell_type"].values == "Cd8"),
        "ave_cd4": np.mean(np.asarray(adata[:, "CD4"].X.todense()).flatten()),
        "ave_cd8": np.mean(adata.obs["CD8_all_genes"].values),
        "n_stimulated": np.sum(adata.obs["condition"].values == "stimulated"),
        "n_unstimulated": np.sum(adata.obs["condition"].values == "unstimulated"),
        "in_ifng_cluster": np.mean(adata.obs["ifng_pos_cluster"].values),
        "ave_ifng+": np.mean(np.asarray(adata[:, "IFNG"].X.todense()).flatten() > 0),
        "ave_ifng+_stimulated": np.mean(np.logical_and(
            adata.obs["condition"].values == "stimulated",
            np.asarray(adata[:, "IFNG"].X.todense()).flatten() > 0,
        )),
        "ave_ifng": np.mean(np.asarray(adata[:, "IFNG"].X.todense()).flatten()),
        "ave_ifng_stimulated": np.mean(np.asarray(adata[:, "IFNG"].X.todense()).flatten()[
            adata.obs["condition"].values == "stimulated",
        ]),
        "lfc_ifng_stimulated": np.log(np.mean(np.asarray(adata[:, "IFNG"].X.todense()).flatten()[
            adata.obs["condition"].values == "stimulated",
        ])+1) - np.log(np.mean(np.asarray(adata[:, "IFNG"].X.todense()).flatten()[
            adata.obs["condition"].values == "unstimulated",
        ])+1),
        "pval_ifng_condition": scipy.stats.ttest_ind(
            a=np.asarray(adata[:, "IFNG"].X.todense()).flatten()[
                adata.obs["condition"].values == "stimulated",
            ],
            b=np.asarray(adata[:, "IFNG"].X.todense()).flatten()[
                adata.obs["condition"].values == "unstimulated",
            ],
            equal_var=False
        )[1],
        "frac_orphan_beta": np.mean(adata.obs["chain_pairing"].values == "Orphan beta"),
    }.items()) + [
        (k, [np.unique(adata.obs[k].values)[np.logical_not(np.isnan(np.unique(adata.obs[k].values)))]])
        if adata.obs[k].values.dtype == np.float 
        else (k, [np.unique(adata.obs[k].values)])
        for k in adata.obs.columns
        if "TRA_" in k or "TRB_" in k
    ] + [
        ("ave_" + k, np.nanmean(adata.obs[k].values))
        for k in adata.obs.columns
        if "_expr" in k  # TRX_X_expr
    ] + [
        ("ave_" + k.lower(), [np.mean(np.asarray(adata[:, k].X.todense())).flatten()[0]])
        if k in adata.var_names else ("ave_" + k.lower(), 0.)
        for k in genes_to_record_in_clonotype_table
    ] + [
        ("ave_" + k.lower() + "_unstimulated", [np.mean(np.asarray(
            adata[adata.obs["condition"].values == "unstimulated", k].X.todense()
        )).flatten()[0]])
        if k in adata.var_names else ("ave_" + k.lower(), 0.)
        for k in genes_to_record_in_clonotype_table
    ] + [
        ("ave_" + k.lower() + "_stimulated", [np.mean(np.asarray(
            adata[adata.obs["condition"].values == "stimulated", k].X.todense()
        )).flatten()[0]])
        if k in adata.var_names else ("ave_" + k.lower(), 0.)
        for k in genes_to_record_in_clonotype_table
    ] + [
        (k.lower() + "_score", np.mean(adata.obs[k.lower() + "_score"].values))
        for k in tcell_signatures.keys()
    ] + [
        (k.lower() + "_score_stim", np.mean(adata.obs.loc[
            adata.obs["condition"].values == "stimulated", 
            k.lower() + "_score"
        ].values))
        for k in tcell_signatures.keys()
    ] + [
        (k.lower() + "_score_unstim", np.mean(adata.obs.loc[
            adata.obs["condition"].values == "unstimulated", 
            k.lower() + "_score"
        ].values))
        for k in tcell_signatures.keys()
    ] + [
        ("pval_" + k.lower() + "_score_cond", scipy.stats.ttest_ind(
            a=adata.obs.loc[
                adata.obs["condition"].values == "stimulated",
                k.lower() + "_score"
            ],
            b=adata.obs.loc[
                adata.obs["condition"].values == "unstimulated",
                k.lower() + "_score"
            ],
            equal_var=False
        )[1])
        for k in tcell_signatures.keys()
    ]), index=[x]))
tab_clonotypes_p1_filtered = pd.concat(tab_clonotypes_p1_filtered, axis=0)
tab_clonotypes_p1_filtered["label_cell_type"] = [
    ["Cd4", "Cd8"][x < y] for x, y in zip(
        tab_clonotypes_p1_filtered["n_cd4"].values,
        tab_clonotypes_p1_filtered["n_cd8"].values
    )
]

map_single_beta = {}
for x, z in [(x, z) for x, y, z in zip(
    tab_clonotypes_p1_filtered["clonotype"].values,
    tab_clonotypes_p1_filtered["TRA_1_cdr3"].values,
    tab_clonotypes_p1_filtered["label_cell_type"].values
) if y == ["None"]]:
    # Check if any other clonotype has same beta chain:
    trb = tab_clonotypes_p1_filtered["TRB_1_cdr3"].values[tab_clonotypes_p1_filtered["clonotype"].values == x][0]
    mapped = list(set(tab_clonotypes_p1_filtered["clonotype"].values[np.logical_and(
        tab_clonotypes_p1_filtered["label_cell_type"].values == z,
        np.logical_or(
            tab_clonotypes_p1_filtered["TRB_1_cdr3"].values == trb,
            tab_clonotypes_p1_filtered["TRB_2_cdr3"].values == trb
        )
    )]) - set([x]))
    if len(mapped) > 0:
        if len(mapped) > 1:
            mapped = list(set(tab_clonotypes_p1_filtered["clonotype"].values[np.logical_and(
                tab_clonotypes_p1_filtered["TRB_1_cdr3"].values == trb,
                tab_clonotypes_p1_filtered["label_cell_type"].values == z,
            )]) - set([x]))
            if len(mapped) > 1:
                map_single_beta[x] = x + "_unclear"
        map_single_beta[x] = mapped[0]
tab_clonotypes_p1_filtered["clonotype_merged"] = [
    map_single_beta[x] if x in map_single_beta.keys() else x
    for x in tab_clonotypes_p1_filtered["clonotype"].values
]
adata_proc_p1.obs["clonotype_merged"] = [
    map_single_beta[x] if x in map_single_beta.keys() else x
    for x in adata_proc_p1.obs["clonotype"].values
]

print(tab_clonotypes_p1_filtered.groupby("label_cell_type").size())

# %% [code]
tab = pd.concat([
    tab_clonotypes_p1_filtered.loc[
        np.logical_and(
            tab_clonotypes_p1_filtered["TRA_1_cdr3"].values == x,
            tab_clonotypes_p1_filtered["TRB_1_cdr3"].values == y
        ), 
        [
            "clonotype", "size", "n_stimulated", "n_unstimulated", 
            "TRA_1_cdr3", "TRB_1_cdr3", 
            "TRA_1_v_gene", "TRB_1_v_gene", 
            "TRB_1_d_gene", 
            "TRA_1_j_gene", "TRB_1_j_gene"
        ]
    ] for x, y in zip(
        [
            ['CAVALGAAGNKLTF'],
            ['CAVRFEDNKLTF'],
            ['CAMREVNTNAGKSTF'],
            ['CAVGDSGSARQLTF'],
            ['CAMRGFNTNAGKSTF'],
            ['CAMREVNTNAGKSTF'],
            ['CAVASRGQKLVF'],
        ],
        [
            ['CASSRTGTGESGAFF'],
            ['CASSNPGQGVTEAFF'],
            ['CSARVEEAGGFSRGYTF'],
            ['CSALTSGNTGELFF'],
            ['CASSESDAADRVGQPQHF'],
            ['CASSSSGTAIYEQYF'],
            ['CASSLKVRGLVVNGSYNSPLHF'],
        ]
    )
], axis=0)
tab.to_csv(dir_tables + "ExtendedDataTable10_validated_reactive_tcrs.csv", sep=";")
tab

# %% [code]
clonotypes_p2 = [
    x for x in np.unique(adata_proc_p2.obs["clonotype"].values)
    if np.sum(np.logical_and(
        adata_proc_p2.obs["clonotype"].values == x,
        adata_proc_p2.obs["condition"].values == "stimulated"
    )) >= 1 and np.sum(np.logical_and(
        adata_proc_p2.obs["clonotype"].values == x,
        adata_proc_p2.obs["condition"].values == "unstimulated"
    )) >= 1
]
tab_clonotypes_p2_filtered = []
adata_temp = adata_proc_p2.copy()
keys = list(adata_temp.uns.keys())
for k in keys:
    del adata_temp.uns[k]
for x in clonotypes_p2:
    adata = adata_temp[adata_temp.obs["clonotype"].values == x]
    tab_clonotypes_p2_filtered.append(pd.DataFrame(dict(list({
        "clonotype": x,
        "clonotypes_all_chains": [np.unique(adata.obs["clonotype_all_chains"].values)],
        "exclusive_clonotype_map": len(np.unique(adata.obs["clonotype_all_chains"].values)) == 1,
        "size": adata.shape[0],
        "n_cd4": np.sum(adata.obs["cell_type"].values == "Cd4"),
        "n_cd8": np.sum(adata.obs["cell_type"].values == "Cd8"),
        "ave_cd4": np.mean(np.asarray(adata[:, "CD4"].X.todense()).flatten()),
        "ave_cd8": np.mean(adata.obs["CD8_all_genes"].values),
        "n_stimulated": np.sum(adata.obs["condition"].values == "stimulated"),
        "n_unstimulated": np.sum(adata.obs["condition"].values == "unstimulated"),
        "in_ifng_cluster": np.mean(adata.obs["ifng_pos_cluster"].values),
        "ave_ifng+": np.mean(np.asarray(adata[:, "IFNG"].X.todense()).flatten() > 0),
        "ave_ifng+_stimulated": np.mean(np.logical_and(
            adata.obs["condition"].values == "stimulated",
            np.asarray(adata[:, "IFNG"].X.todense()).flatten() > 0,
        )),
        "ave_ifng": np.mean(np.asarray(adata[:, "IFNG"].X.todense()).flatten()),
        "ave_ifng_stimulated": np.mean(np.asarray(adata[:, "IFNG"].X.todense()).flatten()[
            adata.obs["condition"].values == "stimulated",
        ]),
        "lfc_ifng_stimulated": np.log(np.mean(np.asarray(adata[:, "IFNG"].X.todense()).flatten()[
            adata.obs["condition"].values == "stimulated",
        ])+1) - np.log(np.mean(np.asarray(adata[:, "IFNG"].X.todense()).flatten()[
            adata.obs["condition"].values == "unstimulated",
        ])+1),
        "pval_ifng_condition": scipy.stats.ttest_ind(
            a=np.asarray(adata[:, "IFNG"].X.todense()).flatten()[
                adata.obs["condition"].values == "stimulated",
            ],
            b=np.asarray(adata[:, "IFNG"].X.todense()).flatten()[
                adata.obs["condition"].values == "unstimulated",
            ],
            equal_var=False
        )[1],
        "frac_orphan_beta": np.mean(adata.obs["chain_pairing"].values == "Orphan beta"),
    }.items()) + [
        (k, [np.unique(adata.obs[k].values)[np.logical_not(np.isnan(np.unique(adata.obs[k].values)))]])
        if adata.obs[k].values.dtype == np.float 
        else (k, [np.unique(adata.obs[k].values)])
        for k in adata.obs.columns
        if "TRA_" in k or "TRB_" in k
    ] + [
        ("ave_" + k, np.nanmean(adata.obs[k].values))
        for k in adata.obs.columns
        if "_expr" in k  # TRX_X_expr
    ] + [
        ("ave_" + k.lower(), [np.mean(np.asarray(adata[:, k].X.todense())).flatten()[0]])
        if k in adata.var_names else ("ave_" + k.lower(), 0.)
        for k in genes_to_record_in_clonotype_table
    ] + [
        ("ave_" + k.lower() + "_unstimulated", [np.mean(np.asarray(
            adata[adata.obs["condition"].values == "unstimulated", k].X.todense()
        )).flatten()[0]])
        if k in adata.var_names else ("ave_" + k.lower(), 0.)
        for k in genes_to_record_in_clonotype_table
    ] + [
        ("ave_" + k.lower() + "_stimulated", [np.mean(np.asarray(
            adata[adata.obs["condition"].values == "stimulated", k].X.todense()
        )).flatten()[0]])
        if k in adata.var_names else ("ave_" + k.lower(), 0.)
        for k in genes_to_record_in_clonotype_table
    ] + [
        (k.lower() + "_score", np.mean(adata.obs[k.lower() + "_score"].values))
        for k in tcell_signatures.keys()
    ] + [
        (k.lower() + "_score_stim", np.mean(adata.obs.loc[
            adata.obs["condition"].values == "stimulated", 
            k.lower() + "_score"
        ].values))
        for k in tcell_signatures.keys()
    ] + [
        (k.lower() + "_score_unstim", np.mean(adata.obs.loc[
            adata.obs["condition"].values == "unstimulated", 
            k.lower() + "_score"
        ].values))
        for k in tcell_signatures.keys()
    ] + [
        ("pval_" + k.lower() + "_score_cond", scipy.stats.ttest_ind(
            a=adata.obs.loc[
                adata.obs["condition"].values == "stimulated",
                k.lower() + "_score"
            ],
            b=adata.obs.loc[
                adata.obs["condition"].values == "unstimulated",
                k.lower() + "_score"
            ],
            equal_var=False
        )[1])
        for k in tcell_signatures.keys()
    ]), index=[x]))
tab_clonotypes_p2_filtered = pd.concat(tab_clonotypes_p2_filtered, axis=0)
tab_clonotypes_p2_filtered["label_cell_type"] = [
    ["Cd4", "Cd8"][x < y] for x, y in zip(
        tab_clonotypes_p2_filtered["n_cd4"].values,
        tab_clonotypes_p2_filtered["n_cd8"].values
    )
]

map_single_beta = {}
for x, z in [(x, z) for x, y, z in zip(
    tab_clonotypes_p2_filtered["clonotype"].values,
    tab_clonotypes_p2_filtered["TRA_1_cdr3"].values,
    tab_clonotypes_p2_filtered["label_cell_type"].values
) if y == ["None"]]:
    # Check if any other clonotype has same beta chain:
    trb = tab_clonotypes_p2_filtered["TRB_1_cdr3"].values[tab_clonotypes_p2_filtered["clonotype"].values == x][0]
    mapped = list(set(tab_clonotypes_p2_filtered["clonotype"].values[np.logical_and(
        tab_clonotypes_p2_filtered["label_cell_type"].values == z,
        np.logical_or(
            tab_clonotypes_p2_filtered["TRB_1_cdr3"].values == trb,
            tab_clonotypes_p2_filtered["TRB_2_cdr3"].values == trb
        )
    )]) - set([x]))
    if len(mapped) > 0:
        if len(mapped) > 1:
            mapped = list(set(tab_clonotypes_p2_filtered["clonotype"].values[np.logical_and(
                tab_clonotypes_p2_filtered["TRB_1_cdr3"].values == trb,
                tab_clonotypes_p2_filtered["label_cell_type"].values == z,
            )]) - set([x]))
            if len(mapped) > 1:
                map_single_beta[x] = x + "_unclear"
        map_single_beta[x] = mapped[0]
tab_clonotypes_p2_filtered["clonotype_merged"] = [
    map_single_beta[x] if x in map_single_beta.keys() else x
    for x in tab_clonotypes_p2_filtered["clonotype"].values
]
adata_proc_p2.obs["clonotype_merged"] = [
    map_single_beta[x] if x in map_single_beta.keys() else x
    for x in adata_proc_p2.obs["clonotype"].values
]

print(tab_clonotypes_p2_filtered.groupby("label_cell_type").size())

# %% [markdown]
#### All chains clonotypes

# %% [code]
clonotype_all_chains_p1 = [
    x for x in np.unique(adata_proc_p1.obs["clonotype_all_chains"].values)
    if (np.sum(np.logical_and(
        adata_proc_p1.obs["clonotype_all_chains"].values == x,
        adata_proc_p1.obs["condition"].values == "stimulated"
    )) >= 1 and np.sum(np.logical_and(
        adata_proc_p1.obs["clonotype_all_chains"].values == x,
        adata_proc_p1.obs["condition"].values == "unstimulated"
    )) >= 1) or x in tab_clonotypes_p1_filtered["clonotypes_all_chains"].values
]
tab_clonotypes_all_chains_p1_filtered = []
adata_temp = adata_proc_p1.copy()
keys = list(adata_temp.uns.keys())
for k in keys:
    del adata_temp.uns[k]
for x in clonotype_all_chains_p1:
    adata = adata_temp[adata_temp.obs["clonotype_all_chains"].values == x]
    tab_clonotypes_all_chains_p1_filtered.append(pd.DataFrame(dict(list({
        "clonotype_all_chains": x,
        "clonotype": [np.unique(adata.obs["clonotype"].values)],
        "exclusive_clonotype_map": len(np.unique(adata.obs["clonotype"].values)) == 1,
        "size": adata.shape[0],
        "n_cd4": np.sum(adata.obs["cell_type"].values == "Cd4"),
        "n_cd8": np.sum(adata.obs["cell_type"].values == "Cd8"),
        "ave_cd4": np.mean(np.asarray(adata[:, "CD4"].X.todense()).flatten()),
        "ave_cd8": np.mean(adata.obs["CD8_all_genes"].values),
        "n_stimulated": np.sum(adata.obs["condition"].values == "stimulated"),
        "n_unstimulated": np.sum(adata.obs["condition"].values == "unstimulated"),
        "in_ifng_cluster": np.mean(adata.obs["ifng_pos_cluster"].values),
        "ave_ifng+": np.mean(np.asarray(adata[:, "IFNG"].X.todense()).flatten() > 0),
        "ave_ifng+_stimulated": np.mean(np.logical_and(
            adata.obs["condition"].values == "stimulated",
            np.asarray(adata[:, "IFNG"].X.todense()).flatten() > 0,
        )),
        "ave_ifng": np.mean(np.asarray(adata[:, "IFNG"].X.todense()).flatten()),
        "ave_ifng_stimulated": np.mean(np.asarray(adata[:, "IFNG"].X.todense()).flatten()[
            adata.obs["condition"].values == "stimulated",
        ]),
        "lfc_ifng_stimulated": np.log(np.mean(np.asarray(adata[:, "IFNG"].X.todense()).flatten()[
            adata.obs["condition"].values == "stimulated",
        ])+1) - np.log(np.mean(np.asarray(adata[:, "IFNG"].X.todense()).flatten()[
            adata.obs["condition"].values == "unstimulated",
        ])+1),
        "pval_ifng_condition": scipy.stats.ttest_ind(
            a=np.asarray(adata[:, "IFNG"].X.todense()).flatten()[
                adata.obs["condition"].values == "stimulated",
            ],
            b=np.asarray(adata[:, "IFNG"].X.todense()).flatten()[
                adata.obs["condition"].values == "unstimulated",
            ],
            equal_var=False
        )[1],
        "frac_orphan_beta": np.mean(adata.obs["chain_pairing"].values == "Orphan beta"),
    }.items()) + [
        (k, [np.unique(adata.obs[k].values)[np.logical_not(np.isnan(np.unique(adata.obs[k].values)))]])
        if adata.obs[k].values.dtype == np.float 
        else (k, [np.unique(adata.obs[k].values)])
        for k in adata.obs.columns
        if "TRA_" in k or "TRB_" in k
    ] + [
        ("ave_" + k, np.nanmean(adata.obs[k].values))
        for k in adata.obs.columns
        if "_expr" in k  # TRX_X_expr
    ] + [
        ("ave_" + k.lower(), [np.mean(np.asarray(adata[:, k].X.todense())).flatten()[0]])
        if k in adata.var_names else ("ave_" + k.lower(), 0.)
        for k in genes_to_record_in_clonotype_table
    ] + [
        ("ave_" + k.lower() + "_unstimulated", [np.mean(np.asarray(
            adata[adata.obs["condition"].values == "unstimulated", k].X.todense()
        )).flatten()[0]])
        if k in adata.var_names else ("ave_" + k.lower(), 0.)
        for k in genes_to_record_in_clonotype_table
    ] + [
        ("ave_" + k.lower() + "_stimulated", [np.mean(np.asarray(
            adata[adata.obs["condition"].values == "stimulated", k].X.todense()
        )).flatten()[0]])
        if k in adata.var_names else ("ave_" + k.lower(), 0.)
        for k in genes_to_record_in_clonotype_table
    ] + [
        (k.lower() + "_score", np.mean(adata.obs[k.lower() + "_score"].values))
        for k in tcell_signatures.keys()
    ] + [
        (k.lower() + "_score_stim", np.mean(adata.obs.loc[
            adata.obs["condition"].values == "stimulated", 
            k.lower() + "_score"
        ].values))
        for k in tcell_signatures.keys()
    ] + [
        (k.lower() + "_score_unstim", np.mean(adata.obs.loc[
            adata.obs["condition"].values == "unstimulated", 
            k.lower() + "_score"
        ].values))
        for k in tcell_signatures.keys()
    ] + [
        ("pval_" + k.lower() + "_score_cond", scipy.stats.ttest_ind(
            a=adata.obs.loc[
                adata.obs["condition"].values == "stimulated",
                k.lower() + "_score"
            ],
            b=adata.obs.loc[
                adata.obs["condition"].values == "unstimulated",
                k.lower() + "_score"
            ],
            equal_var=False
        )[1])
        for k in tcell_signatures.keys()
    ]), index=[x]))
tab_clonotypes_all_chains_p1_filtered = pd.concat(tab_clonotypes_all_chains_p1_filtered, axis=0)
tab_clonotypes_all_chains_p1_filtered["label_cell_type"] = [
    ["Cd4", "Cd8"][x < y] for x, y in zip(
        tab_clonotypes_all_chains_p1_filtered["n_cd4"].values,
        tab_clonotypes_all_chains_p1_filtered["n_cd8"].values
    )
]
print(tab_clonotypes_all_chains_p1_filtered.groupby("label_cell_type").size())

# %% [code]
clonotype_all_chains_p2 = [
    x for x in np.unique(adata_proc_p2.obs["clonotype_all_chains"].values)
    if (np.sum(np.logical_and(
        adata_proc_p2.obs["clonotype_all_chains"].values == x,
        adata_proc_p2.obs["condition"].values == "stimulated"
    )) >= 1 and np.sum(np.logical_and(
        adata_proc_p2.obs["clonotype_all_chains"].values == x,
        adata_proc_p2.obs["condition"].values == "unstimulated"
    )) >= 1) or x in tab_clonotypes_p2_filtered["clonotypes_all_chains"].values
]
tab_clonotypes_all_chains_p2_filtered = []
adata_temp = adata_proc_p2.copy()
keys = list(adata_temp.uns.keys())
for k in keys:
    del adata_temp.uns[k]
for x in clonotypes_p2:
    adata = adata_temp[adata_temp.obs["clonotype_all_chains"].values == x]
    tab_clonotypes_all_chains_p2_filtered.append(pd.DataFrame(dict(list({
        "clonotype_all_chains": x,
        "clonotype": [np.unique(adata.obs["clonotype"].values)],
        "exclusive_clonotype_map": len(np.unique(adata.obs["clonotype"].values)) == 1,
        "size": adata.shape[0],
        "n_cd4": np.sum(adata.obs["cell_type"].values == "Cd4"),
        "n_cd8": np.sum(adata.obs["cell_type"].values == "Cd8"),
        "ave_cd4": np.mean(np.asarray(adata[:, "CD4"].X.todense()).flatten()),
        "ave_cd8": np.mean(adata.obs["CD8_all_genes"].values),
        "n_stimulated": np.sum(adata.obs["condition"].values == "stimulated"),
        "n_unstimulated": np.sum(adata.obs["condition"].values == "unstimulated"),
        "in_ifng_cluster": np.mean(adata.obs["ifng_pos_cluster"].values),
        "ave_ifng+": np.mean(np.asarray(adata[:, "IFNG"].X.todense()).flatten() > 0),
        "ave_ifng+_stimulated": np.mean(np.logical_and(
            adata.obs["condition"].values == "stimulated",
            np.asarray(adata[:, "IFNG"].X.todense()).flatten() > 0,
        )),
        "ave_ifng": np.mean(np.asarray(adata[:, "IFNG"].X.todense()).flatten()),
        "ave_ifng_stimulated": np.mean(np.asarray(adata[:, "IFNG"].X.todense()).flatten()[
            adata.obs["condition"].values == "stimulated",
        ]),
        "lfc_ifng_stimulated": np.log(np.mean(np.asarray(adata[:, "IFNG"].X.todense()).flatten()[
            adata.obs["condition"].values == "stimulated",
        ])+1) - np.log(np.mean(np.asarray(adata[:, "IFNG"].X.todense()).flatten()[
            adata.obs["condition"].values == "unstimulated",
        ])+1),
        "pval_ifng_condition": scipy.stats.ttest_ind(
            a=np.asarray(adata[:, "IFNG"].X.todense()).flatten()[
                adata.obs["condition"].values == "stimulated",
            ],
            b=np.asarray(adata[:, "IFNG"].X.todense()).flatten()[
                adata.obs["condition"].values == "unstimulated",
            ],
            equal_var=False
        )[1],
        "frac_orphan_beta": np.mean(adata.obs["chain_pairing"].values == "Orphan beta"),
    }.items()) + [
        (k, [np.unique(adata.obs[k].values)[np.logical_not(np.isnan(np.unique(adata.obs[k].values)))]])
        if adata.obs[k].values.dtype == np.float 
        else (k, [np.unique(adata.obs[k].values)])
        for k in adata.obs.columns
        if "TRA_" in k or "TRB_" in k
    ] + [
        ("ave_" + k, np.nanmean(adata.obs[k].values))
        for k in adata.obs.columns
        if "_expr" in k  # TRX_X_expr
    ] + [
        ("ave_" + k.lower(), [np.mean(np.asarray(adata[:, k].X.todense())).flatten()[0]])
        if k in adata.var_names else ("ave_" + k.lower(), 0.)
        for k in genes_to_record_in_clonotype_table
    ] + [
        ("ave_" + k.lower() + "_unstimulated", [np.mean(np.asarray(
            adata[adata.obs["condition"].values == "unstimulated", k].X.todense()
        )).flatten()[0]])
        if k in adata.var_names else ("ave_" + k.lower(), 0.)
        for k in genes_to_record_in_clonotype_table
    ] + [
        ("ave_" + k.lower() + "_stimulated", [np.mean(np.asarray(
            adata[adata.obs["condition"].values == "stimulated", k].X.todense()
        )).flatten()[0]])
        if k in adata.var_names else ("ave_" + k.lower(), 0.)
        for k in genes_to_record_in_clonotype_table
    ] + [
        (k.lower() + "_score", np.mean(adata.obs[k.lower() + "_score"].values))
        for k in tcell_signatures.keys()
    ] + [
        (k.lower() + "_score_stim", np.mean(adata.obs.loc[
            adata.obs["condition"].values == "stimulated", 
            k.lower() + "_score"
        ].values))
        for k in tcell_signatures.keys()
    ] + [
        (k.lower() + "_score_unstim", np.mean(adata.obs.loc[
            adata.obs["condition"].values == "unstimulated", 
            k.lower() + "_score"
        ].values))
        for k in tcell_signatures.keys()
    ] + [
        ("pval_" + k.lower() + "_score_cond", scipy.stats.ttest_ind(
            a=adata.obs.loc[
                adata.obs["condition"].values == "stimulated",
                k.lower() + "_score"
            ],
            b=adata.obs.loc[
                adata.obs["condition"].values == "unstimulated",
                k.lower() + "_score"
            ],
            equal_var=False
        )[1])
        for k in tcell_signatures.keys()
    ]), index=[x]))
tab_clonotypes_all_chains_p2_filtered = pd.concat(tab_clonotypes_all_chains_p2_filtered, axis=0)
tab_clonotypes_all_chains_p2_filtered["label_cell_type"] = [
    ["Cd4", "Cd8"][x < y] for x, y in zip(
        tab_clonotypes_all_chains_p2_filtered["n_cd4"].values,
        tab_clonotypes_all_chains_p2_filtered["n_cd8"].values
    )
]
print(tab_clonotypes_all_chains_p2_filtered.groupby("label_cell_type").size())

# %% [markdown]
### Create clonotype summary tables with expression per cell

# %% [code]
table_marker_keys = [
    "IFNG",
] + list(tcell_signatures.keys())

# %% [code]
clonotypes_p1 = [
    x for x in np.unique(adata_proc_p1.obs["clonotype"].values)
    if np.sum(np.logical_and(
        adata_proc_p1.obs["clonotype"].values == x,
        adata_proc_p1.obs["condition"].values == "stimulated"
    )) >= 3 and np.sum(np.logical_and(
        adata_proc_p1.obs["clonotype"].values == x,
        adata_proc_p1.obs["condition"].values == "unstimulated"
    )) >= 3
]
tab_clonotypes_p1_stim_filtered_bycell = {
    "Cd4": dict([
        ("_".join(k.lower().split(" ")) + "_score", [])
        if k in list(tcell_signatures.keys())
        else (k, [])
        for k in table_marker_keys
    ]),
    "Cd8": dict([
        ("_".join(k.lower().split(" ")) + "_score", [])
        if k in list(tcell_signatures.keys())
        else (k, [])
        for k in table_marker_keys
    ]),
}
tab_clonotypes_p1_unstim_filtered_bycell = {
    "Cd4": dict([
        ("_".join(k.lower().split(" ")) + "_score", [])
        if k in list(tcell_signatures.keys())
        else (k, [])
        for k in table_marker_keys
    ]),
    "Cd8": dict([
        ("_".join(k.lower().split(" ")) + "_score", [])
        if k in list(tcell_signatures.keys())
        else (k, [])
        for k in table_marker_keys
    ]),
}
adata_temp = adata_proc_p1.copy()
for k in keys:
    del adata_temp.uns[k]
ncell_cols_stim = np.max(tab_clonotypes_p1_filtered["n_stimulated"].values)
ncell_cols_unstim = np.max(tab_clonotypes_p1_filtered["n_unstimulated"].values)
for x in clonotypes_p1:
    adata = adata_temp[adata_temp.obs["clonotype"].values == x]
    ncells_stim = np.sum(adata.obs["condition"].values == "stimulated")
    ncells_unstim = np.sum(adata.obs["condition"].values == "unstimulated")
    cell_type = tab_clonotypes_p1_filtered.loc[
        tab_clonotypes_p1_filtered["clonotype"].values == x,
        "label_cell_type"
    ].values[0]
    for k, kk in zip(
        table_marker_keys, 
        list(tab_clonotypes_p1_unstim_filtered_bycell["Cd4"].keys())
    ):
        # stimulated
        tab_clonotypes_p1_stim_filtered_bycell[cell_type][kk].append(pd.DataFrame(dict(list({
            "clonotype": x,
            "clonotypes_all_chains": [np.unique(adata.obs["clonotype_all_chains"].values)],
            "size": adata.shape[0],
            "n_stimulated": np.sum(adata.obs["condition"].values == "stimulated"),
            "n_unstimulated": np.sum(adata.obs["condition"].values == "unstimulated"),
        }.items()) + [
            (
                "cell_" + str(i),
                adata.obs[k.lower() + "_score"].values[adata.obs["condition"].values == "stimulated"][i]
                if (k.lower() + "_score" in adata.obs.keys() and i < ncells_stim) else 
                np.asarray(adata[adata.obs["condition"].values == "stimulated", k].X.todense()).flatten()[i]
                if i < ncells_stim else np.nan
            )
            for i in range(ncell_cols_stim)
        ]), index=[x]))
        # unstimulated
        tab_clonotypes_p1_unstim_filtered_bycell[cell_type][kk].append(pd.DataFrame(dict(list({
            "clonotype": x,
            "clonotypes_all_chains": [np.unique(adata.obs["clonotype_all_chains"].values)],
            "size": adata.shape[0],
            "n_stimulated": np.sum(adata.obs["condition"].values == "stimulated"),
            "n_unstimulated": np.sum(adata.obs["condition"].values == "unstimulated"),
        }.items()) + [
            (
                "cell_" + str(i),
                adata.obs[k.lower() + "_score"].values[adata.obs["condition"].values == "unstimulated"][i]
                if (k.lower() + "_score" in adata.obs.keys() and i < ncells_unstim) else 
                np.asarray(adata[adata.obs["condition"].values == "unstimulated", k].X.todense()).flatten()[i]
                if i < ncells_unstim else np.nan
            )
            for i in range(ncell_cols_unstim)
        ]), index=[x]))
for k in ["Cd4", "Cd8"]:
    for kk in tab_clonotypes_p1_unstim_filtered_bycell[k].keys():
        tab_clonotypes_p1_unstim_filtered_bycell[k][kk] = pd.concat(tab_clonotypes_p1_unstim_filtered_bycell[k][kk], axis=0)
        tab_clonotypes_p1_unstim_filtered_bycell[k][kk].sort_values("size").iloc[::-1].to_csv(dir_tables+'tab_clonotypes_p1_' + k.lower() + '_' + kk.lower() + '_unstim_filtered_bycell.csv', sep=";")

        tab_clonotypes_p1_stim_filtered_bycell[k][kk] = pd.concat(tab_clonotypes_p1_stim_filtered_bycell[k][kk], axis=0)
        tab_clonotypes_p1_stim_filtered_bycell[k][kk].sort_values("size").iloc[::-1].to_csv(dir_tables+'tab_clonotypes_p1_' + k.lower() + '_'  + kk.lower() + '_stim_filtered_bycell.csv', sep=";")
tab_clonotypes_p1_unstim_filtered_bycell["Cd8"]["cd8_cytotoxic_score"].sort_values("size").iloc[::-1].to_csv(dir_tables+'sfig4a1_cd8_unstim.csv', sep=";")
tab_clonotypes_p1_unstim_filtered_bycell["Cd8"]["cd8_cytokine_score"].sort_values("size").iloc[::-1].to_csv(dir_tables+'sfig4a2_cd8_unstim.csv', sep=";")
tab_clonotypes_p1_unstim_filtered_bycell["Cd8"]["IFNG"].sort_values("size").iloc[::-1].to_csv(dir_tables+'sfig4a3_cd8_unstim.csv', sep=";")
tab_clonotypes_p1_stim_filtered_bycell["Cd8"]["cd8_cytotoxic_score"].sort_values("size").iloc[::-1].to_csv(dir_tables+'sfig4a1_cd8_stim.csv', sep=";")
tab_clonotypes_p1_stim_filtered_bycell["Cd8"]["cd8_cytokine_score"].sort_values("size").iloc[::-1].to_csv(dir_tables+'sfig4a2_cd8_stim.csv', sep=";")
tab_clonotypes_p1_stim_filtered_bycell["Cd8"]["IFNG"].sort_values("size").iloc[::-1].to_csv(dir_tables+'sfig4a3_cd8_stim.csv', sep=";")

tab_clonotypes_p1_unstim_filtered_bycell["Cd4"]["ifn_response_score"].sort_values("size").iloc[::-1].to_csv(dir_tables+'sfig4c1_cd4_unstim.csv', sep=";")
tab_clonotypes_p1_unstim_filtered_bycell["Cd4"]["proliferation_score"].sort_values("size").iloc[::-1].to_csv(dir_tables+'sfig4c2_cd4_unstim.csv', sep=";")
tab_clonotypes_p1_unstim_filtered_bycell["Cd4"]["IFNG"].sort_values("size").iloc[::-1].to_csv(dir_tables+'sfig4c3_cd4_unstim.csv', sep=";")
tab_clonotypes_p1_stim_filtered_bycell["Cd4"]["ifn_response_score"].sort_values("size").iloc[::-1].to_csv(dir_tables+'sfig4c1_cd4_stim.csv', sep=";")
tab_clonotypes_p1_stim_filtered_bycell["Cd4"]["proliferation_score"].sort_values("size").iloc[::-1].to_csv(dir_tables+'sfig4c2_cd4_stim.csv', sep=";")
tab_clonotypes_p1_stim_filtered_bycell["Cd4"]["IFNG"].sort_values("size").iloc[::-1].to_csv(dir_tables+'sfig4c3_cd4_stim.csv', sep=";")

# %% [code]
clonotypes_p1 = [
    x for x in np.unique(adata_proc_p1.obs["clonotype_merged"].values)
    if np.sum(np.logical_and(
        adata_proc_p1.obs["clonotype_merged"].values == x,
        adata_proc_p1.obs["condition"].values == "stimulated"
    )) >= 3 and np.sum(np.logical_and(
        adata_proc_p1.obs["clonotype_merged"].values == x,
        adata_proc_p1.obs["condition"].values == "unstimulated"
    )) >= 3
]
tab_clonotypes_p1_stim_filtered_bycell = {
    "Cd4": dict([
        ("_".join(k.lower().split(" ")) + "_score", [])
        if k in list(tcell_signatures.keys())
        else (k, [])
        for k in table_marker_keys
    ]),
    "Cd8": dict([
        ("_".join(k.lower().split(" ")) + "_score", [])
        if k in list(tcell_signatures.keys())
        else (k, [])
        for k in table_marker_keys
    ]),
}
tab_clonotypes_p1_unstim_filtered_bycell = {
    "Cd4": dict([
        ("_".join(k.lower().split(" ")) + "_score", [])
        if k in list(tcell_signatures.keys())
        else (k, [])
        for k in table_marker_keys
    ]),
    "Cd8": dict([
        ("_".join(k.lower().split(" ")) + "_score", [])
        if k in list(tcell_signatures.keys())
        else (k, [])
        for k in table_marker_keys
    ]),
}
adata_temp = adata_proc_p1.copy()
for k in keys:
    del adata_temp.uns[k]
ncell_cols_stim = np.max(tab_clonotypes_p1_filtered["n_stimulated"].values)
ncell_cols_unstim = np.max(tab_clonotypes_p1_filtered["n_unstimulated"].values)
for x in clonotypes_p1:
    adata = adata_temp[adata_temp.obs["clonotype_merged"].values == x]
    ncells_stim = np.sum(adata.obs["condition"].values == "stimulated")
    ncells_unstim = np.sum(adata.obs["condition"].values == "unstimulated")
    cell_type = tab_clonotypes_p1_filtered.loc[
        tab_clonotypes_p1_filtered["clonotype_merged"].values == x,
        "label_cell_type"
    ].values[0]
    for k, kk in zip(
        table_marker_keys, 
        list(tab_clonotypes_p1_unstim_filtered_bycell["Cd4"].keys())
    ):
        # stimulated
        tab_clonotypes_p1_stim_filtered_bycell[cell_type][kk].append(pd.DataFrame(dict(list({
            "clonotype_merged": x,
            "clonotypes_all_chains": [np.unique(adata.obs["clonotype_all_chains"].values)],
            "size": adata.shape[0],
            "n_stimulated": np.sum(adata.obs["condition"].values == "stimulated"),
            "n_unstimulated": np.sum(adata.obs["condition"].values == "unstimulated"),
        }.items()) + [
            (
                "cell_" + str(i),
                adata.obs[k.lower() + "_score"].values[adata.obs["condition"].values == "stimulated"][i]
                if (k.lower() + "_score" in adata.obs.keys() and i < ncells_stim) else 
                np.asarray(adata[adata.obs["condition"].values == "stimulated", k].X.todense()).flatten()[i]
                if i < ncells_stim else np.nan
            )
            for i in range(ncell_cols_stim)
        ]), index=[x]))
        # unstimulated
        tab_clonotypes_p1_unstim_filtered_bycell[cell_type][kk].append(pd.DataFrame(dict(list({
            "clonotype_merged": x,
            "clonotypes_all_chains": [np.unique(adata.obs["clonotype_all_chains"].values)],
            "size": adata.shape[0],
            "n_stimulated": np.sum(adata.obs["condition"].values == "stimulated"),
            "n_unstimulated": np.sum(adata.obs["condition"].values == "unstimulated"),
        }.items()) + [
            (
                "cell_" + str(i),
                adata.obs[k.lower() + "_score"].values[adata.obs["condition"].values == "unstimulated"][i]
                if (k.lower() + "_score" in adata.obs.keys() and i < ncells_unstim) else 
                np.asarray(adata[adata.obs["condition"].values == "unstimulated", k].X.todense()).flatten()[i]
                if i < ncells_unstim else np.nan
            )
            for i in range(ncell_cols_unstim)
        ]), index=[x]))
for k in ["Cd4", "Cd8"]:
    for kk in tab_clonotypes_p1_unstim_filtered_bycell[k].keys():
        tab_clonotypes_p1_unstim_filtered_bycell[k][kk] = pd.concat(tab_clonotypes_p1_unstim_filtered_bycell[k][kk], axis=0)
        tab_clonotypes_p1_stim_filtered_bycell[k][kk] = pd.concat(tab_clonotypes_p1_stim_filtered_bycell[k][kk], axis=0)
tab_clonotypes_p1_unstim_filtered_bycell["Cd8"]["IFNG"].sort_values("size").iloc[::-1].to_csv(dir_tables+'fig2a_cd8_unstim.csv', sep=";")
tab_clonotypes_p1_unstim_filtered_bycell["Cd4"]["IFNG"].sort_values("size").iloc[::-1].to_csv(dir_tables+'fig2b_cd4_unstim.csv', sep=";")
tab_clonotypes_p1_stim_filtered_bycell["Cd8"]["IFNG"].sort_values("size").iloc[::-1].to_csv(dir_tables+'fig2a_cd8_stim.csv', sep=";")
tab_clonotypes_p1_stim_filtered_bycell["Cd4"]["IFNG"].sort_values("size").iloc[::-1].to_csv(dir_tables+'fig2b_cd4_stim.csv', sep=";")

# %% [code]
import statsmodels.stats.weightstats

clonotype_label = "clonotype_merged"
cell_type = "Cd4"
panel_id = "fig2b_cd4"
idx = np.array([
    x in tab_clonotypes_p1_filtered.loc[
        tab_clonotypes_p1_filtered["label_cell_type"].values == cell_type, 
        clonotype_label
    ].values and y != "None" and z != "None" and
    tab_clonotypes_p1_filtered.loc[x, "n_stimulated"] >= 3 and
    tab_clonotypes_p1_filtered.loc[x, "n_unstimulated"] >= 3
    for x, y, z in zip(
        adata_proc_p1.obs[clonotype_label].values,
        adata_proc_p1.obs["TRA_1_cdr3"].values,
        adata_proc_p1.obs["TRB_1_cdr3"].values
    )
])
tab = adata_proc_p1[idx, :].obs.copy()
tab["IFNG"] = np.asarray(adata_proc_p1[idx, "IFNG"].X.todense()).flatten()
categ = tab.groupby(clonotype_label).size().reset_index().sort_values(0).iloc[::-1][clonotype_label].values.tolist()
tab[clonotype_label] = pd.Categorical(
    tab[clonotype_label].values.tolist(),
    categories=categ
)

significance = pd.DataFrame({
    "clonotype": tab[clonotype_label].values.categories,
    "TRA_1_cdr3": [
        np.unique(tab.loc[tab[clonotype_label].values == x, "TRA_1_cdr3"].values)
        for x in tab[clonotype_label].values.categories
    ],
    "TRB_1_cdr3": [
        np.unique(tab.loc[tab[clonotype_label].values == x, "TRB_1_cdr3"].values)
        for x in tab[clonotype_label].values.categories
    ],
    "TRA_1_v_gene": [
        np.unique(tab.loc[tab[clonotype_label].values == x, "TRA_1_v_gene"].values)
        for x in tab[clonotype_label].values.categories
    ],
    "TRB_1_v_gene": [
        np.unique(tab.loc[tab[clonotype_label].values == x, "TRB_1_v_gene"].values)
        for x in tab[clonotype_label].values.categories
    ],
    "TRA_1_d_gene": [
        np.unique(tab.loc[tab[clonotype_label].values == x, "TRA_1_d_gene"].values)
        for x in tab[clonotype_label].values.categories
    ],
    "TRB_1_d_gene": [
        np.unique(tab.loc[tab[clonotype_label].values == x, "TRB_1_d_gene"].values)
        for x in tab[clonotype_label].values.categories
    ],
    "TRA_1_j_gene": [
        np.unique(tab.loc[tab[clonotype_label].values == x, "TRA_1_j_gene"].values)
        for x in tab[clonotype_label].values.categories
    ],
    "TRB_1_j_gene": [
        np.unique(tab.loc[tab[clonotype_label].values == x, "TRB_1_j_gene"].values)
        for x in tab[clonotype_label].values.categories
    ],
    "n_stimulated": [
        np.sum(np.logical_and(
            tab[clonotype_label].values == x,
            tab["condition"].values == "stimulated"
        ))
        for x in tab[clonotype_label].values.categories
    ],
    "n_unstimulated": [
        np.sum(np.logical_and(
            tab[clonotype_label].values == x,
            tab["condition"].values == "unstimulated"
        ))
        for x in tab[clonotype_label].values.categories
    ],
    "pval": [
        statsmodels.stats.weightstats.ttest_ind(
            x1=tab.loc[np.logical_and(
                tab[clonotype_label].values == x,
                tab["condition"].values == "unstimulated"
            ), "IFNG"].values,
            x2=tab.loc[np.logical_and(
                tab[clonotype_label].values == x,
                tab["condition"].values == "stimulated"
            ), "IFNG"].values,
            alternative='smaller', usevar='pooled'
        )[1]
        for x in tab[clonotype_label].values.categories
    ]
})
significance["qval"] = de.stats.correct(pvals=significance["pval"].values, method="fdr_bh")
significance["stars"] = [
    "" if x >= 0.05 or np.isnan(x)
    else "*" if x >= 0.01
    else "**" if x >= 0.001
    else "***"
    for x in significance["qval"].values
]
tab[clonotype_label] = [
    x if significance.loc[significance["clonotype"].values == x, "stars"].values[0] == ""
    else "(" + significance.loc[significance["clonotype"].values == x, "stars"].values[0] + ") " + x
    for x in tab[clonotype_label].values
]
categ = tab.groupby(clonotype_label).size().reset_index().sort_values(0).iloc[::-1][clonotype_label].values.tolist()
tab[clonotype_label] = pd.Categorical(
    tab[clonotype_label].values.tolist(),
    categories=categ
)

fig, ax = plt.subplots(
    nrows=3, ncols=1, figsize=(10,7), 
    gridspec_kw={'height_ratios': [1, 1, 3]}
)
sb.stripplot(
    data=tab,
    x=clonotype_label,
    hue="condition",
    y="IFNG",
    ax=ax[2],
    palette=["darkred", "grey"],
)
ax[2].set_xticklabels(ax[2].get_xticklabels(), rotation=90)
ax[2].set_xlabel("clonotype")
sb.boxplot(data=tab,
    x=clonotype_label,
    hue="condition",
    y="IFNG",
    ax=ax[1],
    palette=["darkred", "grey"],
)
ax[1].set_xticklabels([])
ax[1].set_xlabel("")
ax[1].legend_.remove()
tab2 = tab.groupby(clonotype_label).size().reset_index()
tab2[clonotype_label] = pd.Categorical(
    tab2[clonotype_label].values.tolist(),
    categories=tab[clonotype_label].values.categories.tolist()
)
tab2["size"] = tab2.loc[:, 0].values
sb.barplot(
    data=tab2,
    x=clonotype_label,
    y="size",
    palette="viridis",
    ax=ax[0],
)
ax[0].set_xticklabels([])
ax[0].set_xlabel("")
ax[2].legend(bbox_to_anchor=(1.2, 1))
plt.tight_layout()
plt.subplots_adjust(hspace=0.05)
plt.savefig(dir_panels + panel_id + ".pdf")
plt.show()
significance.to_csv(dir_panels + panel_id + ".csv")
significance.loc[significance["qval"].values < 1.1, :]

# %% [code]
import statsmodels.stats.weightstats

clonotype_label = "clonotype_merged"
cell_type = "Cd8"
panel_id = "fig2a_cd8"
idx = np.array([
    x in tab_clonotypes_p1_filtered.loc[
        tab_clonotypes_p1_filtered["label_cell_type"].values == cell_type, 
        clonotype_label
    ].values and y != "None" and z != "None" and
    tab_clonotypes_p1_filtered.loc[x, "n_stimulated"] >= 3 and
    tab_clonotypes_p1_filtered.loc[x, "n_unstimulated"] >= 3
    for x, y, z in zip(
        adata_proc_p1.obs[clonotype_label].values,
        adata_proc_p1.obs["TRA_1_cdr3"].values,
        adata_proc_p1.obs["TRB_1_cdr3"].values
    )
])
tab = adata_proc_p1[idx, :].obs.copy()
tab["IFNG"] = np.asarray(adata_proc_p1[idx, "IFNG"].X.todense()).flatten()
categ = tab.groupby(clonotype_label).size().reset_index().sort_values(0).iloc[::-1][clonotype_label].values.tolist()
tab[clonotype_label] = pd.Categorical(
    tab[clonotype_label].values.tolist(),
    categories=categ
)

significance = pd.DataFrame({
    "clonotype": tab[clonotype_label].values.categories,
    "TRA_1_cdr3": [
        np.unique(tab.loc[tab[clonotype_label].values == x, "TRA_1_cdr3"].values)
        for x in tab[clonotype_label].values.categories
    ],
    "TRB_1_cdr3": [
        np.unique(tab.loc[tab[clonotype_label].values == x, "TRB_1_cdr3"].values)
        for x in tab[clonotype_label].values.categories
    ],
    "TRA_1_v_gene": [
        np.unique(tab.loc[tab[clonotype_label].values == x, "TRA_1_v_gene"].values)
        for x in tab[clonotype_label].values.categories
    ],
    "TRB_1_v_gene": [
        np.unique(tab.loc[tab[clonotype_label].values == x, "TRB_1_v_gene"].values)
        for x in tab[clonotype_label].values.categories
    ],
    "TRA_1_d_gene": [
        np.unique(tab.loc[tab[clonotype_label].values == x, "TRA_1_d_gene"].values)
        for x in tab[clonotype_label].values.categories
    ],
    "TRB_1_d_gene": [
        np.unique(tab.loc[tab[clonotype_label].values == x, "TRB_1_d_gene"].values)
        for x in tab[clonotype_label].values.categories
    ],
    "TRA_1_j_gene": [
        np.unique(tab.loc[tab[clonotype_label].values == x, "TRA_1_j_gene"].values)
        for x in tab[clonotype_label].values.categories
    ],
    "TRB_1_j_gene": [
        np.unique(tab.loc[tab[clonotype_label].values == x, "TRB_1_j_gene"].values)
        for x in tab[clonotype_label].values.categories
    ],
    "n_stimulated": [
        np.sum(np.logical_and(
            tab[clonotype_label].values == x,
            tab["condition"].values == "stimulated"
        ))
        for x in tab[clonotype_label].values.categories
    ],
    "n_unstimulated": [
        np.sum(np.logical_and(
            tab[clonotype_label].values == x,
            tab["condition"].values == "unstimulated"
        ))
        for x in tab[clonotype_label].values.categories
    ],
    "pval": [
        statsmodels.stats.weightstats.ttest_ind(
            x1=tab.loc[np.logical_and(
                tab[clonotype_label].values == x,
                tab["condition"].values == "unstimulated"
            ), "IFNG"].values,
            x2=tab.loc[np.logical_and(
                tab[clonotype_label].values == x,
                tab["condition"].values == "stimulated"
            ), "IFNG"].values,
            alternative='smaller', usevar='pooled'
        )[1]
        for x in tab[clonotype_label].values.categories
    ]
})
significance["qval"] = de.stats.correct(pvals=significance["pval"].values, method="fdr_bh")
significance["stars"] = [
    "" if x >= 0.05 or np.isnan(x)
    else "*" if x >= 0.01
    else "**" if x >= 0.001
    else "***"
    for x in significance["qval"].values
]
tab[clonotype_label] = [
    x if significance.loc[significance["clonotype"].values == x, "stars"].values[0] == ""
    else "(" + significance.loc[significance["clonotype"].values == x, "stars"].values[0] + ") " + x
    for x in tab[clonotype_label].values
]
categ = tab.groupby(clonotype_label).size().reset_index().sort_values(0).iloc[::-1][clonotype_label].values.tolist()
tab[clonotype_label] = pd.Categorical(
    tab[clonotype_label].values.tolist(),
    categories=categ
)

fig, ax = plt.subplots(
    nrows=3, ncols=1, figsize=(16,7), 
    gridspec_kw={'height_ratios': [1, 1, 3]}
)
sb.stripplot(
    data=tab,
    x=clonotype_label,
    hue="condition",
    y="IFNG",
    ax=ax[2],
    palette=["darkred", "grey"],
)
ax[2].set_xticklabels(ax[2].get_xticklabels(), rotation=90)
ax[2].set_xlabel("clonotype")
sb.boxplot(data=tab,
    x=clonotype_label,
    hue="condition",
    y="IFNG",
    ax=ax[1],
    palette=["darkred", "grey"],
)
ax[1].set_xticklabels([])
ax[1].set_xlabel("")
ax[1].legend_.remove()
tab2 = tab.groupby(clonotype_label).size().reset_index()
tab2[clonotype_label] = pd.Categorical(
    tab2[clonotype_label].values.tolist(),
    categories=tab[clonotype_label].values.categories.tolist()
)
tab2["size"] = tab2.loc[:, 0].values
sb.barplot(
    data=tab2,
    x=clonotype_label,
    y="size",
    palette="viridis",
    ax=ax[0],
)
ax[0].set_xticklabels([])
ax[0].set_xlabel("")
ax[2].legend(bbox_to_anchor=(1.1, 1))
plt.tight_layout()
plt.subplots_adjust(hspace=0.05)
plt.savefig(dir_panels + panel_id + ".pdf")
plt.show()
significance.to_csv(dir_panels + panel_id + ".csv")
significance.loc[significance["qval"].values < 0.05, :]

# %% [code]
import statsmodels.stats.weightstats

clonotype_label = "clonotype"
cell_type = "Cd4"
panel_id = "sfig4b_cd4"
scores = ['ifn response_score', 'proliferation_score']
idx = np.array([
    x in tab_clonotypes_p1_filtered.loc[
        tab_clonotypes_p1_filtered["label_cell_type"].values == cell_type, 
        clonotype_label
    ].values and 
    tab_clonotypes_p1_filtered.loc[x, "n_stimulated"] >= 3 and
    tab_clonotypes_p1_filtered.loc[x, "n_unstimulated"] >= 3
    for x in adata_proc_p1.obs[clonotype_label].values
])
tab = adata_proc_p1[idx, :].obs.copy()
tab["IFNG"] = np.asarray(adata_proc_p1[idx, "IFNG"].X.todense()).flatten()
categ = tab.groupby(clonotype_label).size().reset_index().sort_values(0).iloc[::-1][clonotype_label].values.tolist()
tab[clonotype_label] = pd.Categorical(
    tab[clonotype_label].values.tolist(),
    categories=categ
)

significance = pd.DataFrame({
    "clonotype": tab[clonotype_label].values.categories,
    "TRA_1_cdr3": [
        np.unique(tab.loc[tab[clonotype_label].values == x, "TRA_1_cdr3"].values)
        for x in tab[clonotype_label].values.categories
    ],
    "TRB_1_cdr3": [
        np.unique(tab.loc[tab[clonotype_label].values == x, "TRB_1_cdr3"].values)
        for x in tab[clonotype_label].values.categories
    ],
    "TRA_1_v_gene": [
        np.unique(tab.loc[tab[clonotype_label].values == x, "TRA_1_v_gene"].values)
        for x in tab[clonotype_label].values.categories
    ],
    "TRB_1_v_gene": [
        np.unique(tab.loc[tab[clonotype_label].values == x, "TRB_1_v_gene"].values)
        for x in tab[clonotype_label].values.categories
    ],
    "TRA_1_d_gene": [
        np.unique(tab.loc[tab[clonotype_label].values == x, "TRA_1_d_gene"].values)
        for x in tab[clonotype_label].values.categories
    ],
    "TRB_1_d_gene": [
        np.unique(tab.loc[tab[clonotype_label].values == x, "TRB_1_d_gene"].values)
        for x in tab[clonotype_label].values.categories
    ],
    "TRA_1_j_gene": [
        np.unique(tab.loc[tab[clonotype_label].values == x, "TRA_1_j_gene"].values)
        for x in tab[clonotype_label].values.categories
    ],
    "TRB_1_j_gene": [
        np.unique(tab.loc[tab[clonotype_label].values == x, "TRB_1_j_gene"].values)
        for x in tab[clonotype_label].values.categories
    ],
    "n_stimulated": [
        np.sum(np.logical_and(
            tab[clonotype_label].values == x,
            tab["condition"].values == "stimulated"
        ))
        for x in tab[clonotype_label].values.categories
    ],
    "n_unstimulated": [
        np.sum(np.logical_and(
            tab[clonotype_label].values == x,
            tab["condition"].values == "unstimulated"
        ))
        for x in tab[clonotype_label].values.categories
    ],
    "pval_score1": [
        statsmodels.stats.weightstats.ttest_ind(
            x1=tab.loc[np.logical_and(
                tab[clonotype_label].values == x,
                tab["condition"].values == "unstimulated"
            ), scores[0]].values,
            x2=tab.loc[np.logical_and(
                tab[clonotype_label].values == x,
                tab["condition"].values == "stimulated"
            ), scores[0]].values,
            alternative='smaller', usevar='pooled'
        )[1]
        for x in tab[clonotype_label].values.categories
    ],
    "pval_score2": [
        statsmodels.stats.weightstats.ttest_ind(
            x1=tab.loc[np.logical_and(
                tab[clonotype_label].values == x,
                tab["condition"].values == "unstimulated"
            ), scores[1]].values,
            x2=tab.loc[np.logical_and(
                tab[clonotype_label].values == x,
                tab["condition"].values == "stimulated"
            ), scores[1]].values,
            alternative='smaller', usevar='pooled'
        )[1]
        for x in tab[clonotype_label].values.categories
    ],
    "pval_ifng": [
        statsmodels.stats.weightstats.ttest_ind(
            x1=tab.loc[np.logical_and(
                tab[clonotype_label].values == x,
                tab["condition"].values == "unstimulated"
            ), "IFNG"].values,
            x2=tab.loc[np.logical_and(
                tab[clonotype_label].values == x,
                tab["condition"].values == "stimulated"
            ), "IFNG"].values,
            alternative='smaller', usevar='pooled'
        )[1]
        for x in tab[clonotype_label].values.categories
    ]
})
significance["qval_ifng"] = de.stats.correct(pvals=significance["pval_ifng"].values, method="fdr_bh")
significance["qval_score1"] = de.stats.correct(pvals=significance["pval_score1"].values, method="fdr_bh")
significance["qval_score2"] = de.stats.correct(pvals=significance["pval_score2"].values, method="fdr_bh")
significance["stars_ifng"] = [
    "" if x >= 0.05 or np.isnan(x)
    else "*" if x >= 0.01
    else "**" if x >= 0.001
    else "***"
    for x in significance["qval_ifng"].values
]
significance["stars_score1"] = [
    "" if x >= 0.05 or np.isnan(x)
    else "*" if x >= 0.01
    else "**" if x >= 0.001
    else "***"
    for x in significance["qval_score1"].values
]
significance["stars_score2"] = [
    "" if x >= 0.05 or np.isnan(x)
    else "*" if x >= 0.01
    else "**" if x >= 0.001
    else "***"
    for x in significance["qval_score2"].values
]
tab["stars_ifng"] = [
    significance.loc[significance["clonotype"].values == x, "stars_ifng"].values[0]
    for x in tab[clonotype_label].values
]
tab["stars_score1"] = [
    significance.loc[significance["clonotype"].values == x, "stars_score1"].values[0]
    for x in tab[clonotype_label].values
]
tab["stars_score2"] = [
    significance.loc[significance["clonotype"].values == x, "stars_score2"].values[0]
    for x in tab[clonotype_label].values
]
tab[clonotype_label] = [
    x if significance.loc[significance["clonotype"].values == x, "stars_ifng"].values[0] == ""
    else "(" + significance.loc[significance["clonotype"].values == x, "stars_ifng"].values[0] + ") " + x
    for x in tab[clonotype_label].values
]
categ = tab.groupby(clonotype_label).size().reset_index().sort_values(0).iloc[::-1][clonotype_label].values.tolist()
tab[clonotype_label] = pd.Categorical(
    tab[clonotype_label].values.tolist(),
    categories=categ
)
tab = tab.sort_values(clonotype_label).copy()

fig, ax = plt.subplots(
    nrows=7, ncols=1, figsize=(10,14), 
    gridspec_kw={'height_ratios': [1, 1, 2, 1, 2, 1, 2]}
)
tab2 = tab.groupby(clonotype_label).size().reset_index()
tab2[clonotype_label] = pd.Categorical(
    tab2[clonotype_label].values.tolist(),
    categories=tab[clonotype_label].values.categories.tolist()
)
tab2["size"] = tab2.loc[:, 0].values
sb.barplot(
    data=tab2,
    x=clonotype_label,
    y="size",
    palette="viridis",
    ax=ax[0],
)
ax[0].set_xticklabels([])
ax[0].set_xlabel("")

for i, x in enumerate(scores + ["IFNG"]):
    sb.boxplot(data=tab,
        x=clonotype_label,
        hue="condition",
        y=x,
        ax=ax[1+i*2],
        palette=["darkred", "grey"],
    )
    ax[1+i*2].set_xticklabels([])
    ax[1+i*2].set_xlabel("")
    ax[1+i*2].legend_.remove()

    sb.stripplot(
        data=tab,
        x=clonotype_label,
        hue="condition",
        y=x,
        ax=ax[2+i*2],
        palette=["darkred", "grey"]
    )
    if x == 'ifn response_score':
        ax[1+i*2].set_ylabel("")
        ax[2+i*2].set_ylabel("IFN response score")
    if x == 'proliferation_score':
        ax[1+i*2].set_ylabel("")
        ax[2+i*2].set_ylabel("Proliferation score")
    if x == 'IFNG':
        ax[1+i*2].set_ylabel("")
        ax[2+i*2].set_ylabel("IFNG expression")
    if i == 0:
        ax[2+i*2].set_xticklabels(significance["stars_score1"].values, rotation=0)
        ax[2+i*2].set_xlabel("")
    elif i == 1:
        ax[2+i*2].set_xticklabels(significance["stars_score2"].values, rotation=0)
        ax[2+i*2].set_xlabel("")
    else:
        ax[2+i*2].set_xticklabels(ax[2+i*2].get_xticklabels(), rotation=90)
        ax[2+i*2].set_xlabel("clonotype")
        
    ax[2+i*2].legend(bbox_to_anchor=(1.2, 1))
plt.tight_layout()
plt.subplots_adjust(hspace=0.15)
plt.savefig(dir_panels + panel_id + ".pdf")
plt.show()
significance.to_csv(dir_panels + panel_id + ".csv")

# %% [code]
import statsmodels.stats.weightstats

clonotype_label = "clonotype"
cell_type = "Cd8"
panel_id = "sfig4a_cd8"
scores = ['cd8 cytotoxic_score', 'cd8 cytokine_score']
idx = np.array([
    x in tab_clonotypes_p1_filtered.loc[
        tab_clonotypes_p1_filtered["label_cell_type"].values == cell_type, 
        clonotype_label
    ].values and 
    tab_clonotypes_p1_filtered.loc[x, "n_stimulated"] >= 3 and
    tab_clonotypes_p1_filtered.loc[x, "n_unstimulated"] >= 3
    for x in adata_proc_p1.obs[clonotype_label].values
])
tab = adata_proc_p1[idx, :].obs.copy()
tab["IFNG"] = np.asarray(adata_proc_p1[idx, "IFNG"].X.todense()).flatten()
categ = tab.groupby(clonotype_label).size().reset_index().sort_values(0).iloc[::-1][clonotype_label].values.tolist()
tab[clonotype_label] = pd.Categorical(
    tab[clonotype_label].values.tolist(),
    categories=categ
)

significance = pd.DataFrame({
    "clonotype": tab[clonotype_label].values.categories,
    "TRA_1_cdr3": [
        np.unique(tab.loc[tab[clonotype_label].values == x, "TRA_1_cdr3"].values)
        for x in tab[clonotype_label].values.categories
    ],
    "TRB_1_cdr3": [
        np.unique(tab.loc[tab[clonotype_label].values == x, "TRB_1_cdr3"].values)
        for x in tab[clonotype_label].values.categories
    ],
    "TRA_1_v_gene": [
        np.unique(tab.loc[tab[clonotype_label].values == x, "TRA_1_v_gene"].values)
        for x in tab[clonotype_label].values.categories
    ],
    "TRB_1_v_gene": [
        np.unique(tab.loc[tab[clonotype_label].values == x, "TRB_1_v_gene"].values)
        for x in tab[clonotype_label].values.categories
    ],
    "TRA_1_d_gene": [
        np.unique(tab.loc[tab[clonotype_label].values == x, "TRA_1_d_gene"].values)
        for x in tab[clonotype_label].values.categories
    ],
    "TRB_1_d_gene": [
        np.unique(tab.loc[tab[clonotype_label].values == x, "TRB_1_d_gene"].values)
        for x in tab[clonotype_label].values.categories
    ],
    "TRA_1_j_gene": [
        np.unique(tab.loc[tab[clonotype_label].values == x, "TRA_1_j_gene"].values)
        for x in tab[clonotype_label].values.categories
    ],
    "TRB_1_j_gene": [
        np.unique(tab.loc[tab[clonotype_label].values == x, "TRB_1_j_gene"].values)
        for x in tab[clonotype_label].values.categories
    ],
    "n_stimulated": [
        np.sum(np.logical_and(
            tab[clonotype_label].values == x,
            tab["condition"].values == "stimulated"
        ))
        for x in tab[clonotype_label].values.categories
    ],
    "n_unstimulated": [
        np.sum(np.logical_and(
            tab[clonotype_label].values == x,
            tab["condition"].values == "unstimulated"
        ))
        for x in tab[clonotype_label].values.categories
    ],
    "pval_score1": [
        statsmodels.stats.weightstats.ttest_ind(
            x1=tab.loc[np.logical_and(
                tab[clonotype_label].values == x,
                tab["condition"].values == "unstimulated"
            ), scores[0]].values,
            x2=tab.loc[np.logical_and(
                tab[clonotype_label].values == x,
                tab["condition"].values == "stimulated"
            ), scores[0]].values,
            alternative='smaller', usevar='pooled'
        )[1]
        for x in tab[clonotype_label].values.categories
    ],
    "pval_score2": [
        statsmodels.stats.weightstats.ttest_ind(
            x1=tab.loc[np.logical_and(
                tab[clonotype_label].values == x,
                tab["condition"].values == "unstimulated"
            ), scores[1]].values,
            x2=tab.loc[np.logical_and(
                tab[clonotype_label].values == x,
                tab["condition"].values == "stimulated"
            ), scores[1]].values,
            alternative='smaller', usevar='pooled'
        )[1]
        for x in tab[clonotype_label].values.categories
    ],
    "pval_ifng": [
        statsmodels.stats.weightstats.ttest_ind(
            x1=tab.loc[np.logical_and(
                tab[clonotype_label].values == x,
                tab["condition"].values == "unstimulated"
            ), "IFNG"].values,
            x2=tab.loc[np.logical_and(
                tab[clonotype_label].values == x,
                tab["condition"].values == "stimulated"
            ), "IFNG"].values,
            alternative='smaller', usevar='pooled'
        )[1]
        for x in tab[clonotype_label].values.categories
    ]
})
significance["qval_ifng"] = de.stats.correct(pvals=significance["pval_ifng"].values, method="fdr_bh")
significance["qval_score1"] = de.stats.correct(pvals=significance["pval_score1"].values, method="fdr_bh")
significance["qval_score2"] = de.stats.correct(pvals=significance["pval_score2"].values, method="fdr_bh")
significance["stars_ifng"] = [
    "" if x >= 0.05 or np.isnan(x)
    else "*" if x >= 0.01
    else "**" if x >= 0.001
    else "***"
    for x in significance["qval_ifng"].values
]
significance["stars_score1"] = [
    "" if x >= 0.05 or np.isnan(x)
    else "*" if x >= 0.01
    else "**" if x >= 0.001
    else "***"
    for x in significance["qval_score1"].values
]
significance["stars_score2"] = [
    "" if x >= 0.05 or np.isnan(x)
    else "*" if x >= 0.01
    else "**" if x >= 0.001
    else "***"
    for x in significance["qval_score2"].values
]
tab["stars_ifng"] = [
    significance.loc[significance["clonotype"].values == x, "stars_ifng"].values[0]
    for x in tab[clonotype_label].values
]
tab["stars_score1"] = [
    significance.loc[significance["clonotype"].values == x, "stars_score1"].values[0]
    for x in tab[clonotype_label].values
]
tab["stars_score2"] = [
    significance.loc[significance["clonotype"].values == x, "stars_score2"].values[0]
    for x in tab[clonotype_label].values
]
tab[clonotype_label] = [
    x if significance.loc[significance["clonotype"].values == x, "stars_ifng"].values[0] == ""
    else "(" + significance.loc[significance["clonotype"].values == x, "stars_ifng"].values[0] + ") " + x
    for x in tab[clonotype_label].values
]
categ = tab.groupby(clonotype_label).size().reset_index().sort_values(0).iloc[::-1][clonotype_label].values.tolist()
tab[clonotype_label] = pd.Categorical(
    tab[clonotype_label].values.tolist(),
    categories=categ
)
tab = tab.sort_values(clonotype_label).copy()

fig, ax = plt.subplots(
    nrows=7, ncols=1, figsize=(20,14), 
    gridspec_kw={'height_ratios': [1, 1, 2, 1, 2, 1, 2]}
)
tab2 = tab.groupby(clonotype_label).size().reset_index()
tab2[clonotype_label] = pd.Categorical(
    tab2[clonotype_label].values.tolist(),
    categories=tab[clonotype_label].values.categories.tolist()
)
tab2["size"] = tab2.loc[:, 0].values
sb.barplot(
    data=tab2,
    x=clonotype_label,
    y="size",
    palette="viridis",
    ax=ax[0],
)
ax[0].set_xticklabels([])
ax[0].set_xlabel("")

for i, x in enumerate(scores + ["IFNG"]):
    sb.boxplot(data=tab,
        x=clonotype_label,
        hue="condition",
        y=x,
        ax=ax[1+i*2],
        palette=["darkred", "grey"],
    )
    ax[1+i*2].set_xticklabels([])
    ax[1+i*2].set_xlabel("")
    ax[1+i*2].legend_.remove()

    sb.stripplot(
        data=tab,
        x=clonotype_label,
        hue="condition",
        y=x,
        ax=ax[2+i*2],
        palette=["darkred", "grey"]
    )
    if x == 'cd8 cytotoxic_score':
        ax[1+i*2].set_ylabel("")
        ax[2+i*2].set_ylabel("CD8 cytotoxic score")
    if x == 'cd8 cytokine_score':
        ax[1+i*2].set_ylabel("")
        ax[2+i*2].set_ylabel("CD8 cytokine score")
    if x == 'IFNG':
        ax[1+i*2].set_ylabel("")
        ax[2+i*2].set_ylabel("IFNG expression")
    if i == 0:
        ax[2+i*2].set_xticklabels(significance["stars_score1"].values, rotation=0)
        ax[2+i*2].set_xlabel("")
    elif i == 1:
        ax[2+i*2].set_xticklabels(significance["stars_score2"].values, rotation=0)
        ax[2+i*2].set_xlabel("")
    else:
        ax[2+i*2].set_xticklabels(ax[2+i*2].get_xticklabels(), rotation=90)
        ax[2+i*2].set_xlabel("clonotype")
        
    ax[2+i*2].legend(bbox_to_anchor=(1.1, 1))
plt.tight_layout()
plt.subplots_adjust(hspace=0.15)
plt.savefig(dir_panels + panel_id + ".pdf")
plt.show()
significance.to_csv(dir_panels + panel_id + ".csv")

# %% [code]
clonotypes_p2 = [
    x for x in np.unique(adata_proc_p2.obs["clonotype"].values)
    if np.sum(np.logical_and(
        adata_proc_p2.obs["clonotype"].values == x,
        adata_proc_p2.obs["condition"].values == "stimulated"
    )) >= 1 and np.sum(np.logical_and(
        adata_proc_p2.obs["clonotype"].values == x,
        adata_proc_p2.obs["condition"].values == "unstimulated"
    )) >= 1
]
tab_clonotypes_p2_stim_filtered_bycell = {
    "Cd4": dict([
        ("_".join(k.lower().split(" ")) + "_score", [])
        if k in list(tcell_signatures.keys())
        else (k, [])
        for k in table_marker_keys
    ]),
    "Cd8": dict([
        ("_".join(k.lower().split(" ")) + "_score", [])
        if k in list(tcell_signatures.keys())
        else (k, [])
        for k in table_marker_keys
    ]),
}
tab_clonotypes_p2_unstim_filtered_bycell = {
    "Cd4": dict([
        ("_".join(k.lower().split(" ")) + "_score", [])
        if k in list(tcell_signatures.keys())
        else (k, [])
        for k in table_marker_keys
    ]),
    "Cd8": dict([
        ("_".join(k.lower().split(" ")) + "_score", [])
        if k in list(tcell_signatures.keys())
        else (k, [])
        for k in table_marker_keys
    ]),
}

adata_temp = adata_proc_p2.copy()
for k in keys:
    del adata_temp.uns[k]
ncell_cols_stim = np.max(tab_clonotypes_p2_filtered["n_stimulated"].values)
ncell_cols_unstim = np.max(tab_clonotypes_p2_filtered["n_unstimulated"].values)
for x in clonotypes_p2:
    adata = adata_temp[adata_temp.obs["clonotype"].values == x]
    ncells_stim = np.sum(adata.obs["condition"].values == "stimulated")
    ncells_unstim = np.sum(adata.obs["condition"].values == "unstimulated")
    cell_type = tab_clonotypes_p2_filtered.loc[
        tab_clonotypes_p2_filtered["clonotype"].values == x,
        "label_cell_type"
    ].values[0]
    for k, kk in zip(
        table_marker_keys, 
        list(tab_clonotypes_p1_unstim_filtered_bycell["Cd4"].keys())
    ):
        # stimulated
        tab_clonotypes_p2_stim_filtered_bycell[cell_type][kk].append(pd.DataFrame(dict(list({
            "clonotype": x,
            "clonotypes_all_chains": [np.unique(adata.obs["clonotype_all_chains"].values)],
            "size": adata.shape[0],
            "n_stimulated": np.sum(adata.obs["condition"].values == "stimulated"),
            "n_unstimulated": np.sum(adata.obs["condition"].values == "unstimulated"),
        }.items()) + [
            (
                "cell_" + str(i),
                adata.obs[k.lower() + "_score"].values[adata.obs["condition"].values == "stimulated"][i]
                if (k.lower() + "_score" in adata.obs.keys() and i < ncells_stim) else 
                np.asarray(adata[adata.obs["condition"].values == "stimulated", k].X.todense()).flatten()[i]
                if i < ncells_stim else np.nan
            )
            for i in range(ncell_cols_stim)
        ]), index=[x]))
        # unstimulated
        tab_clonotypes_p2_unstim_filtered_bycell[cell_type][kk].append(pd.DataFrame(dict(list({
            "clonotype": x,
            "clonotypes_all_chains": [np.unique(adata.obs["clonotype_all_chains"].values)],
            "size": adata.shape[0],
            "n_stimulated": np.sum(adata.obs["condition"].values == "stimulated"),
            "n_unstimulated": np.sum(adata.obs["condition"].values == "unstimulated"),
        }.items()) + [
            (
                "cell_" + str(i),
                adata.obs[k.lower() + "_score"].values[adata.obs["condition"].values == "unstimulated"][i]
                if (k.lower() + "_score" in adata.obs.keys() and i < ncells_unstim) else 
                np.asarray(adata[adata.obs["condition"].values == "unstimulated", k].X.todense()).flatten()[i]
                if i < ncells_unstim else np.nan
            )
            for i in range(ncell_cols_unstim)
        ]), index=[x]))
for k in ["Cd4", "Cd8"]:
    for kk in tab_clonotypes_p2_unstim_filtered_bycell[k].keys():
        tab_clonotypes_p2_unstim_filtered_bycell[k][kk] = pd.concat(tab_clonotypes_p2_unstim_filtered_bycell[k][kk], axis=0)
        tab_clonotypes_p2_unstim_filtered_bycell[k][kk].sort_values("size").iloc[::-1].to_csv(dir_tables+'tab_clonotypes_p2_' + k.lower() + '_' + kk.lower() + '_unstim_filtered_bycell.csv', sep=";")

        tab_clonotypes_p2_stim_filtered_bycell[k][kk] = pd.concat(tab_clonotypes_p2_stim_filtered_bycell[k][kk], axis=0)
        tab_clonotypes_p2_stim_filtered_bycell[k][kk].sort_values("size").iloc[::-1].to_csv(dir_tables+'tab_clonotypes_p2_' + k.lower() + '_' + kk.lower() + '_stim_filtered_bycell.csv', sep=";")

# %% [markdown]
### Rest

# %% [code]
ir.tl.repertoire_overlap(adata_proc_p1, "sample_rna")
ir.tl.repertoire_overlap(adata_proc_p2, "sample_rna")

# %% [markdown]
## Sanitize cell type assignments based on clonotypes

# %% [markdown]
Only sanitize selected clonotypes.

# %% [markdown]
### P1

# %% [code]
fig, ax = plt.subplots(figsize=(30, 4))
sb.barplot(
    data=adata_proc_p1[[
        x in tab_clonotypes_p1_filtered.loc[
            tab_clonotypes_p1_filtered["label_cell_type"].values == "Cd4",
            "clonotype"
        ].values
        for x in adata_proc_p1.obs["clonotype"].values
    ]].obs.groupby(["clonotype", "cell_type"]).size().reset_index(),
    x="clonotype",
    hue="cell_type",
    y=0,
    ax=ax
)
ax.set_title("P1 Cd4 clonotypes")
plt.show()

fig, ax = plt.subplots(figsize=(30, 4))
sb.barplot(
    data=adata_proc_p1[[
        x in tab_clonotypes_p1_filtered.loc[
            tab_clonotypes_p1_filtered["label_cell_type"].values == "Cd8",
            "clonotype"
        ].values
        for x in adata_proc_p1.obs["clonotype"].values
    ]].obs.groupby(["clonotype", "cell_type"]).size().reset_index(),
    x="clonotype",
    hue="cell_type",
    y=0,
    ax=ax
)
ax.set_title("P1 Cd8 clonotypes")
plt.show()

# %% [markdown]
### P2

# %% [code]
fig, ax = plt.subplots(figsize=(30, 4))
sb.barplot(
    data=adata_proc_p2[[
        x in tab_clonotypes_p2_filtered.loc[
            tab_clonotypes_p2_filtered["label_cell_type"].values == "Cd4",
            "clonotype"
        ].values
        for x in adata_proc_p2.obs["clonotype"].values
    ]].obs.groupby(["clonotype", "cell_type"]).size().reset_index(),
    x="clonotype",
    hue="cell_type",
    y=0,
    ax=ax
)
ax.set_title("P2 Cd4 clonotypes")
plt.show()

fig, ax = plt.subplots(figsize=(30, 4))
sb.barplot(
    data=adata_proc_p2[[
        x in tab_clonotypes_p2_filtered.loc[
            tab_clonotypes_p2_filtered["label_cell_type"].values == "Cd8",
            "clonotype"
        ].values
        for x in adata_proc_p2.obs["clonotype"].values
    ]].obs.groupby(["clonotype", "cell_type"]).size().reset_index(),
    x="clonotype",
    hue="cell_type",
    y=0,
    ax=ax
)
ax.set_title("P2 Cd8 clonotypes")
plt.show()

# %% [markdown]
### Sanitize clonotype table

# %% [markdown]
#### P1

# %% [code]
clonotypes_to_sanitize_p1 = [
    y for x, y in zip(
        tab_clonotypes_p1_filtered["label_cell_type"].values,
        tab_clonotypes_p1_filtered["clonotype"].values
    )
    if np.unique(adata_proc_p1.obs["cell_type"].values[adata_proc_p1.obs["clonotype"].values == y]).shape[0] > 1
]

# %% [code]
fig, ax = plt.subplots(nrows=2, ncols=2, figsize=(10, 10))
sc.pl.umap(
    adata_proc_p1,
    color="cell_type",
    ax=ax[0, 0],
    show=False,
)
sc.pl.umap(
    adata_proc_p1,
    ax=ax[0, 1],
    show=False,
)
sc.pl.umap(
    adata_proc_p1[[
        x in clonotypes_to_sanitize_p1
        for x in adata_proc_p1.obs["clonotype"].values
    ]],
    size=20,
    color="clonotype",
    ax=ax[0, 1],
    show=False,
)
sc.pl.umap(
    adata_proc_p1,
    color="CD8_all_genes",
    ax=ax[1, 0],
    show=False,
)
sc.pl.umap(
    adata_proc_p1,
    color="CD8_all_genes",
    ax=ax[1, 1],
    show=False,
)
plt.tight_layout()
plt.show()

# %% [code]
tab_clonotypes_p1_filtered["label_cell_type_sanitised"] = [
    "Cd4" if y in clonotypes_to_sanitize_p1
    and np.mean(adata_proc_p1.obs["cell_type"].values[adata_proc_p1.obs["clonotype"].values == y] == "Cd4") > 0.5
    else x
    for x, y in zip(
        tab_clonotypes_p1_filtered["label_cell_type"].values,
        tab_clonotypes_p1_filtered["clonotype"].values
    )
]

# %% [markdown]
#### P2

# %% [code]
clonotypes_to_sanitize_p2 = [
    y for x, y in zip(
        tab_clonotypes_p2_filtered["label_cell_type"].values,
        tab_clonotypes_p2_filtered["clonotype"].values
    )
    if np.unique(adata_proc_p2.obs["cell_type"].values[adata_proc_p2.obs["clonotype"].values == y]).shape[0] > 1
]

# %% [code]
fig, ax = plt.subplots(nrows=2, ncols=2, figsize=(10, 10))
sc.pl.umap(
    adata_proc_p2,
    color="cell_type",
    ax=ax[0, 0],
    show=False,
)
sc.pl.umap(
    adata_proc_p2,
    ax=ax[0, 1],
    show=False,
)
sc.pl.umap(
    adata_proc_p2[[
        x in clonotypes_to_sanitize_p2
        for x in adata_proc_p2.obs["clonotype"].values
    ]],
    size=20,
    color="clonotype",
    ax=ax[0, 1],
    show=False,
)
sc.pl.umap(
    adata_proc_p2,
    color="CD8_all_genes",
    ax=ax[1, 0],
    show=False,
)
sc.pl.umap(
    adata_proc_p2,
    color="CD8_all_genes",
    ax=ax[1, 1],
    show=False,
)
plt.tight_layout()
plt.show()

# %% [code]
tab_clonotypes_p2_filtered["label_cell_type_sanitised"] = [
    "Cd4" if y in clonotypes_to_sanitize_p2 
    and np.mean(adata_proc_p2.obs["cell_type"].values[adata_proc_p2.obs["clonotype"].values == y] == "Cd4") > 0.5
    else x
    for x, y in zip(
        tab_clonotypes_p2_filtered["label_cell_type"].values,
        tab_clonotypes_p2_filtered["clonotype"].values
    )
]

# %% [markdown]
### Sanitize adata

# %% [code]
adata_proc_p1.obs["cell_type"] = [
    tab_clonotypes_p1_filtered["label_cell_type_sanitised"].values[
        tab_clonotypes_p1_filtered["clonotype"].values == y
    ][0]
    if y in clonotypes_to_sanitize_p1 else x
    for x, y in zip(
        adata_proc_p1.obs["cell_type"].values,
        adata_proc_p1.obs["clonotype"].values
    )
]

adata_proc_p1_unstim.obs["cell_type"] = [
    tab_clonotypes_p1_filtered["label_cell_type_sanitised"].values[
        tab_clonotypes_p1_filtered["clonotype"].values == y
    ][0]
    if y in clonotypes_to_sanitize_p1 else x
    for x, y in zip(
        adata_proc_p1_unstim.obs["cell_type"].values,
        adata_proc_p1_unstim.obs["clonotype"].values
    )
]

# %% [code]
adata_proc_p2.obs["cell_type"] = [
    tab_clonotypes_p2_filtered["label_cell_type_sanitised"].values[
        tab_clonotypes_p2_filtered["clonotype"].values == y
    ][0] 
    if y in clonotypes_to_sanitize_p2 else x
    for x, y in zip(
        adata_proc_p2.obs["cell_type"].values,
        adata_proc_p2.obs["clonotype"].values
    )
]

adata_proc_p2_unstim.obs["cell_type"] = [
    tab_clonotypes_p2_filtered["label_cell_type_sanitised"].values[
        tab_clonotypes_p2_filtered["clonotype"].values == y
    ][0] 
    if y in clonotypes_to_sanitize_p2 else x
    for x, y in zip(
        adata_proc_p2_unstim.obs["cell_type"].values,
        adata_proc_p2_unstim.obs["clonotype"].values
    )
]

# %% [code]
fig, axs = plt.subplots(nrows=1, ncols=3, figsize=(12, 4))
sc.pl.umap(adata_proc_p1, color=["leiden"], ax=axs[0], show=False)
sc.pl.umap(adata_proc_p1, color=["cell_type"], ax=axs[1], show=False)
sc.pl.umap(adata_proc_p1, color=["condition"], ax=axs[2], show=False)
axs[0].set_title("Leiden cluster")
axs[1].set_title("T cell type")
axs[2].set_title("Condition")
axs[0].set_xlabel(" ")
axs[1].set_xlabel(" ")
axs[2].set_xlabel(" ")
axs[0].set_ylabel(" ")
axs[1].set_ylabel(" ")
axs[2].set_ylabel(" ")
axs[0].legend_.remove()
axs[1].legend_.remove()
axs[2].legend_.remove()
plt.subplots_adjust(wspace=0.05)
plt.savefig(dir_panels + "fig1b.pdf")
plt.show()

# %% [code]
sc.pl.umap(
    adata_proc_p1, 
    color=["cell_type"],
)
sc.pl.umap(
    adata_proc_p1, 
    color=["CD8_all_genes", "CD8A", "CD8B"]
)

# %% [code]
fig, axs = plt.subplots(nrows=1, ncols=3, figsize=(12, 4))
sc.pl.umap(adata_proc_p2, color=["leiden"], ax=axs[0], show=False)
sc.pl.umap(adata_proc_p2, color=["cell_type"], ax=axs[1], show=False)
sc.pl.umap(adata_proc_p2, color=["condition"], ax=axs[2], show=False)
axs[0].set_title("Leiden cluster")
axs[1].set_title("T cell type")
axs[2].set_title("Condition")
axs[0].set_xlabel(" ")
axs[1].set_xlabel(" ")
axs[2].set_xlabel(" ")
axs[0].set_ylabel(" ")
axs[1].set_ylabel(" ")
axs[2].set_ylabel(" ")
axs[0].legend_.remove()
axs[1].legend_.remove()
axs[2].legend_.remove()
plt.subplots_adjust(wspace=0.05)
plt.savefig(dir_panels + "sfig2a.pdf")
plt.show()

# %% [code]
sc.pl.umap(
    adata_proc_p2, 
    color=["cell_type"]
)
sc.pl.umap(
    adata_proc_p2, 
    color=["CD8_all_genes", "CD8A", "CD8B"]
)

# %% [markdown]
## Recluster sanititzed Cd4 and Cd8 cells separately in each patient

# %% [markdown]
### Compute clusterings

# %% [code]
if bool_recomp:    
    for x in ["Cd4", "Cd8"]:
        adata_temp = adata_proc_p1_unstim[adata_proc_p1_unstim.obs["cell_type"].values == x].copy()
        #adata_temp.X = adata_proc_p1[adata_temp.obs_names].raw.X
        #sc.pp.filter_genes(adata_temp, min_cells=10)
        #sc.pp.normalize_per_cell(adata_temp, counts_per_cell_after=1000)
        #sc.pp.log1p(adata_temp)
        sc.pp.highly_variable_genes(adata_temp)
        sc.pp.pca(adata_temp, use_highly_variable=True)
        sc.pp.neighbors(adata_temp, knn=50)
        if bool_recluster == True:
            sc.tl.leiden(adata_temp, resolution=0.8, random_state=0)
            pd.DataFrame(adata_temp.obs).to_csv(path_or_buf=dir_anndata+"adata_proc_p1_unstim_"+x+"_obs.csv")
        else:
            obs = pd.read_csv(dir_anndata+"adata_proc_p1_unstim_"+x+"_obs.csv")
            adata_temp.obs['leiden'] = pd.Categorical(
                obs['leiden'].values.astype(str), 
                categories=np.sort(np.unique(obs['leiden'].values.astype(str)))
            )
        adata_proc_p1_unstim.obs["leiden_" + x] = [
            adata_temp.obs.loc[x, "leiden"] if x in adata_temp.obs_names else "nan"
            for x in adata_proc_p1_unstim.obs_names
        ]
        adata_proc_p1.obs["leiden_" + x] = [
            adata_temp.obs.loc[x, "leiden"] if x in adata_temp.obs_names else "nan"
            for x in adata_proc_p1.obs_names
        ]

        adata_temp = adata_proc_p2_unstim[adata_proc_p2_unstim.obs["cell_type"].values == x].copy()
        #adata_temp.X = adata_proc_p2[adata_temp.obs_names].raw.X
        #sc.pp.filter_genes(adata_temp, min_cells=10)
        #sc.pp.normalize_per_cell(adata_temp, counts_per_cell_after=1000)
        #sc.pp.log1p(adata_temp)
        sc.pp.highly_variable_genes(adata_temp)
        sc.pp.pca(adata_temp, use_highly_variable=True)
        sc.pp.neighbors(adata_temp, knn=50)
        if bool_recluster == True:
            sc.tl.leiden(adata_temp, resolution=1., random_state=0)
            pd.DataFrame(adata_temp.obs).to_csv(path_or_buf=dir_anndata+"adata_proc_p2_unstim_"+x+"_obs.csv")
        else:
            obs = pd.read_csv(dir_anndata+"adata_proc_p2_unstim_"+x+"_obs.csv")
            adata_temp.obs['leiden'] = pd.Categorical(
                obs['leiden'].values.astype(str), 
                categories=np.sort(np.unique(obs['leiden'].values.astype(str)))
            )
        adata_proc_p2_unstim.obs["leiden_" + x] = [
            adata_temp.obs.loc[x, "leiden"] if x in adata_temp.obs_names else "nan"
            for x in adata_proc_p2_unstim.obs_names
        ]
        adata_proc_p2.obs["leiden_" + x] = [
            adata_temp.obs.loc[x, "leiden"] if x in adata_temp.obs_names else "nan"
            for x in adata_proc_p2.obs_names
        ]

# %% [markdown]
### Cd4 cell states

# %% [code]
gene_ids = adata_proc_p1.var_names[np.asarray(adata_proc_p1[
    np.logical_and(
        adata_proc_p1.obs["leiden_Cd4"].values != "nan",
        adata_proc_p1.obs["condition"] == "unstimulated"
    )
].X.mean(axis=0)).flatten() > np.log(1.0+0.5)]  # >logp1(mean = 0.5)
tab = pd.concat([
    adata_proc_p1[
        np.logical_and(
            adata_proc_p1.obs["leiden_Cd4"].values != "nan",
            adata_proc_p1.obs["condition"] == "unstimulated"
        )
    ].obs["leiden_Cd4"],
    pd.DataFrame(
        adata_proc_p1[
            np.logical_and(
                adata_proc_p1.obs["leiden_Cd4"].values != "nan",
                adata_proc_p1.obs["condition"] == "unstimulated"
            ),
            gene_ids
        ].X.toarray(),
        columns=gene_ids,
        index=adata_proc_p1[np.logical_and(
            adata_proc_p1.obs["leiden_Cd4"].values != "nan",
            adata_proc_p1.obs["condition"] == "unstimulated"
        )].obs.index
    ),
], axis=1).groupby("leiden_Cd4").mean().reset_index().sort_values("leiden_Cd4")
tab.index = tab["leiden_Cd4"]
leiden_clusters_p1_cd4 = np.unique(tab["leiden_Cd4"].values)
g = sb.clustermap(tab.iloc[:, 1:], figsize=(20,20))
plt.close()
idx_leiden_p1_cd4 = g.dendrogram_row.reordered_ind
leiden_clusters_p1_cd4_ordered = np.array(leiden_clusters_p1_cd4)[idx_leiden_p1_cd4]

adata_proc_p1.obs["leiden_Cd4"] = pd.Categorical(
    adata_proc_p1.obs["leiden_Cd4"].values,
    categories=np.concatenate([leiden_clusters_p1_cd4_ordered, ["nan"]])
)
adata_proc_p1_unstim.obs["leiden_Cd4"] = pd.Categorical(
    adata_proc_p1_unstim.obs["leiden_Cd4"].values,
    categories=np.concatenate([leiden_clusters_p1_cd4_ordered, ["nan"]])
)

# %% [code]
fig, ax = plt.subplots(figsize=(5,5))
sc.pl.umap(
    adata_proc_p1, 
    ax=ax, show=False
)
adatatemp = adata_proc_p1[np.logical_and(
    adata_proc_p1.obs["condition"].values == "unstimulated",
    adata_proc_p1.obs["leiden_Cd4"].values != "nan"
)].copy()
adatatemp.obs["leiden_Cd4"] = pd.Categorical(
    adatatemp.obs["leiden_Cd4"].values,
    categories=np.sort(np.unique(adatatemp.obs["leiden_Cd4"].values))
)
sc.pl.umap(
    adatatemp, 
    color=["leiden_Cd4"], 
    ax=ax, show=False
)
plt.tight_layout()
plt.savefig(dir_panels+"fig3c6.pdf")
plt.savefig(dir_panels+"sfig7c2.pdf")
plt.show()

# %% [code]
gene_ids = adata_proc_p2.var_names[np.asarray(adata_proc_p2[
    np.logical_and(
        adata_proc_p2.obs["leiden_Cd4"].values != "nan",
        adata_proc_p2.obs["condition"] == "unstimulated"
    )
].X.mean(axis=0)).flatten() > np.log(1.0+0.5)]  # >logp1(mean = 0.5)
tab = pd.concat([
    adata_proc_p2[
        np.logical_and(
            adata_proc_p2.obs["leiden_Cd4"].values != "nan",
            adata_proc_p2.obs["condition"] == "unstimulated"
        )
    ].obs["leiden_Cd4"],
    pd.DataFrame(
        adata_proc_p2[
            np.logical_and(
                adata_proc_p2.obs["leiden_Cd4"].values != "nan",
                adata_proc_p2.obs["condition"] == "unstimulated"
            ),
            gene_ids
        ].X.toarray(),
        columns=gene_ids,
        index=adata_proc_p2[np.logical_and(
            adata_proc_p2.obs["leiden_Cd4"].values != "nan",
            adata_proc_p2.obs["condition"] == "unstimulated"
        )].obs.index
    ),
], axis=1).groupby("leiden_Cd4").mean().reset_index().sort_values("leiden_Cd4")
tab.index = tab["leiden_Cd4"]
leiden_clusters_p2_cd4 = np.unique(tab["leiden_Cd4"].values)
g = sb.clustermap(tab.iloc[:, 1:], figsize=(20,20))
plt.close()
idx_leiden_p2_cd4 = g.dendrogram_row.reordered_ind
leiden_clusters_p2_cd4_ordered = np.array(leiden_clusters_p2_cd4)[idx_leiden_p2_cd4]

adata_proc_p2.obs["leiden_Cd4"] = pd.Categorical(
    adata_proc_p2.obs["leiden_Cd4"].values,
    categories=np.concatenate([
        leiden_clusters_p2_cd4_ordered,
        ["nan"]
    ])
)
adata_proc_p2_unstim.obs["leiden_Cd4"] = pd.Categorical(
    adata_proc_p2_unstim.obs["leiden_Cd4"].values,
    categories=np.concatenate([
        leiden_clusters_p2_cd4_ordered,
        ["nan"]
    ])
)

# %% [code]
fig, ax = plt.subplots(figsize=(5,5))
sc.pl.umap(
    adata_proc_p2, 
    ax=ax, show=False
)
sc.pl.umap(
    adata_proc_p2[np.logical_and(
        adata_proc_p2.obs["condition"].values == "unstimulated",
        adata_proc_p2.obs["leiden_Cd4"].values != "nan"
    )], 
    color=["leiden_Cd4"], 
    ax=ax, show=False
)
plt.show()

# %% [code]
for k, v in markers_cd4.items():
    sc.pl.dotplot(
        adata_proc_p1[adata_proc_p1.obs["cell_type"].values == "Cd4"], 
        var_names=[x for x in v if x in adata_proc_p1.var_names], 
        groupby="leiden_Cd4", 
        use_raw=False,
        save="sfig7f_pbmc_cd4_tcells_leiden_" + k + ".pdf",
        show=False
    )

# %% [code]
fig, ax = plt.subplots(figsize=(5,5))
sc.pl.umap(
    adata_proc_p1, 
    ax=ax, show=False
)
sc.pl.umap(
    adata_proc_p1[np.logical_and(
        adata_proc_p1.obs["condition"].values == "unstimulated",
        adata_proc_p1.obs["cell_type"].values == "Cd4"
    )], 
    color=["IFNGR2"], use_raw=False,
    vmin=np.min(adata_proc_p1[adata_proc_p1.obs["cell_type"].values == "Cd4", "IFNGR2"].X),
    vmax=np.max(adata_proc_p1[adata_proc_p1.obs["cell_type"].values == "Cd4", "IFNGR2"].X),
    ax=ax, show=False
)
plt.tight_layout()
plt.savefig(dir_panels+"sfig1a1.pdf")
plt.show()

fig, ax = plt.subplots(figsize=(5,5))
sc.pl.umap(
    adata_proc_p1, 
    ax=ax, show=False
)
sc.pl.umap(
    adata_proc_p1[np.logical_and(
        adata_proc_p1.obs["condition"].values == "stimulated",
        adata_proc_p1.obs["cell_type"].values == "Cd4"
    )], 
    color=["IFNGR2"], use_raw=False,
    vmin=np.min(adata_proc_p1[adata_proc_p1.obs["cell_type"].values == "Cd4", "IFNGR2"].X),
    vmax=np.max(adata_proc_p1[adata_proc_p1.obs["cell_type"].values == "Cd4", "IFNGR2"].X),
    ax=ax, show=False
)
plt.tight_layout()
plt.savefig(dir_panels+"sfig1a2.pdf")
plt.show()

# %% [code]
fig, ax = plt.subplots(figsize=(5,5))
sc.pl.umap(
    adata_proc_p1, 
    ax=ax, show=False
)
sc.pl.umap(
    adata_proc_p1[np.logical_and(
        adata_proc_p1.obs["condition"].values == "unstimulated",
        adata_proc_p1.obs["cell_type"].values == "Cd4"
    )], 
    color=["CCR7"], use_raw=False,
    vmin=np.min(adata_proc_p1[adata_proc_p1.obs["cell_type"].values == "Cd4", "CCR7"].X),
    vmax=np.max(adata_proc_p1[adata_proc_p1.obs["cell_type"].values == "Cd4", "CCR7"].X),
    ax=ax, show=False
)
plt.tight_layout()
plt.savefig(dir_panels+"sfig7e1.pdf")
plt.show()

fig, ax = plt.subplots(figsize=(5,5))
sc.pl.umap(
    adata_proc_p1, 
    ax=ax, show=False
)
sc.pl.umap(
    adata_proc_p1[np.logical_and(
        adata_proc_p1.obs["condition"].values == "unstimulated",
        adata_proc_p1.obs["cell_type"].values == "Cd4"
    )], 
    color=["IL7R"], use_raw=False,
    vmin=np.min(adata_proc_p1[adata_proc_p1.obs["cell_type"].values == "Cd4", "IL7R"].X),
    vmax=np.max(adata_proc_p1[adata_proc_p1.obs["cell_type"].values == "Cd4", "IL7R"].X),
    ax=ax, show=False
)
plt.tight_layout()
plt.savefig(dir_panels+"sfig7e2.pdf")
plt.show()

# %% [markdown]
#### Differential expression

# %% [code]
adata_temp = adata_proc_p1[
    np.logical_and(
        adata_proc_p1.obs["leiden_Cd4"].values != "nan",
        adata_proc_p1.obs["condition"] == "unstimulated"
    )
]
sc.tl.rank_genes_groups(adata_temp, groupby="leiden_Cd4")
sc.pl.rank_genes_groups(adata_temp)
del adata_temp

# %% [code]
adata_temp = adata_proc_p2[
    np.logical_and(
        adata_proc_p2.obs["leiden_Cd4"].values != "nan",
        adata_proc_p2.obs["condition"] == "unstimulated"
    )
]
sc.tl.rank_genes_groups(adata_temp, groupby="leiden_Cd4")
sc.pl.rank_genes_groups(adata_temp)
del adata_temp

# %% [markdown]
### Cd8 cell states

# %% [code]
fig, ax = plt.subplots(figsize=(5,5))
sc.pl.umap(
    adata_proc_p1, 
    ax=ax, show=False
)
adatatemp = adata_proc_p1[np.logical_and(
    adata_proc_p1.obs["condition"].values == "unstimulated",
    adata_proc_p1.obs["leiden_Cd8"].values != "nan"
)].copy()
adatatemp.obs["leiden_Cd4"] = pd.Categorical(
    adatatemp.obs["leiden_Cd4"].values,
    categories=np.sort(np.unique(adatatemp.obs["leiden_Cd4"].values))
)
sc.pl.umap(
    adatatemp, 
    color=["leiden_Cd8"], 
    ax=ax, show=False
)
plt.tight_layout()
plt.savefig(dir_panels+"fig3c3.pdf")
plt.savefig(dir_panels+"sfig7c1.pdf")
plt.show()

# %% [code]
gene_ids = adata_proc_p1.var_names[np.asarray(adata_proc_p1[
    np.logical_and(
        adata_proc_p1.obs["leiden_Cd8"].values != "nan",
        adata_proc_p1.obs["condition"] == "unstimulated"
    )
].X.mean(axis=0)).flatten() > np.log(1.0+0.5)]  # >logp1(mean = 0.5)
tab = pd.concat([
    adata_proc_p1[
        np.logical_and(
            adata_proc_p1.obs["leiden_Cd8"].values != "nan",
            adata_proc_p1.obs["condition"] == "unstimulated"
        )
    ].obs["leiden_Cd8"],
    pd.DataFrame(
        adata_proc_p1[
            np.logical_and(
                adata_proc_p1.obs["leiden_Cd8"].values != "nan",
                adata_proc_p1.obs["condition"] == "unstimulated"
            ),
            gene_ids
        ].X.toarray(),
        columns=gene_ids,
        index=adata_proc_p1[np.logical_and(
            adata_proc_p1.obs["leiden_Cd8"].values != "nan",
            adata_proc_p1.obs["condition"] == "unstimulated"
        )].obs.index
    ),
], axis=1).groupby("leiden_Cd8").mean().reset_index().sort_values("leiden_Cd8")
tab.index = tab["leiden_Cd8"]
leiden_clusters_p1_cd8 = np.unique(tab["leiden_Cd8"].values)
g = sb.clustermap(tab.iloc[:, 1:], figsize=(20,20))
plt.close()
idx_leiden_p1_cd8 = g.dendrogram_row.reordered_ind
leiden_clusters_p1_cd8_ordered = np.array(leiden_clusters_p1_cd8)[idx_leiden_p1_cd8]

adata_proc_p1.obs["leiden_Cd8"] = pd.Categorical(
    adata_proc_p1.obs["leiden_Cd8"].values,
    categories=np.concatenate([leiden_clusters_p1_cd8_ordered, ["nan"]])
)
adata_proc_p1_unstim.obs["leiden_Cd8"] = pd.Categorical(
    adata_proc_p1_unstim.obs["leiden_Cd8"].values,
    categories=np.concatenate([leiden_clusters_p1_cd8_ordered, ["nan"]])
)

# %% [code]
fig, ax = plt.subplots(figsize=(5,5))
sc.pl.umap(
    adata_proc_p2, 
    ax=ax, show=False
)
sc.pl.umap(
    adata_proc_p2[np.logical_and(
        adata_proc_p2.obs["condition"].values == "unstimulated",
        adata_proc_p2.obs["leiden_Cd8"].values != "nan"
    )], 
    color=["leiden_Cd8"], 
    ax=ax, show=False
)
plt.tight_layout()
plt.show()

# %% [code]
gene_ids = adata_proc_p2.var_names[np.asarray(adata_proc_p2[
    np.logical_and(
        adata_proc_p2.obs["leiden_Cd8"].values != "nan",
        adata_proc_p2.obs["condition"] == "unstimulated"
    )
].X.mean(axis=0)).flatten() > np.log(1.0+0.5)]  # >logp1(mean = 0.5)
tab = pd.concat([
    adata_proc_p2[
        np.logical_and(
            adata_proc_p2.obs["leiden_Cd8"].values != "nan",
            adata_proc_p2.obs["condition"] == "unstimulated"
        )
    ].obs["leiden_Cd8"],
    pd.DataFrame(
        adata_proc_p2[
            np.logical_and(
                adata_proc_p2.obs["leiden_Cd8"].values != "nan",
                adata_proc_p2.obs["condition"] == "unstimulated"
            ),
            gene_ids
        ].X.toarray(),
        columns=gene_ids,
        index=adata_proc_p2[np.logical_and(
            adata_proc_p2.obs["leiden_Cd8"].values != "nan",
            adata_proc_p2.obs["condition"] == "unstimulated"
        )].obs.index
    ),
], axis=1).groupby("leiden_Cd8").mean().reset_index().sort_values("leiden_Cd8")
tab.index = tab["leiden_Cd8"]
leiden_clusters_p2_cd8 = np.unique(tab["leiden_Cd8"].values)
g = sb.clustermap(tab.iloc[:, 1:], figsize=(20,20))
plt.close()
idx_leiden_p2_cd8 = g.dendrogram_row.reordered_ind
leiden_clusters_p2_cd8_ordered = np.array(leiden_clusters_p2_cd8)[idx_leiden_p2_cd8]

adata_proc_p2.obs["leiden_Cd8"] = pd.Categorical(
    adata_proc_p2.obs["leiden_Cd8"].values,
    categories=np.concatenate([leiden_clusters_p2_cd8_ordered, ["nan"]])
)
adata_proc_p2_unstim.obs["leiden_Cd8"] = pd.Categorical(
    adata_proc_p2_unstim.obs["leiden_Cd8"].values,
    categories=np.concatenate([leiden_clusters_p2_cd8_ordered, ["nan"]])
)

# %% [code]
fig, ax = plt.subplots(figsize=(5,5))
sc.pl.umap(
    adata_proc_p1, 
    ax=ax, show=False
)
sc.pl.umap(
    adata_proc_p1[np.logical_and(
        adata_proc_p1.obs["condition"].values == "unstimulated",
        adata_proc_p1.obs["leiden_Cd8"].values != "nan"
    )], 
    color=["leiden_Cd8"], 
    ax=ax, show=False
)
plt.show()

# %% [code]
for k, v in markers_cd8.items():
    sc.pl.dotplot(
        adata_proc_p1[adata_proc_p1.obs["cell_type"].values == "Cd8"], 
        var_names=[x for x in v if x in adata_proc_p1.var_names], 
        groupby="leiden_Cd8", 
        use_raw=False,
        save="sfig7g_pbmc_cd8_tcells_leiden_" + k + ".pdf",
        show=False
    )

# %% [markdown]
#### Differential expression

# %% [code]
adata_temp = adata_proc_p1[
    np.logical_and(
        adata_proc_p1.obs["leiden_Cd8"].values != "nan",
        adata_proc_p1.obs["condition"] == "unstimulated"
    )
]
sc.tl.rank_genes_groups(adata_temp, groupby="leiden_Cd8")
sc.pl.rank_genes_groups(adata_temp)
del adata_temp

# %% [code]
adata_temp = adata_proc_p2[
    np.logical_and(
        adata_proc_p2.obs["leiden_Cd8"].values != "nan",
        adata_proc_p2.obs["condition"] == "unstimulated"
    )
]
sc.tl.rank_genes_groups(adata_temp, groupby="leiden_Cd8")
sc.pl.rank_genes_groups(adata_temp)
del adata_temp

# %% [markdown]
### Assign global sub states

# %% [code]
adata_proc_p1.obs["leiden_Cdall"] = [
    "cd8_" + str(adata_proc_p1_unstim[x].obs["leiden_Cd8"].values[0])
    if x in adata_proc_p1_unstim.obs_names and adata_proc_p1_unstim[x].obs["leiden_Cd8"].values != "nan"
    else "cd4_" + str(adata_proc_p1_unstim[x].obs["leiden_Cd4"].values[0])
    if x in adata_proc_p1_unstim.obs_names and adata_proc_p1_unstim[x].obs["leiden_Cd4"].values != "nan"
    else "other"
    for x in adata_proc_p1.obs_names
]

# %% [code]
gene_ids = adata_proc_p1.var_names[np.asarray(adata_proc_p1[
    np.logical_and(
        adata_proc_p1.obs["leiden_Cdall"].values != "other",
        adata_proc_p1.obs["condition"] == "unstimulated"
    )
].X.mean(axis=0)).flatten() > np.log(1.0+0.5)]  # >logp1(mean = 0.5)
tab = pd.concat([
    adata_proc_p1[
        np.logical_and(
            adata_proc_p1.obs["leiden_Cdall"].values != "other",
            adata_proc_p1.obs["condition"] == "unstimulated"
        )
    ].obs["leiden_Cdall"],
    pd.DataFrame(
        adata_proc_p1[
            np.logical_and(
                adata_proc_p1.obs["leiden_Cdall"].values != "other",
                adata_proc_p1.obs["condition"] == "unstimulated"
            ),
            gene_ids
        ].X.toarray(),
        columns=gene_ids,
        index=adata_proc_p1[np.logical_and(
            adata_proc_p1.obs["leiden_Cdall"].values != "other",
            adata_proc_p1.obs["condition"] == "unstimulated"
        )].obs.index
    ),
], axis=1).groupby("leiden_Cdall").mean().reset_index().sort_values("leiden_Cdall")
tab.index = tab["leiden_Cdall"]
leiden_clusters_p1_cdall = np.unique(tab["leiden_Cdall"].values)
g = sb.clustermap(tab.iloc[:, 1:], figsize=(20,20))
plt.close()
idx_leiden_p1_cdall = g.dendrogram_row.reordered_ind
leiden_clusters_p1_cdall_ordered = np.array(leiden_clusters_p1_cdall)[idx_leiden_p1_cdall]

adata_proc_p1.obs["leiden_Cdall"] = pd.Categorical(
    adata_proc_p1.obs["leiden_Cdall"].values,
    categories=np.concatenate([leiden_clusters_p1_cdall_ordered, ["nan"]])
)

# %% [markdown]
#### UMAPs

# %% [code]
fig, ax = plt.subplots(figsize=(5,5))
sc.pl.umap(
    adata_proc_p1, 
    ax=ax, show=False
)
sc.pl.umap(
    adata_proc_p1[adata_proc_p1.obs["condition"].values == "unstimulated"], 
    color=["cell_type"], 
    ax=ax, show=False
)
plt.tight_layout()
plt.savefig(dir_panels+"fig3c1.pdf")
plt.show()

# %% [code]
fig, ax = plt.subplots(figsize=(5,5))
sc.pl.umap(
    adata_proc_p1, 
    ax=ax, show=False
)
sc.pl.umap(
    adata_proc_p1[adata_proc_p1.obs["condition"].values == "unstimulated"], 
    color=["leiden_Cdall"], 
    ax=ax, show=False
)
plt.show()

# %% [markdown]
# Stimulation vectors

# %% [code]
n_high_genes = 2000
idx_highly_expressed_p1 = np.argsort(np.asarray(adata_proc_p1.X.mean(axis=0)).flatten())[::-1][:n_high_genes]
idx_highly_expressed_p2 = np.argsort(np.asarray(adata_proc_p2.X.mean(axis=0)).flatten())[::-1][:n_high_genes]

# %% [markdown]
## By cell type

# %% [code]
stim_vec_p1 = {}

idx_stim = np.where(np.logical_and(
    adata_proc_p1.obs["condition"].values == "stimulated",
    adata_proc_p1.obs["cell_type"].values == "Cd4"
))[0]
idx_unstim = np.where(np.logical_and(
    adata_proc_p1.obs["condition"].values == "unstimulated",
    adata_proc_p1.obs["cell_type"].values == "Cd4"
))[0]
stim_vec_p1["Cd4"] = np.asarray(adata_proc_p1[idx_stim, :].X[:, idx_highly_expressed_p1].mean(axis=0)).flatten() - \
    (np.asarray(adata_proc_p1[idx_unstim, :].X[:, idx_highly_expressed_p1].mean(axis=0)).flatten()+1e-8)

idx_stim = np.where(np.logical_and(
    adata_proc_p1.obs["condition"].values == "stimulated",
    adata_proc_p1.obs["cell_type"].values == "Cd8"
))[0]
idx_unstim = np.where(np.logical_and(
    adata_proc_p1.obs["condition"].values == "unstimulated",
    adata_proc_p1.obs["cell_type"].values == "Cd8"
))[0]
stim_vec_p1["Cd8"] = np.asarray(adata_proc_p1[idx_stim, :].X[:, idx_highly_expressed_p1].mean(axis=0)).flatten() - \
    (np.asarray(adata_proc_p1[idx_unstim, :].X[:, idx_highly_expressed_p1].mean(axis=0)).flatten()+1e-8)

# %% [code]
stim_vec_p2 = {}

idx_stim = np.where(np.logical_and(
    adata_proc_p2.obs["condition"].values == "stimulated",
    adata_proc_p2.obs["cell_type"].values == "Cd4"
))[0]
idx_unstim = np.where(np.logical_and(
    adata_proc_p2.obs["condition"].values == "unstimulated",
    adata_proc_p2.obs["cell_type"].values == "Cd4"
))[0]
stim_vec_p2["Cd4"] = np.asarray(adata_proc_p2[idx_stim, :].X[:, idx_highly_expressed_p2].mean(axis=0)).flatten() - \
    (np.asarray(adata_proc_p2[idx_unstim, :].X[:, idx_highly_expressed_p2].mean(axis=0)).flatten()+1e-8)

idx_stim = np.where(np.logical_and(
    adata_proc_p2.obs["condition"].values == "stimulated",
    adata_proc_p2.obs["cell_type"].values == "Cd8"
))[0]
idx_unstim = np.where(np.logical_and(
    adata_proc_p2.obs["condition"].values == "unstimulated",
    adata_proc_p2.obs["cell_type"].values == "Cd8"
))[0]
stim_vec_p2["Cd8"] = np.asarray(adata_proc_p2[idx_stim, :].X[:, idx_highly_expressed_p2].mean(axis=0)).flatten() - \
    (np.asarray(adata_proc_p2[idx_unstim, :].X[:, idx_highly_expressed_p2].mean(axis=0)).flatten()+1e-8)

# %% [markdown]
## By clonotype

# %% [code]
vecs_p1 = {}
for x, y, z in zip(
    tab_clonotypes_p1_filtered["clonotype_merged"].values,
    tab_clonotypes_p1_filtered["label_cell_type"].values,
    tab_clonotypes_p1_filtered["clonotype"].values,
):  # computes vectors of clonotypes that were merged multiple times so that vecs still has dimensionality of tab_clonotypes_p1_filtered
    idx_stim = np.where(np.logical_and(
        np.logical_and(
            adata_proc_p1.obs["condition"].values == "stimulated",
            adata_proc_p1.obs["clonotype_merged"].values == x,
        ),
        adata_proc_p1.obs["cell_type"].values == y
    ))[0]
    idx_unstim = np.where(np.logical_and(
        np.logical_and(
            adata_proc_p1.obs["condition"].values == "unstimulated",
            adata_proc_p1.obs["clonotype_merged"].values == x,
        ),
        adata_proc_p1.obs["cell_type"].values == y
    ))[0]
    vecs_p1[z] = np.asarray(adata_proc_p1[idx_stim, :].X[:, idx_highly_expressed_p1].mean(axis=0)).flatten() - \
        (np.asarray(adata_proc_p1[idx_unstim, :].X[:, idx_highly_expressed_p1].mean(axis=0)).flatten()+1e-8)

# %% [code]
mean_vec_p1 = np.mean(
    np.concatenate([np.expand_dims(x, axis=0) for x in list(vecs_p1.values())], axis=0), 
    axis=0
)
tab_clonotypes_p1_filtered["r2"] = [
    np.corrcoef(
        x=v,
        y=mean_vec_p1
    )[0,1] for k, v in vecs_p1.items()
]

# %% [code]
vecs_p2 = {}
for x, y in zip(
    tab_clonotypes_p2_filtered["clonotype"].values,
    tab_clonotypes_p2_filtered["label_cell_type"].values
):
    idx_stim = np.where(np.logical_and(
        np.logical_and(
            adata_proc_p2.obs["condition"].values == "stimulated",
            adata_proc_p2.obs["clonotype"].values == x,
        ),
        adata_proc_p2.obs["cell_type"].values == y
    ))[0]
    idx_unstim = np.where(np.logical_and(
        np.logical_and(
            adata_proc_p2.obs["condition"].values == "unstimulated",
            adata_proc_p2.obs["clonotype"].values == x,
        ),
        adata_proc_p2.obs["cell_type"].values == y
    ))[0]
    vecs_p2[x] = np.asarray(adata_proc_p2[idx_stim, :].X[:, idx_highly_expressed_p2].mean(axis=0)).flatten() - \
        (np.asarray(adata_proc_p2[idx_unstim, :].X[:, idx_highly_expressed_p2].mean(axis=0)).flatten()+1e-8)

# %% [code]
mean_vec_p2 = np.mean(
    np.concatenate([np.expand_dims(x, axis=0) for x in list(vecs_p2.values())], axis=0), 
    axis=0
)
tab_clonotypes_p2_filtered["r2"] = [
    np.corrcoef(
        x=v,
        y=mean_vec_p2
    )[0,1] for k, v in vecs_p2.items()
]

# %% [markdown]
# Summary

# %% [markdown]
## Clonotype summaries

# %% [markdown]
### Full clonotype table

# %% [markdown]
#### Primary chain clonotypes

# %% [code]
tab_clonotypes_p1_filtered.sort_values("size").iloc[::-1].to_csv(dir_tables+'sfig3a.csv', sep=";")
tab_clonotypes_p1_filtered.loc[tab_clonotypes_p1_filtered["label_cell_type"].values == "Cd4"].sort_values("size").iloc[::-1].to_csv(dir_tables+'sfig3b1_cd4.csv', sep=";")
tab_clonotypes_p1_filtered.loc[tab_clonotypes_p1_filtered["label_cell_type"].values == "Cd8"].sort_values("size").iloc[::-1].to_csv(dir_tables+'sfig3b2_cd8.csv', sep=";")

# %% [code]
tab_clonotypes_p1_filtered.loc[tab_clonotypes_p1_filtered["label_cell_type"].values == "Cd4"].sort_values("size").iloc[::-1].to_csv(dir_tables+'tab_clonotypes_p1_cd4_filtered.csv', sep=";")
tab_clonotypes_p1_filtered.loc[tab_clonotypes_p1_filtered["label_cell_type"].values == "Cd8"].sort_values("size").iloc[::-1].to_csv(dir_tables+'tab_clonotypes_p1_cd8_filtered.csv', sep=";")
tab_clonotypes_p2_filtered.loc[tab_clonotypes_p2_filtered["label_cell_type"].values == "Cd4"].sort_values("size").iloc[::-1].to_csv(dir_tables+'tab_clonotypes_p2_cd4_filtered.csv', sep=";")
tab_clonotypes_p2_filtered.loc[tab_clonotypes_p2_filtered["label_cell_type"].values == "Cd8"].sort_values("size").iloc[::-1].to_csv(dir_tables+'tab_clonotypes_p2_cd8_filtered.csv', sep=";")

# %% [code]
tab_clonotypes_p1_filtered.sort_values("size").iloc[::-1].to_csv(dir_tables+'ExtendedDataTable11_P1_filtered_clonotypes.csv', sep=";")
tab_clonotypes_p2_filtered.sort_values("size").iloc[::-1].to_csv(dir_tables+'ExtendedDataTable12_P2_filtered_clonotypes.csv', sep=";")

# %% [code]
clonotypes_p1_cd4_selected = tab_clonotypes_p1_filtered.loc[np.logical_and(
    tab_clonotypes_p1_filtered["label_cell_type"].values == "Cd4",
    np.logical_and(
        np.logical_and(
            tab_clonotypes_p1_filtered["n_stimulated"].values >= 1,
            tab_clonotypes_p1_filtered["n_unstimulated"].values >= 1
        ),
        tab_clonotypes_p1_filtered["in_ifng_cluster"].values > 0.
    )
), "clonotype_merged"].values
clonotypes_p1_cd4_selected = np.unique(clonotypes_p1_cd4_selected)
np.sort(clonotypes_p1_cd4_selected.astype(int))

# %% [code]
clonotypes_p1_cd8_selected = tab_clonotypes_p1_filtered.loc[np.logical_and(
    tab_clonotypes_p1_filtered["label_cell_type"].values == "Cd8",
    np.logical_and(
        np.logical_and(
            tab_clonotypes_p1_filtered["n_stimulated"].values >= 1,
            tab_clonotypes_p1_filtered["n_unstimulated"].values >= 1
        ),
        tab_clonotypes_p1_filtered["in_ifng_cluster"].values > 0.
    )
), "clonotype_merged"].values
clonotypes_p1_cd8_selected = np.unique(clonotypes_p1_cd8_selected)
np.sort(clonotypes_p1_cd8_selected.astype(int))

# %% [code]
clonotypes_p2_cd4_selected = tab_clonotypes_p2_filtered.loc[np.logical_and(
    tab_clonotypes_p2_filtered["label_cell_type"].values == "Cd4",
    np.logical_and(
        np.logical_and(
            tab_clonotypes_p2_filtered["n_stimulated"].values >= 1,
            tab_clonotypes_p2_filtered["n_unstimulated"].values >= 1
        ),
        tab_clonotypes_p2_filtered["in_ifng_cluster"].values > 0.
    )
), "clonotype_merged"].values
clonotypes_p2_cd4_selected = np.unique(clonotypes_p2_cd4_selected)
np.sort(clonotypes_p2_cd4_selected.astype(int))

# %% [code]
clonotypes_p2_cd8_selected = tab_clonotypes_p2_filtered.loc[np.logical_and(
    tab_clonotypes_p2_filtered["label_cell_type"].values == "Cd8",
    np.logical_and(
        np.logical_and(
            tab_clonotypes_p2_filtered["n_stimulated"].values >= 1,
            tab_clonotypes_p2_filtered["n_unstimulated"].values >= 1
        ),
        tab_clonotypes_p2_filtered["in_ifng_cluster"].values > 0.
    )
), "clonotype_merged"].values
clonotypes_p2_cd8_selected = np.unique(clonotypes_p2_cd8_selected)
np.sort(clonotypes_p2_cd8_selected.astype(int))

# %% [code]
fig, ax = plt.subplots(nrows=3, ncols=1, figsize=(4, 12))
sb.regplot(
    data=tab_clonotypes_p1_filtered,
    x="n_stimulated",
    y="n_unstimulated",
    color="black",
    ax=ax[0]
)
r2 = np.corrcoef(
    tab_clonotypes_p1_filtered["n_stimulated"].values,
    tab_clonotypes_p1_filtered["n_unstimulated"].values,
)[0, 1]
ax[0].text(20, 1, "R2="+str(np.round(r2, 2)), style='italic', fontsize=12,
        bbox={'facecolor': 'white', 'alpha': 1., 'pad': 10})
ax[0].set_xlabel("number of stimulated cell")
ax[0].set_ylabel("number of unstimulated cell")
ax[0].set_xscale('log')
ax[0].set_yscale('log')

sb.regplot(
    data=tab_clonotypes_p1_filtered.loc[tab_clonotypes_p1_filtered["label_cell_type"].values == "Cd4"],
    x="ave_ifng_stimulated",
    y="proliferation_score_stim",
    color="black",
    ax=ax[1]
)
r2 = np.corrcoef(
    tab_clonotypes_p1_filtered["ave_ifng_stimulated"].values,
    tab_clonotypes_p1_filtered["proliferation_score_stim"].values,
)[0, 1]
ax[1].text(2, 0.1, "R2="+str(np.round(r2, 2)), style='italic', fontsize=12,
        bbox={'facecolor': 'white', 'alpha': 1., 'pad': 10})
ax[1].set_xlabel("IFNG (stim)")
ax[1].set_ylabel("Proliferation score (stim)")

sb.regplot(
    data=tab_clonotypes_p1_filtered.loc[tab_clonotypes_p1_filtered["label_cell_type"].values == "Cd8"],
    x="ave_ifng_stimulated",
    y="cd8 cytokine_score_stim",
    color="black",
    ax=ax[2]
)
r2 = np.corrcoef(
    tab_clonotypes_p1_filtered["ave_ifng_stimulated"].values,
    tab_clonotypes_p1_filtered["cd8 cytokine_score_stim"].values,
)[0, 1]
ax[2].text(0.8, 0.1, "R2="+str(np.round(r2, 2)), style='italic', fontsize=12,
        bbox={'facecolor': 'white', 'alpha': 1., 'pad': 10})
ax[2].set_xlabel("IFNG (stim)")
ax[2].set_ylabel("Cytokine score (stim)")

plt.tight_layout()
plt.savefig(dir_panels + "sfig3.pdf")
plt.show()

# %% [markdown]
#### All chains clonotypes

# %% [code]
tab_clonotypes_all_chains_p1_filtered.sort_values("size").iloc[::-1].to_csv(dir_tables+'tab_clonotypes_all_chains_p1_filtered.csv', sep=";")
tab_clonotypes_all_chains_p2_filtered.sort_values("size").iloc[::-1].to_csv(dir_tables+'tab_clonotypes_all_chains_p2_filtered.csv', sep=";")

# %% [markdown]
## Export cell tables

# %% [markdown]
### Ifng+ cluster

# %% [code]
tab = adata_proc_p1.obs.loc[
    adata_proc_p1.obs["ifng_pos_cluster"].values == True, :
]
for x in ["IFNG"]:
    tab[x] = np.asarray(adata_proc_p1[tab.index, x].X.todense()).flatten()
tab.to_csv(dir_tables+'tab_cells_p1_ifng_pos_cluster.csv', sep=";")

# %% [markdown]
### P1

# %% [code]
gene_list = ["TBX21", "EOMES", "GATA3"]
clonotypes_p1 = [
    x for x in np.unique(adata_proc_p1.obs["clonotype"].values)
    if np.sum(np.logical_and(
        adata_proc_p1.obs["clonotype"].values == x,
        adata_proc_p1.obs["condition"].values == "stimulated"
    )) >= 1 and np.sum(np.logical_and(
        adata_proc_p1.obs["clonotype"].values == x,
        adata_proc_p1.obs["condition"].values == "unstimulated"
    )) >= 1
]
tab_clonotypes_p1_stim_filtered_bycell = {
    "Cd4": {x: [] for x in gene_list},
    "Cd8": {x: [] for x in gene_list}
}
tab_clonotypes_p1_unstim_filtered_bycell = {
    "Cd4": {x: [] for x in gene_list},
    "Cd8": {x: [] for x in gene_list}
}

adata_temp = adata_proc_p1.copy()
for k in keys:
    del adata_temp.uns[k]
ncell_cols_stim = np.max(tab_clonotypes_p1_filtered["n_stimulated"].values)
ncell_cols_unstim = np.max(tab_clonotypes_p1_filtered["n_unstimulated"].values)
for x in clonotypes_p1:
    adata = adata_temp[adata_temp.obs["clonotype"].values == x]
    ncells_stim = np.sum(adata.obs["condition"].values == "stimulated")
    ncells_unstim = np.sum(adata.obs["condition"].values == "unstimulated")
    cell_type = tab_clonotypes_p1_filtered.loc[
        tab_clonotypes_p1_filtered["clonotype"].values == x,
        "label_cell_type"
    ].values[0]
    for k in ["TBX21", "EOMES", "GATA3"]:
        # stimulated
        tab_clonotypes_p1_stim_filtered_bycell[cell_type][k].append(pd.DataFrame(dict(list({
            "clonotype": x,
            "clonotypes_all_chains": [np.unique(adata.obs["clonotype_all_chains"].values)],
            "size": adata.shape[0],
            "n_stimulated": np.sum(adata.obs["condition"].values == "stimulated"),
            "n_unstimulated": np.sum(adata.obs["condition"].values == "unstimulated"),
        }.items()) + [
            (
                "cell_" + str(i),
                np.asarray(adata[adata.obs["condition"].values == "stimulated", k].X.todense()).flatten()[i]
                if i < ncells_stim else np.nan
            )
            for i in range(ncell_cols_stim)
        ]), index=[x]))
        # unstimulated
        tab_clonotypes_p1_unstim_filtered_bycell[cell_type][k].append(pd.DataFrame(dict(list({
            "clonotype": x,
            "clonotypes_all_chains": [np.unique(adata.obs["clonotype_all_chains"].values)],
            "size": adata.shape[0],
            "n_stimulated": np.sum(adata.obs["condition"].values == "stimulated"),
            "n_unstimulated": np.sum(adata.obs["condition"].values == "unstimulated"),
        }.items()) + [
            (
                "cell_" + str(i),
                np.asarray(adata[adata.obs["condition"].values == "unstimulated", k].X.todense()).flatten()[i]
                if i < ncells_unstim else np.nan
            )
            for i in range(ncell_cols_unstim)
        ]), index=[x]))
for k in ["Cd4", "Cd8"]:
    for kk in tab_clonotypes_p1_unstim_filtered_bycell[k].keys():
        tab_clonotypes_p1_unstim_filtered_bycell[k][kk] = pd.concat(tab_clonotypes_p1_unstim_filtered_bycell[k][kk], axis=0)
        tab_clonotypes_p1_unstim_filtered_bycell[k][kk].sort_values("size").iloc[::-1].to_csv(dir_tables+'tab_clonotypes_p1_' + k.lower() + '_' + kk.lower() + '_unstim_filtered_bycell.csv', sep=";")

        tab_clonotypes_p1_stim_filtered_bycell[k][kk] = pd.concat(tab_clonotypes_p1_stim_filtered_bycell[k][kk], axis=0)
        tab_clonotypes_p1_stim_filtered_bycell[k][kk].sort_values("size").iloc[::-1].to_csv(dir_tables+'tab_clonotypes_p1_' + k.lower() + '_'  + kk.lower() + '_stim_filtered_bycell.csv', sep=";")

# %% [markdown]
# Select reactive clonotypes

# %% [markdown]
## Define reactive clonotypes

# %% [markdown]
### Show selected clonotypes on UMAP

# %% [markdown]
#### P1

# %% [code]
clono_type_list = ["19", "574", "244", "13"]
nrows = len(clono_type_list)
ncols = 2
vmax = adata_proc_p1[[x in clono_type_list for x in adata_proc_p1.obs["clonotype_merged"].values], "IFNG"].X.max()
fig, axs = plt.subplots(nrows=nrows, ncols=ncols, figsize=(ncols*3, 3*nrows))
for i, x in enumerate(clono_type_list):
    sc.pl.umap(adata_proc_p1, ax=axs[i, 0], show=False)
    sc.pl.umap(
        adata_proc_p1[np.logical_and(
            adata_proc_p1.obs["clonotype_merged"].values == x,
            adata_proc_p1.obs["condition"].values == "unstimulated"
        )], 
        color="IFNG",
        vmin=0,
        vmax=vmax,
        size=40,
        use_raw=False,
        ax=axs[i, 0], 
        show=False
    )
    axs[i, 0].set_ylabel(x)
    
    sc.pl.umap(adata_proc_p1, ax=axs[i, 1], show=False)
    sc.pl.umap(
        adata_proc_p1[np.logical_and(
            adata_proc_p1.obs["clonotype_merged"].values == x,
            adata_proc_p1.obs["condition"].values == "stimulated"
        )], 
        color="IFNG",
        vmin=0,
        vmax=vmax,
        size=40,
        use_raw=False,
        ax=axs[i, 1], 
        show=False
    )
    axs[i, 1].set_ylabel(" ")
    
    if i < len(clono_type_list) - 1:
        axs[i, 0].set_xlabel(" ")
        axs[i, 1].set_xlabel(" ")
    if i == 0:
        axs[i, 0].set_title("unstimulated")
        axs[i, 1].set_title("stimulated")
    else:
        axs[i, 0].set_title(" ")
        axs[i, 1].set_title(" ")
#plt.tight_layout()
plt.subplots_adjust(wspace=0.1, hspace=0.1)
plt.savefig(dir_panels + "fig1e.pdf")
plt.close()

# %% [code]
clono_type_list = ["1373", "1904"]
ncols = len(clono_type_list)
nrows = 2
vmax = adata_proc_p1[[x in clono_type_list for x in adata_proc_p1.obs["clonotype"].values], "IFNG"].X.max()
fig, axs = plt.subplots(nrows=nrows, ncols=ncols, figsize=(ncols*3, 3*nrows))
for i, x in enumerate(clono_type_list):
    sc.pl.umap(adata_proc_p1, ax=axs[i, 0], show=False)
    sc.pl.umap(
        adata_proc_p1[np.logical_and(
            adata_proc_p1.obs["clonotype"].values == x,
            adata_proc_p1.obs["condition"].values == "unstimulated"
        )], 
        color="IFNG",
        use_raw=False,
        vmin=0,
        vmax=vmax,
        size=40,
        ax=axs[i, 0], 
        show=False
    )
    axs[i, 0].set_ylabel(x)
    
    sc.pl.umap(adata_proc_p1, ax=axs[i, 1], show=False)
    sc.pl.umap(
        adata_proc_p1[np.logical_and(
            adata_proc_p1.obs["clonotype"].values == x,
            adata_proc_p1.obs["condition"].values == "stimulated"
        )], 
        color="IFNG",
        use_raw=False,
        vmin=0,
        vmax=vmax,
        size=40,
        ax=axs[i, 1], 
        show=False
    )
    axs[i, 1].set_ylabel(" ")
    
    axs[i, 0].set_xlabel(" ")
    axs[i, 1].set_xlabel(" ")
    if i == 0:
        axs[i, 0].set_title("unstimulated")
        axs[i, 1].set_title("stimulated")
    else:
        axs[i, 0].set_title(" ")
        axs[i, 1].set_title(" ")
#plt.tight_layout()
plt.subplots_adjust(wspace=0.15, hspace=0.1)
plt.savefig(dir_panels + "sfig5d.pdf")
plt.show()

# %% [code]
clono_type_list = ["81", "651", "244", "548"]
nrows = len(clono_type_list)
ncols = 2
vmax = adata_proc_p1[[x in clono_type_list for x in adata_proc_p1.obs["clonotype"].values], "IFNG"].X.max()
fig, axs = plt.subplots(nrows=nrows, ncols=ncols, figsize=(ncols*3, 3*nrows))
for i, x in enumerate(clono_type_list):
    sc.pl.umap(adata_proc_p1, ax=axs[i, 0], show=False)
    sc.pl.umap(
        adata_proc_p1[np.logical_and(
            adata_proc_p1.obs["clonotype"].values == x,
            adata_proc_p1.obs["condition"].values == "unstimulated"
        )], 
        color="IFNG",
        vmin=0,
        vmax=vmax,
        size=40,
        use_raw=False,
        ax=axs[i, 0], 
        show=False
    )
    axs[i, 0].set_ylabel(x)
    
    sc.pl.umap(adata_proc_p1, ax=axs[i, 1], show=False)
    sc.pl.umap(
        adata_proc_p1[np.logical_and(
            adata_proc_p1.obs["clonotype"].values == x,
            adata_proc_p1.obs["condition"].values == "stimulated"
        )], 
        color="IFNG",
        vmin=0,
        vmax=vmax,
        size=40,
        use_raw=False,
        ax=axs[i, 1], 
        show=False
    )
    axs[i, 1].set_ylabel(" ")
    
    if i < len(clono_type_list) - 1:
        axs[i, 0].set_xlabel(" ")
        axs[i, 1].set_xlabel(" ")
    if i == 0:
        axs[i, 0].set_title("unstimulated")
        axs[i, 1].set_title("stimulated")
    else:
        axs[i, 0].set_title(" ")
        axs[i, 1].set_title(" ")
#plt.tight_layout()
plt.subplots_adjust(wspace=0.1, hspace=0.1)
plt.savefig(dir_panels + "sfig4b.pdf")
plt.close()

# %% [markdown]
#### P2

# %% [code]
clono_type_list = tab_clonotypes_p2_filtered[
    tab_clonotypes_p2_filtered["label_cell_type"].values == "Cd4"
].sort_values("size").iloc[::-1, :]["clonotype_merged"].values[:20].tolist()
clono_type_list = [
    x for x in clonotypes_p2_cd4_selected
    if x not in clono_type_list
] + clono_type_list
# nrows = len(clono_type_list) // 4 + int(len(clono_type_list) % 4 > 0)
ncols = 4
nrows = len(clono_type_list)
fig, axs = plt.subplots(nrows=nrows, ncols=ncols, figsize=(ncols*3, 3*nrows))
axs = axs.ravel()
for i, x in enumerate(clono_type_list):
    sc.pl.umap(adata_proc_p2, ax=axs[i*ncols], show=False)
    sc.pl.umap(
        adata_proc_p2[np.logical_and(
            adata_proc_p2.obs["clonotype_merged"].values == x,
            adata_proc_p2.obs["condition"].values == "unstimulated"
        )], 
        color="IFNG",
        vmin=0,
        vmax=adata_proc_p2[adata_proc_p2.obs["clonotype_merged"].values == x, "IFNG"].X.max(),
        size=40,
        use_raw=False,
        ax=axs[i*ncols], 
        show=False
    )
    axs[i*ncols].set_ylabel("%s\nUMAP2" % x)
    
    sc.pl.umap(adata_proc_p2, ax=axs[i*ncols+1], show=False)
    sc.pl.umap(
        adata_proc_p2[np.logical_and(
            adata_proc_p2.obs["clonotype_merged"].values == x,
            adata_proc_p2.obs["condition"].values == "stimulated"
        )], 
        color="IFNG",
        vmin=0,
        vmax=adata_proc_p2[adata_proc_p2.obs["clonotype_merged"].values == x, "IFNG"].X.max(),
        size=40,
        use_raw=False,
        ax=axs[i*ncols+1], 
        show=False
    )
    
    sc.pl.umap(adata_proc_p2, ax=axs[i*ncols+2], show=False)
    sc.pl.umap(
        adata_proc_p2[np.logical_and(
            adata_proc_p2.obs["clonotype_merged"].values == x,
            adata_proc_p2.obs["condition"].values == "unstimulated"
        )], 
        color="IL2",
        vmin=0,
        vmax=adata_proc_p2[adata_proc_p2.obs["clonotype_merged"].values == x, "IL2"].X.max(),
        size=40,
        use_raw=False,
        ax=axs[i*ncols+2], 
        show=False
    )
    
    sc.pl.umap(adata_proc_p2, ax=axs[i*ncols+3], show=False)
    sc.pl.umap(
        adata_proc_p2[np.logical_and(
            adata_proc_p2.obs["clonotype_merged"].values == x,
            adata_proc_p2.obs["condition"].values == "stimulated"
        )], 
        color="IL2",
        vmin=0,
        vmax=adata_proc_p2[adata_proc_p2.obs["clonotype_merged"].values == x, "IL2"].X.max(),
        size=40,
        use_raw=False,
        ax=axs[i*ncols+3], 
        show=False
    )
    if i == 0:
        axs[i*ncols].set_title("unstimulated (IFNG)")
        axs[i*ncols+1].set_title("stimulated (IFNG)")
        axs[i*ncols+2].set_title("unstimulated (IL2)")
        axs[i*ncols+3].set_title("stimulated (IL2)")
    else:
        axs[i*ncols].set_title(" ")
        axs[i*ncols+1].set_title(" ")
        axs[i*ncols+2].set_title(" ")
        axs[i*ncols+3].set_title(" ")
plt.tight_layout()
plt.savefig(dir_panels + "umap_p2_cd4_marker_clonotypes.pdf")
plt.close()

# %% [code]
clono_type_list = tab_clonotypes_p2_filtered[
    tab_clonotypes_p2_filtered["label_cell_type"].values == "Cd8"
].sort_values("size").iloc[::-1, :]["clonotype_merged"].values[:20].tolist()
clono_type_list = [
    x for x in clonotypes_p2_cd8_selected
    if x not in clono_type_list
] + clono_type_list
# nrows = len(clono_type_list) // 4 + int(len(clono_type_list) % 4 > 0)
ncols = 4
nrows = len(clono_type_list)
fig, axs = plt.subplots(nrows=nrows, ncols=ncols, figsize=(ncols*3, 3*nrows))
axs = axs.ravel()
for i, x in enumerate(clono_type_list):
    sc.pl.umap(adata_proc_p2, ax=axs[i*ncols], show=False)
    sc.pl.umap(
        adata_proc_p2[np.logical_and(
            adata_proc_p2.obs["clonotype_merged"].values == x,
            adata_proc_p2.obs["condition"].values == "unstimulated"
        )], 
        color="IFNG",
        vmin=0,
        vmax=adata_proc_p2[adata_proc_p2.obs["clonotype_merged"].values == x, "IFNG"].X.max(),
        size=40,
        use_raw=False,
        ax=axs[i*ncols], 
        show=False
    )
    axs[i*ncols].set_ylabel("%s\nUMAP2" % x)
    
    sc.pl.umap(adata_proc_p2, ax=axs[i*ncols+1], show=False)
    sc.pl.umap(
        adata_proc_p2[np.logical_and(
            adata_proc_p2.obs["clonotype_merged"].values == x,
            adata_proc_p2.obs["condition"].values == "stimulated"
        )], 
        color="IFNG",
        vmin=0,
        vmax=adata_proc_p2[adata_proc_p2.obs["clonotype_merged"].values == x, "IFNG"].X.max(),
        size=40,
        use_raw=False,
        ax=axs[i*ncols+1], 
        show=False
    )
    
    sc.pl.umap(adata_proc_p2, ax=axs[i*ncols+2], show=False)
    sc.pl.umap(
        adata_proc_p2[np.logical_and(
            adata_proc_p2.obs["clonotype_merged"].values == x,
            adata_proc_p2.obs["condition"].values == "unstimulated"
        )], 
        color="IL2",
        vmin=0,
        vmax=adata_proc_p2[adata_proc_p2.obs["clonotype_merged"].values == x, "IL2"].X.max(),
        size=40,
        use_raw=False,
        ax=axs[i*ncols+2], 
        show=False
    )
    
    sc.pl.umap(adata_proc_p2, ax=axs[i*ncols+3], show=False)
    sc.pl.umap(
        adata_proc_p2[np.logical_and(
            adata_proc_p2.obs["clonotype_merged"].values == x,
            adata_proc_p2.obs["condition"].values == "stimulated"
        )], 
        color="IL2",
        vmin=0,
        vmax=adata_proc_p2[adata_proc_p2.obs["clonotype_merged"].values == x, "IL2"].X.max(),
        size=40,
        use_raw=False,
        ax=axs[i*ncols+3], 
        show=False
    )
    if i == 0:
        axs[i*ncols].set_title("unstimulated (IFNG)")
        axs[i*ncols+1].set_title("stimulated (IFNG)")
        axs[i*ncols+2].set_title("unstimulated (IL2)")
        axs[i*ncols+3].set_title("stimulated (IL2)")
    else:
        axs[i*ncols].set_title(" ")
        axs[i*ncols+1].set_title(" ")
        axs[i*ncols+2].set_title(" ")
        axs[i*ncols+3].set_title(" ")
plt.tight_layout()
plt.savefig(dir_panels + "umap_p2_cd8_marker_clonotypes.pdf")
plt.close()

# %% [markdown]
### Show scores in stimulated condition in cells from same clonotype in unstim

# %% [code]
score_to_plot = [k.lower() + "_score" for k in tcell_signatures.keys()]
for k in score_to_plot:
    adata_proc_p1.obs[k + "_in_mapped_stim"] = [
        tab_clonotypes_p1_filtered.loc[tab_clonotypes_p1_filtered["clonotype"].values == x, k + "_stim"].values[0]
        if x in tab_clonotypes_p1_filtered["clonotype"].values
        else np.nan
        for i, x in enumerate(adata_proc_p1.obs["clonotype"].values)
    ]
genes_to_plot = ["IFNG", "IL2", "TNF", "NME1", "IFIT1", "IL2RA", "FOXP3"]
for k in genes_to_plot:
    adata_proc_p1.obs[k + "_in_mapped_stim"] = [
        tab_clonotypes_p1_filtered.loc[tab_clonotypes_p1_filtered["clonotype"].values == x, "ave_" + k.lower() + "_stimulated"].values[0]
        if x in tab_clonotypes_p1_filtered["clonotype"].values
        else np.nan
        for i, x in enumerate(adata_proc_p1.obs["clonotype"].values)
    ]
x_to_plot = score_to_plot + genes_to_plot
fig, ax = plt.subplots(nrows=1, ncols=len(x_to_plot), figsize=(len(x_to_plot)*4,4))
for i, k in enumerate(x_to_plot):
    sc.pl.umap(
        adata_proc_p1, 
        ax=ax[i], show=False
    )
    sc.pl.umap(
        adata_proc_p1[adata_proc_p1.obs["condition"]=="unstimulated", :], 
        color=k + "_in_mapped_stim", 
        ax=ax[i], show=False
    )
plt.tight_layout()
plt.savefig(dir_panels+"umap_p1_unstim_score_map_from_stim_via_clonotype.pdf")
plt.close()

# %% [code]
score_to_plot = [k.lower() + "_score" for k in tcell_signatures.keys()]
for k in score_to_plot:
    adata_proc_p2.obs[k + "_in_mapped_stim"] = [
        tab_clonotypes_p2_filtered.loc[tab_clonotypes_p2_filtered["clonotype"].values == x, k + "_stim"].values[0]
        if x in tab_clonotypes_p2_filtered["clonotype"].values
        else np.nan
        for i, x in enumerate(adata_proc_p2.obs["clonotype"].values)
    ]
genes_to_plot = ["IFNG", "IL2", "TNF", "NME1", "IFIT1", "IL2RA", "FOXP3"]
for k in genes_to_plot:
    adata_proc_p2.obs[k + "_in_mapped_stim"] = [
        tab_clonotypes_p2_filtered.loc[tab_clonotypes_p2_filtered["clonotype"].values == x, "ave_" + k.lower() + "_stimulated"].values[0]
        if x in tab_clonotypes_p2_filtered["clonotype"].values
        else np.nan
        for i, x in enumerate(adata_proc_p2.obs["clonotype"].values)
    ]
x_to_plot = score_to_plot + genes_to_plot
fig, ax = plt.subplots(nrows=1, ncols=len(x_to_plot), figsize=(len(x_to_plot)*4,4))
for i, k in enumerate(x_to_plot):
    sc.pl.umap(
        adata_proc_p2, 
        ax=ax[i], show=False
    )
    sc.pl.umap(
        adata_proc_p2[adata_proc_p2.obs["condition"]=="unstimulated", :], 
        color=k + "_in_mapped_stim", 
        ax=ax[i], show=False
    )
plt.tight_layout()
plt.savefig(dir_panels+"umap_p2_unstim_score_map_from_stim_via_clonotype.pdf")
plt.close()

# %% [markdown]
### Manual definition

# %% [code]
#clonotypes_p1_cd4_selected_final = clonotypes_p1_cd4_selected
clonotypes_p1_cd4_selected_final = np.array(["19", "136", "138", "256", "574"])
clonotypes_p1_cd4_selected_final_merged = np.array(["19", "138", "256", "574"])
print([x for x in clonotypes_p1_cd4_selected_final if x not in clonotypes_p1_cd4_selected])

#clonotypes_p1_cd8_selected_final = clonotypes_p1_cd8_selected
clonotypes_p1_cd8_selected_final = np.array(["81", "104", "152", "244", "304", "548", "651"])
clonotypes_p1_cd8_selected_final_merged = np.array(["81", "104", "152", "244", "304"])
print([x for x in clonotypes_p1_cd8_selected_final if x not in clonotypes_p1_cd8_selected])

# %% [code]
clonotypes_p1_cd4_counterselected_final = tab_clonotypes_p1_filtered.loc[np.logical_and(
    tab_clonotypes_p1_filtered["label_cell_type"].values == "Cd4",
    np.logical_and(
        np.logical_and(
            tab_clonotypes_p1_filtered["n_stimulated"].values >= 3,
            tab_clonotypes_p1_filtered["n_unstimulated"].values >= 3
        ),
        [x not in clonotypes_p1_cd4_selected_final for x in tab_clonotypes_p1_filtered["clonotype"].values]
    )
), "clonotype"].values
clonotypes_p1_cd8_counterselected_final = tab_clonotypes_p1_filtered.loc[np.logical_and(
    tab_clonotypes_p1_filtered["label_cell_type"].values == "Cd8",
    np.logical_and(
        np.logical_and(
            tab_clonotypes_p1_filtered["n_stimulated"].values >= 3,
            tab_clonotypes_p1_filtered["n_unstimulated"].values >= 3
        ),
        [x not in clonotypes_p1_cd8_selected_final for x in tab_clonotypes_p1_filtered["clonotype"].values]
    )
), "clonotype"].values

# %% [code]
print(
    (
        "P1 CD4+:\ncells in manually selected clonotypes %i cells,\n" +
        "cells in clonotypes that are in IFNG+cluster and appear in both conditions %i cells,\n" +
        "cells in clonotypes that are not in IFNG+ cluster and appear in both conditions >=3 %i cells.\n"
    ) % (
        tab_clonotypes_p1_filtered.loc[
            [x in clonotypes_p1_cd4_selected_final for x in tab_clonotypes_p1_filtered["clonotype"].values], 
            "size"
        ].values.sum(),
        tab_clonotypes_p1_filtered.loc[
            [x in clonotypes_p1_cd4_selected for x in tab_clonotypes_p1_filtered["clonotype"].values], 
            "size"
        ].values.sum(),
        tab_clonotypes_p1_filtered.loc[
            [x in clonotypes_p1_cd4_counterselected_final for x in tab_clonotypes_p1_filtered["clonotype"].values], 
            "size"
        ].values.sum(),
    )
)
print(
    (
        "P1 CD8+:\ncells in manually selected clonotypes %i cells,\n" +
        "cells in clonotypes that are in IFNG+cluster and appear in both conditions %i cells,\n" +
        "cells in clonotypes that are not in IFNG+ cluster and appear in both conditions >=3 %i cells.\n"
    ) % (
        tab_clonotypes_p1_filtered.loc[
            [x in clonotypes_p1_cd8_selected_final for x in tab_clonotypes_p1_filtered["clonotype"].values], 
            "size"
        ].values.sum(),
        tab_clonotypes_p1_filtered.loc[
            [x in clonotypes_p1_cd8_selected for x in tab_clonotypes_p1_filtered["clonotype"].values], 
            "size"
        ].values.sum(),
        tab_clonotypes_p1_filtered.loc[
            [x in clonotypes_p1_cd8_counterselected_final for x in tab_clonotypes_p1_filtered["clonotype"].values], 
            "size"
        ].values.sum(),
    )
)

# %% [code]
print(tab_clonotypes_p1_filtered.loc[
    [x in clonotypes_p1_cd4_selected_final for x in tab_clonotypes_p1_filtered["clonotype"].values], 
    ["clonotype", "clonotype_merged", "in_ifng_cluster", "ave_ifng+"]
])
print(tab_clonotypes_p1_filtered.loc[
    [x in clonotypes_p1_cd8_selected_final for x in tab_clonotypes_p1_filtered["clonotype"].values], 
    ["clonotype", "clonotype_merged", "in_ifng_cluster", "ave_ifng+"]
])

# %% [code]
clonotypes_p1_selected = clonotypes_p1_cd4_selected_final.tolist() + clonotypes_p1_cd8_selected_final.tolist()
clonotypes_p2_selected = clonotypes_p2_cd4_selected.tolist() + clonotypes_p2_cd8_selected.tolist()

# %% [code]
fig, axs = plt.subplots(nrows=1, ncols=1, figsize=(5, 5))
sc.pl.umap(adata_proc_p1, ax=axs, show=False)
sc.pl.umap(
    adata_proc_p1[np.logical_and(
        np.array([
            xx in clonotypes_p1_cd8_selected_final and y != "None" and z != "None"
            for xx, y, z in zip(
                adata_proc_p1.obs["clonotype_merged"].values,
                adata_proc_p1.obs["TRA_1_cdr3"].values,
                adata_proc_p1.obs["TRB_1_cdr3"].values
            )
        ]),
        adata_proc_p1.obs["condition"].values == "unstimulated"
    )], 
    color="clonotype_merged",
    size=40,
    use_raw=False,
    ax=axs, 
    show=False
)
plt.tight_layout()
plt.savefig(dir_panels + "fig3c2.pdf")
plt.show()

fig, axs = plt.subplots(nrows=1, ncols=1, figsize=(5,5))
sc.pl.umap(adata_proc_p1, ax=axs, show=False)
sc.pl.umap(
    adata_proc_p1[np.logical_and(
        np.array([
            xx in clonotypes_p1_cd4_selected_final and y != "None" and z != "None"
            for xx, y, z in zip(
                adata_proc_p1.obs["clonotype_merged"].values,
                adata_proc_p1.obs["TRA_1_cdr3"].values,
                adata_proc_p1.obs["TRB_1_cdr3"].values
            )
        ]),
        adata_proc_p1.obs["condition"].values == "unstimulated"
    )], 
    color="clonotype_merged",
    size=40,
    use_raw=False,
    ax=axs, 
    show=False
)
plt.tight_layout()
plt.savefig(dir_panels + "fig3c5.pdf")
plt.show()

# %% [code]
marker_keys = [
    "IFNG",
] 

ncols = 2
nrows = len(marker_keys)
fig, axs = plt.subplots(nrows=nrows, ncols=ncols, figsize=(ncols*4, 4*nrows))
axs = axs.ravel()

i=0
sc.pl.umap(adata_proc_p1, ax=axs[i*ncols], show=False)
sc.pl.umap(
    adata_proc_p1[np.logical_and(
        np.array([
            xx in clonotypes_p1_cd4_selected_final 
            for xx in adata_proc_p1.obs["clonotype"].values
        ]),
        adata_proc_p1.obs["condition"].values == "unstimulated"
    )], 
    color="clonotype",
    size=40,
    use_raw=False,
    ax=axs[i*ncols], 
    show=False
)
axs[i*ncols].set_ylabel("%s\nUMAP2" % x)

sc.pl.umap(adata_proc_p1, ax=axs[i*ncols+1], show=False)
sc.pl.umap(
    adata_proc_p1[np.logical_and(
        np.array([
            xx in clonotypes_p1_cd4_selected_final 
            for xx in adata_proc_p1.obs["clonotype"].values
        ]),
        adata_proc_p1.obs["condition"].values == "stimulated"
    )], 
    color="clonotype",
    size=40,
    use_raw=False,
    ax=axs[i*ncols+1], 
    show=False
)
axs[i*ncols].set_title("unstimulated")
axs[i*ncols+1].set_title("stimulated")
plt.tight_layout()
plt.savefig(dir_panels + "sfig7b1.pdf")
plt.show()

ncols = 2
nrows = len(marker_keys)
fig, axs = plt.subplots(nrows=nrows, ncols=ncols, figsize=(ncols*4, 4*nrows))
axs = axs.ravel()

for i, x in enumerate(marker_keys):
    sc.pl.umap(adata_proc_p1, ax=axs[i*ncols], show=False)
    sc.pl.umap(
        adata_proc_p1[np.logical_and(
            np.array([
                xx in clonotypes_p1_cd4_selected_final.tolist()
                for xx in adata_proc_p1.obs["clonotype"].values
            ]),
            adata_proc_p1.obs["condition"].values == "unstimulated"
        )], 
        color=x,
        vmin=0,
        vmax=adata_proc_p1[
            np.array([
                xx in clonotypes_p1_cd4_selected_final.tolist()
                for xx in adata_proc_p1.obs["clonotype"].values
            ]), x
        ].X.max(),
        size=40,
        use_raw=False,
        ax=axs[i*ncols], 
        show=False
    )
    axs[i*ncols].set_ylabel("%s\nUMAP2" % x)
    
    sc.pl.umap(adata_proc_p1, ax=axs[i*ncols+1], show=False)
    sc.pl.umap(
        adata_proc_p1[np.logical_and(
            np.array([
                xx in clonotypes_p1_cd4_selected_final.tolist()
                for xx in adata_proc_p1.obs["clonotype"].values
            ]),
            adata_proc_p1.obs["condition"].values == "stimulated"
        )], 
        color=x,
        vmin=0,
        vmax=adata_proc_p1[
            np.array([
                xx in clonotypes_p1_cd4_selected_final.tolist()
                for xx in adata_proc_p1.obs["clonotype"].values
            ]), x
        ].X.max(),
        size=40,
        use_raw=False,
        ax=axs[i*ncols+1], 
        show=False
    )
    if i == 0:
        axs[i*ncols].set_title("unstimulated")
        axs[i*ncols+1].set_title("stimulated")
    else:
        axs[i*ncols].set_title(" ")
        axs[i*ncols+1].set_title(" ")
plt.tight_layout()
plt.savefig(dir_panels + "sfig7a1.pdf")
plt.show()

# %% [code]
marker_keys = [
    "IFNG",
] 

ncols = 2
nrows = 1
fig, axs = plt.subplots(nrows=nrows, ncols=ncols, figsize=(ncols*4, 4*nrows))
axs = axs.ravel()

i=0
sc.pl.umap(adata_proc_p1, ax=axs[i*ncols], show=False)
sc.pl.umap(
    adata_proc_p1[np.logical_and(
        np.array([
            xx in clonotypes_p1_cd8_selected_final 
            for xx in adata_proc_p1.obs["clonotype"].values
        ]),
        adata_proc_p1.obs["condition"].values == "unstimulated"
    )], 
    color="clonotype",
    size=40,
    use_raw=False,
    ax=axs[i*ncols], 
    show=False
)
axs[i*ncols].set_ylabel("%s\nUMAP2" % x)

sc.pl.umap(adata_proc_p1, ax=axs[i*ncols+1], show=False)
sc.pl.umap(
    adata_proc_p1[np.logical_and(
        np.array([
            xx in clonotypes_p1_cd8_selected_final 
            for xx in adata_proc_p1.obs["clonotype"].values
        ]),
        adata_proc_p1.obs["condition"].values == "stimulated"
    )], 
    color="clonotype",
    size=40,
    use_raw=False,
    ax=axs[i*ncols+1], 
    show=False
)
axs[i*ncols].set_title("unstimulated")
axs[i*ncols+1].set_title("stimulated")
plt.tight_layout()
plt.savefig(dir_panels + "sfig7b2.pdf")
plt.show()

ncols = 2
nrows = len(marker_keys)
fig, axs = plt.subplots(nrows=nrows, ncols=ncols, figsize=(ncols*4, 4*nrows))
axs = axs.ravel()

for i, x in enumerate(marker_keys):
    sc.pl.umap(adata_proc_p1, ax=axs[i*ncols], show=False)
    sc.pl.umap(
        adata_proc_p1[np.logical_and(
            np.array([
                xx in clonotypes_p1_cd8_selected_final.tolist() 
                for xx in adata_proc_p1.obs["clonotype"].values
            ]),
            adata_proc_p1.obs["condition"].values == "unstimulated"
        )], 
        color=x,
        vmin=0,
        vmax=adata_proc_p1[
            np.array([
                xx in clonotypes_p1_cd8_selected_final.tolist()
                for xx in adata_proc_p1.obs["clonotype"].values
            ]), x
        ].X.max(),
        size=40,
        use_raw=False,
        ax=axs[i*ncols], 
        show=False
    )
    axs[i*ncols].set_ylabel("%s\nUMAP2" % x)
    
    sc.pl.umap(adata_proc_p1, ax=axs[i*ncols+1], show=False)
    sc.pl.umap(
        adata_proc_p1[np.logical_and(
            np.array([
                xx in clonotypes_p1_cd8_selected_final.tolist()
                for xx in adata_proc_p1.obs["clonotype"].values
            ]),
            adata_proc_p1.obs["condition"].values == "stimulated"
        )], 
        color=x,
        vmin=0,
        vmax=adata_proc_p1[
            np.array([
                xx in clonotypes_p1_cd8_selected_final.tolist() 
                for xx in adata_proc_p1.obs["clonotype"].values
            ]), x
        ].X.max(),
        size=40,
        use_raw=False,
        ax=axs[i*ncols+1], 
        show=False
    )
    if i == 0:
        axs[i*ncols].set_title("unstimulated")
        axs[i*ncols+1].set_title("stimulated")
    else:
        axs[i*ncols].set_title(" ")
        axs[i*ncols+1].set_title(" ")
plt.tight_layout()
plt.savefig(dir_panels + "sfig7a2.pdf")
plt.show()

# %% [markdown]
### Save TCR genes of reactive clonotypes

# %% [code]
tab_clonotypes_p1_filtered.loc[clonotypes_p1_cd4_selected_final, [x for x in tab_clonotypes_p1_filtered.columns if x.split("_")[-1] == "gene"]]\
.to_csv(dir_tables + "p1_cd4_reactive_clonotypes_tcr_genes.csv", sep=";")
tab_clonotypes_p1_filtered.loc[clonotypes_p1_cd8_selected_final, [x for x in tab_clonotypes_p1_filtered.columns if x.split("_")[-1] == "gene"]]\
.to_csv(dir_tables + "p1_cd8_reactive_clonotypes_tcr_genes.csv", sep=";")

# %% [markdown]
### Store in adata

# %% [code]
adata_proc_p1.obs["reactive"] = [
    "reactive" if x in clonotypes_p1_cd4_selected_final.tolist() + clonotypes_p1_cd8_selected_final.tolist()
    else "nonreactive" if x in clonotypes_p1_cd4_counterselected_final.tolist() + clonotypes_p1_cd8_counterselected_final.tolist()
    else "few cells"
    for x in adata_proc_p1.obs["clonotype"].values
]
adata_proc_p2.obs["reactive"] = [
    "reactive" if x in clonotypes_p2_cd4_selected.tolist() + clonotypes_p2_cd8_selected.tolist()
    else "nonreactive"
    for x in adata_proc_p2.obs["clonotype"].values
]
adata_proc_p1.obs["reactive_bool"] = adata_proc_p1.obs["reactive"].values == "reactive"
adata_proc_p2.obs["reactive_bool"] = adata_proc_p2.obs["reactive"].values == "reactive"

# %% [code]
adata_proc_p1.obs.groupby(["reactive", "condition", "cell_type"]).size()

# %% [markdown]
## Plot marker expression by reactive and non-reactive clonotype group

# %% [code]
adata_proc_p1.obs["reactive_condition"] = [
    y+"_"+str(x) if x in ["reactive", "nonreactive"] else "other" for x, y in zip(
        adata_proc_p1.obs["reactive"].values,
        adata_proc_p1.obs["condition"].values
    )
]
adata_proc_p2.obs["reactive_condition"] = [
    y+"_"+str(x) if x in ["reactive", "nonreactive"] else "other" for x, y in zip(
        adata_proc_p2.obs["reactive"].values,
        adata_proc_p2.obs["condition"].values
    )
]

# %% [markdown]
### Cd4

# %% [code]
adata_proc_p1[np.logical_and(
    adata_proc_p1.obs["cell_type"] == "Cd4",
    [x in ["reactive", "nonreactive"] for x in adata_proc_p1.obs["reactive"]]
)].obs["reactive_condition"]

# %% [code]
adata_temp = adata_proc_p1[np.logical_and(
    adata_proc_p1.obs["cell_type"] == "Cd4",
    [x in ["reactive", "nonreactive"] for x in adata_proc_p1.obs["reactive"]]
)].copy()
adata_temp.obs["reactive_condition"] = pd.Categorical(
    adata_temp.obs["reactive_condition"].values,
    categories=[
        'stimulated_reactive',
        'unstimulated_reactive',
        'unstimulated_nonreactive',
        'stimulated_nonreactive', 
    ]
)
for k, v in markers_2c.items():
    sc.pl.dotplot(
        adata_temp, 
        var_names=[x for x in v if x in adata_temp.var_names], 
        groupby="reactive_condition", 
        use_raw=False,
        save="fig3a_cd4_" + k.lower() + ".pdf",
        show=False
    )
sc.pl.dotplot(
    adata_temp, 
    var_names=[x for x in markers_s6ab["cd4"] if x in adata_temp.var_names], 
    groupby="reactive_condition", 
    use_raw=False,
    save="sfig6a_cd4.pdf",
    show=False
)
for k, v in markers_cd4_ta.items():
    sc.pl.dotplot(
        adata_temp, 
        var_names=[x for x in v if x in adata_temp.var_names], 
        groupby="reactive_condition" ,
        use_raw=False,
        save="sfigx_pbmc_" + "Cd4".lower() + "_tcells_reactive_condition_" + k + ".pdf",
        show=False
    )
del adata_temp

# %% [markdown]
### Cd8

# %% [code]
adata_temp = adata_proc_p1[np.logical_and(
    adata_proc_p1.obs["cell_type"] == "Cd8",
    [x in ["reactive", "nonreactive"] for x in adata_proc_p1.obs["reactive"]]
)].copy()
adata_temp.obs["reactive_condition"] = pd.Categorical(
    adata_temp.obs["reactive_condition"].values,
    categories=[
        'stimulated_reactive',
        'unstimulated_reactive',
        'unstimulated_nonreactive',
        'stimulated_nonreactive', 
    ]
)
sc.pl.dotplot(
    adata_temp, 
    var_names=[x for x in markers_s6ab["cd8"] if x in adata_temp.var_names], 
    groupby="reactive_condition", 
    use_raw=False,
    save="sfig6b_cd8.pdf",
    show=False
)
for k, v in markers_cd8_ta.items():
    sc.pl.dotplot(
        adata_temp, 
        var_names=[x for x in v if x in adata_temp.var_names], 
        groupby="reactive_condition", 
        use_raw=False,
        save="sfigx_pbmc_" + "Cd8".lower() + "_tcells_reactive_condition_" + k + ".pdf",
        show=False
    )
del adata_temp

# %% [markdown]
## Differential expression across joint responsive clonotypes

# %% [code]
tcr_genes_p1 = np.sort(np.unique([
    x for x in adata_proc_p1.var_names.tolist()
    if np.any([
        x.startswith(xx) for xx in [
            "TRAV", #"TRAJ", #"TRAC", 
            "TRBV", #"TRBJ", #"TRBC", "
        ]
    ]) 
]))
print(tcr_genes_p1)

# %% [markdown]
### Ifng+ cluster vs rest

# %% [code]
det = de.test.t_test(
    adata_proc_p1[
        np.logical_and(
            adata_proc_p1.obs["cell_type"].values == "Cd4",
            adata_proc_p1.obs["condition"].values == "stimulated",
        ),
        [x not in tcr_genes_p1 for x in adata_proc_p1.var_names]
    ],
    grouping="ifng_pos_cluster", 
    is_logged=True
)
tab = det.summary()
print(np.unique(det.grouping))
tab["log2fc["+str(np.unique(det.grouping)[1])+"]"] = tab["log2fc"].values
idx = np.logical_and(
    tab["qval"].values < 0.05,
    np.logical_and(
        tab["mean"].values >= np.log(0.1),
        np.abs(tab["log2fc"].values) >= np.log(1.1)
    )
)
tab.loc[idx, :].sort_values("log2fc").to_csv(dir_tables+'tab_de_p1_cd4_infg_cluster.csv', sep=";")
tab.sort_values("log2fc").to_csv(dir_tables+'tab_de_p1_cd4_infg_cluster_all.csv', sep=";")
tab.sort_values("log2fc").to_csv(dir_tables+'ExtendedDataTable3_Cd4Tcells_testfor_IFNG_cluster.csv', sep=";")

# %% [code]
det = de.test.t_test(
    adata_proc_p1[
        np.logical_and(
            adata_proc_p1.obs["cell_type"].values == "Cd8",
            adata_proc_p1.obs["condition"].values == "stimulated",
        ),
        [x not in tcr_genes_p1 for x in adata_proc_p1.var_names]
    ],
    grouping="ifng_pos_cluster", 
    is_logged=True
)
tab = det.summary()
print(np.unique(det.grouping))
tab["log2fc["+str(np.unique(det.grouping)[1])+"]"] = tab["log2fc"].values
idx = np.logical_and(
    tab["qval"].values < 0.05,
    np.logical_and(
        tab["mean"].values >= np.log(0.1),
        np.abs(tab["log2fc"].values) >= np.log(1.1)
    )
)
tab.loc[idx, :].sort_values("log2fc").to_csv(dir_tables+'tab_de_p1_cd8_infg_cluster.csv', sep=";")
tab.sort_values("log2fc").to_csv(dir_tables+'tab_de_p1_cd8_infg_cluster_all.csv', sep=";")
tab.sort_values("log2fc").to_csv(dir_tables+'ExtendedDataTable2_Cd8Tcells_testfor_IFNG_cluster.csv', sep=";")

# %% [markdown]
### Responsive vs non-responsive in unstim

# %% [code]
det = de.test.t_test(
    adata_proc_p1[
        np.logical_and(
            adata_proc_p1.obs["cell_type"].values == "Cd4",
            np.logical_and(
                adata_proc_p1.obs["condition"].values == "unstimulated",
                [x in ["reactive", "nonreactive"] for x in adata_proc_p1.obs["reactive"]]
            )
        ),
        [x not in tcr_genes_p1 for x in adata_proc_p1.var_names]
    ],
    grouping="reactive_bool", 
    is_logged=True
)
tab = det.summary()
print(np.unique(det.grouping))
tab["log2fc["+str(np.unique(det.grouping)[1])+"]"] = tab["log2fc"].values
de_genes_p1_cd4_responsive_unstim = tab.loc[tab["qval"].values < 0.01, "gene"].values
et = de.enrich.test(
    ref=rs,
    det=det,
    threshold=0.01,
)
significant_set_ids_p1_cd4_responsive_unstim = et.significant_set_ids()
print(len(de_genes_p1_cd4_responsive_unstim))
idx = np.logical_and(
    tab["qval"].values < 0.05,
    np.logical_and(
        tab["mean"].values >= np.log(0.1),
        np.abs(tab["log2fc"].values) >= np.log(1.1)
    )
)
tab.loc[idx, :].sort_values("log2fc").to_csv(dir_tables+'tab_de_p1_cd4_unstimulated_reactiveness.csv', sep=";")
tab.sort_values("log2fc").to_csv(dir_tables+'tab_de_p1_cd4_unstimulated_reactiveness_all.csv', sep=";")
tab.sort_values("log2fc").to_csv(dir_tables+'ExtendedDataTable6_unstimulated_Cd4Tcells_testfor_reactiveness.csv', sep=";")
tab_figure = tab[["gene", "log2fc", "qval"]].copy()
tab_figure["qval_significant"] = [
    str(x) if y else ""
    for x, y in zip(tab_figure["qval"].values, idx)
]
tab_figure["qval_nonsignificant"] = [
    "" if y else str(x)
    for x, y in zip(tab_figure["qval"].values, idx)
]
tab_figure.sort_values("log2fc").to_csv(dir_tables+'fig3d2_cd4_unstim.csv', sep=";")
tab_figure.sort_values("log2fc").to_csv(dir_tables+'sfig8a_cd4_unstim.csv', sep=";")

de_genes_unstim_cd4_responsive_all = tab.loc[idx, "gene"].values
direction_de_genes_unstim_cd4_responsive_all = ((tab.loc[idx, "log2fc"].values > 0)-0.5)*2.

# %% [code]
de_genes_unstim_cd4_responsive_all

# %% [code]
adata_proc_p1.obs["score_cd4_unstimulated_reactiveness"] = (np.asarray(
    (
        adata_proc_p1[:, de_genes_unstim_cd4_responsive_all].X - 
        adata_proc_p1[:, de_genes_unstim_cd4_responsive_all].X.mean(axis=0)
    ) / adata_proc_p1[:, de_genes_unstim_cd4_responsive_all].X.todense().std(axis=0)
)*direction_de_genes_unstim_cd4_responsive_all).mean(axis=1).flatten()
sb.catplot(
    data=adata_proc_p1.obs, 
    y="score_cd4_unstimulated_reactiveness", 
    x="condition", 
    hue="reactive_bool", 
    col="cell_type",
    kind="violin",
    split=True,
    ax=ax
)
plt.tight_layout()
plt.savefig(dir_panels + "violinsplit_p1_pbmc_score_cd4_unstimulated_reactiveness.pdf")

# %% [code]
det = de.test.t_test(
    adata_proc_p1[
        np.logical_and(
            adata_proc_p1.obs["cell_type"].values == "Cd8",
            np.logical_and(
                adata_proc_p1.obs["condition"].values == "unstimulated",
                [x in ["reactive", "nonreactive"] for x in adata_proc_p1.obs["reactive"]]
            )
        ),
        [x not in tcr_genes_p1 for x in adata_proc_p1.var_names]
    ],
    grouping="reactive_bool", 
    is_logged=True
)
tab = det.summary()
print(np.unique(det.grouping))
tab["log2fc["+str(np.unique(det.grouping)[1])+"]"] = tab["log2fc"].values
de_genes_p1_cd8_responsive_unstim = tab.loc[tab["qval"].values < 0.01, "gene"].values
et = de.enrich.test(
    ref=rs,
    det=det,
    threshold=0.01,
)
significant_set_ids_p1_cd8_responsive_unstim = et.significant_set_ids()
print(len(de_genes_p1_cd8_responsive_unstim))
idx = np.logical_and(
    tab["qval"].values < 0.05,
    np.logical_and(
        tab["mean"].values >= np.log(0.1),
        np.abs(tab["log2fc"].values) >= np.log(1.1)
    )
)
tab.loc[idx, :].sort_values("log2fc").to_csv(dir_tables+'tab_de_p1_cd8_unstimulated_reactiveness.csv', sep=";")
tab.sort_values("log2fc").to_csv(dir_tables+'tab_de_p1_cd8_unstimulated_reactiveness_all.csv', sep=";")
tab.sort_values("log2fc").to_csv(dir_tables+'ExtendedDataTable7_unstimulated_C84Tcells_testfor_reactiveness.csv', sep=";")

tab_figure = tab[["gene", "log2fc", "qval"]].copy()
tab_figure["qval_significant"] = [
    str(x) if y else ""
    for x, y in zip(tab_figure["qval"].values, idx)
]
tab_figure["qval_nonsignificant"] = [
    "" if y else str(x)
    for x, y in zip(tab_figure["qval"].values, idx)
]
tab_figure.sort_values("log2fc").to_csv(dir_tables+'fig3d1_cd8_unstim.csv', sep=";")
tab_figure.sort_values("log2fc").to_csv(dir_tables+'sfig8b_cd4_unstim.csv', sep=";")

de_genes_unstim_cd8_responsive_all = tab.loc[idx, "gene"].values
direction_de_genes_unstim_cd8_responsive_all = ((tab.loc[idx, "log2fc"].values > 0)-0.5)*2.

# %% [code]
de_genes_unstim_cd8_responsive_all

# %% [code]
adata_proc_p1.obs["score_cd8_unstimulated_reactiveness"] = (np.asarray(
    (
        adata_proc_p1[:, de_genes_unstim_cd8_responsive_all].X - 
        adata_proc_p1[:, de_genes_unstim_cd8_responsive_all].X.mean(axis=0)
    ) / adata_proc_p1[:, de_genes_unstim_cd8_responsive_all].X.todense().std(axis=0)
)*direction_de_genes_unstim_cd8_responsive_all).mean(axis=1).flatten()
sb.catplot(
    data=adata_proc_p1.obs, 
    y="score_cd8_unstimulated_reactiveness", 
    x="condition", 
    hue="reactive_bool", 
    col="cell_type",
    kind="violin",
    split=True,
    ax=ax
)
plt.tight_layout()
plt.savefig(dir_panels + "violinsplit_p1_pbmc_score_cd8_unstimulated_reactiveness.pdf")

# %% [markdown]
### Responsive vs non-responsive in stim

# %% [code]
det = de.test.t_test(
    adata_proc_p1[
        np.logical_and(
            adata_proc_p1.obs["cell_type"].values == "Cd4",
            np.logical_and(
                adata_proc_p1.obs["condition"].values == "stimulated",
                [x in ["reactive", "nonreactive"] for x in adata_proc_p1.obs["reactive"]]
            ),
        ),
        [x not in tcr_genes_p1 for x in adata_proc_p1.var_names]
    ],
    grouping="reactive_bool", 
    is_logged=True
)
tab = det.summary()
print(np.unique(det.grouping))
tab["log2fc["+str(np.unique(det.grouping)[1])+"]"] = tab["log2fc"].values
de_genes_p1_cd4_responsive_stim = tab.loc[tab["qval"].values < 0.01, "gene"].values
et = de.enrich.test(
    ref=rs,
    det=det,
    threshold=0.01,
)
significant_set_ids_p1_cd4_responsive_stim = et.significant_set_ids()
print(len(de_genes_p1_cd4_responsive_stim))
idx = np.logical_and(
    tab["qval"].values < 0.05,
    np.logical_and(
        tab["mean"].values >= np.log(0.1),
        np.abs(tab["log2fc"].values) >= np.log(1.1)
    )
)
tab.loc[idx, :].sort_values("log2fc").to_csv(dir_tables+'tab_de_p1_cd4_stimulated_reactiveness.csv', sep=";")
tab.sort_values("log2fc").to_csv(dir_tables+'tab_de_p1_cd4_stimulated_reactiveness_all.csv', sep=";")
tab.sort_values("log2fc").to_csv(dir_tables+'ExtendedDataTable8_stimulated_Cd4Tcells_testfor_reactiveness.csv', sep=";")

tab_figure = tab[["gene", "log2fc", "qval"]].copy()
tab_figure["qval_significant"] = [
    str(x) if y else ""
    for x, y in zip(tab_figure["qval"].values, idx)
]
tab_figure["qval_nonsignificant"] = [
    "" if y else str(x)
    for x, y in zip(tab_figure["qval"].values, idx)
]
tab_figure.sort_values("log2fc").to_csv(dir_tables+'sfig8c_cd4_stim.csv', sep=";")

de_genes_stim_cd4_responsive_all = tab.loc[idx, "gene"].values
direction_de_genes_stim_cd4_responsive_all = ((tab.loc[idx, "log2fc"].values > 0)-0.5)*2.

# %% [code]
de_genes_stim_cd4_responsive_all

# %% [code]
adata_proc_p1.obs["score_cd4_stimulated_reactiveness"] = (np.asarray(
    (
        adata_proc_p1[:, de_genes_stim_cd4_responsive_all].X - 
        adata_proc_p1[:, de_genes_stim_cd4_responsive_all].X.mean(axis=0)
    ) / adata_proc_p1[:, de_genes_stim_cd4_responsive_all].X.todense().std(axis=0)
)*direction_de_genes_stim_cd4_responsive_all).mean(axis=1).flatten()
sb.catplot(
    data=adata_proc_p1.obs, 
    y="score_cd4_stimulated_reactiveness", 
    x="condition", 
    hue="reactive_bool", 
    col="cell_type",
    kind="violin",
    split=True,
    ax=ax
)
plt.tight_layout()
plt.savefig(dir_panels + "violinsplit_p1_pbmc_score_cd4_stimulated_reactiveness.pdf")

# %% [code]
det = de.test.t_test(
    adata_proc_p1[
        np.logical_and(
            adata_proc_p1.obs["cell_type"].values == "Cd8",
            np.logical_and(
                adata_proc_p1.obs["condition"].values == "stimulated",
                [x in ["reactive", "nonreactive"] for x in adata_proc_p1.obs["reactive"]]
            )
        ),
        [x not in tcr_genes_p1 for x in adata_proc_p1.var_names]
    ],
    grouping="reactive_bool", 
    is_logged=True
)
tab = det.summary()
print(np.unique(det.grouping))
tab["log2fc["+str(np.unique(det.grouping)[1])+"]"] = tab["log2fc"].values
de_genes_p1_cd8_responsive_stim = tab.loc[tab["qval"].values < 0.01, "gene"].values
et = de.enrich.test(
    ref=rs,
    det=det,
    threshold=0.01,
)
significant_set_ids_p1_cd8_responsive_stim = et.significant_set_ids()
print(len(de_genes_p1_cd8_responsive_stim))
idx = np.logical_and(
    tab["qval"].values < 0.05,
    np.logical_and(
        tab["mean"].values >= np.log(0.1),
        np.abs(tab["log2fc"].values) >= np.log(1.1)
    )
)
tab.loc[idx, :].sort_values("log2fc").to_csv(dir_tables+'tab_de_p1_cd8_stimulated_reactiveness.csv', sep=";")
tab.sort_values("log2fc").to_csv(dir_tables+'tab_de_p1_cd8_stimulated_reactiveness_all.csv', sep=";")
tab.sort_values("log2fc").to_csv(dir_tables+'ExtendedDataTable9_stimulated_Cd8Tcells_testfor_reactiveness.csv', sep=";")

tab_figure = tab[["gene", "log2fc", "qval"]].copy()
tab_figure["qval_significant"] = [
    str(x) if y else ""
    for x, y in zip(tab_figure["qval"].values, idx)
]
tab_figure["qval_nonsignificant"] = [
    "" if y else str(x)
    for x, y in zip(tab_figure["qval"].values, idx)
]
tab_figure.sort_values("log2fc").to_csv(dir_tables+'sfig8b_cd8_stim.csv', sep=";")

de_genes_stim_cd8_responsive_all = tab.loc[idx, "gene"].values
direction_de_genes_stim_cd8_responsive_all = ((tab.loc[idx, "log2fc"].values > 0)-0.5)*2.

# %% [code]
de_genes_stim_cd8_responsive_all

# %% [code]
adata_proc_p1.obs["score_cd8_stimulated_reactiveness"] = (np.asarray(
    (
        adata_proc_p1[:, de_genes_stim_cd8_responsive_all].X - 
        adata_proc_p1[:, de_genes_stim_cd8_responsive_all].X.mean(axis=0)
    ) / adata_proc_p1[:, de_genes_stim_cd8_responsive_all].X.todense().std(axis=0)
)*direction_de_genes_stim_cd8_responsive_all).mean(axis=1).flatten()
sb.catplot(
    data=adata_proc_p1.obs, 
    y="score_cd8_stimulated_reactiveness", 
    x="condition", 
    hue="reactive_bool", 
    col="cell_type",
    kind="violin",
    split=True,
    ax=ax
)
plt.tight_layout()
plt.savefig(dir_panels + "violinsplit_p1_pbmc_score_cd8_stimulated_reactiveness.pdf")

# %% [markdown]
### Unstimulated vs stimulated in reactive

# %% [code]
det = de.test.t_test(
    adata_proc_p1[
        np.logical_and(
            adata_proc_p1.obs["cell_type"].values == "Cd4",
            np.array([
                xx in clonotypes_p1_cd4_selected_final 
                for xx in adata_proc_p1.obs["clonotype"].values
            ]),
        ),
        [x not in tcr_genes_p1 for x in adata_proc_p1.var_names]
    ],
    grouping="condition", 
    is_logged=True
)
tab = det.summary()
print(np.unique(det.grouping))
tab["log2fc["+str(np.unique(det.grouping)[1])+"]"] = tab["log2fc"].values
de_genes_p1_cd4_responsive = tab.loc[tab["qval"].values < 0.01, "gene"].values
et = de.enrich.test(
    ref=rs,
    det=det,
    threshold=0.01,
)
significant_set_ids_p1_cd4_responsive = et.significant_set_ids()
print(len(de_genes_p1_cd4_responsive))
idx = np.logical_and(
    tab["qval"].values < 0.05,
    np.logical_and(
        tab["mean"].values >= np.log(0.1),
        np.abs(tab["log2fc"].values) >= np.log(1.1)
    )
)
tab.loc[idx, :].sort_values("log2fc").to_csv(dir_tables+'tab_de_p1_cd4_reactive_condition.csv', sep=";")
tab.sort_values("log2fc").to_csv(dir_tables+'tab_de_p1_cd4_reactive_condition_all.csv', sep=";")
tab.sort_values("log2fc").to_csv(dir_tables+'ExtendedDataTable4_reactive_Cd4Tcells_testfor_condition.csv', sep=";")

tab_figure = tab[["gene", "log2fc", "qval"]].copy()
tab_figure["qval_significant"] = [
    str(x) if y else ""
    for x, y in zip(tab_figure["qval"].values, idx)
]
tab_figure["qval_nonsignificant"] = [
    "" if y else str(x)
    for x, y in zip(tab_figure["qval"].values, idx)
]
tab_figure.sort_values("log2fc").to_csv(dir_tables+'sfig6c_cd4.csv', sep=";")

de_genes_cd4_responsive_all = tab.loc[idx, "gene"].values
direction_de_genes_cd4_responsive_all = -((tab.loc[idx, "log2fc"].values > 0)-0.5)*2.

# %% [code]
de_genes_cd4_responsive_all

# %% [code]
adata_proc_p1.obs["score_cd4_reactive_condition"] = (np.asarray(
    (
        adata_proc_p1[:, de_genes_cd4_responsive_all].X - 
        adata_proc_p1[:, de_genes_cd4_responsive_all].X.mean(axis=0)
    ) / adata_proc_p1[:, de_genes_cd4_responsive_all].X.todense().std(axis=0)
)*direction_de_genes_cd4_responsive_all).mean(axis=1).flatten()
sb.catplot(
    data=adata_proc_p1.obs, 
    y="score_cd4_reactive_condition", 
    hue="condition", 
    x="reactive_bool", 
    col="cell_type",
    kind="violin",
    split=True,
    ax=ax
)
plt.tight_layout()
plt.savefig(dir_panels + "violinsplit_p1_pbmc_score_cd4_reactive_condition.pdf")

# %% [code]
det = de.test.t_test(
    adata_proc_p1[
        np.logical_and(
            adata_proc_p1.obs["cell_type"].values == "Cd8",
            np.array([
                xx in clonotypes_p1_cd8_selected_final 
                for xx in adata_proc_p1.obs["clonotype"].values
            ]),
        ),
        [x not in tcr_genes_p1 for x in adata_proc_p1.var_names]
    ],
    grouping="condition", 
    is_logged=True
)
tab = det.summary()
print(np.unique(det.grouping))
tab["log2fc["+str(np.unique(det.grouping)[1])+"]"] = tab["log2fc"].values
de_genes_p1_cd8_responsive = tab.loc[tab["qval"].values < 0.01, "gene"].values
et = de.enrich.test(
    ref=rs,
    det=det,
    threshold=0.01,
)
significant_set_ids_p1_cd8_responsive = et.significant_set_ids()
print(len(de_genes_p1_cd8_responsive))
idx = np.logical_and(
    tab["qval"].values < 0.05,
    np.logical_and(
        tab["mean"].values >= np.log(0.1),
        np.abs(tab["log2fc"].values) >= np.log(1.1)
    )
)
tab.loc[idx, :].sort_values("log2fc").to_csv(dir_tables+'tab_de_p1_cd8_reactive_condition.csv', sep=";")
tab.sort_values("log2fc").to_csv(dir_tables+'tab_de_p1_cd8_reactive_condition_all.csv', sep=";")
tab.sort_values("log2fc").to_csv(dir_tables+'ExtendedDataTable5_reactive_Cd8Tcells_testfor_condition.csv', sep=";")

tab_figure = tab[["gene", "log2fc", "qval"]].copy()
tab_figure["qval_significant"] = [
    str(x) if y else ""
    for x, y in zip(tab_figure["qval"].values, idx)
]
tab_figure["qval_nonsignificant"] = [
    "" if y else str(x)
    for x, y in zip(tab_figure["qval"].values, idx)
]
tab_figure.sort_values("log2fc").to_csv(dir_tables+'sfig6d_cd8.csv', sep=";")

de_genes_cd8_responsive_all = tab.loc[idx, "gene"].values
direction_de_genes_cd8_responsive_all = -((tab.loc[idx, "log2fc"].values > 0)-0.5)*2.

# %% [code]
de_genes_cd8_responsive_all

# %% [code]
adata_proc_p1.obs["score_cd8_reactive_condition"] = (np.asarray(
    (
        adata_proc_p1[:, de_genes_cd8_responsive_all].X - 
        adata_proc_p1[:, de_genes_cd8_responsive_all].X.mean(axis=0)
    ) / adata_proc_p1[:, de_genes_cd8_responsive_all].X.todense().std(axis=0)
)*direction_de_genes_cd8_responsive_all).mean(axis=1).flatten()
sb.catplot(
    data=adata_proc_p1.obs, 
    y="score_cd8_reactive_condition", 
    hue="condition", 
    x="reactive_bool", 
    col="cell_type",
    kind="violin",
    split=True,
    ax=ax
)
plt.tight_layout()
plt.savefig(dir_panels + "violinsplit_p1_pbmc_score_cd8_reactive_condition.pdf")

# %% [markdown]
### Unstimulated vs stimulated in non-reactive

# %% [code]
det = de.test.t_test(
    adata_proc_p1[
        np.logical_and(
            adata_proc_p1.obs["cell_type"].values == "Cd4",
            np.array([
                xx not in clonotypes_p1_cd4_selected_final 
                for xx in adata_proc_p1.obs["clonotype"].values
            ]),
        ),
        [x not in tcr_genes_p1 for x in adata_proc_p1.var_names]
    ],
    grouping="condition", 
    is_logged=True
)
tab = det.summary()
print(np.unique(det.grouping))
tab["log2fc["+str(np.unique(det.grouping)[1])+"]"] = tab["log2fc"].values
de_genes_p1_cd4_unresponsive = tab.loc[tab["qval"].values < 0.01, "gene"].values
et = de.enrich.test(
    ref=rs,
    det=det,
    threshold=0.01,
)
significant_set_ids_p1_cd4_unresponsive = et.significant_set_ids()
print(len(de_genes_p1_cd4_unresponsive))
idx = np.logical_and(
    tab["qval"].values < 0.05,
    np.logical_and(
        tab["mean"].values >= np.log(0.1),
        np.abs(tab["log2fc"].values) >= np.log(1.1)
    )
)
tab.loc[idx, :].sort_values("log2fc").to_csv(dir_tables+'tab_de_p1_cd4_nonreactive_condition.csv', sep=";")
tab.sort_values("log2fc").to_csv(dir_tables+'tab_de_p1_cd4_nonreactive_condition_all.csv', sep=";")

de_genes_cd4_unresponsive_all = tab.loc[idx, "gene"].values
direction_de_genes_cd4_unresponsive_all = -((tab.loc[idx, "log2fc"].values > 0)-0.5)*2.

# %% [code]
de_genes_cd4_unresponsive_all

# %% [code]
adata_proc_p1.obs["score_cd4_nonreactive_condition"] = (np.asarray(
    (
        adata_proc_p1[:, de_genes_cd4_unresponsive_all].X - 
        adata_proc_p1[:, de_genes_cd4_unresponsive_all].X.mean(axis=0)
    ) / adata_proc_p1[:, de_genes_cd4_unresponsive_all].X.todense().std(axis=0)
)*direction_de_genes_cd4_unresponsive_all).mean(axis=1).flatten()
sb.catplot(
    data=adata_proc_p1.obs, 
    y="score_cd4_nonreactive_condition", 
    hue="condition", 
    x="reactive_bool", 
    col="cell_type",
    kind="violin",
    split=True,
    ax=ax
)
plt.tight_layout()
plt.savefig(dir_panels + "violinsplit_p1_pbmc_score_cd4_nonreactive_condition.pdf")

# %% [code]
det = de.test.t_test(
    adata_proc_p1[
        np.logical_and(
            adata_proc_p1.obs["cell_type"].values == "Cd8",
            np.array([
                xx not in clonotypes_p1_cd8_selected_final 
                for xx in adata_proc_p1.obs["clonotype"].values
            ]),
        ),
        [x not in tcr_genes_p1 for x in adata_proc_p1.var_names]
    ],
    grouping="condition", 
    is_logged=True
)
tab = det.summary()
print(np.unique(det.grouping))
tab["log2fc["+str(np.unique(det.grouping)[1])+"]"] = tab["log2fc"].values
de_genes_p1_cd8_unresponsive = tab.loc[tab["qval"].values < 0.01, "gene"].values
et = de.enrich.test(
    ref=rs,
    det=det,
    threshold=0.01,
)
significant_set_ids_p1_cd8_unresponsive = et.significant_set_ids()
print(len(de_genes_p1_cd8_unresponsive))
idx = np.logical_and(
    tab["qval"].values < 0.05,
    np.logical_and(
        tab["mean"].values >= np.log(0.1),
        np.abs(tab["log2fc"].values) >= np.log(1.1)
    )
)
tab.loc[idx, :].sort_values("log2fc").to_csv(dir_tables+'tab_de_p1_cd8_nonreactive_condition.csv', sep=";")
tab.sort_values("log2fc").to_csv(dir_tables+'tab_de_p1_cd8_nonreactive_condition_all.csv', sep=";")

de_genes_cd8_unresponsive_all = tab.loc[idx, "gene"].values
direction_de_genes_cd8_unresponsive_all = -((tab.loc[idx, "log2fc"].values > 0)-0.5)*2.

# %% [code]
de_genes_cd8_unresponsive_all

# %% [code]
adata_proc_p1.obs["score_cd8_nonreactive_condition"] = (np.asarray(
    (
        adata_proc_p1[:, de_genes_cd8_unresponsive_all].X - 
        adata_proc_p1[:, de_genes_cd8_unresponsive_all].X.mean(axis=0)
    ) / adata_proc_p1[:, de_genes_cd8_unresponsive_all].X.todense().std(axis=0)
)*direction_de_genes_cd8_unresponsive_all).mean(axis=1).flatten()
sb.catplot(
    data=adata_proc_p1.obs, 
    y="score_cd8_nonreactive_condition", 
    hue="condition", 
    x="reactive_bool", 
    col="cell_type",
    kind="violin",
    split=True,
    ax=ax
)
plt.tight_layout()
plt.savefig(dir_panels + "violinsplit_p1_pbmc_score_cd8_nonreactive_condition.pdf")

# %% [markdown]
### Overlap DE gene sets

# %% [code]
import matplotlib_venn

# %% [code]
matplotlib_venn.venn2([
        set(list(de_genes_unstim_cd4_responsive_all)),
        set(list(de_genes_cd4_responsive_all))
], set_labels = ['unstim\n(reactive\n-nonreactive)', 'reactive\n(unstim\n-stim)'], set_colors = ['grey', 'red'])
plt.tight_layout()
plt.savefig(dir_panels+"venn_de_genes_cd4", format="pdf")
pd.DataFrame({
    "genes": list(set(list(de_genes_unstim_cd4_responsive_all)).
                  intersection(set(list(de_genes_cd4_responsive_all))))
}).to_csv(dir_tables + "tab_set_cd4_unstim.reactive-nonreactive_reactive.unstim-stim_intersection.csv", sep=";")
pd.DataFrame({
    "genes": list(set(list(de_genes_unstim_cd4_responsive_all)).
                  difference(set(list(de_genes_cd4_responsive_all))))
}).to_csv(dir_tables + "tab_set_cd4_unstim.reactive-nonreactive_reactive.unstim-stim_1only.csv", sep=";")
pd.DataFrame({
    "genes": list(set(list(de_genes_cd4_responsive_all)).
                  difference(set(list(de_genes_unstim_cd4_responsive_all))))
}).to_csv(dir_tables + "tab_set_cd4_unstim.reactive-nonreactive_reactive.unstim-stim_2only.csv", sep=";")

# %% [code]
matplotlib_venn.venn2([
        set(list(de_genes_unstim_cd8_responsive_all)),
        set(list(de_genes_cd8_responsive_all))
], set_labels = ['unstim\n(reactive-\nnonreactive)', 'reactive\n(unstim-\nstim)'], set_colors = ['grey', 'red'])
plt.tight_layout()
plt.savefig(dir_panels+"venn_de_genes_cd8", format="pdf")
pd.DataFrame({
    "genes": list(set(list(de_genes_unstim_cd8_responsive_all)).
                  intersection(set(list(de_genes_cd8_responsive_all))))
}).to_csv(dir_tables + "tab_set_cd8_unstim.reactive-nonreactive_reactive.unstim-stim_intersection.csv", sep=";")
pd.DataFrame({
    "genes": list(set(list(de_genes_unstim_cd8_responsive_all)).
                  difference(set(list(de_genes_cd8_responsive_all))))
}).to_csv(dir_tables + "tab_set_cd8_unstim.reactive-nonreactive_reactive.unstim-stim_1only.csv", sep=";")
pd.DataFrame({
    "genes": list(set(list(de_genes_cd8_responsive_all)).
                  difference(set(list(de_genes_unstim_cd8_responsive_all))))
}).to_csv(dir_tables + "tab_set_cd8_unstim.reactive-nonreactive_reactive.unstim-stim_2only.csv", sep=";")

# %% [code]
matplotlib_venn.venn2([
        set(list(de_genes_unstim_cd4_responsive_all)),
        set(list(de_genes_stim_cd4_responsive_all))
], set_labels = ['unstim\n(reactive\n-nonreactive)', 'stim\n(reactive\n-nonreactive)'], set_colors = ['grey', 'red'])
plt.tight_layout()
plt.savefig(dir_panels+"venn_de_cd4_genes_reactive_nonreactive", format="pdf")
plt.savefig(dir_panels+"sfig8e_cd4", format="pdf")
plt.show()

pd.DataFrame({
    "genes": list(set(list(de_genes_unstim_cd4_responsive_all)).
                  intersection(set(list(de_genes_stim_cd4_responsive_all))))
}).to_csv(dir_tables + "sfig8e_cd4_unstim.reactive-nonreactive_stim.reactive-nonreactive_intersection.csv", sep=";")
pd.DataFrame({
    "genes": list(set(list(de_genes_unstim_cd4_responsive_all)).
                  difference(set(list(de_genes_stim_cd4_responsive_all))))
}).to_csv(dir_tables + "sfig8e_cd4_unstim.reactive-nonreactive_stim.reactive-nonreactive_1only.csv", sep=";")
pd.DataFrame({
    "genes": list(set(list(de_genes_stim_cd4_responsive_all)).
                  difference(set(list(de_genes_unstim_cd4_responsive_all))))
}).to_csv(dir_tables + "sfig8e_cd4_unstim.reactive-nonreactive_stim.reactive-nonreactive_2only.csv", sep=";")

pd.DataFrame({
    "genes": list(set(list(de_genes_unstim_cd4_responsive_all)).
                  intersection(set(list(de_genes_stim_cd4_responsive_all))))
}).to_csv(dir_tables + "tab_set_cd4_unstim.reactive-nonreactive_stim.reactive-nonreactive_intersection.csv", sep=";")
pd.DataFrame({
    "genes": list(set(list(de_genes_unstim_cd4_responsive_all)).
                  difference(set(list(de_genes_stim_cd4_responsive_all))))
}).to_csv(dir_tables + "tab_set_cd4_unstim.reactive-nonreactive_stim.reactive-nonreactive_1only.csv", sep=";")
pd.DataFrame({
    "genes": list(set(list(de_genes_stim_cd4_responsive_all)).
                  difference(set(list(de_genes_unstim_cd4_responsive_all))))
}).to_csv(dir_tables + "tab_set_cd4_unstim.reactive-nonreactive_stim.reactive-nonreactive_2only.csv", sep=";")

# %% [code]
matplotlib_venn.venn2([
        set(list(de_genes_unstim_cd8_responsive_all)),
        set(list(de_genes_stim_cd8_responsive_all))
], set_labels = ['unstim\n(reactive\n-nonreactive)', 'stim\n(reactive\n-nonreactive)'], set_colors = ['grey', 'red'])
plt.tight_layout()
plt.savefig(dir_panels+"venn_de_cd8_genes_reactive_nonreactive", format="pdf")
plt.savefig(dir_panels+"sfig8f_cd8", format="pdf")
plt.show()

pd.DataFrame({
    "genes": list(set(list(de_genes_unstim_cd8_responsive_all)).
                  intersection(set(list(de_genes_stim_cd8_responsive_all))))
}).to_csv(dir_tables + "sfig8f_cd8_unstim.reactive-nonreactive_stim.reactive-nonreactive_intersection.csv", sep=";")
pd.DataFrame({
    "genes": list(set(list(de_genes_unstim_cd8_responsive_all)).
                  difference(set(list(de_genes_stim_cd8_responsive_all))))
}).to_csv(dir_tables + "sfig8f_cd8_unstim.reactive-nonreactive_stim.reactive-nonreactive_1only.csv", sep=";")
pd.DataFrame({
    "genes": list(set(list(de_genes_stim_cd8_responsive_all)).
                  difference(set(list(de_genes_unstim_cd8_responsive_all))))
}).to_csv(dir_tables + "sfig8f_cd8_unstim.reactive-nonreactive_stim.reactive-nonreactive_2only.csv", sep=";")

print("GZMB in unstim: %s, in stim: %s" % (
    "GZMB" in de_genes_unstim_cd8_responsive_all,
    "GZMB" in de_genes_stim_cd8_responsive_all,
))
print("CCL5 in unstim: %s, in stim: %s" % (
    "CCL5" in de_genes_unstim_cd8_responsive_all,
    "CCL5" in de_genes_stim_cd8_responsive_all,
))
print("CXCR4 in unstim: %s, in stim: %s" % (
    "CXCR4" in de_genes_unstim_cd8_responsive_all,
    "CXCR4" in de_genes_stim_cd8_responsive_all,
))

pd.DataFrame({
    "genes": list(set(list(de_genes_unstim_cd8_responsive_all)).
                  intersection(set(list(de_genes_stim_cd8_responsive_all))))
}).to_csv(dir_tables + "tab_set_cd8_unstim.reactive-nonreactive_stim.reactive-nonreactive_intersection.csv", sep=";")
pd.DataFrame({
    "genes": list(set(list(de_genes_unstim_cd8_responsive_all)).
                  difference(set(list(de_genes_stim_cd8_responsive_all))))
}).to_csv(dir_tables + "tab_set_cd8_unstim.reactive-nonreactive_stim.reactive-nonreactive_1only.csv", sep=";")
pd.DataFrame({
    "genes": list(set(list(de_genes_stim_cd8_responsive_all)).
                  difference(set(list(de_genes_unstim_cd8_responsive_all))))
}).to_csv(dir_tables + "tab_set_cd8_unstim.reactive-nonreactive_stim.reactive-nonreactive_2only.csv", sep=";")

# %% [code]
matplotlib_venn.venn2([
        set(list(de_genes_cd4_unresponsive_all)),
        set(list(de_genes_cd4_responsive_all))
], set_labels = ['non-reactive\n(unstim\n-stim)', 'reactive\n(unstim\n-stim)'], set_colors = ['grey', 'red'])
plt.tight_layout()
plt.savefig(dir_panels+"venn_de_genes_cd4", format="pdf")
pd.DataFrame({
    "genes": list(set(list(de_genes_cd4_unresponsive_all)).
                  intersection(set(list(de_genes_cd4_responsive_all))))
}).to_csv(dir_tables + "tab_set_cd4_nonreactive.unstim-stim_reactive.unstim-stim_intersection.csv", sep=";")
pd.DataFrame({
    "genes": list(set(list(de_genes_cd4_unresponsive_all)).
                  difference(set(list(de_genes_cd4_responsive_all))))
}).to_csv(dir_tables + "tab_set_cd4_nonreactive.unstim-stim_reactive.unstim-stim_1only.csv", sep=";")
pd.DataFrame({
    "genes": list(set(list(de_genes_cd4_responsive_all)).
                  difference(set(list(de_genes_cd4_unresponsive_all))))
}).to_csv(dir_tables + "tab_set_cd4_nonreactive.unstim-stim_reactive.unstim-stim_2only.csv", sep=";")

# %% [markdown]
### Compute gene-set based scores

# %% [code]
cell_type = "Cd4"
score = {
    "score_cd4_nonreactive_stimulation": de_genes_cd4_unresponsive_all,
    "score_cd4_reactive_stimulation": de_genes_cd4_responsive_all,
    "score_cd4_unstimulated_reactiveness": de_genes_unstim_cd4_responsive_all,
    "score_cd4_stimulated_reactiveness": de_genes_stim_cd4_responsive_all,
}
idx = adata_proc_p1.obs["cell_type"].values == cell_type
for k, v in score.items():
    atemp = adata_proc_p1[idx, v]
    atemp.obs[k] = np.asarray(
        (
            atemp.X - 
            atemp.X.mean(axis=0)
        ) / atemp.X.todense().std(axis=0)
    ).mean(axis=1).flatten()
    adata_proc_p1.obs[k] = [
        atemp.obs.loc[x, k] if x in atemp.obs_names else np.nan for x in adata_proc_p1.obs_names
    ]
fig, ax = plt.subplots(nrows=len(score.keys()), ncols=1, figsize=(4, 4*len(score.keys())))
for i, (k, v) in enumerate(score.items()):
    sc.pl.umap(adata_proc_p1, ax=ax[i], show=False)
    sc.pl.umap(adata_proc_p1, color=k, ax=ax[i], show=False)
plt.tight_layout()
plt.savefig(dir_panels+ "umap_p1_pbmc_cd4_score_pairwise.pdf")
plt.show()

# %% [code]
cell_type = "Cd8"
score = {
    "score_cd8_nonreactive_stimulation": de_genes_cd8_unresponsive_all,
    "score_cd8_reactive_stimulation": de_genes_cd8_responsive_all,
    "score_cd8_unstimulated_reactiveness": de_genes_unstim_cd8_responsive_all,
    "score_cd8_stimulated_reactiveness": de_genes_stim_cd8_responsive_all,
}
idx = adata_proc_p1.obs["cell_type"].values == cell_type
for k, v in score.items():
    atemp = adata_proc_p1[idx, v]
    atemp.obs[k] = np.asarray(
        (
            atemp.X - 
            atemp.X.mean(axis=0)
        ) / atemp.X.todense().std(axis=0)
    ).mean(axis=1).flatten()
    adata_proc_p1.obs[k] = [
        atemp.obs.loc[x, k] if x in atemp.obs_names else np.nan for x in adata_proc_p1.obs_names
    ]
fig, ax = plt.subplots(nrows=len(score.keys()), ncols=1, figsize=(4, 4*len(score.keys())))
for i, (k, v) in enumerate(score.items()):
    sc.pl.umap(adata_proc_p1, ax=ax[i], show=False)
    sc.pl.umap(adata_proc_p1, color=k, ax=ax[i], show=False)
plt.tight_layout()
plt.savefig(dir_panels+ "umap_p1_pbmc_cd8_score_pairwise.pdf")
plt.show()

# %% [code]
def compute_score(adata, celltype, genes):
    genes = [x for x in genes if not isinstance(x, str) and not np.isnan(x)]
    a = adata_proc_p1[adata_proc_p1.obs["cell_type"].values == celltype].copy()
    sc.tl.score_genes(
        adata=a, 
        gene_list=de_genes_cd4_unresponsive_all, 
        ctrl_size=50, 
        n_bins=25, 
        score_name="score", 
        random_state=0, 
        use_raw=False
    )
    return [
        a.obs.loc[x, "score"] if x in a.obs_names else np.nan 
        for x in adata_proc_p1.obs_names
    ]

# %% [markdown]
does not work well

# %% [markdown]
### Overlay on TA

# %% [code]
sc.pl.umap(adata_p1_withta, color=["cell_type", "leiden_all_ta"])

# %% [code]
adata_p1_withta.obs.groupby(["cell_type", "identifier"]).size()

# %% [markdown]
#### Per patient

# %% [code]
cell_type = "Cd4"
score = {
    "score_cd4_nonreactive_stimulation": de_genes_cd4_unresponsive_all,
    "score_cd4_reactive_stimulation": de_genes_cd4_responsive_all,
    "score_cd4_unstimulated_reactiveness": de_genes_unstim_cd4_responsive_all,
    "score_cd4_stimulated_reactiveness": de_genes_stim_cd4_responsive_all,
}
partitions = ["Muc_10x_GT_5", "Muc_10x_GH_7"] #np.unique(adata_p1_withta.obs["identifier"].values)
for k, v in score.items():
    atemps = []
    for partition in partitions:
        idx = np.where(np.logical_and(
            adata_p1_withta.obs["cell_type"].values == cell_type,
            adata_p1_withta.obs["identifier"].values == partition
        ))[0]
        atemp = adata_p1_withta[idx, v].copy()
        atemp.obs[k] = np.asarray(
            (atemp.layers["lognormalised"] - atemp.layers["lognormalised"].mean(axis=0)) / 
            (atemp.layers["lognormalised"].std(axis=0) + 1e-8)
        ).mean(axis=1).flatten()
        atemps.append(atemp)
    atemps = atemps[0].concatenate(atemps[1:])
    atemps.obs_names = ["-".join(x.split("-")[:-1]) for x in atemps.obs_names]
    adata_p1_withta.obs[k] = [
        atemps.obs.loc[x, k] if x in atemps.obs_names else np.nan for x in adata_p1_withta.obs_names
    ]
fig, ax = plt.subplots(
    nrows=len(score.keys()), 
    ncols=len(partitions), 
    figsize=(4*len(partitions), 4*len(score.keys()))
)
for j, partition in enumerate(partitions):
    for i, (k, v) in enumerate(score.items()):
        sc.pl.umap(adata_p1_withta, ax=ax[i, j], size=20, show=False)
        sc.pl.umap(adata_p1_withta[np.logical_and(
            adata_p1_withta.obs["cell_type"].values == cell_type,
            adata_p1_withta.obs["identifier"].values == partition
        )], color=k, ax=ax[i, j], size=20, show=False)
        if i == 0:
            ax[i, j].set_title(partition)
        else:
            ax[i, j].set_title("")
        if j == 0:
            ax[i, j].set_ylabel(k+"\nUMAP2")
        else:
            ax[i, j].set_ylabel("")
        if i < len(score.items())-1:
            ax[i, j].set_xlabel("")
        else:
            ax[i, j].set_xlabel("UMAP1")
plt.tight_layout()
plt.savefig(dir_panels+ "umap_p1_pbmcta_cd4_score_pair.pdf")
plt.show()

fig, ax = plt.subplots(
    nrows=len(score.items()), 
    ncols=1, 
    figsize=(4, 4*len(score.items()))
)
for i, (k, v) in enumerate(score.items()):
    sb.boxplot(
        data=adata_p1_withta[np.logical_and(
            np.logical_and(
                adata_p1_withta.obs["cell_type"].values == cell_type,
                adata_p1_withta.obs["data_set"].values == "TA",
            ),
            [x in partitions for x in adata_p1_withta.obs["identifier"].values]
        )].obs,
        x="leiden_all_ta",
        #hue="identifier",
        y=k,
        ax=ax[i]
    )
    ax[i].set_title("")
    if i < len(score.items())-1:
        ax[i].set_xlabel("")
    else:
        ax[i].set_xlabel("cluster")
plt.tight_layout()
plt.show()

# %% [code]
cell_type = "Cd4"
separate_scoring = True
print(adata_p1_withta.obs.groupby(["cell_type", "data_set"]).size())
score = {
    "score_cd4_nonreactive_stimulation": (de_genes_cd4_unresponsive_all, direction_de_genes_cd4_unresponsive_all),
    "score_cd4_reactive_stimulation": (de_genes_cd4_responsive_all, direction_de_genes_cd4_responsive_all),
    "score_cd4_unstimulated_reactiveness": (de_genes_unstim_cd4_responsive_all, direction_de_genes_unstim_cd4_responsive_all),
    "score_cd4_stimulated_reactiveness": (de_genes_stim_cd4_responsive_all, direction_de_genes_stim_cd4_responsive_all),
}
fig, ax = plt.subplots(
    nrows=4, 
    ncols=1, 
    figsize=(4, 4*4)
)
for i, (k, v) in enumerate(score.items()):
    atemps = []
    for x in ["TA", "PBMC"]:
        if x == "TA":
            idx = np.where(np.logical_and(
                adata_p1_withta.obs["cell_type"].values == cell_type,
                adata_p1_withta.obs["data_set"].values == x
            ))[0]
            atemp = adata_p1_withta[
                idx, 
                v[0]
            ].copy()
            atemp.obs[k] = (np.asarray(
                (atemp.layers["lognormalised"] - atemp.layers["lognormalised"].mean(axis=0)) / 
                (atemp.layers["lognormalised"].std(axis=0) + 1e-8)) * v[1]
            ).mean(axis=1).flatten()
            atemp.X = atemp.layers["lognormalised"]
            atemp.obs["reactive"] = "TA"
        else:
            idx = np.where(np.logical_and(
                adata_proc_p1.obs["cell_type"].values == cell_type,
                [
                    x in ["reactive", "nonreactive"] 
                    for x in adata_proc_p1.obs["reactive"].values
                ]
            ))[0]
            atemp = adata_proc_p1[
                idx, 
                v[0]
            ].copy()
            atemp.obs[k] = (np.asarray(
                (atemp.X - atemp.X.mean(axis=0)) / 
                (atemp.X.todense().std(axis=0) + 1e-8)) * v[1]
            ).mean(axis=1).flatten()
            atemp.obs["data_set"] = "PBMC"
            atemp.obs["reactive"] = atemp.obs["reactive"]
        atemps.append(atemp)
    atemps = atemps[0].concatenate([atemps[1]])
    atemps.obs_names = ["-".join(x.split("-")[:-1]) for x in atemps.obs_names]
    if not separate_scoring:
        atemps.obs[k] = (np.asarray(
            (atemps.X - atemps.X.mean(axis=0)) / 
            (atemps.X.todense().std(axis=0) + 1e-8)) * v[1]
        ).mean(axis=1).flatten()
    sb.violinplot(
        data=atemps.obs,
        x="data_set",
        hue="reactive",
        y=k,
        ax=ax[i]
    )
plt.tight_layout()
plt.savefig(dir_panels+ "violin_p1_pbmcta_cd4_score_pair.pdf")
plt.show()

# %% [code]
cell_type = "Cd8"
score = {
    "score_cd8_nonreactive_stimulation": de_genes_cd8_unresponsive_all,
    "score_cd8_reactive_stimulation": de_genes_cd8_responsive_all,
    "score_cd8_unstimulated_reactiveness": de_genes_unstim_cd8_responsive_all,
    "score_cd8_stimulated_reactiveness": de_genes_stim_cd8_responsive_all,
}
partitions = ["Muc_10x_GT_5", "Muc_10x_GH_7", "muc17887"] #np.unique(adata_p1_withta.obs["identifier"].values)
for k, v in score.items():
    atemps = []
    for partition in partitions:
        idx = np.where(np.logical_and(
            adata_p1_withta.obs["cell_type"].values == cell_type,
            adata_p1_withta.obs["identifier"].values == partition
        ))[0]
        atemp = adata_p1_withta[idx, v].copy()
        atemp.obs[k] = np.asarray(
            (atemp.layers["lognormalised"] - atemp.layers["lognormalised"].mean(axis=0)) / 
            (atemp.layers["lognormalised"].std(axis=0) + 1e-8)
        ).mean(axis=1).flatten()
        atemps.append(atemp)
    atemps = atemps[0].concatenate(atemps[1:])
    atemps.obs_names = ["-".join(x.split("-")[:-1]) for x in atemps.obs_names]
    adata_p1_withta.obs[k] = [
        atemps.obs.loc[x, k] if x in atemps.obs_names else np.nan for x in adata_p1_withta.obs_names
    ]
fig, ax = plt.subplots(
    nrows=len(score.keys()), 
    ncols=len(partitions), 
    figsize=(4*len(partitions), 4*len(score.keys()))
)
for j, partition in enumerate(partitions):
    for i, (k, v) in enumerate(score.items()):
        sc.pl.umap(adata_p1_withta, ax=ax[i, j], size=20, show=False)
        sc.pl.umap(adata_p1_withta[np.logical_and(
            adata_p1_withta.obs["cell_type"].values == cell_type,
            adata_p1_withta.obs["identifier"].values == partition
        )], color=k, ax=ax[i, j], size=20, show=False)
        if i == 0:
            ax[i, j].set_title(partition)
        else:
            ax[i, j].set_title("")
        if j == 0:
            ax[i, j].set_ylabel(k+"\nUMAP2")
        else:
            ax[i, j].set_ylabel("")
        if i < len(score.items())-1:
            ax[i, j].set_xlabel("")
        else:
            ax[i, j].set_xlabel("UMAP1")
plt.tight_layout()
plt.savefig(dir_panels+ "umap_p1_pbmcta_cd8_score_pairwise.pdf")
plt.show()

fig, ax = plt.subplots(
    nrows=len(score.items()), 
    ncols=1, 
    figsize=(4, 4*len(score.items()))
)
for i, (k, v) in enumerate(score.items()):
    sb.boxplot(
        data=adata_p1_withta[np.logical_and(
            np.logical_and(
                adata_p1_withta.obs["cell_type"].values == cell_type,
                adata_p1_withta.obs["data_set"].values == "TA",
            ),
            [x in partitions for x in adata_p1_withta.obs["identifier"].values]
        )].obs,
        x="leiden_all_ta",
        #hue="identifier",
        y=k,
        ax=ax[i]
    )
    ax[i].set_title("")
    if i < len(score.items())-1:
        ax[i].set_xlabel("")
    else:
        ax[i].set_xlabel("cluster")
plt.tight_layout()
plt.show()

# %% [code]
cell_type = "Cd8"
separate_scoring = True
print(adata_p1_withta.obs.groupby(["cell_type", "data_set"]).size())
score = {
    "score_cd8_nonreactive_stimulation": (de_genes_cd8_unresponsive_all, direction_de_genes_cd8_unresponsive_all),
    "score_cd8_reactive_stimulation": (de_genes_cd8_responsive_all, direction_de_genes_cd8_responsive_all),
    "score_cd8_unstimulated_reactiveness": (de_genes_unstim_cd8_responsive_all, direction_de_genes_unstim_cd8_responsive_all),
    "score_cd8_stimulated_reactiveness": (de_genes_stim_cd8_responsive_all, direction_de_genes_stim_cd8_responsive_all),
}
fig, ax = plt.subplots(
    nrows=4, 
    ncols=1, 
    figsize=(4, 4*4)
)
for i, (k, v) in enumerate(score.items()):
    atemps = []
    for x in ["TA", "PBMC"]:
        if x == "TA":
            idx = np.where(np.logical_and(
                adata_p1_withta.obs["cell_type"].values == cell_type,
                adata_p1_withta.obs["data_set"].values == x
            ))[0]
            atemp = adata_p1_withta[
                idx, 
                v[0]
            ].copy()
            atemp.obs[k] = (np.asarray(
                (atemp.layers["lognormalised"] - atemp.layers["lognormalised"].mean(axis=0)) / 
                (atemp.layers["lognormalised"].std(axis=0) + 1e-8)) * v[1]
            ).mean(axis=1).flatten()
            atemp.X = atemp.layers["lognormalised"]
            atemp.obs["reactive"] = "TA"
        else:
            idx = np.where(np.logical_and(
                adata_proc_p1.obs["cell_type"].values == cell_type,
                [
                    x in ["reactive", "nonreactive"] 
                    for x in adata_proc_p1.obs["reactive"].values
                ]
            ))[0]
            atemp = adata_proc_p1[
                idx, 
                v[0]
            ].copy()
            atemp.obs[k] = (np.asarray(
                (atemp.X - atemp.X.mean(axis=0)) / 
                (atemp.X.todense().std(axis=0) + 1e-8)) * v[1]
            ).mean(axis=1).flatten()
            atemp.obs["data_set"] = "PBMC"
            atemp.obs["reactive"] = atemp.obs["reactive"]
        atemps.append(atemp)
    atemps = atemps[0].concatenate([atemps[1]])
    atemps.obs_names = ["-".join(x.split("-")[:-1]) for x in atemps.obs_names]
    if not separate_scoring:
        atemps.obs[k] = (np.asarray(
            (atemps.X - atemps.X.mean(axis=0)) / 
            (atemps.X.todense().std(axis=0) + 1e-8)) * v[1]
        ).mean(axis=1).flatten()
    sb.violinplot(
        data=atemps.obs,
        x="data_set",
        hue="reactive",
        y=k,
        ax=ax[i]
    )
plt.tight_layout()
plt.savefig(dir_panels+ "violin_p1_pbmcta_cd8_score_pair.pdf")
plt.show()

# %% [markdown]
#### Global

# %% [code]
adata_p1_withta.obs["data_set_sfig9a"] = pd.Categorical([
    "TA (same patient)" if x == "TA" and z in ['Muc_10x_GT_6', 'muc17888']
    else "TA" if x == "TA"
    else "PBMC (stim)" if y == "stimulated"
    else "PBMC (unstim)"
    for x, y, z in zip(
        adata_p1_withta.obs["data_set"].values,
        adata_p1_withta.obs["condition"].values,
        adata_p1_withta.obs["identifier"].values,
    )
], categories = ["TA", "TA (same patient)", "PBMC (stim)", "PBMC (unstim)"])
adata_p1_withta.uns["data_set_sfig9a_colors"] = ["purple", "pink", "#1f77b4", "#ff7f0e"]

adata_p1_withta.obs["data_set_sfig9d"] = pd.Categorical([
    "TA" if x == "TA"
    else "PBMC stimulated" if y == "stimulated"
    else "PBMC unstimulated"
    for x, y, z in zip(
        adata_p1_withta.obs["data_set"].values,
        adata_p1_withta.obs["condition"].values,
        adata_p1_withta.obs["identifier"].values,
    )
], categories = ["TA", "PBMC stimulated", "PBMC unstimulated"])

# %% [code]
fig, ax = plt.subplots(
    nrows=1, 
    ncols=3, 
    figsize=(4*3, 4)
)
sc.pl.umap(adata_p1_withta, color="data_set_sfig9a", ax=ax[0], size=20, show=False)
sc.pl.umap(adata_p1_withta, color="cell_type", ax=ax[1], size=20, show=False)

sc.pl.umap(adata_p1_withta, color="T_cells", ax=ax[1], alpha=0.5, size=20, show=False)
sc.pl.umap(adata_p1_withta[
    adata_p1_withta.obs["data_set"].values == 'PBMC'
], color="T_cells", ax=ax[1], size=20, show=False)

sc.pl.umap(adata_p1_withta, ax=ax[2], size=20, show=False)
sc.pl.umap(adata_p1_withta[np.logical_and(
    adata_p1_withta.obs["reactive"].values != 'un-reactive',
    adata_p1_withta.obs["condition"].values == 'stimulated'
)], color="T_cells", ax=ax[2], size=20, show=False)
sc.pl.umap(adata_p1_withta[np.logical_and(
    adata_p1_withta.obs["reactive"].values != 'un-reactive',
    adata_p1_withta.obs["condition"].values == 'unstimulated'
)], color="T_cells", alpha=0.4, ax=ax[2], size=40, show=False)

ax[0].legend_.remove()
ax[1].legend_.remove()
ax[2].legend_.remove()
ax[1].set_ylabel("")
ax[2].set_ylabel("")

ax[0].set_title("")
ax[1].set_title("")
ax[2].set_title("")
plt.tight_layout()
plt.savefig(dir_panels+ "sfig9a.pdf")
plt.show()

# %% [code]
partitions = ["PBMC unstimulated", "PBMC stimulated", "TA"]
score = ["IFNG", "Proliferation score", "Cytokine score"]
fig, ax = plt.subplots(
    nrows=len(score), 
    ncols=len(partitions), 
    figsize=(4*len(partitions), 4*len(score))
)
for i, k in enumerate(score):
    for j, kk in enumerate(partitions):
        sc.pl.umap(adata_p1_withta, ax=ax[i, j], size=20, show=False)
        sc.pl.umap(
            adata_p1_withta[adata_p1_withta.obs["data_set_sfig9d"].values == kk], 
            color=k, 
            ax=ax[i, j], 
            size=20, 
            show=False,
            cmap="viridis"
        )
        if i == 0:
            ax[i, j].set_title(kk)
        else:
            ax[i, j].set_title("")
        if j == 0:
            ax[i, j].set_ylabel(k)
        else:
            ax[i, j].set_ylabel("")
        ax[i, j].set_xlabel("")
plt.tight_layout()
plt.savefig(dir_panels+"sfig9d.pdf")
plt.show()

fig, ax = plt.subplots(
    nrows=len(score), 
    ncols=len(partitions), 
    figsize=(4*len(partitions), 4*len(score))
)
for i, k in enumerate(score):
    for j, kk in enumerate(partitions):
        sc.pl.umap(adata_p1_withta, ax=ax[i, j], size=20, show=False)
        sc.pl.umap(
            adata_p1_withta[adata_p1_withta.obs["data_set_sfig9d"].values == kk], 
            color=k, 
            vmin=np.min(adata_p1_withta.obs[k].values) if k in adata_p1_withta.obs.columns else np.min(adata_p1_withta[:, k].X.todense()),
            vmax=np.max(adata_p1_withta.obs[k].values) if k in adata_p1_withta.obs.columns else np.max(adata_p1_withta[:, k].X.todense()),
            ax=ax[i, j], 
            size=20, 
            show=False,
            cmap="viridis"
        )
        if i == 0:
            ax[i, j].set_title(kk)
        else:
            ax[i, j].set_title("")
        if j == 0:
            ax[i, j].set_ylabel(k)
        else:
            ax[i, j].set_ylabel("")
        ax[i, j].set_xlabel("")
plt.tight_layout()
plt.savefig(dir_panels+"sfig9d_scaled.pdf")
plt.show()

# %% [code]
cell_type = "Cd4"
score = {
    #"score_cd4_nonreactive_stimulation": de_genes_cd4_unresponsive_all,
    #"score_cd4_reactive_stimulation": de_genes_cd4_responsive_all,
    "score_cd4_unstimulated_reactiveness": de_genes_unstim_cd4_responsive_all,
    "score_cd4_stimulated_reactiveness": de_genes_stim_cd4_responsive_all,
}
partitions = ["TA", "PBMC"]
print(adata_p1_withta.obs.groupby(["data_set", "cell_type"]).size())
for k, v in score.items():
    atemps = []
    for partition in partitions:
        idx = np.where(np.logical_and(
            adata_p1_withta.obs["cell_type"].values == cell_type,
            adata_p1_withta.obs["data_set"].values == partition
        ))[0]
        atemp = adata_p1_withta[idx, v].copy()
        if partition == "TA":
            atemp.obs[k] = np.asarray(
                (atemp.layers["lognormalised"] - atemp.layers["lognormalised"].mean(axis=0)) / 
                (atemp.layers["lognormalised"].std(axis=0) + 1e-8)
            ).mean(axis=1).flatten()
        else:
            atemp.obs[k] = np.asarray(
                (atemp.X - atemp.X.mean(axis=0)) / 
                (atemp.X.todense().std(axis=0) + 1e-8)
            ).mean(axis=1).flatten()
        atemps.append(atemp)
    atemps = atemps[0].concatenate(atemps[1:])
    atemps.obs_names = ["-".join(x.split("-")[:-1]) for x in atemps.obs_names]
    adata_p1_withta.obs[k] = [
        atemps.obs.loc[x, k] if x in atemps.obs_names else np.nan for x in adata_p1_withta.obs_names
    ]
partitions = ["TA", "PBMC"]
fig, ax = plt.subplots(
    nrows=1, 
    ncols=len(score.keys()), 
    figsize=(4*len(score.keys()), 4)
)
for i, (k, v) in enumerate(score.items()):
    sc.pl.umap(adata_p1_withta, ax=ax[i], size=20, show=False)
    sc.pl.umap(
        adata_p1_withta[adata_p1_withta.obs["cell_type"].values == cell_type], 
        color=k, ax=ax[i], size=20, show=False, cmap="Blues"
    )
    ax[i].set_title(k)
    ax[i].set_ylabel("")
    ax[i].set_xlabel("")
ax[0].set_ylabel("")
plt.tight_layout()
plt.savefig(dir_panels+"sfig9b.pdf")
plt.show()

# %% [code]
cell_type = "Cd8"
score = {
    #"score_cd8_nonreactive_stimulation": de_genes_cd8_unresponsive_all,
    #"score_cd8_reactive_stimulation": de_genes_cd8_responsive_all,
    "score_cd8_unstimulated_reactiveness": de_genes_unstim_cd8_responsive_all,
    "score_cd8_stimulated_reactiveness": de_genes_stim_cd8_responsive_all,
}
partitions = ["TA", "PBMC"]
print(adata_p1_withta.obs.groupby(["data_set", "cell_type"]).size())
for k, v in score.items():
    atemps = []
    for partition in partitions:
        idx = np.where(np.logical_and(
            adata_p1_withta.obs["cell_type"].values == cell_type,
            adata_p1_withta.obs["data_set"].values == partition
        ))[0]
        atemp = adata_p1_withta[idx, v].copy()
        if partition == "TA":
            atemp.obs[k] = np.asarray(
                (atemp.layers["lognormalised"] - atemp.layers["lognormalised"].mean(axis=0)) / 
                (atemp.layers["lognormalised"].std(axis=0) + 1e-8)
            ).mean(axis=1).flatten()
        else:
            atemp.obs[k] = np.asarray(
                (atemp.X - atemp.X.mean(axis=0)) / 
                (atemp.X.todense().std(axis=0) + 1e-8)
            ).mean(axis=1).flatten()
        atemps.append(atemp)
    atemps = atemps[0].concatenate(atemps[1:])
    atemps.obs_names = ["-".join(x.split("-")[:-1]) for x in atemps.obs_names]
    adata_p1_withta.obs[k] = [
        atemps.obs.loc[x, k] if x in atemps.obs_names else np.nan for x in adata_p1_withta.obs_names
    ]
partitions = ["TA", "PBMC"]
fig, ax = plt.subplots(
    nrows=1, 
    ncols=len(score.keys()), 
    figsize=(4*len(score.keys()), 4)
)
for i, (k, v) in enumerate(score.items()):
    sc.pl.umap(adata_p1_withta, ax=ax[i], size=20, show=False)
    sc.pl.umap(
        adata_p1_withta[adata_p1_withta.obs["cell_type"].values == cell_type], 
        color=k, ax=ax[i], size=20, show=False, cmap="Blues"
    )
    ax[i].set_title(k)
    ax[i].set_ylabel("")
    ax[i].set_xlabel("")
ax[0].set_ylabel("")
plt.tight_layout()
plt.savefig(dir_panels+"sfig9c.pdf")
plt.show()

# %% [code]
idx_genes = [x for x in residence_score_gene_list if x in adata_p1_withta.var_names]
pbmc_mean = adata_p1_withta[
    adata_p1_withta.obs["data_set"].values == "PBMC", 
    idx_genes
].X.mean(axis=0)
pbmc_std = np.maximum(adata_p1_withta[
    adata_p1_withta.obs["data_set"].values == "PBMC", 
    idx_genes
].X.todense().std(axis=0), 1e-8)

ta_mean = adata_p1_withta[
    adata_p1_withta.obs["data_set"].values == "TA", 
    idx_genes
].X.mean(axis=0)
ta_std = np.maximum(adata_p1_withta[
    adata_p1_withta.obs["data_set"].values == "TA", 
    idx_genes
].X.todense().std(axis=0), 1e-8)

means = np.concatenate([
    pbmc_mean if y == "PBMC"
    else ta_mean
    for i, y in enumerate(adata_p1_withta.obs["data_set"].values)
], axis=0)
stds = np.concatenate([
    pbmc_std if y == "PBMC"
    else ta_std
    for i, y in enumerate(adata_p1_withta.obs["data_set"].values)
], axis=0)

adata_p1_withta.obs["tissue_residency_score"] = np.asarray(np.mean(
    (adata_p1_withta[:, idx_genes].X - means) / stds, 
    axis=1
)).flatten()

# %% [code]
fig, ax = plt.subplots(1,1,figsize=(12, 4))
sc.pl.violin(adata_p1_withta, groupby="reactive_combined", keys="tissue_residency_score", ax=ax, show=False)
ax.set_xticklabels(ax.get_xticklabels(), rotation=90)
plt.savefig(dir_panels+"sfig9f.pdf")
plt.show()

# %% [code]
partitions = ["PBMC unstimulated", "PBMC stimulated", "TA"]
score = ["tissue_residency_score"]
fig, ax = plt.subplots(
    nrows=len(score), 
    ncols=len(partitions), 
    figsize=(4*len(partitions), 4*len(score))
)
for j, k in enumerate(score):
    for i, kk in enumerate(partitions):
        sc.pl.umap(adata_p1_withta, ax=ax[i], size=20, show=False)
        sc.pl.umap(
            adata_p1_withta[adata_p1_withta.obs["data_set_sfig9d"].values == kk], 
            color=k, 
            ax=ax[i], 
            size=20, 
            show=False,
            cmap="viridis"
        )
        if j == 0:
            ax[i].set_title(kk)
        else:
            ax[i].set_title("")
        if i == 0:
            ax[i].set_ylabel(k)
        else:
            ax[i].set_ylabel("")
        ax[i].set_xlabel("")
plt.tight_layout()
plt.savefig(dir_panels+"sfig9e.pdf")
plt.show()

# %% [markdown]
## Marker genes

# %% [markdown]
### Cd4

# %% [code]
atemp = adata_proc_p1[
    np.logical_and(
        adata_proc_p1.obs["cell_type"].values == "Cd4",
        [x in ["reactive", "nonreactive"] for x in adata_proc_p1.obs["reactive"]]
    ),
    [x not in tcr_genes_p1 for x in adata_proc_p1.var_names]
].copy()
atemp.obs["reactive_condition"] = pd.Categorical(
    atemp.obs["reactive_condition"].values,
    categories=np.sort(np.unique(atemp.obs["reactive_condition"].values))
)
print(atemp.obs.groupby("reactive_condition").size())
sc.tl.rank_genes_groups(
    atemp,
    groupby="reactive_condition", 
    use_raw=False, 
    log=True, 
    groups='all', 
    reference='rest',
    method="wilcoxon",
    key_added='rank_genes_groups',
)

# %% [code]
sc.tl.filter_rank_genes_groups(
    atemp, 
    key='rank_genes_groups', 
    groupby=None, 
    use_raw=False, 
    log=True, 
    key_added='rank_genes_groups_filtered', 
    min_in_group_fraction=.1, 
    min_fold_change=1.2, 
    max_out_group_fraction=1.
)

# %% [code]
sc.pl.rank_genes_groups_dotplot(
    atemp, 
    key='rank_genes_groups_filtered',
    use_raw=False
)

# %% [code]
if False:  # does not work
    cell_type = "Cd4"
    score = {
        "score_cd4_marker_stimulated_nonreactive": atemp.uns['rank_genes_groups_filtered']["names"]["stimulated_nonreactive"].tolist(),
        "score_cd4_marker_stimulated_reactive": atemp.uns['rank_genes_groups_filtered']["names"]["stimulated_reactive"].tolist(),
        "score_cd4_marker_unstimulated_nonreactive": atemp.uns['rank_genes_groups_filtered']["names"]["unstimulated_nonreactive"].tolist(),
        "score_cd4_marker_unstimulated_reactive": atemp.uns['rank_genes_groups_filtered']["names"]["unstimulated_reactive"].tolist(),
    }
    for k, v in score.items():
        adata_proc_p1.obs[k] = compute_score(
            adata=adata_proc_p1, 
            celltype=cell_type, 
            genes=v
        )
    fig, ax = plt.subplots(nrows=len(score.keys()), ncols=2, figsize=(8, 4*len(score.keys())))
    for i, (k, v) in enumerate(score.items()):
        sc.pl.umap(adata_proc_p1, ax=ax[i, 0], show=False)
        sc.pl.umap(adata_proc_p1, color=k, ax=ax[i, 0], show=False)
        sc.pl.umap(adata_proc_p1, ax=ax[i, 1], show=False)
        sc.pl.umap(adata_proc_p1[adata_proc_p1.obs["cell_type"].values == cell_type], color="condition", ax=ax[i, 1], show=False)
    plt.tight_layout()
    plt.show()

# %% [markdown]
### Cd8

# %% [code]
atemp = adata_proc_p1[
    np.logical_and(
        adata_proc_p1.obs["cell_type"].values == "Cd8",
        [x in ["reactive", "nonreactive"] for x in adata_proc_p1.obs["reactive"]]
    ),
    [x not in tcr_genes_p1 for x in adata_proc_p1.var_names]
].copy()
atemp.obs["reactive_condition"] = pd.Categorical(
    atemp.obs["reactive_condition"].values,
    categories=np.sort(np.unique(atemp.obs["reactive_condition"].values))
)
print(atemp.obs.groupby("reactive_condition").size())
sc.tl.rank_genes_groups(
    atemp,
    groupby="reactive_condition", 
    use_raw=False, 
    log=True, 
    groups='all', 
    reference='rest',
    method="t-test_overestim_var",
    key_added='rank_genes_groups',
)

# %% [code]
sc.tl.filter_rank_genes_groups(
    atemp, 
    key='rank_genes_groups', 
    groupby=None, 
    use_raw=False, 
    log=True, 
    key_added='rank_genes_groups_filtered', 
    min_in_group_fraction=0.1, 
    min_fold_change=1.2, 
    max_out_group_fraction=1.
)

# %% [code]
sc.pl.rank_genes_groups_dotplot(
    atemp, 
    key='rank_genes_groups_filtered',
    use_raw=False
)

# %% [markdown]
## Full linear model DE

# %% [markdown]
### Cd4

# %% [code]
atemp = adata_proc_p1[np.logical_and(
    adata_proc_p1.obs["cell_type"].values == "Cd4",
    [x in ["reactive", "nonreactive"] for x in adata_proc_p1.obs["reactive"]]
)].copy()
print(atemp.obs.groupby("reactive_condition").size())
atemp.obs["condition"] = pd.Categorical(
    atemp.obs["condition"].values, 
    categories=['unstimulated', 'stimulated']
)
atemp = atemp[:, atemp.var["n_cells"].values > 100].copy()
atemp.obs["sf"] = atemp.obs["n_counts"] / 10000
det_cd4 = de.test.wald(
    data=atemp.raw.X,
    sample_description=atemp.obs,
    gene_names=atemp.raw.var_names,
    size_factors="sf",
    formula_loc="~1+condition+reactive+condition:reactive",
    factor_loc_totest="reactive", 
)

# %% [code]
det_cd4.model_estim.input_data.loc_names

# %% [code]
tab_responsive = det_cd4.summary()
tab_stimulated = de.test.wald_repeated(
    det_cd4,
    coef_to_test='condition[T.stimulated]', 
).summary()
tab_stimulatedresponsive = de.test.wald_repeated(
    det_cd4,
    coef_to_test='condition[T.stimulated]:reactive[T.reactive]', 
).summary()

# %% [code]
mean_threshold = 0.1
tab_responsive_de = tab_responsive[np.logical_and(
    tab_responsive["qval"].values <= 0.01,
    np.logical_and(
        tab_responsive["mean"].values >= mean_threshold,
        tab_responsive["log2fc"].values >= np.log(1.5)
    )
)]
tab_stimulated_de = tab_stimulated[np.logical_and(
    tab_stimulated["qval"].values <= 0.01,
    np.logical_and(
        tab_stimulated["mean"].values >= mean_threshold,
        tab_stimulated["log2fc"].values >= np.log(1.5)
    )
)]
tab_stimulatedresponsive_de = tab_stimulatedresponsive[np.logical_and(
    tab_stimulatedresponsive["qval"].values <= 0.01,
    np.logical_and(
        tab_stimulatedresponsive["mean"].values >= mean_threshold,
        tab_stimulatedresponsive["log2fc"].values >= np.log(1.5)
    )
)]
tab_responsive_de.to_csv(dir_tables+'tab_set_p1_cd4_glm_reactive.csv', sep=";")
tab_stimulated_de.to_csv(dir_tables+'tab_set_p1_cd4_glm_stimulated.csv', sep=";")
tab_stimulatedresponsive_de.to_csv(dir_tables+'tab_set_p1_cd4_glm_stimulatedreactive.csv', sep=";")

# %% [code]
print(tab_responsive_de.shape)
print(tab_stimulated_de.shape)
print(tab_stimulatedresponsive_de.shape)

# %% [code]
matplotlib_venn.venn3([
    set(list(tab_responsive_de["gene"].values)),
    set(list(tab_stimulated_de["gene"].values)),
    set(list(tab_stimulatedresponsive_de["gene"].values))
], set_labels = [
    'reactive', 
    'stimulated',
    'reactive_stimulated'
], 
    set_colors = ['grey', 'orange', 'blue'])
plt.tight_layout()
#plt.savefig(dir_panels+"venn_de_genes_cd4", format="pdf")

# %% [code]
marker_genes_responsive = list(
    set(list(tab_responsive_de["gene"].values)).difference(
    set(list(tab_stimulated_de["gene"].values))).difference(
    set(list(tab_stimulatedresponsive_de["gene"].values))
))
marker_genes_stim = list(
    set(list(tab_stimulated_de["gene"].values)).difference(
    set(list(tab_responsive_de["gene"].values))).difference(
    set(list(tab_stimulatedresponsive_de["gene"].values))
))
marker_genes_responsive_stim = list(
    set(list(tab_stimulatedresponsive_de["gene"].values)).difference(
    set(list(tab_responsive_de["gene"].values))).difference(
    set(list(tab_stimulated_de["gene"].values))
))
print(len(marker_genes_responsive))
print(len(marker_genes_stim))
print(len(marker_genes_responsive_stim))

# %% [code]
cell_type = "Cd4"
score = {
    "score_cd4_glm_reactive": marker_genes_responsive,
    "score_cd4_glm_stimulated": marker_genes_stim,
    "score_cd4_glm_reactive_stimulated": marker_genes_responsive_stim,
}
idx = adata_proc_p1.obs["cell_type"].values == cell_type
for k, v in score.items():
    atemp = adata_proc_p1[idx, v]
    atemp.obs[k] = np.asarray(
        (
            atemp.X - 
            atemp.X.mean(axis=0)
        ) / atemp.X.todense().std(axis=0)
    ).mean(axis=1).flatten()
    adata_proc_p1.obs[k] = [
        atemp.obs.loc[x, k] if x in atemp.obs_names else np.nan for x in adata_proc_p1.obs_names
    ]
fig, ax = plt.subplots(nrows=len(score.keys()), ncols=1, figsize=(4, 4*len(score.keys())))
for i, (k, v) in enumerate(score.items()):
    sc.pl.umap(adata_proc_p1, ax=ax[i], show=False)
    sc.pl.umap(adata_proc_p1, color=k, ax=ax[i], show=False)
plt.tight_layout()
plt.savefig(dir_panels+ "umap_p1_pbmc_cd4_score_glm.pdf")
plt.show()

# %% [markdown]
### Cd8

# %% [code]
atemp = adata_proc_p1[np.logical_and(
    adata_proc_p1.obs["cell_type"].values == "Cd8",
    [x in ["reactive", "nonreactive"] for x in adata_proc_p1.obs["reactive"]]
)].copy()
print(atemp.obs.groupby("reactive_condition").size())
atemp.obs["condition"] = pd.Categorical(
    atemp.obs["condition"].values, 
    categories=['unstimulated', 'stimulated']
)
atemp = atemp[:, atemp.var["n_cells"].values > 100].copy()
atemp.obs["sf"] = atemp.obs["n_counts"] / 10000
det_cd8 = de.test.wald(
    data=atemp.raw.X,
    sample_description=atemp.obs,
    gene_names=atemp.raw.var_names,
    size_factors="sf",
    formula_loc="~1+condition+reactive+condition:reactive",
    factor_loc_totest="reactive", 
)

# %% [code]
det_cd8.model_estim.input_data.loc_names

# %% [code]
tab_responsive = det_cd8.summary()
tab_stimulated = de.test.wald_repeated(
    det_cd8,
    coef_to_test='condition[T.stimulated]', 
).summary()
tab_stimulatedresponsive = de.test.wald_repeated(
    det_cd8,
    coef_to_test='condition[T.stimulated]:reactive[T.reactive]', 
).summary()

# %% [code]
mean_threshold = 0.1
tab_responsive_de = tab_responsive[np.logical_and(
    tab_responsive["qval"].values <= 0.01,
    np.logical_and(
        tab_responsive["mean"].values >= mean_threshold,
        tab_responsive["log2fc"].values >= np.log(1.5)
    )
)]
tab_stimulated_de = tab_stimulated[np.logical_and(
    tab_stimulated["qval"].values <= 0.01,
    np.logical_and(
        tab_stimulated["mean"].values >= mean_threshold,
        tab_stimulated["log2fc"].values >= np.log(1.5)
    )
)]
tab_stimulatedresponsive_de = tab_stimulatedresponsive[np.logical_and(
    tab_stimulatedresponsive["qval"].values <= 0.01,
    np.logical_and(
        tab_stimulatedresponsive["mean"].values >= mean_threshold,
        tab_stimulatedresponsive["log2fc"].values >= np.log(1.5)
    )
)]
tab_responsive_de.to_csv(dir_tables+'tab_set_p1_cd8_glm_reactive.csv', sep=";")
tab_stimulated_de.to_csv(dir_tables+'tab_set_p1_cd8_glm_stimulated.csv', sep=";")
tab_stimulatedresponsive_de.to_csv(dir_tables+'tab_set_p1_cd8_glm_stimulatedreactive.csv', sep=";")

# %% [code]
print(tab_responsive_de.shape)
print(tab_stimulated_de.shape)
print(tab_stimulatedresponsive_de.shape)

# %% [code]
matplotlib_venn.venn3([
    set(list(tab_responsive_de["gene"].values)),
    set(list(tab_stimulated_de["gene"].values)),
    set(list(tab_stimulatedresponsive_de["gene"].values))
], set_labels = [
    'reactive', 
    'stimulated',
    'reactive_stimulated'
], 
    set_colors = ['grey', 'orange', 'blue'])
plt.tight_layout()
#plt.savefig(dir_panels+"venn_de_genes_cd4", format="pdf")

# %% [code]
marker_genes_responsive = list(
    set(list(tab_responsive_de["gene"].values)).difference(
    set(list(tab_stimulated_de["gene"].values))).difference(
    set(list(tab_stimulatedresponsive_de["gene"].values))
))
marker_genes_stim = list(
    set(list(tab_stimulated_de["gene"].values)).difference(
    set(list(tab_responsive_de["gene"].values))).difference(
    set(list(tab_stimulatedresponsive_de["gene"].values))
))
marker_genes_responsive_stim = list(
    set(list(tab_stimulatedresponsive_de["gene"].values)).difference(
    set(list(tab_responsive_de["gene"].values))).difference(
    set(list(tab_stimulated_de["gene"].values))
))
print(len(marker_genes_responsive))
print(len(marker_genes_stim))
print(len(marker_genes_responsive_stim))

# %% [code]
cell_type = "Cd8"
score = {
    "score_cd8_glm_reactive": marker_genes_responsive,
    "score_cd8_glm_stimulated": marker_genes_stim,
    "score_cd8_glm_reactive_stimulated": marker_genes_responsive_stim,
}
idx = adata_proc_p1.obs["cell_type"].values == cell_type
for k, v in score.items():
    atemp = adata_proc_p1[idx, v]
    atemp.obs[k] = np.asarray(
        (
            atemp.X - 
            atemp.X.mean(axis=0)
        ) / atemp.X.todense().std(axis=0)
    ).mean(axis=1).flatten()
    adata_proc_p1.obs[k] = [
        atemp.obs.loc[x, k] if x in atemp.obs_names else np.nan for x in adata_proc_p1.obs_names
    ]
fig, ax = plt.subplots(nrows=len(score.keys()), ncols=1, figsize=(4, 4*len(score.keys())))
for i, (k, v) in enumerate(score.items()):
    sc.pl.umap(adata_proc_p1, ax=ax[i], show=False)
    sc.pl.umap(adata_proc_p1, color=k, ax=ax[i], show=False)
plt.tight_layout()
plt.savefig(dir_panels+ "umap_p1_pbmc_cd8_score_glm.pdf")
plt.show()

# %% [markdown]
### Overlay on TA

# %% [code]
sc.pl.umap(adata_p1_withta, color=["cell_type", "leiden_all_ta"])

# %% [code]
adata_p1_withta.obs.groupby(["cell_type", "identifier"]).size()

# %% [code]
cell_type = "Cd4"
tab_responsive_de = pd.read_csv(dir_tables+'tab_set_p1_cd4_glm_reactive.csv', sep=";")
tab_stimulated_de = pd.read_csv(dir_tables+'tab_set_p1_cd4_glm_stimulated.csv', sep=";")
tab_stimulatedresponsive_de = pd.read_csv(dir_tables+'tab_set_p1_cd4_glm_stimulatedreactive.csv', sep=";")
score = {
    "score_cd4_glm_reactive": tab_responsive_de["gene"].values,
    "score_cd4_glm_stimulated": tab_stimulated_de["gene"].values,
    "score_cd4_glm_reactive_stimulated": tab_stimulatedresponsive_de["gene"].values,
}
partitions = ["Muc_10x_GT_5", "Muc_10x_GH_7"] #np.unique(adata_p1_withta.obs["identifier"].values)
for k, v in score.items():
    atemps = []
    for partition in partitions:
        idx = np.where(np.logical_and(
            adata_p1_withta.obs["cell_type"].values == cell_type,
            adata_p1_withta.obs["identifier"].values == partition
        ))[0]
        atemp = adata_p1_withta[idx, v].copy()
        atemp.obs[k] = np.asarray(
            (atemp.layers["lognormalised"] - atemp.layers["lognormalised"].mean(axis=0)) / 
            (atemp.layers["lognormalised"].std(axis=0) + 1e-8)
        ).mean(axis=1).flatten()
        atemps.append(atemp)
    atemps = atemps[0].concatenate(atemps[1:])
    atemps.obs_names = ["-".join(x.split("-")[:-1]) for x in atemps.obs_names]
    adata_p1_withta.obs[k] = [
        atemps.obs.loc[x, k] if x in atemps.obs_names else np.nan for x in adata_p1_withta.obs_names
    ]
fig, ax = plt.subplots(
    nrows=len(score.keys()), 
    ncols=len(partitions), 
    figsize=(4*len(partitions), 4*len(score.keys()))
)
for j, partition in enumerate(partitions):
    for i, (k, v) in enumerate(score.items()):
        sc.pl.umap(adata_p1_withta, ax=ax[i, j], size=20, show=False)
        sc.pl.umap(adata_p1_withta[np.logical_and(
            adata_p1_withta.obs["cell_type"].values == cell_type,
            adata_p1_withta.obs["identifier"].values == partition
        )], color=k, ax=ax[i, j], size=20, show=False)
        if i == 0:
            ax[i, j].set_title(partition)
        else:
            ax[i, j].set_title("")
        if j == 0:
            ax[i, j].set_ylabel(k+"\nUMAP2")
        else:
            ax[i, j].set_ylabel("")
        if i < len(score.items())-1:
            ax[i, j].set_xlabel("")
        else:
            ax[i, j].set_xlabel("UMAP1")
plt.tight_layout()
plt.savefig(dir_panels+ "umap_p1_pbmcta_cd4_score_glm.pdf")
plt.show()

fig, ax = plt.subplots(
    nrows=len(score.items()), 
    ncols=1, 
    figsize=(4, 4*len(score.items()))
)
for i, (k, v) in enumerate(score.items()):
    sb.boxplot(
        data=adata_p1_withta[np.logical_and(
            np.logical_and(
                adata_p1_withta.obs["cell_type"].values == cell_type,
                adata_p1_withta.obs["data_set"].values == "TA",
            ),
            [x in partitions for x in adata_p1_withta.obs["identifier"].values]
        )].obs,
        x="leiden_all_ta",
        #hue="identifier",
        y=k,
        ax=ax[i]
    )
    ax[i].set_title("")
    if i < len(score.items())-1:
        ax[i].set_xlabel("")
    else:
        ax[i].set_xlabel("cluster")
plt.tight_layout()
plt.show()

# %% [code]
tab = pd.read_csv(dir_tables+'tab_set_p1_cd8_glm_stimulatedreactive.csv', sep=";")
cell_type = "Cd4"
separate_scoring = True
print(adata_p1_withta.obs.groupby(["cell_type", "data_set"]).size())
tab_responsive_de = pd.read_csv(dir_tables+'tab_set_p1_cd4_glm_reactive.csv', sep=";")
tab_stimulated_de = pd.read_csv(dir_tables+'tab_set_p1_cd4_glm_stimulated.csv', sep=";")
tab_stimulatedresponsive_de = pd.read_csv(dir_tables+'tab_set_p1_cd4_glm_stimulatedreactive.csv', sep=";")
score = {
    "score_cd4_glm_reactive": tab_responsive_de["gene"].values,
    "score_cd4_glm_stimulated": tab_stimulated_de["gene"].values,
    "score_cd4_glm_reactive_stimulated": tab_stimulatedresponsive_de["gene"].values,
}
fig, ax = plt.subplots(
    nrows=3, 
    ncols=1, 
    figsize=(4, 4*3)
)
for i, (k, v) in enumerate(score.items()):
    atemps = []
    for x in ["TA", "PBMC"]:
        if x == "TA":
            idx = np.where(np.logical_and(
                adata_p1_withta.obs["cell_type"].values == cell_type,
                adata_p1_withta.obs["data_set"].values == x
            ))[0]
            atemp = adata_p1_withta[
                idx, 
                v
            ].copy()
            atemp.obs[k] = np.asarray(
                (atemp.layers["lognormalised"] - atemp.layers["lognormalised"].mean(axis=0)) / 
                (atemp.layers["lognormalised"].std(axis=0) + 1e-8)
            ).mean(axis=1).flatten()
            atemp.X = atemp.layers["lognormalised"]
            atemp.obs["reactive"] = "TA"
        else:
            idx = np.where(np.logical_and(
                adata_proc_p1.obs["cell_type"].values == cell_type,
                [
                    x in ["reactive", "nonreactive"] 
                    for x in adata_proc_p1.obs["reactive"].values
                ]
            ))[0]
            atemp = adata_proc_p1[
                idx, 
                v
            ].copy()
            atemp.obs[k] = np.asarray(
                (atemp.X - atemp.X.mean(axis=0)) / 
                (atemp.X.todense().std(axis=0) + 1e-8)
            ).mean(axis=1).flatten()
            atemp.obs["data_set"] = "PBMC"
            atemp.obs["reactive"] = atemp.obs["reactive"]
        atemps.append(atemp)
    atemps = atemps[0].concatenate([atemps[1]])
    atemps.obs_names = ["-".join(x.split("-")[:-1]) for x in atemps.obs_names]
    if not separate_scoring:
        atemps.obs[k] = np.asarray(
            (atemps.X - atemps.X.mean(axis=0)) / 
            (atemps.X.todense().std(axis=0) + 1e-8)
        ).mean(axis=1).flatten()
    sb.violinplot(
        data=atemps.obs,
        x="data_set",
        hue="reactive",
        y=k,
        ax=ax[i]
    )
plt.tight_layout()
plt.savefig(dir_panels+ "violin_p1_pbmcta_cd4_score_glm.pdf")
plt.show()

# %% [code]
cell_type = "Cd8"
tab_responsive_de = pd.read_csv(dir_tables+'tab_set_p1_cd8_glm_reactive.csv', sep=";")
tab_stimulated_de = pd.read_csv(dir_tables+'tab_set_p1_cd8_glm_stimulated.csv', sep=";")
tab_stimulatedresponsive_de = pd.read_csv(dir_tables+'tab_set_p1_cd8_glm_stimulatedreactive.csv', sep=";")
score = {
    "score_cd8_glm_reactive": tab_responsive_de["gene"].values,
    "score_cd8_glm_stimulated": tab_stimulated_de["gene"].values,
    "score_cd8_glm_reactive_stimulated": tab_stimulatedresponsive_de["gene"].values,
}
partitions = ["Muc_10x_GT_5", "Muc_10x_GH_7", "muc17887"] #np.unique(adata_p1_withta.obs["identifier"].values)
for k, v in score.items():
    atemps = []
    for partition in partitions:
        idx = np.where(np.logical_and(
            adata_p1_withta.obs["cell_type"].values == cell_type,
            adata_p1_withta.obs["identifier"].values == partition
        ))[0]
        atemp = adata_p1_withta[idx, v].copy()
        atemp.obs[k] = np.asarray(
            (atemp.layers["lognormalised"] - atemp.layers["lognormalised"].mean(axis=0)) / 
            (atemp.layers["lognormalised"].std(axis=0) + 1e-8)
        ).mean(axis=1).flatten()
        atemps.append(atemp)
    atemps = atemps[0].concatenate(atemps[1:])
    atemps.obs_names = ["-".join(x.split("-")[:-1]) for x in atemps.obs_names]
    adata_p1_withta.obs[k] = [
        atemps.obs.loc[x, k] if x in atemps.obs_names else np.nan for x in adata_p1_withta.obs_names
    ]
fig, ax = plt.subplots(
    nrows=len(score.keys()), 
    ncols=len(partitions), 
    figsize=(4*len(partitions), 4*len(score.keys()))
)
for j, partition in enumerate(partitions):
    for i, (k, v) in enumerate(score.items()):
        sc.pl.umap(adata_p1_withta, ax=ax[i, j], size=20, show=False)
        sc.pl.umap(adata_p1_withta[np.logical_and(
            adata_p1_withta.obs["cell_type"].values == cell_type,
            adata_p1_withta.obs["identifier"].values == partition
        )], color=k, ax=ax[i, j], size=20, show=False)
        if i == 0:
            ax[i, j].set_title(partition)
        else:
            ax[i, j].set_title("")
        if j == 0:
            ax[i, j].set_ylabel(k+"\nUMAP2")
        else:
            ax[i, j].set_ylabel("")
        if i < len(score.items())-1:
            ax[i, j].set_xlabel("")
        else:
            ax[i, j].set_xlabel("UMAP1")
plt.tight_layout()
plt.savefig(dir_panels+ "umap_p1_pbmcta_cd8_score_glm.pdf")
plt.show()

# %% [code]
fig, ax = plt.subplots(
    nrows=len(score.items()), 
    ncols=1, 
    figsize=(4, 4*len(score.items()))
)
for i, (k, v) in enumerate(score.items()):
    sb.boxplot(
        data=adata_p1_withta[np.logical_and(
            np.logical_and(
                adata_p1_withta.obs["cell_type"].values == cell_type,
                adata_p1_withta.obs["data_set"].values == "TA",
            ),
            [x in partitions for x in adata_p1_withta.obs["identifier"].values]
        )].obs,
        x="leiden_all_ta",
        #hue="identifier",
        y=k,
        ax=ax[i]
    )
    ax[i].set_title("")
    if i < len(score.items())-1:
        ax[i].set_xlabel("")
    else:
        ax[i].set_xlabel("cluster")
plt.tight_layout()
plt.show()

# %% [code]
tab = pd.read_csv(dir_tables+'tab_set_p1_cd8_glm_stimulatedreactive.csv', sep=";")
cell_type = "Cd8"
separate_scoring = True
print(adata_p1_withta.obs.groupby(["cell_type", "data_set"]).size())
tab_responsive_de = pd.read_csv(dir_tables+'tab_set_p1_cd8_glm_reactive.csv', sep=";")
tab_stimulated_de = pd.read_csv(dir_tables+'tab_set_p1_cd8_glm_stimulated.csv', sep=";")
tab_stimulatedresponsive_de = pd.read_csv(dir_tables+'tab_set_p1_cd8_glm_stimulatedreactive.csv', sep=";")
score = {
    "score_cd8_glm_reactive": tab_responsive_de["gene"].values,
    "score_cd8_glm_stimulated": tab_stimulated_de["gene"].values,
    "score_cd8_glm_reactive_stimulated": tab_stimulatedresponsive_de["gene"].values,
}
fig, ax = plt.subplots(
    nrows=3, 
    ncols=1, 
    figsize=(4, 4*3)
)
for i, (k, v) in enumerate(score.items()):
    atemps = []
    for x in ["TA", "PBMC"]:
        if x == "TA":
            idx = np.where(np.logical_and(
                adata_p1_withta.obs["cell_type"].values == cell_type,
                adata_p1_withta.obs["data_set"].values == x
            ))[0]
            atemp = adata_p1_withta[
                idx, 
                v
            ].copy()
            atemp.obs[k] = np.asarray(
                (atemp.layers["lognormalised"] - atemp.layers["lognormalised"].mean(axis=0)) / 
                (atemp.layers["lognormalised"].std(axis=0) + 1e-8)
            ).mean(axis=1).flatten()
            atemp.obs["reactive"] = "TA"
            atemp.X = atemp.layers["lognormalised"]
        else:
            idx = np.where(np.logical_and(
                adata_proc_p1.obs["cell_type"].values == cell_type,
                [
                    x in ["reactive", "nonreactive"] 
                    for x in adata_proc_p1.obs["reactive"].values
                ]
            ))[0]
            atemp = adata_proc_p1[
                idx, 
                v
            ].copy()
            atemp.obs[k] = np.asarray(
                (atemp.X - atemp.X.mean(axis=0)) / 
                (atemp.X.todense().std(axis=0) + 1e-8)
            ).mean(axis=1).flatten()
            atemp.obs["data_set"] = "PBMC"
            atemp.obs["reactive"] = atemp.obs["reactive"]
        atemps.append(atemp)
    atemps = atemps[0].concatenate([atemps[1]])
    atemps.obs_names = ["-".join(x.split("-")[:-1]) for x in atemps.obs_names]
    if not separate_scoring:
        atemps.obs[k] = np.asarray(
            (atemps.X - atemps.X.mean(axis=0)) / 
            (atemps.X.todense().std(axis=0) + 1e-8)
        ).mean(axis=1).flatten()
    sb.violinplot(
        data=atemps.obs,
        x="data_set",
        hue="reactive",
        y=k,
        ax=ax[i]
    )
plt.tight_layout()
plt.savefig(dir_panels+ "violin_p1_pbmcta_cd8_score_glm.pdf")
plt.show()

# %% [markdown]
## Similarity of selected clonotypes

# %% [markdown]
### Condition response

# %% [code]
for k, v in {"low":(10,10), "high": (28, 28)}.items():  #{"low":(20,20), "high": (100, 100)}.items():
    idx = np.array([
        y >= 3 and z >= 3 for y, z in zip(
            tab_clonotypes_p1_filtered["n_stimulated"].values,
            tab_clonotypes_p1_filtered["n_unstimulated"].values
        )
    ])
    clonotypes_temp = np.unique(tab_clonotypes_p1_filtered["clonotype_merged"].values[idx])
    idx = np.array([
        tab_clonotypes_p1_filtered["clonotype_merged"].values.tolist().index(x) 
        for x in clonotypes_temp
    ])  # select first occurrence only, entries in vecs_p1 are the same
    g = sb.clustermap(
        data=pd.DataFrame(
            np.corrcoef(
                x=np.concatenate([
                    np.concatenate([
                        np.expand_dims(vecs_p1[k], axis=0) 
                        for k in clonotypes_temp
                    ], axis=0)
                ], axis=0)
            ),
            index=\
                ["" for x in clonotypes_temp]
                if k == "low" else clonotypes_temp,
            columns=\
                ["" for x in clonotypes_temp]
                if k == "low" else clonotypes_temp,
        ),
        row_colors=pd.DataFrame({
            "Cd4": [
                ["white", "darkorange"][int(x == "Cd4")] for x in tab_clonotypes_p1_filtered["label_cell_type"].values[idx]
            ],
            "Cd8": [
                ["white", "blue"][int(x == "Cd8")] for x in tab_clonotypes_p1_filtered["label_cell_type"].values[idx]
            ],
            "in_27": [
                ["white", "lightcoral"][int(x > 0)] for x in tab_clonotypes_p1_filtered["in_ifng_cluster"].values[idx]
            ],
            "selected": [
                ["white", "darkred"][int(x in clonotypes_p1_selected)] for x in clonotypes_temp
            ],
        }, index=\
            ["" for x in clonotypes_temp]
            if k == "low" else clonotypes_temp
        ),
        colors_ratio=0.02,
        figsize=v,
    )
    plt.tight_layout()
    g.savefig(dir_panels + f"sfig5c_{k}.pdf")
    plt.close()

# %% [markdown]
### Sequence

# %% [code]
idx_clonotype_p1 = np.array([
    np.where(adata_proc_p1.obs["clonotype"] == x)[0][0]  # take first cell
    for x in clonotypes_p1_selected
])
g = sb.clustermap(
    data=pd.DataFrame(
        adata_proc_p1.uns['tcr_neighbors_aa_alignment']['distances'][idx_clonotype_p1, :][:, idx_clonotype_p1].todense(),
        index=clonotypes_p1_selected,
        columns=clonotypes_p1_selected,
    ),
    figsize=(10, 10),
)
g.savefig(dir_panels + "tcrdistance_heatmap_p1_all_selected_clonotypes.pdf")

# %% [markdown]
## Distribution of reactive clonotypes across clusters

# %% [markdown]
### P1

# %% [code]
adata_temp = adata_proc_p1[adata_proc_p1.obs["leiden_Cd4"].values != "nan"].copy()
adata_temp.obs["clonotype"] = [
    x if y == "reactive" else "others"
    for x, y in zip(
        adata_temp.obs["clonotype"].values,
        adata_temp.obs["reactive"].values
    )
]
tab = adata_temp.obs.groupby(["clonotype", "leiden_Cd4"]).size().reset_index().pivot(index='clonotype', columns='leiden_Cd4')
tab = pd.DataFrame(
    tab.values / np.expand_dims(tab.sum(axis=1).values, axis=1),
    columns=[x[1] for x in tab.columns],
    index=tab.index
)
fig, ax = plt.subplots(1,1,figsize=(8, 5))
sb.heatmap(tab, ax=ax, annot=False, cmap="viridis")
ax.set_xlabel("cluster")
ax.set_yticklabels(ax.get_yticklabels(), rotation=0)
plt.savefig(dir_panels + "sfig7d2_cd4.pdf")
plt.show()
tab.to_csv(dir_tables + "sfig7d2_cd4.csv", sep=";")
del adata_temp

# %% [code]
adata_temp = adata_proc_p1[adata_proc_p1.obs["leiden_Cd8"].values != "nan"].copy()
adata_temp.obs["clonotype_merged"] = [
    x if y == "reactive" else "others"
    for x, y in zip(
        adata_temp.obs["clonotype_merged"].values,
        adata_temp.obs["reactive"].values
    )
]
tab = adata_temp.obs.groupby(["clonotype_merged", "leiden_Cd8"]).size().reset_index().pivot(index='clonotype_merged', columns='leiden_Cd8')
tab = pd.DataFrame(
    tab.values / np.expand_dims(tab.sum(axis=1).values, axis=1),
    columns=[x[1] for x in tab.columns],
    index=tab.index
)
fig, ax = plt.subplots(1,1,figsize=(8, 5))
sb.heatmap(tab, ax=ax, annot=False, cmap="viridis")
ax.set_xlabel("cluster")
ax.set_yticklabels(ax.get_yticklabels(), rotation=0)
plt.savefig(dir_panels + "sfig7d1_cd8.pdf")
plt.show()
tab.to_csv(dir_tables + "sfig7d1_cd8.csv", sep=";")
del adata_temp

# %% [markdown]
## Marker positive cells in clonotypes

# %% [code]
gene_ids = ["PDCD1", "CXCR4"]
idx = np.where([
    x in clonotypes_p1_cd4_selected_final
    for x in adata_proc_p1.obs["clonotype"].values
])[0]
tab = adata_proc_p1[idx].obs.copy()
for x in gene_ids:
    tab[x] = np.asarray(adata_proc_p1[idx, x].X.todense()).flatten() > 0
tab = tab.groupby(["clonotype", "condition"])[gene_ids].mean().reset_index()
print(tab)
adata_temp = adata_proc_p1[idx].copy()
adata_temp.obs["clonotype_condition"] = [
    x + "_" + y
    for x, y in zip(
        adata_temp.obs["clonotype"].values,
        adata_temp.obs["condition"].values
    )
]
sc.pl.dotplot(
    adata_temp,
    var_names=gene_ids,
    groupby="clonotype_condition",
)
tab.to_csv(dir_tables + "fig3b1.csv", sep=";")

# %% [markdown]
## TBX21, GATA3 correlation

# %% [markdown]
### Discrete

# %% [code]
p_tbx21pos = (np.asarray(adata_proc_p1[adata_proc_p1.obs["cell_type"].values == "Cd4", "TBX21"].X.todense()).flatten() > 0).mean()
p_gata3pos = (np.asarray(adata_proc_p1[adata_proc_p1.obs["cell_type"].values == "Cd4", "GATA3"].X.todense()).flatten() > 0).mean()

# %% [code]
tab = pd.DataFrame({
    "TBX21+GATA3+": np.logical_and(
        np.asarray(adata_proc_p1[adata_proc_p1.obs["cell_type"].values == "Cd4", "TBX21"].X.todense()).flatten() > 0,
        np.asarray(adata_proc_p1[adata_proc_p1.obs["cell_type"].values == "Cd4", "GATA3"].X.todense()).flatten() > 0
    ),
    "TBX21+GATA3-": np.logical_and(
        np.asarray(adata_proc_p1[adata_proc_p1.obs["cell_type"].values == "Cd4", "TBX21"].X.todense()).flatten() > 0,
        np.asarray(adata_proc_p1[adata_proc_p1.obs["cell_type"].values == "Cd4", "GATA3"].X.todense()).flatten() == 0
    ),
    "TBX21-GATA3+": np.logical_and(
        np.asarray(adata_proc_p1[adata_proc_p1.obs["cell_type"].values == "Cd4", "TBX21"].X.todense()).flatten() == 0,
        np.asarray(adata_proc_p1[adata_proc_p1.obs["cell_type"].values == "Cd4", "GATA3"].X.todense()).flatten() > 0
    ),
    "TBX21-GATA3-": np.logical_and(
        np.asarray(adata_proc_p1[adata_proc_p1.obs["cell_type"].values == "Cd4", "TBX21"].X.todense()).flatten() == 0,
        np.asarray(adata_proc_p1[adata_proc_p1.obs["cell_type"].values == "Cd4", "GATA3"].X.todense()).flatten() == 0
    )
})
tab.mean().reset_index()

# %% [code]
tab_exptected = pd.DataFrame({
    "TBX21+GATA3+": p_tbx21pos * p_gata3pos,
    "TBX21+GATA3-": p_tbx21pos * (1.-p_gata3pos),
    "TBX21-GATA3+": (1.-p_tbx21pos) * p_gata3pos,
    "TBX21-GATA3-": (1.-p_tbx21pos) * (1.-p_gata3pos),
}, index=["expected"])
tab_exptected

# %% [markdown]
#### Ifng expression

# %% [code]
clonotypes = ["138", "19", "256", "573"]
idx = np.logical_and(
    np.array([x in clonotypes for x in adata_proc_p1.obs["clonotype_merged"].values]),
    adata_proc_p1.obs["cell_type"].values == "Cd4"
)
tab = pd.DataFrame({
    "group": pd.Categorical(np.array(["TBX21+GATA3+", "TBX21+GATA3-", "TBX21-GATA3+", "TBX21-GATA3-"])[np.where(np.vstack([
        np.logical_and(
            np.asarray(adata_proc_p1[idx, "TBX21"].X.todense()).flatten() > 0,
            np.asarray(adata_proc_p1[idx, "GATA3"].X.todense()).flatten() > 0
        ),
        np.logical_and(
            np.asarray(adata_proc_p1[idx, "TBX21"].X.todense()).flatten() > 0,
            np.asarray(adata_proc_p1[idx, "GATA3"].X.todense()).flatten() == 0
        ),
        np.logical_and(
            np.asarray(adata_proc_p1[idx, "TBX21"].X.todense()).flatten() == 0,
            np.asarray(adata_proc_p1[idx, "GATA3"].X.todense()).flatten() > 0
        ),
        np.logical_and(
            np.asarray(adata_proc_p1[idx, "TBX21"].X.todense()).flatten() == 0,
            np.asarray(adata_proc_p1[idx, "GATA3"].X.todense()).flatten() == 0
        ),
    ]))[0]]),
    "IFNG": np.asarray(adata_proc_p1[idx, "IFNG"].X.todense()).flatten(),
    "clonotype": adata_proc_p1[idx].obs["clonotype"].values,
    "condition": adata_proc_p1[idx].obs["condition"].values,
})
tab.to_csv(dir_tables + "fig3b2.csv", sep=";")

fig, axs = plt.subplots(2,2,figsize=(16, 8))
axs = axs.ravel()
for i, x in enumerate(clonotypes):
    sb.boxplot(data=tab.loc[tab["clonotype"].values == x, :], x="group", y="IFNG", hue="condition", ax=axs[i])
    axs[i].set_title(x)
plt.tight_layout()
plt.show()

# %% [markdown]
### Continuous

# %% [code]
tbx21 = np.asarray(adata_proc_p1[adata_proc_p1.obs["cell_type"].values == "Cd4", "TBX21"].X.todense()).flatten()
gata3 = np.asarray(adata_proc_p1[adata_proc_p1.obs["cell_type"].values == "Cd4", "GATA3"].X.todense()).flatten()
x = np.vstack([
    tbx21,
    gata3
]).T
x.shape

# %% [code]
from sklearn.covariance import LedoitWolf, OAS, ShrunkCovariance, \
    log_likelihood, empirical_covariance
from sklearn.model_selection import GridSearchCV

# %% [code]
lw = LedoitWolf().fit(x)
oa = OAS().fit(x)

# %% [code]
print(lw.covariance_)
print(oa.covariance_)

# %% [markdown]
# Save objects

# %% [code]
if bool_recomp:
    for k in ["repertoire_overlap", "clonotype_imbalance"]:  # cannot save
        if k in adata_proc_p1.uns.keys():
            del adata_proc_p1.uns[k]
        if k in adata_proc_p2.uns.keys():
            del adata_proc_p2.uns[k]
    sc.write(dir_anndata+'adata_proc_p1.h5ad', adata_proc_p1)
    sc.write(dir_anndata+'adata_proc_p2.h5ad', adata_proc_p2)
```
