## Extended Data File 1 for "Single-cell RNA sequencing reveals *in vivo* signatures of SARS-CoV-2-reactive T cells through ‘reverse phenotyping’": 2_pseudotime_analysis.ipynb

```
# %% [markdown]
# Load packages

# %% [code]
import anndata
import diffxpy.api as de
import numpy as np
import scanpy as sc
import scirpy as ir
import scipy.stats
import sys
import pandas as pd
import seaborn as sb
import matplotlib.pyplot as plt
from matplotlib import rcParams
from matplotlib import colors

np.random.seed(seed=1)

# %% [code]
import warnings
warnings.filterwarnings('ignore')

# %% [code]
sc.settings.verbosity = 3 # amount of output
pd.options.display.max_columns = 999

# Put your name here and comment other out.
user = "david"

# Directories on your machine with backslash at the end.
if user == "david_server":
    data_dir_vdj = '/storage/scratch/users/xavier.pastor/projects/20200727_Covid19/data/10x_vdj/cellranger_vdj/'
    data_dir_rna = '/storage/groups/bcf/projects/SingleCell/10x/projects/schiller/200724_Covid19_stimulated/cellranger/'
    dir_out = '/storage/groups/ml01/workspace/david.fischer/covid19tcr/results/'
    dir_data = '/storage/groups/ml01/workspace/david.fischer/covid19tcr/data/'
    dir_data_liao = '/storage/groups/ml01/workspace/david.fischer/covid19tcr/data/GSE145926_RAW/'
    dir_data_lee = '/storage/groups/ml01/workspace/david.fischer/covid19tcr/data/2020_Lee_CovidFlu/'
elif user == "david":
    data_dir_vdj = None
    data_dir_rna = '/Users/david.fischer/phd/data/covidtcr/data/ta/cellranger/'
    dir_out = '/Users/david.fischer/phd/data/covidtcr/results/'
    dir_data = '/Users/david.fischer/phd/data/covidtcr/data/'
    dir_data_liao = '/Users/david.fischer/phd/datasets/2020_Liao_SARSCOV2_TCR/GSE145926_RAW/'
    dir_data_lee = '/Users/david.fischer/phd/datasets/2020_Lee_CovidFlu/'

# Make sure that these folders exist:
dir_panels = dir_out + 'panels/'
dir_tables = dir_out + 'tables/'
dir_anndata = dir_out + 'anndata/'
dir_genesets = dir_data + "genesets/"
dir_reference = dir_data + "reference/"

sc.logging.print_versions()
sc.settings.set_figure_params(dpi=80, scanpy=True)
sc.settings.figdir = dir_panels
print(sys.version)

# %% [code]
bool_recomp = True

# %% [markdown]
# Read data

# %% [markdown]
## Processed PBMCs

# %% [code]
adata_proc_p1 = sc.read_h5ad(dir_anndata+"adata_proc_p1.h5ad")

# %% [markdown]
## Clonotype tables

# %% [code]
tab_clonotypes_p1_filtered_cd4 = pd.read_csv(dir_tables+'tab_clonotypes_p1_cd4_filtered.csv', sep=";", index_col=0, )
tab_clonotypes_p1_filtered_cd8 = pd.read_csv(dir_tables+'tab_clonotypes_p1_cd8_filtered.csv', sep=";", index_col=0)
tab_clonotypes_p1_filtered_cd4.index = [str(x) for x in tab_clonotypes_p1_filtered_cd4.index]
tab_clonotypes_p1_filtered_cd8.index = [str(x) for x in tab_clonotypes_p1_filtered_cd8.index]
tab_clonotypes_p1_filtered_cd4["clonotype"] = [str(x) for x in tab_clonotypes_p1_filtered_cd4["clonotype"].values]
tab_clonotypes_p1_filtered_cd8["clonotype"] = [str(x) for x in tab_clonotypes_p1_filtered_cd8["clonotype"].values]
tab_clonotypes_p1_filtered = pd.concat([
    tab_clonotypes_p1_filtered_cd4, 
    tab_clonotypes_p1_filtered_cd8
], axis=0)

# %% [markdown]
# Pseudotime analysis of response

# %% [code]
sc.pl.umap(adata_proc_p1, color="ifng_pos_cluster")

# %% [markdown]
## Cd4 cells

# %% [markdown]
### Stimulated only

# %% [code]
sc.pl.umap(adata_proc_p1, color="proliferation_score")

# %% [markdown]
#### Proliferation-maximum centred pseudotime

# %% [code]
adata_temp = adata_proc_p1[np.logical_and(
    adata_proc_p1.obs["cell_type"].values == "Cd4",
    adata_proc_p1.obs["condition"].values == "stimulated"
)].copy()
sc.pp.pca(adata_temp)
sc.pp.neighbors(adata_temp, n_neighbors=100)
adata_temp.uns['iroot'] = np.where(adata_temp.obs["leiden_subclustered"].values == "subclustered_2")[0][
    np.argmax(adata_temp[adata_temp.obs["leiden_subclustered"].values == "subclustered_2"].obsm['X_umap'][:,1])
]
sc.tl.diffmap(adata_temp)
sc.tl.dpt(adata_temp)
adata_proc_p1.obs["dpt1_cd4_stim"] = [
    1. - adata_temp[x].obs["dpt_pseudotime"].values[0]
    if x in adata_temp.obs_names else np.nan
    for x in adata_proc_p1.obs_names
]

# %% [code]
fig, ax = plt.subplots(1,1)
ax = sc.pl.umap(adata_proc_p1, ax=ax, show=False)
ax = sc.pl.umap(adata_proc_p1, color="dpt1_cd4_stim", ax=ax, show=False)
plt.savefig(dir_panels + "umap_pseudotime1_cd4_stim.pdf")
plt.show()

sb.distplot(adata_proc_p1.obs["dpt1_cd4_stim"].values)
fig, ax = plt.subplots(1,1)
sb.scatterplot(
    x=adata_proc_p1.obs["dpt1_cd4_stim"].values,
    y=adata_proc_p1.obs["proliferation_score"].values,
    ax=ax
)
ax.set_xlabel("pseudotime score")
ax.set_ylabel("proliferation_score")
plt.savefig(dir_panels + "scatter_pseudotime1_cd4_stim_proliferation_score.pdf")
plt.show()

# %% [code]
tab = adata_proc_p1.obs.loc[np.logical_not(np.isnan(adata_proc_p1.obs["dpt1_cd4_stim"].values)),:].copy()
tab["IFNG"] = np.asarray(adata_proc_p1[np.logical_not(np.isnan(adata_proc_p1.obs["dpt1_cd4_stim"].values)), "IFNG"].X.todense()).flatten()
tab = tab.loc[[
    (tab_clonotypes_p1_filtered.loc[x, "n_stimulated"] >= 3 and \
    tab_clonotypes_p1_filtered.loc[x, "n_unstimulated"] >= 3)
    if x in tab_clonotypes_p1_filtered.index else False
    for x in tab["clonotype_merged"].values
]].copy()
tab["clonotype_merged"] = pd.Categorical(
    tab["clonotype_merged"].values,
    categories=np.unique(tab["clonotype_merged"].values)
)
tab["clonotype_merged"] = pd.Categorical(
    tab["clonotype_merged"].values,
    categories=tab.groupby("clonotype_merged")["dpt1_cd4_stim"].mean().reset_index().\
        sort_values("dpt1_cd4_stim")["clonotype_merged"].values[:].tolist()
)
tab = tab.sort_values("clonotype_merged")

cmap = plt.get_cmap("viridis")
my_pal = dict([
    (x, y[:-1])
    for x, y in zip(
        tab.groupby("clonotype_merged")["dpt1_cd4_stim"].mean().reset_index()["clonotype_merged"],
        cmap(tab.groupby("clonotype_merged")["dpt1_cd4_stim"].mean().reset_index()["dpt1_cd4_stim"])
    )
])

fig, ax = plt.subplots(nrows=2, ncols=1, figsize=(15, 10))
sb.boxplot(
    data=tab,
    x="clonotype_merged",
    y="proliferation_score",
    palette=my_pal,
    ax=ax[0]
)
ax[0].set_xticklabels(ax[0].get_xticklabels(), rotation=90)
sb.boxplot(
    data=tab,
    x="clonotype_merged",
    y="IFNG",
    palette=my_pal,
    ax=ax[1]
)
ax[1].set_xticklabels(ax[1].get_xticklabels(), rotation=90)
plt.show()

# %% [markdown]
#### Cytokine-maximum centred pseudotime

# %% [code]
adata_temp = adata_proc_p1[np.logical_and(
    adata_proc_p1.obs["cell_type"].values == "Cd4",
    adata_proc_p1.obs["condition"].values == "stimulated"
)].copy()
sc.pp.pca(adata_temp)
sc.pp.neighbors(adata_temp, n_neighbors=100)
adata_temp.uns['iroot'] = np.argmax(adata_temp.obsm['X_umap'][:,1])
sc.tl.diffmap(adata_temp)
sc.tl.dpt(adata_temp)
adata_proc_p1.obs["dpt2_cd4_stim"] = [
    1. - adata_temp[x].obs["dpt_pseudotime"].values[0]
    if x in adata_temp.obs_names else np.nan
    for x in adata_proc_p1.obs_names
]

# %% [code]
fig, ax = plt.subplots(1,1)
ax = sc.pl.umap(adata_proc_p1, ax=ax, show=False)
ax = sc.pl.umap(adata_proc_p1, color="dpt2_cd4_stim", ax=ax, show=False)
plt.savefig(dir_panels + "fig1d1.pdf")
plt.show()

sb.distplot(adata_proc_p1.obs["dpt2_cd4_stim"].values)
sb.scatterplot(
    x=adata_proc_p1.obs["dpt2_cd4_stim"].values,
    y=adata_proc_p1.obs["proliferation_score"].values,
    ax=ax
)
ax.set_xlabel("pseudotime score")
ax.set_ylabel("proliferation_score")
plt.show()

# %% [code]
tab = adata_proc_p1.obs.loc[np.logical_not(np.isnan(adata_proc_p1.obs["dpt2_cd4_stim"].values)),:].copy()
tab["IFNG"] = np.asarray(adata_proc_p1[np.logical_not(np.isnan(adata_proc_p1.obs["dpt2_cd4_stim"].values)), "IFNG"].X.todense()).flatten()
tab = tab.loc[[
    (tab_clonotypes_p1_filtered.loc[x, "n_stimulated"] >= 1 and \
    tab_clonotypes_p1_filtered.loc[x, "n_unstimulated"] >= 1)
    if x in tab_clonotypes_p1_filtered.index else False
    for x in tab["clonotype"].values
]].copy()
tab["clonotype"] = pd.Categorical(
    tab["clonotype"].values,
    categories=np.unique(tab["clonotype"].values)
)
tab["clonotype"] = pd.Categorical(
    tab["clonotype"].values,
    categories=tab.groupby("clonotype")["dpt2_cd4_stim"].mean().reset_index().\
        sort_values("dpt2_cd4_stim")["clonotype"].values[:].tolist()
)
tab = tab.sort_values("clonotype")


# %% [code]
cmap = plt.get_cmap("viridis")

my_pal = dict([
    (x, y)
    for x, y in zip(
        tab["clonotype"].values,
        cmap(tab["dpt2_cd4_stim"].values)
    )
])

# %% [code]
tab = adata_proc_p1.obs.loc[np.logical_not(np.isnan(adata_proc_p1.obs["dpt2_cd4_stim"].values)),:].copy()
tab["IFNG"] = np.asarray(adata_proc_p1[np.logical_not(np.isnan(adata_proc_p1.obs["dpt2_cd4_stim"].values)), "IFNG"].X.todense()).flatten()
tab = tab.loc[[
    (tab_clonotypes_p1_filtered.loc[x, "n_stimulated"] >= 1 and \
    tab_clonotypes_p1_filtered.loc[x, "n_unstimulated"] >= 1)
    if x in tab_clonotypes_p1_filtered.index else False
    for x in tab["clonotype_merged"].values
]].copy()
tab["clonotype_merged"] = pd.Categorical(
    tab["clonotype_merged"].values,
    categories=np.unique(tab["clonotype_merged"].values)
)
tab["clonotype_merged"] = pd.Categorical(
    tab["clonotype_merged"].values,
    categories=tab.groupby("clonotype_merged")["dpt2_cd4_stim"].mean().reset_index().\
        sort_values("dpt2_cd4_stim")["clonotype_merged"].values[:].tolist()
)
tab = tab.sort_values("clonotype_merged")

cmap = plt.get_cmap("viridis")
my_pal = dict([
    (x, y[:-1])
    for x, y in zip(
        tab.groupby("clonotype_merged")["dpt2_cd4_stim"].mean().reset_index()["clonotype_merged"],
        cmap(tab.groupby("clonotype_merged")["dpt2_cd4_stim"].mean().reset_index()["dpt2_cd4_stim"])
    )
])

fig, ax = plt.subplots(nrows=2, ncols=1, figsize=(40, 10))
sb.boxplot(
    data=tab,
    x="clonotype_merged",
    y="proliferation_score",
    palette=my_pal,
    ax=ax[0]
)
ax[0].set_xticklabels(ax[0].get_xticklabels(), rotation=90)
sb.boxplot(
    data=tab,
    x="clonotype_merged",
    y="IFNG",
    palette=my_pal,
    ax=ax[1]
)
ax[1].set_xticklabels(ax[1].get_xticklabels(), rotation=90)
plt.show()

fig, ax = plt.subplots(nrows=1, ncols=1, figsize=(5, 50))
sb.boxplot(
    data=tab,
    y="clonotype_merged",
    x="IFNG",
    palette=my_pal,
    ax=ax
)
ax.set_xticklabels(ax.get_xticklabels(), rotation=90)
plt.tight_layout()
plt.savefig(dir_panels + "sfig5a.pdf")
plt.close()

# %% [markdown]
## Cd8 cells

# %% [markdown]
### Stimulated only

# %% [code]
adata_temp = adata_proc_p1[np.logical_and(
    adata_proc_p1.obs["cell_type"].values == "Cd8",
    adata_proc_p1.obs["condition"].values == "stimulated"
)].copy()
sc.pp.pca(adata_temp)
sc.pp.neighbors(adata_temp, n_neighbors=100)
adata_temp.uns['iroot'] = np.argmax(adata_temp.obsm['X_umap'][:,1])
sc.tl.diffmap(adata_temp)
sc.tl.dpt(adata_temp)
adata_proc_p1.obs["dpt_cd8_stim"] = [
    1. - adata_temp[x].obs["dpt_pseudotime"].values[0]
    if x in adata_temp.obs_names else np.nan
    for x in adata_proc_p1.obs_names
]

# %% [code]
fig, ax = plt.subplots(1,1)
ax = sc.pl.umap(adata_proc_p1, ax=ax, show=False)
ax = sc.pl.umap(adata_proc_p1, color=["dpt_cd8_stim"], ax=ax, show=False)
plt.savefig(dir_panels + "fig1d2.pdf")
plt.show()

sb.distplot(adata_proc_p1.obs["dpt_cd8_stim"].values)
plt.show()
fig, ax = plt.subplots(1,1)
sb.scatterplot(
    x=adata_proc_p1.obs["dpt_cd8_stim"].values,
    y=adata_proc_p1.obs["cd8 cytokine_score"].values,
    ax=ax
)
ax.set_xlabel("pseudotime score")
ax.set_ylabel("cd8 cytokine_score")
plt.show()

sb.scatterplot(
    x=adata_proc_p1.obs["dpt_cd8_stim"].values,
    y=adata_proc_p1.obs["cd8 cytotoxic_score"].values,
    ax=ax
)
ax.set_xlabel("pseudotime score")
plt.show()

# %% [code]
tab = adata_proc_p1.obs.loc[np.logical_not(np.isnan(adata_proc_p1.obs["dpt_cd8_stim"].values)),:].copy()
tab["IFNG"] = np.asarray(adata_proc_p1[np.logical_not(np.isnan(adata_proc_p1.obs["dpt_cd8_stim"].values)), "IFNG"].X.todense()).flatten()
tab = tab.loc[[
    (tab_clonotypes_p1_filtered.loc[x, "n_stimulated"] >= 1 and \
    tab_clonotypes_p1_filtered.loc[x, "n_unstimulated"] >= 1)
    if x in tab_clonotypes_p1_filtered.index else False
    for x in tab["clonotype"].values
]].copy()
tab["clonotype_merged"] = pd.Categorical(
    tab["clonotype_merged"].values,
    categories=np.unique(tab["clonotype_merged"].values)
)
tab["clonotype_merged"] = pd.Categorical(
    tab["clonotype_merged"].values,
    categories=tab.groupby("clonotype_merged")["dpt_cd8_stim"].mean().reset_index().\
        sort_values("dpt_cd8_stim")["clonotype_merged"].values[:].tolist()
)
tab = tab.sort_values("clonotype_merged")

cmap = plt.get_cmap("viridis")
my_pal = dict([
    (x, y[:-1])
    for x, y in zip(
        tab.groupby("clonotype_merged")["dpt_cd8_stim"].mean().reset_index()["clonotype_merged"],
        cmap(tab.groupby("clonotype_merged")["dpt_cd8_stim"].mean().reset_index()["dpt_cd8_stim"])
    )
])

fig, ax = plt.subplots(nrows=3, ncols=1, figsize=(50, 15))
sb.boxplot(
    data=tab,
    x="clonotype_merged",
    y="cd8 cytokine_score",
    palette=my_pal,
    ax=ax[0]
)
ax[0].set_xticklabels(ax[0].get_xticklabels(), rotation=90)
sb.boxplot(
    data=tab,
    x="clonotype_merged",
    y="cd8 cytotoxic_score",
    palette=my_pal,
    ax=ax[1]
)
ax[1].set_xticklabels(ax[1].get_xticklabels(), rotation=90)
sb.boxplot(
    data=tab,
    x="clonotype_merged",
    y="IFNG",
    palette=my_pal,
    ax=ax[2]
)
ax[2].set_xticklabels(ax[2].get_xticklabels(), rotation=90)
plt.tight_layout()
plt.show()

fig, ax = plt.subplots(nrows=1, ncols=1, figsize=(5, 50))
sb.boxplot(
    data=tab,
    y="clonotype_merged",
    x="IFNG",
    palette=my_pal,
    ax=ax
)
ax.set_xticklabels(ax.get_xticklabels(), rotation=90)
plt.tight_layout()
plt.savefig(dir_panels + "sfig5b.pdf")
plt.close()

# %% [code]
tab_clonotypes_p1_filtered["dpt_cd8_stim"] = [
    adata_proc_p1[adata_proc_p1.obs["clonotype_merged"].values == x].obs["dpt_cd8_stim"].values.mean()
    for x in tab_clonotypes_p1_filtered["clonotype_merged"].values
]
```
