## Extended Data File 1 for "Single-cell RNA sequencing reveals *in vivo* signatures of SARS-CoV-2-reactive T cells through ‘reverse phenotyping’": chicago_BALF_convert_scanpy_seurat.ipynb

```
# %% [code]
library(Seurat)
library(SeuratDisk)

# %% [code]
## read data
chicago_BALF <- Convert("chicago_BALF.h5ad", dest = "h5seurat", overwrite = TRUE)

# %% [code]
## convert data
chicago_BALF <- LoadH5Seurat("chicago_BALF.h5seurat")

# %% [code]
## save data
saveRDS(chicago_BALF, "chicago_BALF.rds")
```
