## Extended Data File 1 for "Single-cell RNA sequencing reveals *in vivo* signatures of SARS-CoV-2-reactive T cells through ‘reverse phenotyping’": Macrophage_CD4_Tcells_responsive_vs_unresponsive_stim.ipynb

```
# %% [markdown]
# COVID19: Macrophage - CD4 T cell interaction analysis with NicheNet

# %% [code]
library(Seurat)
library(nichenetr)
library(igraph)
library(tidyverse)
library(ggplot2)
library(readr)
library(patchwork)
library(RColorBrewer)
library(data.table)

# %% [code]
## set working directory
setwd("/home/agando/Documents/Niklas/outputs/Tcell_macrophage_loop_KILIAN/")

## directory for outputs
out.dir <- "Macrophage_CD4_Tcells_responsive_vs_unresponsive_stim/"
dir.create(out.dir)

# %% [code]
## load Chicago data from Grant et al.
chicago_BALF <- readRDS("chicago_BALF.rds")

# %% [code]
## global plotting parameters
options(repr.plot.width=16, repr.plot.height=10)

# %% [code]
## summarize CD4 T cells and proliferating CD4 T cells as CD4 T cells
# create data table storing cell barcodes and the corresponding cell type
cell.metadata <- data.table(barcode = colnames(chicago_BALF),
                            celltype = chicago_BALF$Cluster)
# add meta cell types
cell.metadata[, meta_celltype := ifelse(celltype %in% c("CD4 T cells", "Proliferating CD4 T cells"), "CD4 T cells", paste0(celltype))]
cell.metadata[, meta_celltype := ifelse(celltype %in% c("MoAM1, IL1R2"), "MoAM1", paste0(meta_celltype))]
cell.metadata[, meta_celltype := ifelse(celltype %in% c("MoAM2, CCL18"), "MoAM2", paste0(meta_celltype))]
cell.metadata[, meta_celltype := ifelse(celltype %in% c("MoAM3, VCAN"), "MoAM3", paste0(meta_celltype))]
# add meta data to seurat object
cell.metadata <- data.frame(cell.metadata, row.names = cell.metadata$barcode) # convert to data.frame since Seurat requires a data.frame
cell.metadata$celltype <- NULL
cell.metadata$barcode <- NULL
chicago_BALF <- AddMetaData(chicago_BALF, cell.metadata, col.name =  "meta_celltype")

# add meta cell types as Idents of Seurat object
Idents(chicago_BALF) <- "meta_celltype"

# add rename cell type meta data column
chicago_BALF$celltype <- chicago_BALF$meta_celltype

# %% [code]
## setorder of celltypes for plotting
chicago_BALF$celltype <- factor(chicago_BALF$celltype)
chicago_BALF$celltype <- factor(chicago_BALF$celltype, levels(chicago_BALF$celltype)[c(10,11,12,16,17,
                                                                                       1,2,3,4,5,6,7,8,9,
                                                                                       13,14,15,
                                                                                       18)])

# %% [markdown]
## Perform NicheNet analysis

# %% [code]
## load NicheNet data, model and networks ##
# load NicheNet’s model
ligand_target_matrix = readRDS(url("https://zenodo.org/record/3260758/files/ligand_target_matrix.rds"))

# load receptor-ligand network
lr_network = readRDS(url("https://zenodo.org/record/3260758/files/lr_network.rds"))

# load weighted integrated networks
weighted_networks = readRDS(url("https://zenodo.org/record/3260758/files/weighted_networks.rds"))
weighted_networks_lr = weighted_networks$lr_sig %>% inner_join(lr_network %>% distinct(from,to), by = c("from","to"))

# %% [markdown]
### Define SENDER and RECEIVER cells of cell-cell communication

# %% [code]
## define RECEIVER cells
receiver = "CD4 T cells"
# and retrieve their gene expresssion profiles
expressed_genes_receiver = get_expressed_genes(receiver, chicago_BALF, pct = 0.10)
background_expressed_genes = expressed_genes_receiver %>% .[. %in% rownames(ligand_target_matrix)]

## define SENDER cells
sender_celltypes = c("TRAM1", "TRAM2", "MoAM1", "MoAM2", "MoAM3")
# and retrieve their gene expression profiles
list_expressed_genes_sender = sender_celltypes %>% unique() %>% lapply(get_expressed_genes, chicago_BALF, 0.10) # lapply to get the expressed genes of every sender cell type separately here
expressed_genes_sender = list_expressed_genes_sender %>% unlist() %>% unique()

# %% [markdown]
### Define Gene Set of Interest
DE genes between reactive stimulated and unreactive stimulated CD4 T cells

# %% [code]
## load gene set of interest
scoring_geneset <- read.table("tab_p1_cd4_stimulated_reactiveness.csv", sep = ";", header = TRUE)
geneset_oi <- as.vector(scoring_geneset$gene)
length(geneset_oi)

# %% [markdown]
### Define possible Ligands and Receptors

# %% [code]
## define a set of potential ligands
ligands = lr_network %>% pull(from) %>% unique()
receptors = lr_network %>% pull(to) %>% unique()
## retrieve ligands and receptors expressed in SENDERS and RECEIVERS, respectively 
expressed_ligands = intersect(ligands,expressed_genes_sender)
expressed_receptors = intersect(receptors,expressed_genes_receiver)
## select potential ligands based on the expressed receptors
potential_ligands = lr_network %>% filter(from %in% expressed_ligands & to %in% expressed_receptors) %>% pull(from) %>% unique()

# %% [markdown]
### STEP 1: Ligand activity analysis

# %% [code]
## which ligands are the best predictors of the target genes of the expressed receptors?
ligand_activities = predict_ligand_activities(geneset = geneset_oi, background_expressed_genes = background_expressed_genes, ligand_target_matrix = ligand_target_matrix, potential_ligands = potential_ligands)
ligand_activities = ligand_activities %>% arrange(-pearson) %>% mutate(rank = rank(desc(pearson)))
head(ligand_activities, 10)

# print top hits
best_upstream_ligands = ligand_activities %>% top_n(12, pearson) %>% arrange(-pearson) %>% pull(test_ligand) %>% unique()

# %% [code]
## visualization of expression of top ligands
top.ligands <- DotPlot(subset(chicago_BALF, celltype %in% sender_celltypes), features = best_upstream_ligands %>% rev(), group.by = "celltype", cols = "RdYlBu") + RotatedAxis()
top.ligands

# %% [markdown]
### STEP 2: Target Gene Prediction

# %% [code]
## identify ligand targets
active_ligand_target_links_df = best_upstream_ligands %>% lapply(get_weighted_ligand_target_links,geneset = geneset_oi, ligand_target_matrix = ligand_target_matrix, n = 100) %>% bind_rows() %>% drop_na() # default n = 200
active_ligand_target_links = prepare_ligand_target_visualization(ligand_target_df = active_ligand_target_links_df, ligand_target_matrix = ligand_target_matrix, cutoff = 0.33) # default cutoff = 0.33
## prepare for plotting
order_ligands = intersect(best_upstream_ligands, colnames(active_ligand_target_links)) %>% rev() %>% make.names()
order_targets = active_ligand_target_links_df$target %>% unique() %>% intersect(rownames(active_ligand_target_links)) %>% make.names()
rownames(active_ligand_target_links) = rownames(active_ligand_target_links) %>% make.names() # make.names() for heatmap visualization of genes like H2-T23
colnames(active_ligand_target_links) = colnames(active_ligand_target_links) %>% make.names() # make.names() for heatmap visualization of genes like H2-T23
## plot top ligands based on their targets
vis_ligand_target = active_ligand_target_links[order_targets,order_ligands] %>% t()
p_ligand_target_network = vis_ligand_target %>% make_heatmap_ggplot("Prioritized ligands","Predicted target genes", color = "purple",legend_position = "top", x_axis_position = "top",legend_title = "Regulatory potential")  + theme(axis.text.x = element_text(face = "italic")) + scale_fill_gradient2(low = "whitesmoke",  high = "purple", breaks = c(0,0.0045,0.0090))
p_ligand_target_network

# %% [markdown]
### STEP 3: Target Receptor Prediction

# %% [code]
## infer receptors of top-ranked ligands
lr_network_top = lr_network %>% filter(from %in% best_upstream_ligands & to %in% expressed_receptors) %>% distinct(from,to)
best_upstream_receptors = lr_network_top %>% pull(to) %>% unique()

lr_network_top_df_large = weighted_networks_lr %>% filter(from %in% best_upstream_ligands)
lr_network_top_df_large = lr_network_top_df_large %>% filter(weight > 0.4 )

lr_network_top_df = lr_network_top_df_large %>% spread("from","weight",fill = 0)
lr_network_top_matrix = lr_network_top_df %>% select(-to) %>% as.matrix() %>% magrittr::set_rownames(lr_network_top_df$to)

dist_receptors = dist(lr_network_top_matrix, method = "binary")
hclust_receptors = hclust(dist_receptors, method = "ward.D2")
order_receptors = hclust_receptors$labels[hclust_receptors$order]

dist_ligands = dist(lr_network_top_matrix %>% t(), method = "binary")
hclust_ligands = hclust(dist_ligands, method = "ward.D2")
order_ligands_receptor = hclust_ligands$labels[hclust_ligands$order]

order_receptors = order_receptors %>% intersect(rownames(lr_network_top_matrix))
order_ligands_receptor = order_ligands_receptor %>% intersect(colnames(lr_network_top_matrix))

vis_ligand_receptor_network = lr_network_top_matrix[order_receptors, order_ligands_receptor]
rownames(vis_ligand_receptor_network) = order_receptors %>% make.names()
colnames(vis_ligand_receptor_network) = order_ligands_receptor %>% make.names()

# plot results
p_ligand_receptor_network = vis_ligand_receptor_network %>% t() %>% make_heatmap_ggplot("Ligands","Receptors", color = "mediumvioletred", x_axis_position = "top",legend_title = "Prior interaction potential")
p_ligand_receptor_network

## save ligand receptor heatmap to outdir
png(paste0(out.dir,"p_ligand_receptor_network.png"), width=1200,height=400,units="px")
print(p_ligand_receptor_network)
dev.off()

# %% [markdown]
### STEP 4: Summary Plot

# %% [code]
## ligand activity heatmap
ligand_pearson_matrix = ligand_activities %>% select(pearson) %>% as.matrix() %>% magrittr::set_rownames(ligand_activities$test_ligand)

rownames(ligand_pearson_matrix) = rownames(ligand_pearson_matrix) %>% make.names()
colnames(ligand_pearson_matrix) = colnames(ligand_pearson_matrix) %>% make.names()

vis_ligand_pearson = ligand_pearson_matrix[order_ligands, ] %>% as.matrix(ncol = 1) %>% magrittr::set_colnames("Pearson")
p_ligand_pearson = vis_ligand_pearson %>% make_heatmap_ggplot("Prioritized ligands","Ligand activity", color = "darkorange",legend_position = "top", x_axis_position = "top", legend_title = "Pearson correlation coefficient\ntarget gene prediction ability") + theme(legend.text = element_text(size = 9))

## ligand expression heatmap
order_ligands_adapted = order_ligands
order_ligands_adapted[order_ligands_adapted == "H2.M3"] = "H2-M3" # cf required use of make.names for heatmap visualization | this is not necessary if these ligands are not in the list of prioritized ligands!
order_ligands_adapted[order_ligands_adapted == "H2.T23"] = "H2-T23" # cf required use of make.names for heatmap visualization | this is not necessary if these ligands are not in the list of prioritized ligands!
rotated_dotplot = DotPlot(chicago_BALF %>% subset(celltype %in% sender_celltypes), features = order_ligands_adapted, group.by = "celltype", cols = "RdYlBu") + coord_flip() + theme(legend.text = element_text(size = 10), legend.title = element_text(size = 12)) # flip of coordinates necessary because we want to show ligands in the rows when combining all plots

## build figure
figures_without_legend = cowplot::plot_grid(
  p_ligand_pearson + theme(legend.position = "none", axis.ticks = element_blank()) + theme(axis.title.x = element_text()),
  rotated_dotplot + theme(legend.position = "none", axis.ticks = element_blank(), axis.title.x = element_text(size = 12), axis.text.y = element_text(face = "italic", size = 9), axis.text.x = element_text(size = 9,  angle = 90,hjust = 0)) + ylab("Expression in Sender") + xlab("") + scale_y_discrete(position = "right"),
  p_ligand_target_network + theme(legend.position = "none", axis.ticks = element_blank()) + ylab(""),
  align = "hv",
  nrow = 1,
  rel_widths = c(ncol(vis_ligand_pearson)+6, 4+6, ncol(vis_ligand_target)))

legends = cowplot::plot_grid(
  ggpubr::as_ggplot(ggpubr::get_legend(p_ligand_pearson)),
  ggpubr::as_ggplot(ggpubr::get_legend(rotated_dotplot)),
  ggpubr::as_ggplot(ggpubr::get_legend(p_ligand_target_network)),
  nrow = 1,
  align = "h", rel_widths = c(1.5, 1, 1, 1))

summary_plot = cowplot::plot_grid(figures_without_legend, legends, rel_heights = c(10,5), nrow = 2, align = "hv")
summary_plot

## save summary plot to outdir
png(paste0(out.dir,"summary_plot.png"), width=1200,height=600,units="px")
print(summary_plot)
dev.off()

# %% [markdown]
### Assemble figure

# %% [markdown]
#### Subfigure1: Ligand Target Pearson's correlation

# %% [code]
## prepare data
rownames(vis_ligand_pearson)[3] <- "HLA-DRA"
LigandPearsonCor_df = vis_ligand_pearson %>% 
    data.frame() %>% 
    rownames_to_column("y") %>% 
    tbl_df() %>% 
    gather(x, "score", -y) %>% 
    mutate(y = factor(y, levels = rownames(vis_ligand_pearson), ordered = TRUE), 
           x = factor(x, levels = colnames(vis_ligand_pearson), ordered = TRUE)) %>% 
    mutate(score = ifelse(score == 0, NA, score))

# %% [code]
plot_LigandPearsonCor <- LigandPearsonCor_df %>% 
    ggplot(aes(x, y, fill = score)) + 
    geom_tile(color = "white", size = 0.5) + 
    coord_equal() +
    scale_fill_gradient(low = "gold", high = "firebrick", 
                        na.value = "whitesmoke") + 
    theme_minimal() + 
    theme(panel.grid.minor = element_line(color = "transparent"), 
          panel.grid.major = element_line(color = "transparent"),
          plot.title = element_text(size = 12, hjust = 0.5),
          legend.position = "bottom", 
          legend.title = element_text(size = 10, angle = 0),
          legend.text = element_text(size = 8, hjust = 1), 
          axis.ticks = element_line(size = 0), 
          axis.text.x.top = element_text(angle = 90, hjust = 0), 
          axis.text.x = element_text(angle = 90, hjust = 1), 
          axis.title.y = element_text(size = 12), 
          axis.text.y = element_text()) + 
          scale_x_discrete(position = "top", labels = "Pearson") + 
          xlab(paste0("")) + 
          ylab(paste0("Prioritized ligands")) + 
    labs(title = paste0("Ligand activity")) +
    guides(fill = guide_colourbar("Pearson correlation coefficient\ntarget gene prediction ability", title.position = "top", angle = 90))
plot_LigandPearsonCor

# %% [code]
## save subfigure as pdf
pdf(paste0(out.dir,"figure_sub_1.pdf"), paper = "a4")
plot_LigandPearsonCor
dev.off()

# %% [markdown]
#### Subfigure 2: Expression of Ligands in Macrophage subpopulations

# %% [code]
## retrieve average expression of top ligands
top_ligands_avg_expresssion <- AverageExpression(chicago_BALF, features = rownames(vis_ligand_pearson), verbose = F)
top_ligands_avg_expresssion <- top_ligands_avg_expresssion$RNA

# %% [code]
## filter macrophages
top_ligands_avg_expresssion <- top_ligands_avg_expresssion %>% select(all_of(sender_celltypes))

# %% [code]
## prepare data
top_ligands_avg_expresssion <- data.frame(t(top_ligands_avg_expresssion))
colnames(top_ligands_avg_expresssion) <- rev(best_upstream_ligands)
top_ligands_avg_expresssion$celltype <- rownames(top_ligands_avg_expresssion)

# %% [code]
## center and scale expression values per gene
center_and_scale <- function(x, na.rm = FALSE) (x - mean(x, na.rm = na.rm)) / sd(x, na.rm)
top_ligands_avg_expresssion <- top_ligands_avg_expresssion %>% mutate_at(rev(best_upstream_ligands), center_and_scale)

# %% [code]
## reformat data
rownames(top_ligands_avg_expresssion) <- top_ligands_avg_expresssion$celltype
top_ligands_avg_expresssion$celltype <- NULL
top_ligands_avg_expresssion <- t(top_ligands_avg_expresssion)

# %% [code]
## visualize data
LigandExpr_df = top_ligands_avg_expresssion %>% 
    data.frame() %>% 
    rownames_to_column("y") %>% 
    tbl_df() %>% 
    gather(x, "score", -y) %>% 
    mutate(y = factor(y, levels = rownames(top_ligands_avg_expresssion), ordered = TRUE), 
           x = factor(x, levels = colnames(top_ligands_avg_expresssion), ordered = TRUE)) %>% 
    mutate(score = ifelse(score == 0, NA, score))

plot_LigandExpr <- LigandExpr_df %>% 
    ggplot(aes(x, y, fill = score)) + 
    geom_tile(color = "white", size = 0.5) + 
    coord_equal() +
    scale_fill_gradient2(low = 'deepskyblue3', mid = 'white', high = 'red3') +
    theme_minimal() +
    theme(panel.grid.minor = element_line(color = "transparent"), 
          panel.grid.major = element_line(color = "transparent"),
          plot.title = element_text(size = 12, hjust = 0.5),
          legend.position = "bottom",
          legend.title = element_text(size = 10, angle = 0),
          legend.text = element_text(size = 8, hjust = 1),  
          axis.ticks = element_line(size = 0), 
          axis.text.x.top = element_text(angle = 90, hjust = 0), 
          axis.text.x = element_text(angle = 90, hjust = 1), 
          axis.title = element_text(size = 12), 
          axis.text.y = element_text()) + 
          scale_x_discrete(position = "top") + 
          xlab(paste0("")) + 
          ylab(paste0("")) + 
    labs(title = paste0("Relative expression of ligands")) +
    guides(fill = guide_colourbar("Relative expression of ligands\n(centered and scaled)", title.position = "top"))
plot_LigandExpr

# %% [code]
## save subfigure as pdf
pdf(paste0(out.dir,"figure_sub_2.pdf"), paper = "a4")
plot_LigandExpr
dev.off()

# %% [markdown]
#### Subfigure 3: Ligand Target Heatmap

# %% [code]
## prepare data
LigandTarget_df = vis_ligand_target %>%
    data.frame() %>% 
    rownames_to_column("y") %>% 
    tbl_df() %>% 
    gather(x, "score", -y) %>% 
    mutate(y = factor(y, levels = rownames(vis_ligand_target), ordered = TRUE), 
           x = factor(x, levels = colnames(vis_ligand_target), ordered = TRUE)) %>% 
    mutate(score = ifelse(score == 0, NA, score))

# %% [code]
plot_LigandTarget <- LigandTarget_df %>% 
    ggplot(aes(x, y, fill = score)) + 
    geom_tile(color = "white", size = 0.5) + 
    coord_equal() +
    scale_fill_gradient(low = "#E8EAF6", high = "magenta", na.value = "whitesmoke")  + 
    theme_minimal() + 
    theme(panel.grid.minor = element_line(color = "transparent"), 
          panel.grid.major = element_line(color = "transparent"),
          plot.title = element_text(size = 12, hjust = 0.5),
          legend.position = "bottom",
          legend.title = element_text(size = 10, angle = 0),
          legend.text = element_text(size = 8, hjust = 1),  
          axis.ticks = element_line(size = 0), 
          axis.text.x.top = element_text(angle = 90, hjust = 0), 
          axis.text.x = element_text(angle = 90, hjust = 1),
          axis.title = element_text(size = 12), 
          axis.text.y = element_text()) + 
          scale_x_discrete(position = "top") + 
          xlab(paste0("")) + 
          ylab(paste0("")) + 
    labs(title = paste0("Genes distinguishing reactive from unreactive stimulated CD4 T cells")) +
    guides(fill = guide_colourbar("Regulatory potential", title.position = "top"))
plot_LigandTarget

# %% [code]
## save subfigure as pdf
pdf(paste0(out.dir,"figure_sub_3.pdf"), width = 2000, paper = "a4r")
plot_LigandTarget
dev.off()

# %% [markdown]
#### Subfigure 4: log2FC of target genes in ex vivo PBMCs 

# %% [code]
## extracts data from scoring list
targets_logFC <- data.frame(gene = scoring_geneset$gene,
                            reactive = scoring_geneset$log2fc,
                            unreactive = -(scoring_geneset$log2fc))
targets_logFC <- targets_logFC %>% filter(gene %in% colnames(vis_ligand_target))
targets_logFC <- targets_logFC %>% mutate(gene =  factor(gene, levels = colnames(vis_ligand_target))) %>% arrange(gene)
rownames(targets_logFC) <- targets_logFC$gene
targets_logFC$gene <- NULL

# %% [code]
## reformat data
## visualize data
TargetslogFC_df = targets_logFC %>% 
    data.frame() %>% 
    rownames_to_column("y") %>% 
    tbl_df() %>% 
    gather(x, "score", -y) %>% 
    mutate(y = factor(y, levels = rownames(targets_logFC), ordered = TRUE), 
           x = factor(x, levels = colnames(targets_logFC), ordered = TRUE)) %>% 
    mutate(score = ifelse(score == 0, NA, score))

# %% [code]
## visualize data
plot_TargetslogFC <- TargetslogFC_df %>% 
    ggplot(aes(y,x, fill = score)) + 
    geom_tile(color = "white", size = 0.5) + 
    coord_equal() +
    scale_fill_gradient2(low = 'deepskyblue3', mid = 'white', high = 'red3') +
    theme_minimal() + 
    theme(panel.grid.minor = element_line(color = "transparent"), 
          panel.grid.major = element_line(color = "transparent"),
          plot.title = element_text(size = 12, hjust = 0.5),
          legend.position = "bottom", 
          legend.title = element_text(size = 10, angle = 0),
          legend.text = element_text(size = 8, hjust = 1),
          axis.ticks = element_line(size = 0), 
          axis.text.x.top = element_text(angle = 90, hjust = 0), 
          axis.text.x = element_text(angle = 90, hjust = 1),   
          axis.text.y = element_text(),
          axis.title.x = element_text(hjust = 0.5), 
          axis.title = element_text(size = 12)) + 
          scale_x_discrete(position = "top") +
          xlab(paste0("")) + 
          ylab(paste0("")) + 
    labs(title = paste0("log2FC of top predicted targets in ex vivo stimulated CD4 T cells")) + 
    guides(fill = guide_colourbar("log2FC", title.position = "top"))
plot_TargetslogFC

# %% [code]
## save subfigure as pdf
pdf(paste0(out.dir,"figure_sub_4.pdf"), width = 2000, paper = "a4r")
plot_TargetslogFC
dev.off()

# %% [markdown]
#### Subfigure 5: Percentage of virus infected cells per Macrophage cluster

# %% [code]
## subset data
chicago_macrophages <- subset(chicago_BALF, subset = meta_celltype %in% c("MoAM1", "MoAM2", "MoAM3", "TRAM1", "TRAM2"))
chicago_macrophages$Cluster <- factor(chicago_macrophages$Cluster)
chicago_macrophages$Cluster <- factor(chicago_macrophages$Cluster, c("TRAM1", "TRAM2","MoAM1, IL1R2", "MoAM2, CCL18", "MoAM3, VCAN"))

# %% [code]
## table with n cells (total, infected, uninfected)
n_cells <- as.numeric(table(chicago_macrophages$Cluster)[1:5])
n_uninfected <- as.numeric(table(chicago_macrophages$No.of.SARS.CoV.2.genes, chicago_macrophages$Cluster)[1,])
n_infected <- n_cells - n_uninfected

# %% [code]
## calculate rel frequencies
pct_uninfected <- (n_uninfected/n_cells)*100
pct_uninfected <- round(pct_uninfected, 2)
pct_infected <- (n_infected/n_cells)*100
pct_infected <- round(pct_infected, 2)

# %% [code]
## create data.frame
macrophages_viralgenes <- data.frame(n_cells = n_cells,
                                     n_uninfected = n_uninfected,
                                     n_infected = n_infected,
                                     pct_uninfected = pct_uninfected,
                                     pct_infected = pct_infected)
rownames(macrophages_viralgenes) <- c("TRAM1", "TRAM2","MoAM1", "MoAM2", "MoAM3")

# %% [code]
## dataframe for plotting
macrophages_pct_infected <- macrophages_viralgenes %>% select(pct_infected)

# %% [code]
macrophages_pct_infected

# %% [code]
## reformat data
macrophages_pct_infected = macrophages_pct_infected %>% 
    data.frame() %>% 
    rownames_to_column("y") %>% 
    tbl_df() %>% 
    gather(x, "score", -y) %>% 
    mutate(y = factor(y, levels = rownames(macrophages_pct_infected), ordered = TRUE), 
           x = factor(x, levels = colnames(macrophages_pct_infected), ordered = TRUE)) %>% 
    mutate(score = ifelse(score == 0, NA, score))

# %% [code]
macrophages_pct_infected

# %% [code]
## visualize data
plot_pct_infected <- macrophages_pct_infected %>% 
    ggplot(aes(y, x, fill = score)) + 
    geom_tile(color = "white", size = 0.5) + 
    coord_equal() +
    scale_fill_gradientn(colors = brewer.pal(9,"YlGn"),
                        breaks=seq(0,60,10),
                        limits=c(0,65)) +
    theme_minimal() + 
    theme(panel.grid.minor = element_line(color = "transparent"), 
          panel.grid.major = element_line(color = "transparent"),
          plot.caption = element_text(size = 12, hjust = 0.5),
          legend.position = "bottom", 
          legend.title = element_text(size = 10, angle = 0),
          legend.text = element_text(size = 8, hjust = 1), 
          axis.ticks = element_line(size = 0), 
          axis.text.x.top = element_text(angle = 90, hjust = 0), 
          axis.text.x = element_text(angle = 90, hjust = 1), 
          axis.text.y = element_text()) +  
          scale_x_discrete(position = "bottom") + 
          scale_y_discrete(labels = c("% infected")) +
          ylab(paste0("")) +
          xlab(paste0("")) +  
    labs(caption = paste0("Percentage of Sars-CoV2 infected cells per cluster")) +
    guides(fill = guide_colourbar("% of Sars-CoV2 infected cells", title.position = "top", angle = 0))
plot_pct_infected

# %% [markdown]
#### Composing summary figure 1

# %% [code]
## create summary fig no.1
summary_figure_1 <- plot_LigandPearsonCor + plot_LigandExpr + plot_LigandTarget + plot_layout(guides = 'collect')
summary_figure_1

# %% [code]
## create summary fig no.1 as svg
svg(paste0(out.dir,"figure_complete_1.svg"), width = 16, height = 6)
summary_figure_1
dev.off()

# %% [code]
## save summary fig no.1 as pdf
pdf(paste0(out.dir,"figure_complete_1.pdf"), width = 1600, height = 600, paper = "a4r")
summary_figure_1
dev.off()

# %% [markdown]
#### Composing summary figure 2

# %% [code]
## save summary fig no.2
summary_figure_2 <- plot_LigandTarget / plot_TargetslogFC
summary_figure_2

# %% [code]
## save summary fig no.2 as svg
svg(paste0(out.dir,"figure_complete_2.svg"), width = 10, height = 16)
summary_figure_2
dev.off()

# %% [code]
## save summary fig no.2 as pdf
pdf(paste0(out.dir,"figure_complete_2.pdf"), width = 1600, height = 600, paper = "a4r")
summary_figure_2
dev.off()

# %% [markdown]
#### Composing summary figure 3

# %% [code]
## save summary fig no.3
summary_figure_3 <- plot_LigandExpr / plot_pct_infected + plot_layout(guides = "collect")
summary_figure_3

# %% [code]
## save summary fig no.3 as svg
svg(paste0(out.dir,"figure_complete_3.svg"), width = 10, height = 16)
summary_figure_3
dev.off()

# %% [code]
## save summary fig no.3 as pdf
pdf(paste0(out.dir,"figure_complete_3.pdf"), width = 1600, height = 600, paper = "a4r")
summary_figure_3
dev.off()
```
