## Extended Data File 1 for "Single-cell RNA sequencing reveals *in vivo* signatures of SARS-CoV-2-reactive T cells through ‘reverse phenotyping’": README.txt

Description of individual notebooks 

- 1_basic_analysis.ipynb
Basic analysis of PBMC data of cohort presented in this manuscript, including transcriptomic and TCR analysis.

- 2_pseudotime_analysis.ipynb
Pseudonyme analysis of cohort presented in this manuscript.

- chicago_BALF_convert_scanpy_seurat.ipynb
converts the original anndata object from the 'Chicago cohort' into a SeuratObject (prerequisite to reproduce ext. data figures 16 and 17)

- Macrophage_CD4_Tcells_responsive_unresponsive_stim.ipynb 
performs NicheNet analysis to analyse cell-cell communication between Macrophages ('sender') and CD4 T cells ('receiver') using the responsive vs unresponsive (stim) 'in situ signature score' as geneset of interest (ext. data fig. 16 top panel)
- Macrophage_CD4_Tcells_stim_unstim_unresponsive.ipynb
performs NicheNet analysis to analyse cell-cell communication between Macrophages ('sender') and CD4 T cells ('receiver') using the stimulated vs unstimulated (unresponsive) 'in situ signature score' as geneset of interest(ext. data fig. 16 bottom panel)

- Macrophage_CD8_Tcells_responsive_unresponsive_stim.ipynb
performs NicheNet analysis to analyse cell-cell communication between Macrophages ('sender') and CD8 T cells ('receiver') using the responsive vs unresponsive (stim) 'in situ signature score' as geneset of interest(ext. data fig. 17 top panel)
- Macrophage_CD8_Tcells_stim_unstim_unresponsive.ipynb
performs NicheNet analysis to analyse cell-cell communication between Macrophages ('sender') and CD8 T cells ('receiver') using the stimulated vs unstimulated (unresponsive) 'in situ signature score' as geneset of interest (ext. data fig. 17 bottom panel)

- chicago_BALF_tcells_subclustered_convert_scanpy_seurat.ipynb
converts the 'Chicago cohort' subset of the integrated dataset into a SeuratObject (prerequisite to reproduce figures 14 and 15)

- Tcells_TRAM.ipynb
performs NicheNet analysis to analyse cell-cell communication between Tcell subsets ('sender') and TRAMs ('receiver') using the DE genes between SARS-CoV-2 positive (TRAM2) and negative (TRAM1) TRAMs as geneset of interest (ext. data fig. 14)
- Tcells_MoAM.ipynb
performs NicheNet analysis to analyse cell-cell communication between Tcell subsets ('sender') and MoAMs ('receiver') using the DE genes between SARS-CoV-2 positive (MoAM1) and negative (MoAM2/3) MoAMs as geneset of interest (ext. data fig. 15)
