## Extended Data File 1 for "Single-cell RNA sequencing reveals *in vivo* signatures of SARS-CoV-2-reactive T cells through ‘reverse phenotyping’": Tcells_MoAM.ipynb

```
# %% [markdown]
# COVID19: T cell - MoAM interaction analysis with NicheNet

# %% [code]
library(Seurat)
library(nichenetr)
library(igraph)
library(tidyverse)
library(ggplot2)
library(readr)
library(patchwork)
library(RColorBrewer)
library(viridis)
library(data.table)

# %% [code]
## set working directory
setwd("/home/agando/Documents/Niklas/outputs/Tcell_macrophage_loop_KILIAN/")

## directory for outputs
out.dir <- "Tcells_MoAM/"
dir.create(out.dir)

# %% [code]
## load MoAMs from Chicago data (Grant et al.)
chicago_BALF <- readRDS("chicago_BALF.rds")
## subset only MoAM from Chicago data
chicago_MoAM <- subset(chicago_BALF, subset = Cluster %in% c("MoAM1, IL1R2","MoAM2, CCL18", "MoAM3, VCAN"))
rm(chicago_BALF)
## load T cells from Chicago data (Grant et al.)
chicago_tcells <- readRDS("chicago_BALF_tcells_subclustered.rds")

# %% [code]
## summarize MoAM1 as immature MoAMs and MoAM2/3 as mature MoAMs
# create data table storing cell barcodes and the corresponding cell type
cell.metadata <- data.table(barcode = colnames(chicago_MoAM),
                            celltype = chicago_MoAM$Cluster)
# add meta cell types
cell.metadata[, meta_celltype := ifelse(celltype %in% c("MoAM1, IL1R2"), "immature MoAM", paste0(celltype))]
cell.metadata[, meta_celltype := ifelse(celltype %in% c("MoAM2, CCL18", "MoAM3, VCAN"), "mature MoAM", paste0(meta_celltype))]
# add meta data to seurat object
cell.metadata <- data.frame(cell.metadata, row.names = cell.metadata$barcode) # convert to data.frame since Seurat requires a data.frame
cell.metadata$celltype <- NULL
cell.metadata$barcode <- NULL
chicago_MoAM <- AddMetaData(chicago_MoAM, cell.metadata, col.name =  "meta_celltype")

# add meta cell types as Idents of Seurat object
Idents(chicago_MoAM) <- "meta_celltype"

# add rename cell type meta data column
chicago_MoAM$celltype <- chicago_MoAM$meta_celltype

# %% [code]
## summarize CD4 and CD8 meta T cells based on subclusterings
# create data table storing cell barcodes and the corresponding cell type
cell.metadata <- data.table(barcode = colnames(chicago_tcells),
                            subcluster = chicago_tcells$louvain_2)
# add meta cell types
cell.metadata[, meta_celltype := paste0("Cluster", subcluster)]

# add meta data to seurat object
cell.metadata <- data.frame(cell.metadata, row.names = cell.metadata$barcode) # convert to data.frame since Seurat requires a data.frame
cell.metadata$subcluster <- NULL
cell.metadata$barcode <- NULL
chicago_tcells <- AddMetaData(chicago_tcells, cell.metadata, col.name =  "meta_celltype")

# add meta cell types as Idents of Seurat object
Idents(chicago_tcells) <- "meta_celltype"

# add rename cell type meta data column
chicago_tcells$celltype <- chicago_tcells$meta_celltype

# %% [code]
table(Idents(chicago_tcells))

# %% [code]
## setorder of sender celltypes for plotting 
chicago_tcells$celltype <- factor(chicago_tcells$celltype)
chicago_tcells$celltype <- factor(chicago_tcells$celltype, levels(chicago_tcells$celltype)[c(3,8,11,1,2,13,12,17,5,6,14,4,9,7,15,10,16)])

# %% [code]
levels(chicago_tcells$celltype)

# %% [code]
## global plotting parameters
options(repr.plot.width=16, repr.plot.height=10)

# %% [markdown]
## Perform NicheNet analysis

# %% [code]
## load NicheNet data, model and networks ##
# load NicheNet’s model
ligand_target_matrix = readRDS(url("https://zenodo.org/record/3260758/files/ligand_target_matrix.rds"))

# load receptor-ligand network
lr_network = readRDS(url("https://zenodo.org/record/3260758/files/lr_network.rds"))

# load weighted integrated networks
weighted_networks = readRDS(url("https://zenodo.org/record/3260758/files/weighted_networks.rds"))
weighted_networks_lr = weighted_networks$lr_sig %>% inner_join(lr_network %>% distinct(from,to), by = c("from","to"))

# %% [markdown]
### Define SENDER and RECEIVER cells of cell-cell communication

# %% [code]
## define RECEIVER cells
receiver = c("immature MoAM", "mature MoAM")
# and retrieve their gene expresssion profiles
expressed_genes_receiver = get_expressed_genes(receiver, chicago_MoAM, pct = 0.10)
background_expressed_genes = expressed_genes_receiver %>% .[. %in% rownames(ligand_target_matrix)]

## define SENDER cells
sender_celltypes = paste0("Cluster", seq(0,16,1))
# and retrieve their gene expression profiles
list_expressed_genes_sender = sender_celltypes %>% unique() %>% lapply(get_expressed_genes, chicago_tcells, 0.10) # lapply to get the expressed genes of every sender cell type separately here
expressed_genes_sender = list_expressed_genes_sender %>% unlist() %>% unique()

# %% [markdown]
### Define Gene Set of Interest
DE genes between immature and mature MoAMs

# %% [code]
# select reference for gene set of interest
condition_oi = "mature MoAM" 
condition_reference = "immature MoAM" 
# compute DE genes between immature and mature MoAMs
DE_table_receiver = FindMarkers(object = chicago_MoAM, ident.1 = condition_oi, ident.2 = condition_reference, min.pct = 0.10) %>% rownames_to_column("gene") 
# check number of genes
length(DE_table_receiver$gene)

# %% [code]
## filter gene set
geneset_oi = DE_table_receiver %>% filter(p_val_adj <= 0.05 & abs(avg_logFC) >= 0.25) %>% pull(gene)
geneset_oi = geneset_oi %>% .[. %in% rownames(ligand_target_matrix)]
length(geneset_oi)

# %% [markdown]
### Define possible Ligands and Receptors

# %% [code]
## define a set of potential ligands
ligands = lr_network %>% pull(from) %>% unique()
receptors = lr_network %>% pull(to) %>% unique()
## retrieve ligands and receptors expressed in SENDERS and RECEIVERS, respectively 
expressed_ligands = intersect(ligands,expressed_genes_sender)
expressed_receptors = intersect(receptors,expressed_genes_receiver)
## select potential ligands based on the expressed receptors
potential_ligands = lr_network %>% filter(from %in% expressed_ligands & to %in% expressed_receptors) %>% pull(from) %>% unique()

# %% [markdown]
### STEP 1: Ligand activity analysis

# %% [code]
## which ligands are the best predictors of the target genes of the expressed receptors?
ligand_activities = predict_ligand_activities(geneset = geneset_oi, background_expressed_genes = background_expressed_genes, ligand_target_matrix = ligand_target_matrix, potential_ligands = potential_ligands)
ligand_activities = ligand_activities %>% arrange(-pearson) %>% mutate(rank = rank(desc(pearson)))
head(ligand_activities, 10)

# print top hits
best_upstream_ligands = ligand_activities %>% top_n(12, pearson) %>% arrange(-pearson) %>% pull(test_ligand) %>% unique()

# %% [code]
## visualization of expression of top ligands
top.ligands <- DotPlot(subset(chicago_tcells, celltype %in% sender_celltypes), features = best_upstream_ligands %>% rev(), group.by = "celltype", cols = "RdYlBu") + RotatedAxis()
top.ligands

# %% [markdown]
### STEP 2: Target Gene Prediction

# %% [code]
## identify ligand targets
active_ligand_target_links_df = best_upstream_ligands %>% lapply(get_weighted_ligand_target_links,geneset = geneset_oi, ligand_target_matrix = ligand_target_matrix, n = 75) %>% bind_rows() %>% drop_na() # default n = 200
active_ligand_target_links = prepare_ligand_target_visualization(ligand_target_df = active_ligand_target_links_df, ligand_target_matrix = ligand_target_matrix, cutoff = 0.33) # default cutoff = 0.33
## prepare for plotting
order_ligands = intersect(best_upstream_ligands, colnames(active_ligand_target_links)) %>% rev() %>% make.names()
order_targets = active_ligand_target_links_df$target %>% unique() %>% intersect(rownames(active_ligand_target_links)) %>% make.names()
rownames(active_ligand_target_links) = rownames(active_ligand_target_links) %>% make.names() # make.names() for heatmap visualization of genes like H2-T23
colnames(active_ligand_target_links) = colnames(active_ligand_target_links) %>% make.names() # make.names() for heatmap visualization of genes like H2-T23
## plot top ligands based on their targets
vis_ligand_target = active_ligand_target_links[order_targets,order_ligands] %>% t()
p_ligand_target_network = vis_ligand_target %>% make_heatmap_ggplot("Prioritized ligands","Predicted target genes", color = "purple",legend_position = "top", x_axis_position = "top",legend_title = "Regulatory potential")  + theme(axis.text.x = element_text(face = "italic")) + scale_fill_gradient2(low = "whitesmoke",  high = "purple", breaks = c(0,0.0045,0.0090))
p_ligand_target_network

# %% [markdown]
### STEP 3: Target Receptor Prediction

# %% [code]
## infer receptors of top-ranked ligands
lr_network_top = lr_network %>% filter(from %in% best_upstream_ligands & to %in% expressed_receptors) %>% distinct(from,to)
best_upstream_receptors = lr_network_top %>% pull(to) %>% unique()

lr_network_top_df_large = weighted_networks_lr %>% filter(from %in% best_upstream_ligands)
lr_network_top_df_large = lr_network_top_df_large %>% filter(weight > 0.4 )

lr_network_top_df = lr_network_top_df_large %>% spread("from","weight",fill = 0)
lr_network_top_matrix = lr_network_top_df %>% select(-to) %>% as.matrix() %>% magrittr::set_rownames(lr_network_top_df$to)

dist_receptors = dist(lr_network_top_matrix, method = "binary")
hclust_receptors = hclust(dist_receptors, method = "ward.D2")
order_receptors = hclust_receptors$labels[hclust_receptors$order]

dist_ligands = dist(lr_network_top_matrix %>% t(), method = "binary")
hclust_ligands = hclust(dist_ligands, method = "ward.D2")
order_ligands_receptor = hclust_ligands$labels[hclust_ligands$order]

order_receptors = order_receptors %>% intersect(rownames(lr_network_top_matrix))
order_ligands_receptor = order_ligands_receptor %>% intersect(colnames(lr_network_top_matrix))

vis_ligand_receptor_network = lr_network_top_matrix[order_receptors, order_ligands_receptor]
rownames(vis_ligand_receptor_network) = order_receptors %>% make.names()
colnames(vis_ligand_receptor_network) = order_ligands_receptor %>% make.names()

# plot results
p_ligand_receptor_network = vis_ligand_receptor_network %>% t() %>% make_heatmap_ggplot("Ligands","Receptors", color = "mediumvioletred", x_axis_position = "top",legend_title = "Prior interaction potential")
p_ligand_receptor_network

## save ligand receptor heatmap to outdir
png(paste0(out.dir,"p_ligand_receptor_network.png"), width=1200,height=400,units="px")
print(p_ligand_receptor_network)
dev.off()

# %% [markdown]
### STEP 4: Summary Plot

# %% [code]
## ligand activity heatmap
ligand_pearson_matrix = ligand_activities %>% select(pearson) %>% as.matrix() %>% magrittr::set_rownames(ligand_activities$test_ligand)

rownames(ligand_pearson_matrix) = rownames(ligand_pearson_matrix) %>% make.names()
colnames(ligand_pearson_matrix) = colnames(ligand_pearson_matrix) %>% make.names()

vis_ligand_pearson = ligand_pearson_matrix[order_ligands, ] %>% as.matrix(ncol = 1) %>% magrittr::set_colnames("Pearson")
p_ligand_pearson = vis_ligand_pearson %>% make_heatmap_ggplot("Prioritized ligands","Ligand activity", color = "darkorange",legend_position = "top", x_axis_position = "top", legend_title = "Pearson correlation coefficient\ntarget gene prediction ability") + theme(legend.text = element_text(size = 9))

## ligand expression heatmap
order_ligands_adapted = order_ligands
order_ligands_adapted[order_ligands_adapted == "H2.M3"] = "H2-M3" # cf required use of make.names for heatmap visualization | this is not necessary if these ligands are not in the list of prioritized ligands!
order_ligands_adapted[order_ligands_adapted == "H2.T23"] = "H2-T23" # cf required use of make.names for heatmap visualization | this is not necessary if these ligands are not in the list of prioritized ligands!
rotated_dotplot = DotPlot(chicago_tcells %>% subset(celltype %in% sender_celltypes), features = order_ligands_adapted, group.by = "celltype", cols = "RdYlBu") + coord_flip() + theme(legend.text = element_text(size = 10), legend.title = element_text(size = 12)) # flip of coordinates necessary because we want to show ligands in the rows when combining all plots

## build figure
figures_without_legend = cowplot::plot_grid(
  p_ligand_pearson + theme(legend.position = "none", axis.ticks = element_blank()) + theme(axis.title.x = element_text()),
  rotated_dotplot + theme(legend.position = "none", axis.ticks = element_blank(), axis.title.x = element_text(size = 12), axis.text.y = element_text(face = "italic", size = 9), axis.text.x = element_text(size = 9,  angle = 90,hjust = 0)) + ylab("Expression in Sender") + xlab("") + scale_y_discrete(position = "right"),
  p_ligand_target_network + theme(legend.position = "none", axis.ticks = element_blank()) + ylab(""),
  align = "hv",
  nrow = 1,
  rel_widths = c(ncol(vis_ligand_pearson)+8, 16+6, ncol(vis_ligand_target)))

legends = cowplot::plot_grid(
  ggpubr::as_ggplot(ggpubr::get_legend(p_ligand_pearson)),
  ggpubr::as_ggplot(ggpubr::get_legend(rotated_dotplot)),
  ggpubr::as_ggplot(ggpubr::get_legend(p_ligand_target_network)),
  nrow = 1,
  align = "h", rel_widths = c(1.5, 1, 1, 1))

summary_plot = cowplot::plot_grid(figures_without_legend, legends, rel_heights = c(10,5), nrow = 2, align = "hv")
summary_plot

## save summary plot to outdir
png(paste0(out.dir,"summary_plot.png"), width=1200,height=600,units="px")
print(summary_plot)
dev.off()

# %% [markdown]
### Assemble figure

# %% [markdown]
#### Subfigure1: Ligand Target Pearson's correlation

# %% [code]
## prepare data
LigandPearsonCor_df = vis_ligand_pearson %>% 
    data.frame() %>% 
    rownames_to_column("y") %>% 
    tbl_df() %>% 
    gather(x, "score", -y) %>% 
    mutate(y = factor(y, levels = rownames(vis_ligand_pearson), ordered = TRUE), 
           x = factor(x, levels = colnames(vis_ligand_pearson), ordered = TRUE)) %>% 
    mutate(score = ifelse(score == 0, NA, score))

# %% [code]
## visualize data
plot_LigandPearsonCor <- LigandPearsonCor_df %>% 
    ggplot(aes(x, y, fill = score)) + 
    geom_tile(color = "white", size = 0.5) + 
    coord_equal() +
    scale_fill_gradient(low = "gold", high = "firebrick", 
                        na.value = "whitesmoke") + 
    theme_minimal() + 
    theme(panel.grid.minor = element_line(color = "transparent"), 
          panel.grid.major = element_line(color = "transparent"),
          plot.title = element_text(size = 12, hjust = 0.5),
          legend.position = "bottom", 
          legend.title = element_text(size = 10, angle = 0),
          legend.text = element_text(size = 8, hjust = 1), 
          axis.ticks = element_line(size = 0), 
          axis.text.x.top = element_text(angle = 90, hjust = 0), 
          axis.text.x = element_text(angle = 90, hjust = 1), 
          axis.title.y = element_text(size = 12), 
          axis.text.y = element_text()) + 
          scale_x_discrete(position = "top", labels = "Pearson") + 
          xlab(paste0("")) + 
          ylab(paste0("Prioritized ligands")) + 
    labs(title = paste0("Ligand activity")) +
    guides(fill = guide_colourbar("Pearson correlation coefficient\ntarget gene prediction ability", title.position = "top", angle = 90))
plot_LigandPearsonCor

# %% [code]
## save subfigure as pdf
pdf(paste0(out.dir,"figure_sub_1.pdf"), paper = "a4")
plot_LigandPearsonCor
dev.off()

# %% [markdown]
#### Subfigure 2: Expression of Ligands in T cell subsets

# %% [code]
## retrieve average expression of top ligands
top_ligands_avg_expresssion <- AverageExpression(chicago_tcells, features = rownames(vis_ligand_pearson), verbose = F)
top_ligands_avg_expresssion <- top_ligands_avg_expresssion$RNA

# %% [code]
## filter macrophages
top_ligands_avg_expresssion <- top_ligands_avg_expresssion %>% select(all_of(sender_celltypes))

# %% [code]
## prepare data
top_ligands_avg_expresssion <- data.frame(t(top_ligands_avg_expresssion))
colnames(top_ligands_avg_expresssion) <- rev(best_upstream_ligands)
top_ligands_avg_expresssion$celltype <- rownames(top_ligands_avg_expresssion)

# %% [code]
## center and scale expression values per gene
center_and_scale <- function(x, na.rm = FALSE) (x - mean(x, na.rm = na.rm)) / sd(x, na.rm)
top_ligands_avg_expresssion <- top_ligands_avg_expresssion %>% mutate_at(rev(best_upstream_ligands), center_and_scale)

# %% [code]
## reformat data
rownames(top_ligands_avg_expresssion) <- top_ligands_avg_expresssion$celltype
top_ligands_avg_expresssion$celltype <- NULL
top_ligands_avg_expresssion <- t(top_ligands_avg_expresssion)

# %% [code]
## reorder columns (T cell subsets) for plotting
top_ligands_avg_expresssion <- top_ligands_avg_expresssion[, c(11,16,4,1,2,6,5,10,13,14,7,12,17,15,8,3,9)]

# %% [code]
## prepare data for plotting
LigandExpr_df = top_ligands_avg_expresssion %>% 
    data.frame() %>% 
    rownames_to_column("y") %>% 
    tbl_df() %>% 
    gather(x, "score", -y) %>% 
    mutate(y = factor(y, levels = rownames(top_ligands_avg_expresssion), ordered = TRUE), 
           x = factor(x, levels = colnames(top_ligands_avg_expresssion), ordered = TRUE)) %>% 
    mutate(score = ifelse(score == 0, NA, score))

# %% [code]
## visualize data
plot_LigandExpr <- LigandExpr_df %>% 
    ggplot(aes(x, y, fill = score)) + 
    geom_tile(color = "white", size = 0.5) + 
    coord_equal() +
    ggsci::scale_fill_gsea() +
    theme_minimal() +
    theme(panel.grid.minor = element_line(color = "transparent"), 
          panel.grid.major = element_line(color = "transparent"),
          plot.title = element_text(size = 12, hjust = 0.5),
          legend.position = "bottom",
          legend.title = element_text(size = 10, angle = 0),
          legend.text = element_text(size = 8, hjust = 1),  
          axis.ticks = element_line(size = 0), 
          axis.text.x.top = element_text(angle = 90, hjust = 0), 
          axis.text.x = element_text(angle = 90, hjust = 1), 
          axis.title = element_text(size = 12), 
          axis.text.y = element_text()) + 
          scale_x_discrete(labels = as.vector(paste("Louvain", c(10,15,3,0,1,5,4,9,12,13,6,11,16,14,7,2,8))), position = "top") + 
          xlab("") + 
          ylab(paste0("")) + 
    labs(title = paste0("Relative expression of ligands by T cell subsets")) +
    guides(fill = guide_colourbar("Relative expression of ligands\n(centered and scaled)", title.position = "top"))
plot_LigandExpr

# %% [code]
## save subfigure as pdf
pdf(paste0(out.dir,"figure_sub_2.pdf"), paper = "a4")
plot_LigandExpr
dev.off()

# %% [markdown]
#### Subfigure 3: Ligand Target Heatmap

# %% [code]
## prepare data
LigandTarget_df = vis_ligand_target %>%
    data.frame() %>% 
    rownames_to_column("y") %>% 
    tbl_df() %>% 
    gather(x, "score", -y) %>% 
    mutate(y = factor(y, levels = rownames(vis_ligand_target), ordered = TRUE), 
           x = factor(x, levels = colnames(vis_ligand_target), ordered = TRUE)) %>% 
    mutate(score = ifelse(score == 0, NA, score))

# %% [code]
## visualize data
plot_LigandTarget <- LigandTarget_df %>% 
    ggplot(aes(x, y, fill = score)) + 
    geom_tile(color = "white", size = 0.5) + 
    coord_equal() +
    scale_fill_gradient(low = "#E8EAF6", high = "magenta", na.value = "whitesmoke")  + 
    theme_minimal() + 
    theme(panel.grid.minor = element_line(color = "transparent"), 
          panel.grid.major = element_line(color = "transparent"),
          plot.title = element_text(size = 12, hjust = 0.5),
          legend.position = "bottom",
          legend.title = element_text(size = 10, angle = 0),
          legend.text = element_text(size = 8, hjust = 1),  
          axis.ticks = element_line(size = 0), 
          axis.text.x.top = element_text(angle = 90, hjust = 0), 
          axis.text.x = element_text(angle = 90, hjust = 1),
          axis.title = element_text(size = 12), 
          axis.text.y = element_text()) + 
          scale_x_discrete(position = "top") + 
          xlab(paste0("")) + 
          ylab(paste0("")) + 
    labs(title = paste0("Genes distinguishing mature from immature MoAMs")) +
    guides(fill = guide_colourbar("Regulatory potential", title.position = "top"))
plot_LigandTarget

# %% [code]
## save subfigure as pdf
pdf(paste0(out.dir,"figure_sub_3.pdf"), width = 2000, paper = "a4r")
plot_LigandTarget
dev.off()

# %% [markdown]
#### Subfigure 4: log2FC of target genes in mature vs immature MoAMs

# %% [code]
## extracts data from scoring list
targets_logFC <- data.frame(gene = DE_table_receiver$gene,
                            mature = DE_table_receiver$avg_logFC, # higher in mature MoAMs
                            immature = -(DE_table_receiver$avg_logFC))
targets_logFC <- targets_logFC %>% filter(gene %in% colnames(vis_ligand_target))
targets_logFC <- targets_logFC %>% mutate(gene =  factor(gene, levels = colnames(vis_ligand_target))) %>% arrange(gene)
rownames(targets_logFC) <- targets_logFC$gene
targets_logFC$gene <- NULL

# %% [code]
## reformat data
## visualize data
TargetslogFC_df = targets_logFC %>% 
    data.frame() %>% 
    rownames_to_column("y") %>% 
    tbl_df() %>% 
    gather(x, "score", -y) %>% 
    mutate(y = factor(y, levels = rownames(targets_logFC), ordered = TRUE), 
           x = factor(x, levels = colnames(targets_logFC), ordered = TRUE)) %>% 
    mutate(score = ifelse(score == 0, NA, score))

# %% [code]
## visualize data
plot_TargetslogFC <- TargetslogFC_df %>% 
    ggplot(aes(y,x, fill = score)) + 
    geom_tile(color = "white", size = 0.5) + 
    coord_equal() +
    ggsci::scale_fill_gsea() +
    theme_minimal() + 
    theme(panel.grid.minor = element_line(color = "transparent"), 
          panel.grid.major = element_line(color = "transparent"),
          plot.title = element_text(size = 12, hjust = 0.5),
          legend.position = "bottom", 
          legend.title = element_text(size = 10, angle = 0),
          legend.text = element_text(size = 8, hjust = 1),
          axis.ticks = element_line(size = 0), 
          axis.text.x.top = element_text(angle = 90, hjust = 0), 
          axis.text.x = element_text(angle = 90, hjust = 1),   
          axis.text.y = element_text(),
          axis.title.x = element_text(hjust = 0.5), 
          axis.title = element_text(size = 12)) + 
          scale_x_discrete(position = "top") +
          xlab(paste0("")) + 
          ylab(paste0("")) + 
    labs(title = paste0("log2FC of top predicted targets in mature vs immature MoAMs")) + 
    guides(fill = guide_colourbar("log2FC", title.position = "top"))
plot_TargetslogFC

# %% [code]
## save subfigure as pdf
pdf(paste0(out.dir,"figure_sub_4.pdf"), width = 2000, paper = "a4r")
plot_TargetslogFC
dev.off()

# %% [markdown]
#### Subfigure 5: Average score per T cell subset

# %% [code]
## table with barcodes + corresponding scores
all_scores <- data.frame(tcell_subset = chicago_tcells$celltype,
                       cd4_reactive_unreactive_stim = chicago_tcells$X1..stim.reactive.CD4,
                       cd8_reactive_unreactive_stim = chicago_tcells$X2..stim.reactive.CD8,
                       cd4_reactive_unreactive_unstim = chicago_tcells$X3..unstim.reactive.CD4,
                       cd8_reactive_unreactive_unstim = chicago_tcells$X4..unstim.reactive.CD8,
                       cd4_stim_unstim_reactive = chicago_tcells$X5..reactive.cond.CD4,
                       cd8_stim_unstim_reactive = chicago_tcells$X6..reactive.cond.CD8,
                       cd4_stim_unstim_unreactive = chicago_tcells$X7..non.reactive.cond.CD4,
                       cd8_stim_unstim_unreactive = chicago_tcells$X8..non.reactive.cond.CD8)

# %% [code]
## compute stratfied means by T cell subclusters
tcell_scores <- all_scores  %>% group_by(tcell_subset) %>%
  summarise(cd4_reactive_unreactive_stim_mean = mean(cd4_reactive_unreactive_stim, na.rm = TRUE),
            cd8_reactive_unreactive_stim_mean = mean(cd8_reactive_unreactive_stim, na.rm = TRUE),
            cd4_reactive_unreactive_unstim_mean = mean(cd4_reactive_unreactive_unstim, na.rm = TRUE),
            cd8_reactive_unreactive_unstim_mean = mean(cd8_reactive_unreactive_unstim, na.rm = TRUE),
            cd4_stim_unstim_reactive_mean = mean(cd4_stim_unstim_reactive, na.rm = TRUE),
            cd8_stim_unstim_reactive_mean = mean(cd8_stim_unstim_reactive, na.rm = TRUE),
            cd4_stim_unstim_unreactive_mean = mean(cd4_stim_unstim_unreactive, na.rm = TRUE),
            cd8_stim_unstim_unreactive_mean = mean(cd8_stim_unstim_unreactive, na.rm = TRUE)) %>%
  data.frame()

# %% [code]
## center and scale expression values per gene
center_and_scale <- function(x) (x - mean(x, na.rm = TRUE)) / sd(x, na.rm = TRUE)
tcell_scores_normalized <- tcell_scores %>% mutate_at(colnames(tcell_scores)[-1], center_and_scale)

# %% [code]
tcell_scores_normalized$tcell_subset <- NULL

# %% [code]
tcell_scores_normalized <- tcell_scores_normalized[, c(8,6,4,2,7,5,3,1)]

# %% [code]
## reformat data
tcell_scores_df = tcell_scores_normalized %>% 
    data.frame() %>% 
    rownames_to_column("y") %>% 
    tbl_df() %>% 
    gather(x, "score", -y) %>% 
    mutate(y = factor(y, levels = rownames(tcell_scores_normalized), ordered = TRUE), 
           x = factor(x, levels = colnames(tcell_scores_normalized), ordered = TRUE)) %>% 
    mutate(score = ifelse(score == 0, NA, score))

# %% [code]
labels <- c("CD8 stim vs unstim in unreactive", "CD8 stim vs unstim in reactive",
            "CD8 reactive vs unreactive in unstim", "CD8 reactive vs unreactive in stim",
            "CD4 stim vs unstim in unreactive", "CD4 stim vs unstim in reactive",
            "CD4 reactive vs unreactive in unstim", "CD4 reactive vs unreactive in stim")

# %% [code]
## visualize data
plot_avg_scores <- tcell_scores_df %>% 
    ggplot(aes(y, x, fill = score)) + 
    geom_tile(color = "white", size = 0.5) + 
    coord_equal() +
    scale_fill_viridis(option="C", na.value = "lightgrey") +
    #scale_fill_gradientn(colors = viridis_pal()(12), na.value = "whitesmoke",
    #                    breaks=seq(-3,2.2, 1),
    #                    limits=c(-3,2.2)) +
    theme_minimal() + 
    theme(panel.grid.minor = element_line(color = "transparent"), 
          panel.grid.major = element_line(color = "transparent"),
          plot.caption = element_text(size = 12, hjust = 0.5),
          legend.position = "bottom", 
          legend.title = element_text(size = 10, angle = 0),
          legend.text = element_text(size = 8, hjust = 1), 
          axis.ticks = element_line(size = 0), 
          axis.text.x.top = element_text(angle = 90, hjust = 0), 
          axis.text.x = element_text(angle = 90, hjust = 1), 
          axis.text.y = element_text()) +  
          scale_x_discrete(labels = as.vector(paste("Louvain", c(10,15,3,0,1,5,4,9,12,13,6,11,16,14,7,2,8))), position = "bottom") + 
          scale_y_discrete(labels = labels) +
          ylab(paste0("")) +
          xlab(paste0("")) +  
    labs(caption = paste0("Score by T cell subsets")) +
    guides(fill = guide_colourbar("Score\n(centered and scaled)", title.position = "top", angle = 0))
plot_avg_scores

# %% [code]
## save subfigure as pdf
pdf(paste0(out.dir,"figure_sub_5.pdf"), width = 2000, paper = "a4r")
plot_avg_scores
dev.off()

# %% [markdown]
#### Composing summary figure 1

# %% [code]
## create summary fig no.1
summary_figure_1 <- plot_LigandPearsonCor + plot_LigandExpr + plot_LigandTarget + plot_layout(guides = 'collect')
summary_figure_1

# %% [code]
## create summary fig no.1 as svg
svg(paste0(out.dir,"figure_complete_1.svg"), width = 16, height = 6)
summary_figure_1
dev.off()

# %% [code]
## save summary fig no.1 as pdf
pdf(paste0(out.dir,"figure_complete_1.pdf"), width = 1600, height = 600, paper = "a4r")
summary_figure_1
dev.off()

# %% [markdown]
#### Composing summary figure 2

# %% [code]
## save summary fig no.2
summary_figure_2 <- plot_LigandTarget / plot_TargetslogFC
summary_figure_2

# %% [code]
## save summary fig no.2 as svg
svg(paste0(out.dir,"figure_complete_2.svg"), width = 10, height = 16)
summary_figure_2
dev.off()

# %% [code]
## save summary fig no.2 as pdf
pdf(paste0(out.dir,"figure_complete_2.pdf"), width = 1600, height = 600, paper = "a4r")
summary_figure_2
dev.off()

# %% [markdown]
#### Composing summary figure 3

# %% [code]
## save summary fig no.3
summary_figure_3 <- plot_LigandExpr / plot_avg_scores + plot_layout(guides = "collect")
summary_figure_3

# %% [code]
## save summary fig no.3 as svg
svg(paste0(out.dir,"figure_complete_3.svg"), width = 10, height = 16)
summary_figure_3
dev.off()

# %% [code]
## save summary fig no.3 as pdf
pdf(paste0(out.dir,"figure_complete_3.pdf"), width = 1600, height = 600, paper = "a4r")
summary_figure_3
dev.off()

# %% [code]
```
